# Integrative Multi-Omics Analysis of Molecular Associations and Biological Pathways Underlying Neurodevelopmental Conditions in a Twin Cohort

**DOI:** 10.64898/2026.01.16.26344104

**Authors:** Yali Zhang, Bonita van Waardenburg, Karl Lundin Remnélius, Johan Isaksson, Kate Pearse, Andreas Göteson, Marie Vahter, Thomas Bourgeron, Jonathan Swann, Maria Kippler, Mikael Landén, Rochellys Diaz Heijtz, Sven Bölte, Kristiina Tammimies

**Affiliations:** Center of Neurodevelopmental Disorders (KIND), Center for Psychiatry Research, Department of Women’s and Children’s Health, Karolinska Institutet, Stockholm, Sweden; Astrid Lindgren Children’s Hospital, Karolinska University Hospital, Region Stockholm, Stockholm, Sweden; Child and Adolescent Psychiatry Unit, Department of Medical Sciences, Uppsala University, Uppsala, Sweden; School of Human Development and Health, Faculty of Medicine, University of Southampton, UK; Institute of Neuroscience and Physiology, Sahlgrenska Academy at University of Gothenburg, Gothenburg, Sweden; Institute of Environmental Medicine, Karolinska Institutet, Stockholm, Sweden; Human Genetics and Cognitive Functions, Institut Pasteur, Université de Paris, Paris, France; Department of Medical Epidemiology and Biostatistics, Karolinska Institutet, Stockholm, Sweden; Department of Neuroscience, Karolinska Institutet, Stockholm, Sweden; Child and Adolescent Psychiatry, Stockholm Health Care Services, Region Stockholm, Stockholm, Sweden; Curtin Autism Research Group, Curtin School of Allied Health, Curtin University, Perth, Western Australia

## Abstract

Neurodevelopmental conditions (NDCs) arise from complex genetic-environmental interactions, yet their molecular underpinnings remain incompletely defined. We applied an integrative multi-omics approach within a deeply phenotyped twin cohort to identify systemic molecular signatures and pathways associated with NDCs. Our study included 237 twins aged 8-28 years from the Roots of Autism and ADHD Twin Study in Sweden (RATSS), with one or more omics layers available per individual, including serum and cerebrospinal fluid proteomics, urine and fecal metabolomics, blood metallomics, and whole-genome sequencing in a subset of 14 monozygotic (MZ) twin pairs. Using the DIABLO framework, we identified cross-omics features that distinguish individuals with and without NDCs. The identified features converged on enriched metabolic and immune-related pathways, including purine metabolism, lysine degradation, PI3K-Akt, and MAPK signaling. Several key molecules also showed differential abundance at the single-molecule level in generalized estimating equation (GEE) models. ADA protein, pyrimidine-related metabolites (thymidine, glutamine, and 2’-deoxyuridine), and copper showed suggestive associations within NDC-discordant MZ twin pairs, pointing to potential non-shared condition-related effects, while flavin mononucleotide and manganese associations with NDCs were influenced by shared genetic or familial environmental factors. Our findings indicate that NDC-related molecular alterations manifest across multiple biological layers and tissues, and are detectable through integrative systems-level analysis. Our scalable framework provides critical insights into altered metabolic and immune processes in NDCs and highlights candidate features that may inform future biomarker development and mechanistic research in precision psychiatry.

## 1 Introduction

Neurodevelopmental conditions (NDCs), such as autism, attention-deficit/hyperactivity disorder (ADHD), motor and language disorders, are defined by cognitive and behavioral difficulties causing functional challenges in social, academic, and occupational arenas [1]. In high-income countries, approximately 15% of children aged 3 to 17 years are diagnosed with NDCs [2], which exhibit pronounced heterogeneity while sharing clinical features across diagnoses [3–5]. Sex and gender are also important contributors to the biological and behavioral variability [6]. NDC diagnoses frequently co-occur, and individuals with NDCs are also at increased risk for somatic and psychiatric disorders [1, 7]. Although the origins, physiological mechanisms, and consequences of NDCs remain incompletely understood, both genetic and environmental factors contribute substantially to NDC liability, with heritability estimates from 0.66 across conditions to approximately 0.83 for autism specifically in children and adolescents [8, 9]. Metabolic, immune, and inflammatory factors also contribute to NDC likelihood and may interact with genetic susceptibility, further complicating their etiology [10–13].

Single-omics studies have contributed to elucidating the biological underpinnings of NDCs. Genomic research has identified hundreds of NDC-associated genes enriched for rare and de novo variants [14–16], many of which disrupt transcriptional regulation, synaptic function, and brain development [17–19]. Proteomic analyses of peripheral blood and cerebrospinal fluid (CSF) have revealed altered expression profiles linked to mitochondrial dysfunction, neuroinflammation, and metabolic processes in NDCs, with some proteins correlating with clinical symptoms [20, 21]. Because metabolic alterations are sensitive to gene–environment interactions, they offer particular promise for understanding NDC physiology [22, 23]. Metabolomic analyses of non-invasive biospecimens, such as fecal and urine samples, have demonstrated altered microbial-host co-metabolism and amino acid signatures in children with NDCs [24, 25]. Despite the breadth of reported molecular associations, no highly reliable NDC biomarker has been established [26].

Growing omics datasets increasingly enable integration of multiple molecular layers to investigate the complex interplay between biological systems. However, applications in NDC research have only recently emerged [27]. Early efforts typically combined two or three omics layers, most commonly archived genome-wide data with proteomic or transcriptomic data, to assess how genetic variants shape NDC physiology through altered gene or protein expression [28]. Metabolomics has frequently complemented these layers by linking molecular changes to functional pathways. In autism, integrated urine metabolomic and proteomic analyses implicated altered neuroimmune pathways [29], while cross-tissue meta-analyses identified shared molecular signatures across the central nervous system, circulatory system, and non-invasive biofluids [30]. Despite limited sample sizes, integration of placental transcriptomics and metabolomics has shown diagnostic promise, with a machine-learning model classifying NDCs at >95% accuracy using only a few components [31]. Recently, urine metabolomics combined with genomics and epigenomics revealed differential association patterns in ADHD, including strong correlations between specific CpG loci and ADHD polygenic scores [32].

Beyond traditional omics layers, incorporating nutritional status and toxic metal exposure may more directly connect molecular findings to modifiable dietary and environmental factors in the etiology of NDC [33]. Both essential metals (*e.g.,* copper, manganese, and zinc) and toxic metals (*e.g.,* lead, mercury, and arsenic) can influence brain development and function, as altered homeostasis or disrupted metabolic pathways may affect neuronal function and contribute to neurodevelopmental phenotypes [34–37]. However, comprehensive frameworks that integrate multi-tissue, multi-omics data collected within the same cohort, including less-explored domains such as metallomics, remain rare.

Twin study designs provide a uniquely powerful framework for isolating condition-associated biological variation from shared genetic and environmental confounding. By controlling for age and most of the early-life environment and, in monozygotic (MZ) pairs, for essentially the entire inherited genome, within-pair comparisons substantially reduce background sources of heterogeneity. Consequently, differences observed in condition-discordant twin pairs can be more confidently attributed to condition-related processes rather than shared genetic or environmental influences [38, 39].

In this exploratory study, we applied an integrative multi-omics approach with a twin design to identify molecular signatures and convergent biological pathways associated with NDCs across multiple layers of biological organization. We leveraged data from the Roots of Autism and ADHD Twin Study in Sweden (RATSS) cohort [40, 41], which includes twin pairs concordant and discordant for NDCs, as well as typically developing twin pairs. By integrating complementary single-omics datasets generated from the RATSS cohort, including serum and CSF proteomics [21], fecal and urine metabolomics [24], blood metallomics, and whole-genome sequencing (WGS) data from a subset of 14 MZ twin pairs, we systematically examined cross-omics relationships and biological convergence across molecular layers. The most important molecules were examined using across- and within-twin-pair analyses to highlight genetic and environmental contributions.

## 2 Methods and materials

### 2.1 Study participants

The sample was drawn from RATSS [40, 41], a deeply phenotyped cohort of twin pairs with various NDC diagnoses and neurotypically developing twins (Supplementary Fig. S1, Table S1). Detailed recruitment, clinical assessment, and behavioral measures are described in Supplementary Methods S1 and in previous publications [40, 41]. Briefly, twins were recruited from a population-based twin cohort and other sources, targeting autism and ADHD discordant pairs. All twins underwent a comprehensive behavioral assessment at a clinical research center (Supplementary Methods S2), including age-appropriate parent-rated Child/Adult Behavior Checklist (CBCL/ABCL), self-rated Youth/Adult Behavior Checklist (YSR/ASR), Adaptive Behavior Assessment System–Second Edition (ABAS-II), Social Responsiveness Scale 2nd Edition (SRS-2), Wechsler Intelligence Scales (WISC-IV/WAIS-IV) or the Leiter scales. NDCs and psychiatric disorders were assessed by expert-clinician consensus according to DSM-5 and ICD-10 criteria [1]. Zygosity was determined by genotyping or a 48-single-nucleotide polymorphism (SNP) panel [42, 43]. A subset of 14 MZ twin pairs provided WGS data as part of the EU-AIMS initiative [44]. NDC discordance was defined by one twin meeting DSM-5/ICD-10 criteria for any NDC (*e.g.,* autism, ADHD, Intellectual Disability [ID], Tic Disorder, and Specific Learning Disorder) while the co-twin did not. Individuals with one or more NDC diagnoses were assigned to the anyNDC group, and those without any to the noNDC group.

The study was conducted in accordance with the Declaration of Helsinki and was approved by the Swedish Ethical Review Authority (2016/145-231). Informed consent was obtained from all participants and/or their caregivers.

### 2.2 Biological sample processing and measurement

Serum and CSF proteomic data were available for 126 and 86 individuals, respectively. Proteins were measured using multiplex immunoassay panels (Olink Bioscience, Sweden; RRID:SCR_003899), including Inflammation I, CVD (cardiovascular), and Oncology I, covering 203 proteins. More details are provided in Supplementary Methods S3.1 and Smedler et al. [21]. Urine and fecal metabolomic data were available for 112 and 189 individuals, respectively. Urine was profiled by untargeted ^1^H nuclear magnetic resonance (NMR) spectroscopy, and fecal samples by untargeted liquid chromatography-mass spectrometry (Metabolon). More details are provided in Supplementary Methods S3.2, S3.3 and Swann et al. [24]. Metallomic profiling was performed on blood samples from 220 individuals, quantifying essential [calcium (Ca), copper (Cu), iron (Fe), magnesium (Mg), manganese (Mn), molybdenum (Mo), selenium (Se), and zinc (Zn)] and toxic metals [arsenic (As), cadmium (Cd), cobalt (Co), mercury (Hg), and lead (Pb)] by inductively coupled plasma-mass spectrometry (ICP-MS). More details are provided in Supplementary Methods S3.4.

### 2.3 Integrative omics analysis

We applied an integrative multi-omics strategy using the DIABLO (Data Integration Analysis for Biomarker Discovery using Latent Variable Approaches for Omics Studies) framework implemented in the R package mixOmics (version 6.22.0) [45]. DIABLO applied multiblock sparse Partial Least Squares Discriminant Analysis (sPLS-DA) to identify molecules correlated across omics layers while jointly contributing to discriminating anyNDC from noNDC. Through LASSO penalization, a sparse subset of molecules was selected, producing latent components (variates) for each omics block. Each individual received a variate score for each component per block, reflecting their position along the axis of maximal cross-omics covariation associated with NDC status. The average variate 1 and variate 2 scores across omics blocks indicate cross-omics NDC-associated separation at the individual level. We first applied single-omics sPLS-DA models to optimize variable selection parameters for each dataset. To maximize sample coverage across diverse omics combinations, we designed three complementary multiomics models: *MultiOmic-SCFB* (serum-proteomics, CSF-proteomics, fecal-metabolomics, and blood-metallomics datasets) with 72 samples (37 anyNDC, 35 noNDC); *MultiOmic-SFB* (serum-proteomics, fecal-metabolomics, and blood-metallomics datasets) with 109 samples (56 anyNDC, 53 noNDC); *MultiOmic-UFB* (urine-metabolomics, fecal-metabolomics, and blood-metallomics datasets) with 89 samples (29 anyNDC, 60 noNDC) (Fig. 1, Supplementary Fig. S2). Detailed data processing and model construction are described in Supplementary Methods S4 and Fig. S3.

**Fig. 1.**
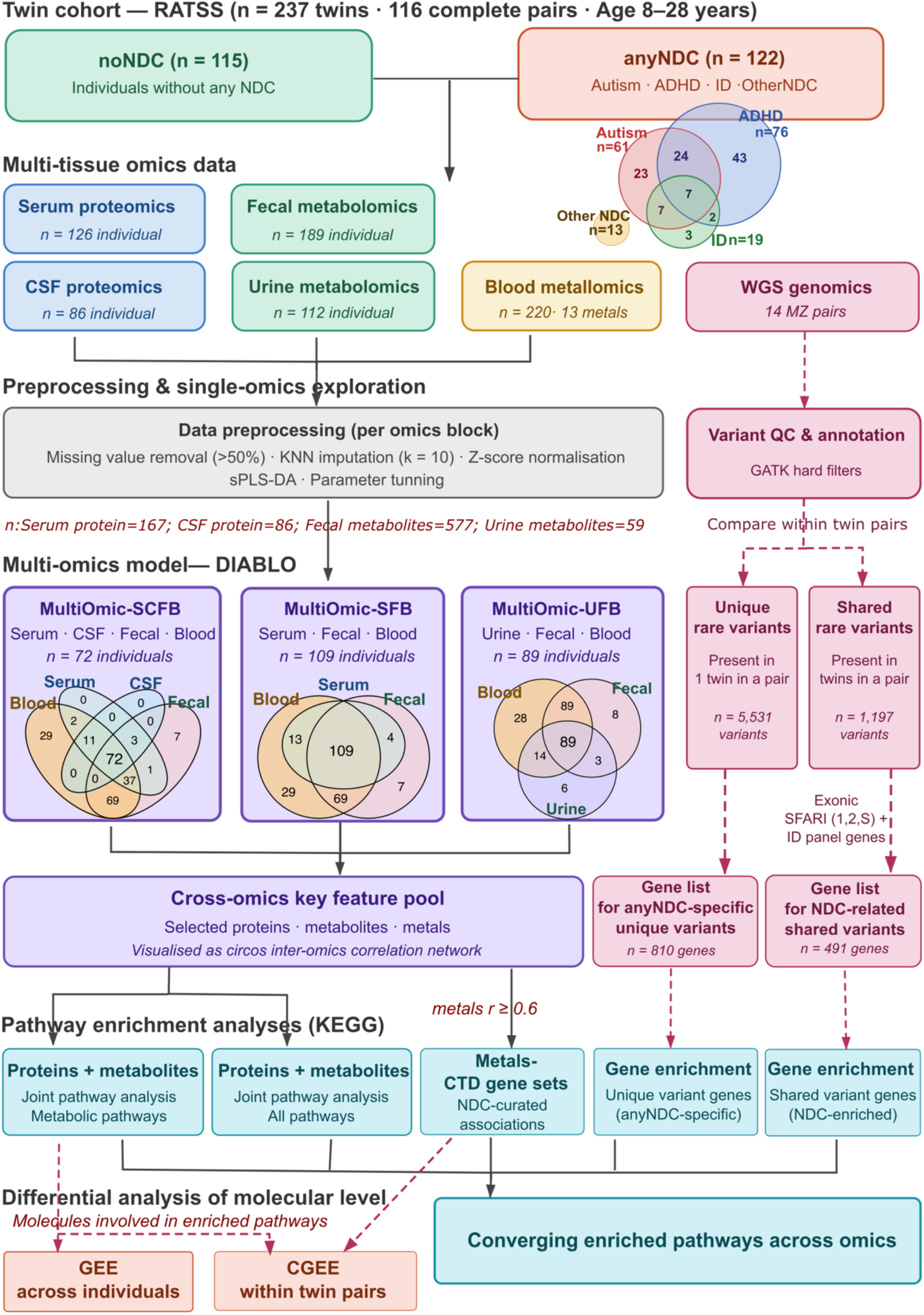
Schematic overview of study design and integrative multi-omics analytical workflow. RATSS twin cohort was stratified into anyNDC (NDC diagnostic subgroups are shown in the Venn diagram) and noNDC groups. Multi-tissue omics data were collected across serum proteomics, CSF proteomics, fecal metabolomics, urine metabolomics, and blood metallomics, with WGS generated for a subset of 14 MZ twin pairs. Each omics block (except for WGS data) underwent independent preprocessing (missing value removal, KNN imputation, z-score normalization) followed by single-omics exploration and sPLS-DA parameter tuning. Three multi-omics DIABLO models (*MultiOmic-SCFB, MultiOmic-SFB, MultiOmic-UFB*), integrating different omics combinations (sample overlap across omics datasets is shown in Venn diagrams), were applied as an exploratory framework to identify a cross-omics key feature pool of molecules correlated across layers and jointly discriminating anyNDC from noNDC. Pathway enrichment analyses (KEGG) explored the converging biological relevance of key molecules, performed jointly for proteins and metabolites and separately for metals via CTD gene sets. In parallel, WGS-derived gene lists from NDC-specific unique and shared rare variants underwent gene enrichment, converging with omics-derived pathways to provide a complementary genomic perspective. Twin-modeling-based differential analyses (GEE across-individual, CGEE within-pair) further accounted for covariates and evaluated whether each molecule was independently associated with NDC status.

Associations between average variate scores and potential confounders and clinical measures were assessed using Generalized Estimating Equation (GEE) models with the R package drgee (version 1.1.10)[46] to account for the family linkage.

### 2.4 Pathway enrichment analysis for proteins and metabolites

We analyzed the enrichment of proteins and metabolites selected from the multi-omics models across Kyoto Encyclopedia of Genes and Genomes (KEGG) pathways [47] using MetaboAnalyst 6.0 web tool (accessed 2025-08-12) [48], via the Joint Pathway Analysis module. Using the default background, we evaluated both metabolic-pathway and all-pathway libraries. Enrichment significance was determined by a hypergeometric test, with degree centrality as a topology measure to weight individual molecular importance. We applied different integration methods for the background: for metabolic pathway analysis, the “*Overall Combine p values”* method, which weights the proportion of each omics; for all pathway analysis, the tighter “*Combine Query”* method, which pools the genes/proteins and metabolites into a single query. The false discovery rate (FDR) method was used to correct for multiple comparisons [49]. Furthermore, we applied Gene-Metabolite Interaction Network analysis in the Network Analysis module (default parameters) to examine interactions between selected proteins and metabolites.

### 2.5 Metallome and NDC-related pathways

We selected seven metals that showed correlation coefficients > 0.6 with features from other omics layers in multi-omics models, including the essential metals Ca, Mg, Fe, Zn, and Mn, as well as the toxic metals Co and Cd. In the Comparative Toxicogenomics Database (CTD, accessed 2025-01-09) [50], we searched the condition keyword “neurodevelopmental disorder” and retrieved curated gene sets for each metal with a documented association with NDCs. Ca was excluded because its association with NDCs in CTD lacked curated evidence. We then conducted KEGG pathway enrichment analysis on the combined gene lists using the R package clusterProfiler (version 4.6.2) [51], with *p*-values adjusted by the FDR method.

### 2.6 Differential analysis of molecular levels

To assess associations between molecular levels and NDC diagnosis, we focused on molecules mapped to significantly enriched metabolic pathways and all 13 metals. The lysine degradation pathway was additionally included as it reached nominal significance in the protein-metabolite pathway analysis and FDR-significance in the metal-related pathway analysis. We first assessed across-individual effects using standard GEE models adjusted for age, sex, BMI, medication (yes/no), and other psychiatric diagnosis (yes/no), with standard errors clustered by twin pair. To control for unmeasured family-constant confounders (*e.g.,* parental psychiatric history, growth environment, and shared genetic factors of twin pairs), we then applied conditional GEE (CGEE) models to estimate within-pair molecular effects. CGEE models were adjusted for cluster-varying covariates (BMI, medication, and other psychiatric diagnoses). Analyses were performed across all twin pairs, MZ twin pairs, and NDC-discordant MZ pairs to isolate the effects of non-shared exposures.

### 2.7 WGS variant analysis and pathway enrichment

We analyzed WGS data from 14 MZ twin pairs (3 NDC-discordant, 6 NDC-concordant, and 5 noNDC-concordant pairs). Zygosity was confirmed by kinship coefficient analysis (Kinship >0.345). We prioritized two variant types to identify potential genetic drivers of discordance and shared liability. First, we identified 5,531 unique rare variants (allele frequency [AF] <0.001 in gnomAD/SweFreq) present in only one twin of a pair and then absent from all other individuals. Second, we identified 1,197 shared rare variants, defined as exonic variants (AF <0.01) shared within a twin pair. Shared variants were restricted to NDC-related genes, including SFARI genes (score 1, 2, or syndromic) and the Intellectual Disability gene panel [52, 53]. Functional impact of genes harboring NDC-specific unique or shared rare variants was assessed by KEGG pathway enrichment analysis using the R package clusterProfiler (version 4.6.2), with FDR-adjusted p-values. Detailed QC and filtering parameters are provided in Supplementary Methods S5.

## 3 Results

Our study included 237 twins from the RATSS cohort with available omics data, including 116 complete twin pairs and five unpaired individuals whose co-twins lacked omics data. Among them, 122 twins (51.5%) were diagnosed with at least one NDC (Fig. 1), and the study population ranged in age from 8 to 28 years (mean 14.7, SD 3.7). When stratified by NDC diagnoses, two groups differed in age, BMI, medication, and psychiatric diagnosis status (Table 1). Within each omics dataset, the proportion of anyNDC and noNDC groups was balanced (anyNDC 49.2-52.4%), except for the urine dataset (anyNDC 33.0%). Detailed demographic characteristics for each omics dataset and individual are presented in Supplementary Fig. S1 and Tables S1-S2. The overall analytical workflow is illustrated in Fig. 1.

**Table 1.** Study population characteristics and descriptive statistics. Difference between noNDC and anyNDC were evaluated by t-test (Age and BMI) and chi-square test (Sex, Any medication and Psychiatric diagnosis).

| **Whole dataset n=237** |  |  |  |
|---|---|---|---|
| NDC diagnosis | noNDC | anyNDC | Difference *p*-value |
| Number (%) | 115 (48.5%) | 122 (51.5%) | / |
| Age (years), range; mean (SD) | 8-25; 15.3 (3.3) | 8-28; 14.1 (4.0) | 0.018 |
| Sex, Female N (%) | 58 (50.4%) | 46 (37.7%) | 0.065 |
| Concordant twin pairs, N (MZ:DZ) | 30:6 | 25:15 | / |
| Discordant NDC twin pairs, N (MZ:DZ) | 23:17 |  | / |
| BMI, mean (SD) | 20.8 (3.1) | 20.2 (3.9) | 0.034 |
| Any medication, N (%) | 18 (15.7%) | 65 (53.3%) | <0.001 |
| Psychiatric diagnosis, N (%) | 11 (9.6%) | 36 (29.5%) | <0.001 |

### 3.1 Identification of multi-omics profile

To incorporate a broader range of omics layers while maximizing the number of samples with complete data, we first integrated the serum-proteomics, CSF-proteomics, fecal-metabolomics, and blood-metallomics datasets (72 individuals: 37 anyNDC, 35 noNDC; Fig. 1, Supplementary Fig. S2). Details of the single-omics investigations are described in Supplementary Results S1. The resulting model, *MultiOmic-SCFB,* identified 106 key molecular features across two latent components. Detailed results for parameter tuning and model optimization are provided in Supplementary Table S3. The model revealed the highest correlation between the first latent components of the serum and CSF datasets (r = 0.54; Supplementary Figs. S4A-B), likely reflecting the shared Olink panel across both tissues, while the modest correlation indicates partially distinct signals highlighting the complementary value of multi-tissue sampling. Projecting twins onto each omics block’s latent components (variate 1 and variate 2 scores) revealed that serum-proteomics provided the strongest single-layer separation of NDC status (Fig. 2A). The average variate scores across all omics blocks, representing the integrated cross-omics signal, showed clearer group separation than any single-omics projection (Fig. 2B). The intermediate region between two groups was enriched for NDC-discordant twin pairs, reflecting molecular similarity potentially shaped by shared environmental or genetic factors within pairs. However, no associations between averaged variate 1 or variate 2 scores and potential confounders (age, sex, BMI, medication use, and other psychiatric diagnoses) survived FDR correction (Supplementary Table S4), indicating that the multi-omics signatures are not driven by these covariates. A global map of the key features and their inter-omics correlations is visualized in a circos plot (Fig. 2C, Supplementary Figs. S4C-D), providing a holistic view of the interconnected molecular architecture of NDCs. More details about multi-omics model results are described in Supplementary Results S2.

**Fig. 2.**
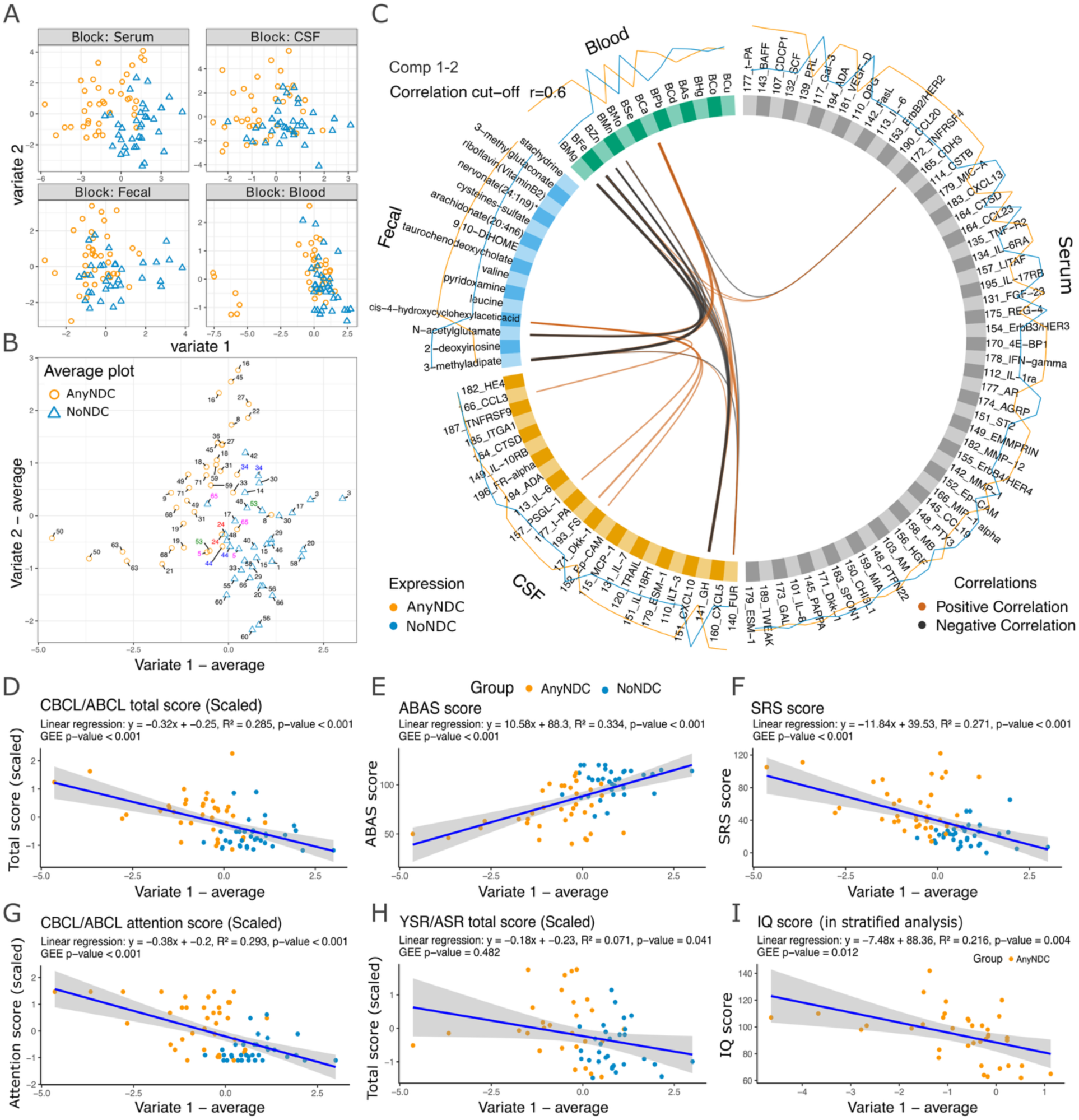
Latent multi-omics profile and clinical correlates of the *MultiOmic-SCFB* model. The model integrated serum-proteomics, CSF-proteomics, fecal-metabolomics, and blood-metallomics. **A**. Sample projection plots for each omics block, displaying individual samples in the space defined by the first two components (variate 1 and variate 2) derived from the model. Each panel corresponds to a different omics dataset and shows the discriminative capacity of each omics layer, along with the alignment between anyNDC and noNDC groups (anyNDC in orange, noNDC in blue). **B**. Averaged component plot across all omics blocks, combining scores from components 1 (variate 1) and 2 (variate 2) into a single integrative sample-space representation. NDC-discordant twin pairs that are projected in close proximity are labeled and color-highlighted to emphasize within-pair similarity. **C**. Circos plot showing pairwise correlations between molecules across different omics datasets, based on merged component loadings. Edges represent cross-omics correlations with an absolute correlation coefficient ≥ 0.6; positive correlations are shown in orange, and negative correlations in grey. The outer rings around the circumference of the plot represent molecule abundance or expression level stratified by NDC status (anyNDC in orange, noNDC in blue). **D-I**. Association between clinical measures and latent multi-omics profile, using the average variate score on component 1 shown in panel B as the x-axis. Two *p*-values are reported for each association: one from linear regression and one from GEE models. GEE models were adjusted for sex, age, BMI, medication status, and other psychiatric diagnosis status, with twin ID included as the clustering variable. Dots are colored by NDC diagnosis status. Plots show associations with: **D**. CBCL/ABCL total score (Child/Adult Behavior Checklist); **E**. ABAS-II score (Adaptive Behavior Assessment System-II); **F**. SRS-2 score (Social Responsiveness Scale-2); **G**. CBCL/ABCL attention score. **H**. YSR/ASR total score (Youth/Adult Self Report); **I**. IQ score (WISC-IV, WAIS-IV, or Leiter) within anyNDC group in stratified analysis.

We further tested whether the omics profile captured in the *MultiOmic-SCFB* model reflected variation in dimensional NDC traits and more general psychiatric symptoms. In GEE models adjusting for sex, age, BMI, medication status and psychiatric diagnosis status, significant but modest correlations (GEE-p<0.001; Fig. 2D-G) were observed between the averaged variate 1 scores and autistic traits (SRS), ADHD traits (CBCL/ABCL attention subscale), adaptive functioning (ABAS), and parent-reported general psychiatric problems (total CBCL/ABCL scores). No association was found with self-reported behavior problems (total YSR/ASR score; GEE-*p*=0.482; Fig. 2H). Stratified analyses for anyNDC and noNDC groups showed no significant associations for CBCL/ABCL, ABAS, and SRS scores, while a significant negative association between IQ scores and averaged variate 1 scores was observed in the anyNDC group (GEE-*p*=0.012; Fig. 2I).

To maximize sample coverage given incomplete omics overlap and improve statistical power, we then integrated serum-proteomics, fecal-metabolomics, and blood-metallomics datasets to build the *MultiOmic-SFB* model (n=109; Supplementary Fig. S5). For urine-metabolomics, we maximized sample retention by integrating it with fecal-metabolomics and blood-metallomics, producing the *MultiOmic-UFB* model (n=89; Supplementary Fig. S6). More details about *MultiOmic-SFB* and *MultiOmic-UFB* models are described in Supplementary Results S2.2 and S2.3. Overall, integrating multiple omics layers tended to improve discrimination between anyNDC and noNDC beyond that achieved by fewer-omics or single-omics datasets (Fig. 2B). We therefore selected molecules identified as key features across the three models to identify enriched biological pathways contributing to the anyNDC and noNDC separation.

### 3.2 Enrichment of key metabolites and proteins

Next, we conducted joint pathway analysis on the 79 metabolites and 63 proteins identified as key features in multi-omics models. Considering all pathways (both metabolic and gene-only), 21 were significantly enriched (FDR-adjusted *p*<0.05; Fig. 3A, Supplementary Table S5). Of these, only two were enriched by both protein and metabolite hits (Central carbon metabolism in cancer and HIF-1 signaling pathway). Notably, the ErbB signaling and EGFR tyrosine kinase inhibitor resistance pathways exhibited the highest topological impact scores of 1.42 and 1.30 (Fig. 3B). Restricting to the metabolic-pathway pool, the “*Combine P-Values*” method revealed seven significantly enriched pathways (Fig. 3C; Supplementary Table S6). Among these, Riboflavin metabolism had the highest topological impact score (0.88; Fig. 3D). Only Purine metabolism was enriched by both protein (i.e., adenosine deaminase [ADA]) and metabolites.

**Fig. 3.**
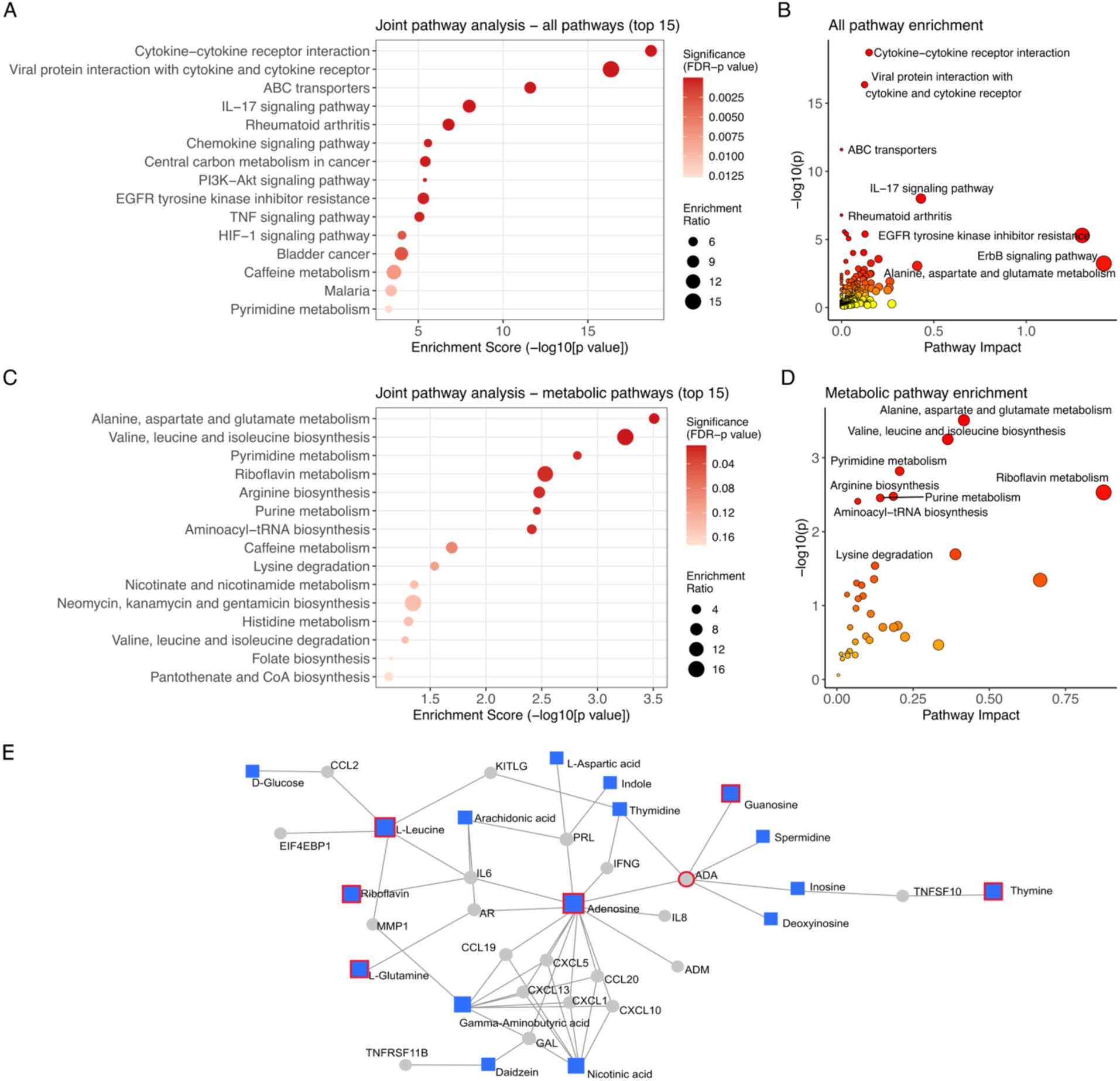
Joint pathway analysis of key metabolites and proteins identified in the MultiOmics models. **A**. Enrichment dot plot of the top 15 enriched pathways, including both metabolic and gene-only pathways. *P*-values were calculated using the *Combine Query* method in *MetaboAnalyst 6.0*. **B**. Scatterplot of pathway analysis results in (**A**). Each circle represents a matched pathway, with color indicating *p*-value and size reflecting pathway impact. **C**. Enrichment dot plot of the top 15 enriched metabolic pathways. *P*-values were derived using the *Combine p-values* method in *Metaboanalyst 6.0*. **D**. Scatterplot of metabolic pathway results in (**C**). Each circle represents a matched pathway, with color indicating *p*-value and size reflecting pathway impact. The full results of joint pathway analysis are listed in Tables S5-S6. **E**. Gene–metabolite interaction network derived from key molecules. The network illustrates functional associations between genes of key proteins and metabolites identified in the MultiOmics models. Interactions were extracted from the STITCH database, filtered to retain only high-confidence associations. Metabolites are shown as squares, and genes are shown as circles. Nodes outlined in red indicate molecules significantly enriched in the purine metabolism pathway.

To further investigate the functional interplay between key proteins and metabolites, we performed Gene–Metabolite Interaction Network analysis, which identified a core functional module centered on Purine metabolism (Fig. 3E). Interestingly, this metabolic hub connected with a cluster of chemokine genes, suggesting a coordinated metabolic-inflammatory axis involving adenosine and nicotinic acid.

### 3.3 Differential analysis of molecules within twin pairs

We then investigated associations between molecular levels and NDC diagnosis utilizing the twin design further for the molecules mapped to seven enriched metabolic pathways (FDR-adjusted *p*<0.05; Fig. 3C) and the lysine degradation pathway, including one protein (ADA; Supplementary Figs. S7-S8), 21 fecal metabolites (Supplementary Fig. S9), and two urine metabolites (Supplementary Fig. S10). Serum ADA levels were consistently and significantly elevated in individuals with NDCs in the whole sample (GEE *p*=0.0002; CGEE *p*=0.015), the MZ subset (GEE *p*=0.0013; CGEE *p*=0.0048), and notably the discordant MZ twins (GEE *p*=0.004; CGEE *p*=0.0009) (Table 2). However, ADA measured in CSF showed no significant difference (Supplementary Table S7).

**Table 2.** Associations between molecular levels and NDC diagnosis across the entire sample and in MZ and discordant MZ subsamples. GEE model adjusting for BMI, medication status, presence of other psychiatric diagnosis, sex, and age; CGEE model adjusting for BMI, medication status, and presence of other psychiatric diagnosis. Abbreviations: GEE, Generalized estimating equation; CGEE, Conditional generalized estimating equation; FDR, False discovery rate; FMN, Flavin mononucleotide; NAA, N-acetylaspartate; CSF, Cerebrospinal fluid; ADA, Adenosine Deaminase; BMn, Blood Manganese; BSe, Blood Selenium; BCu, Blood Copper. Only molecular associations with nominal significance are shown here. The results for all molecules are shown in Supplementary Tables S7-S10.

|  |  | **GEE: across-individual** |  |  |  | **CGEE: within-pair** |  |  |  |
|---|---|---|---|---|---|---|---|---|---|
| Subsamples | Molecules | Estimate | *p* value | FDR-adj *p* value | Sample size | Estimate | *p* value | FDR-adj *p* value | Sample size |
| **Protein (Serum)** |  |  |  |  |  |  |  |  |  |
| All individuals | ADA | 0.656 | **<0.001** | / | 109 | 0.366 | **0.015** | / | 94 |
| All MZ twins | ADA | 0.596 | **0.001** | / | 94 | 0.446 | **0.005** | / | 86 |
| Discordant MZ | ADA | 0.493 | **0.004** | / | 26 | 0.537 | **0.001** | / | 24 |
| **Metabolite (Fecal)** |  |  |  |  |  |  |  |  |  |
| All individuals | 2'-deoxyinosine | -0.448 | **0.014** | 0.145 | 189 | -0.240 | 0.299 | 0.629 | 180 |
|  | FMN | -0.561 | **0.002** | **0.036** |  | 0.017 | 0.940 | 0.956 |  |
|  | deoxycarnitine | -0.190 | 0.320 | 0.448 |  | -0.854 | **0.022** | 0.462 |  |
| All MZ twins | 2'-deoxyinosine | -0.559 | **0.007** | 0.063 | 134 | -0.399 | 0.116 | 0.444 | 128 |
|  | FMN | -0.917 | **<0.001** | **<0.001** |  | -0.176 | 0.536 | 0.920 |  |
|  | 2-aminoadipate | 0.401 | **0.009** | 0.063 |  | 0.450 | 0.071 | 0.444 |  |
|  | glutamine | 0.177 | 0.340 | 0.517 |  | -0.499 | **0.048** | 0.444 |  |
| Discordant MZ | FMN | -0.571 | **0.036** | 0.250 | 42 | 0.190 | 0.519 | 0.748 | 40 |
|  | thymidine | -0.179 | 0.426 | 0.717 |  | -0.407 | **0.002** | **0.029** |  |
|  | glutamine | -0.542 | **0.010** | 0.202 |  | -0.706 | **0.003** | **0.029** |  |
|  | aspartate | -0.486 | 0.092 | 0.484 |  | -0.594 | **0.015** | 0.078 |  |
|  | NAA | -0.368 | **0.035** | 0.250 |  | -0.102 | 0.355 | 0.715 |  |
|  | 2'-deoxyuridine | -0.195 | 0.444 | 0.717 |  | -0.489 | **0.006** | **0.039** |  |
| **Metal (Blood)** |  |  |  |  |  |  |  |  |  |
| All individuals | BMn | 0.456 | **0.005** | 0.067 | 220 | -0.175 | 0.226 | 0.759 | 210 |
|  | BSe | 0.372 | **0.028** | 0.182 |  | -0.117 | 0.236 | 0.759 |  |
| All MZ twins | BMn | 0.716 | **<0.001** | **0.004** | 149 | 0.053 | 0.622 | 0.976 | 146 |
|  | BSe | 0.430 | **0.028** | 0.181 |  | 0.007 | 0.953 | 0.976 |  |
| Discordant MZ | BCu | 0.040 | 0.825 | 0.961 | 44 | 0.367 | **0.003** | **0.036** | 42 |

For fecal metabolites, across-individual GEE models in whole samples (n=189) and MZ twin subset (n=134) identified a significant negative association between flavinmononucleotide (FMN) and NDC diagnosis (FDR-adjusted *p*=0.036 and 4.8 × 10⁻⁵; Supplementary Table S8). However, this association was absent in the within-pair CGEE models (90 and 64 clusters for whole and MZ subsets, respectively), suggesting the FMN signal may be influenced by familial background. Interestingly, for NDC-discordant MZ subsample, within-pair analysis indicated a distinct metabolic signature in which twins with NDC had significantly lower levels of thymidine, glutamine, and 2’-deoxyuridine than their co-twins without NDC (FDR-adjusted *p*=0.029, 0.029, and 0.039, respectively). Results for all fecal molecules are shown in Supplementary Table S8. For urine metabolites, no significant differences were found (Supplementary Table S9).

We also evaluated alterations in 13 blood metals in relation to NDC diagnosis (Supplementary Fig. S11, Table S10). Mn showed a positive association with NDC diagnosis in the MZ subset (FDR-adjusted *p*=0.0039), while Cu showed a significant elevation specifically in NDC-affected twins in the discordant MZ within-pair model (FDR-adjusted *p*=0.036; Table 2). Additional nominally significant results are described in Supplementary Results S3.

Exploratory analyses stratified by ADHD and autism showed heterogeneous modification patterns, reflecting the molecular complexity of individual NDC diagnoses. However, these findings should be interpreted cautiously given the modest and varied sample sizes available across omics datasets and models, and individuals with comorbid diagnoses contributed to subgroup analyses (Supplementary Results S3, Table S11).

### 3.4 Metal exposure and NDC-related pathways

To investigate the functional relevance of the metallomics profile, we focused on metals strongly correlated with other omics layers (Mg, Fe, Zn, Mn, Co, Ca, and Cd; correlation coefficients >0.6) and performed KEGG pathway enrichment using genes associated with both NDCs and at least one metal. Using a set of 306 metal-NDC-associated genes with curated evidence in CTD (Ca-NDC-genes excluded, lacking curated evidence), 100 pathways were significantly enriched (Supplementary Fig. S12, Table S12). More details are described in Supplementary Results S4. Notably, *ADA* was among the genes associated with autism-related Cd and Zn, and autistic disorder-related Zn.

### 3.5 Pathway enrichment for genes harboring unique and shared rare variants in 14 MZ twin pairs

KEGG pathway enrichment of genes harboring NDC-specific unique and shared rare variants revealed convergence with pathways identified in other omics layers (Fig. 4). For 810 genes harboring NDC-exclusive rare unique variants, 15 pathways were significantly enriched (FDR-adjusted *p*<0.05; Supplementary Fig. S13A, Table S13). Among these, the MAPK signaling pathway (FDR-adjusted *p*=0.032) was consistently implicated across enrichment analyses of both proteo-metabolomic and metallomic layers. The chemokine signaling pathway (FDR-adjusted *p*=0.039) was also enriched in the joint pathway analysis of proteins and metabolites. Additionally, several previously significant pathways reappeared at a nominal significance level here (*e.g*., purine metabolism [*p*=0.028] and PI3K-Akt signaling pathway [*p*=0.035]). Analysis of 491 genes harboring only rare exonic shared variants in concordant and discordant NDC twins identified nine significantly enriched pathways (Supplementary Fig. S13B, Table S14). The lysine degradation pathway (FDR-adjusted *p*=0.013) overlapped with previous findings for both key proteins-metabolites and metals. Further narrowing the analysis to genes with variants shared exclusively in concordant NDC twins and absent from all noNDC individuals, the calcium signaling pathway was the sole significantly enriched pathway (FDR-adjusted *p*=0.0097). More results about rare variants identified in WGS data are described in Supplementary Results S5.

**Fig. 4.**
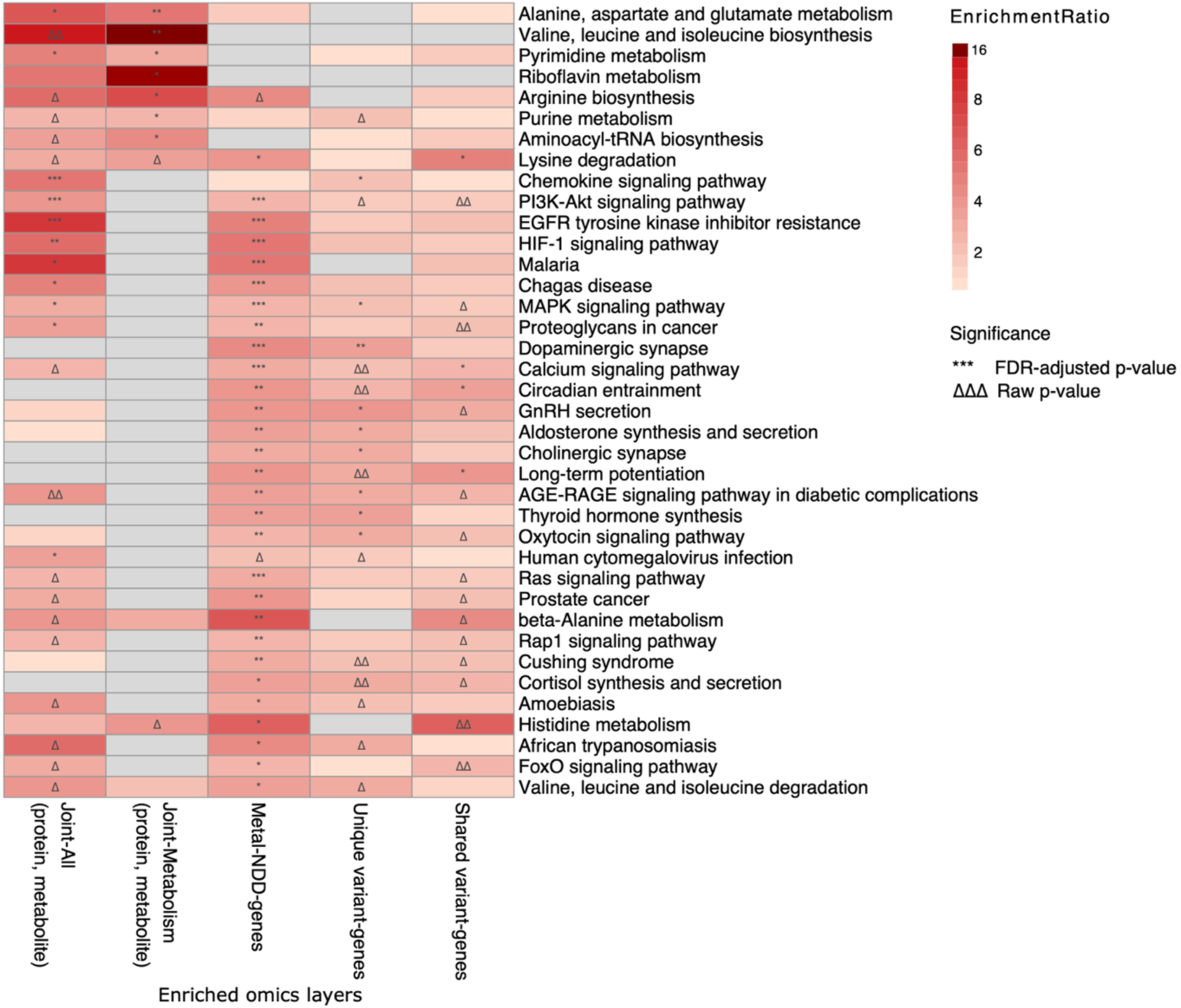
The enriched pathways overlapped in different omics layers. Pathways are shown if they reached significance in at least two omics layers with FDR-adjusted *p*-value*<*0.05, or if they showed one FDR-adjusted significance together with at least two nominal significances (Raw *p*<0.05) in different omics layers. Metabolic pathways identified as significantly enriched in the joint metabolic pathway analysis are also shown. Colors indicate enrichment ratios, with darker shading representing higher ratios. Significances were labeled as FDR-adjusted *p*-value: ***<0.001, **<0.01, *<0.05; Raw *p-*value: ΔΔΔ<0.001, ΔΔ< 0.01, Δ<0.05. The Raw *p*-values were labeled only when the FDR-adjusted *p*-value≥0.05.

## 4 Discussion

In this study, we applied an integrative multi-omics strategy within a twin cohort, combining data-driven multiblock sPLS-DA models with knowledge-based pathway enrichment to investigate molecular signatures and biological pathways associated with NDC. Convergent signals implicated several metabolic and signaling pathways across omics layers. Twin-model-based analyses provided additional context for the molecules prioritized by the integrative framework, indicating that their associations with NDC arose from a combination of shared genetic/familial influences and non-shared individual-specific factors across different molecular features. ADA protein and blood copper showed independent associations with NDC status, while thymidine, glutamine, and 2’-deoxyuridine, involved in pyrimidine metabolism, were consistently decreased in the NDC-discordant MZ subset. Flavin mononucleotide and blood manganese were also associated with NDC, though their associations were modulated by shared genetic or familial environmental influences.

A key value of the integrative approach is that some molecules’ relevance to NDC becomes apparent only through their correlation with other omics layers rather than from single-omics testing alone [54]. In contrast to approaches that enrich each layer separately and overlap the results post hoc [29, 55], our convergent pathway findings reflect coordinated cross-layer relationships captured through joint modeling. This value also stems from the distinct position each omics layer occupies along the genotype-to-phenotype path and its tissue origin [56]. Pathways converge from molecules spanning these compartments, and modeling cross-omics correlation enables more complete identification of coordinated dysregulation. WGS, although available only in 14 MZ twin pairs, complements these insights by capturing rare genetic variation at both somatic and germline levels, offering a genetic-layer perspective on processes also highlighted at the physiological level. Molecules highlighted by both the integrative models and individual-level differential analysis are considered doubly supported and given particular weight in our discussion.

Several pathways were consistently and significantly implicated across different omics layers, highlighting their potential involvement in NDC physiology. Among metabolic pathways, lysine degradation emerged as the most consistently enriched process. Numerous studies have associated lysine degradation with mitochondrial energy metabolism and redox homeostasis; however, direct evidence linking it to NDC remains limited and mainly focused on autism. A recent systematic review reported that autism-associated microRNAs are significantly involved in lysine degradation [57]. Emerging evidence also points to microbiome-mediated mechanisms. Zou et al. found that altered gut microbiota in autistic children was associated with reduced lysine degradation [58], while Hicks et al. observed upregulated microbial RNA enriched in this pathway in the autism oropharynx [59]. Altered lysine metabolism may also affect microbial degradation products such as 5-aminovalerate, which has been implicated in autism-related behaviors [60].

We further observed that one key metabolite in the lysine pathway, 2-aminoadipate, was nominally elevated in individuals with NDCs in the all MZ twin subsamples. Consistent with this, a recent clinical study found higher levels of α-aminoadipic acid in autistic children under five years compared with non-autistic children [61]. The accumulation of 2-aminoadipate is of particular interest because it can act as a neurotoxin by inhibiting the synthesis of kynurenic acid, a neuroprotective metabolite. Such accumulation may occur when aminotransferases that normally metabolize 2-aminoadipate are impaired, a process dependent on the cofactor pyridoxal 5’-phosphate (PLP, vitamin B6). Reduced or highly variable PLP levels have been reported in some autistic children compared with the neurotypical group [60]. Also, early postnatal pyridoxine supplementation has been shown to improve social and repetitive behaviors in the BTBR mouse model [62]. Together, these findings highlight a potential microbial-host metabolic axis linking lysine degradation, vitamin B6-dependent enzymatic activity, and neurobehavioral regulation in NDCs.

The purine metabolism pathway also emerged as a key metabolic focus in our study, enriched across multiple layers. This pathway is critically involved in neurodevelopment, particularly through its regulation of neuroimmune interactions and oxidative stress responses [63]. One of its core enzymes, ADA, regulates extracellular adenosine levels and mediates neuroprotective and anti-inflammatory functions [64]. The *ADA* gene has been listed in the SFARI Gene database as a strong candidate for autism susceptibility [52], with previous genetic studies linking specific allelic variants to ADA deficiency and increased autism likelihood [65, 66]. More recently, transcriptomic evidence has suggested an upregulation of *ADA* expression in autism [67]. Our finding of elevated serum ADA in NDC group parallels this molecular trend and remained robust even after controlling for family-constant confounding. Notably, CSF ADA levels did not differ between groups, reflecting region-specific regulation of ADA in peripheral metabolic disturbances. Together, we extend prior genetic- and transcriptomic-level observations to the protein level and broaden ADA’s relevance across NDC diagnoses, highlighting ADA as a potential peripheral biomarker of purine metabolic imbalance in NDC.

The purine metabolism pathway also appears to interact with broader neuroimmune signaling networks implicated in NDCs. In our metabolite–protein interaction network, adenosine, the central node of purine metabolism, exhibited close topological proximity to several members of the chemokine family. Consistent with this, the chemokine signaling pathway was also significantly enriched in our protein-metabolite joint pathway analysis and in the gene set harboring rare unique variants found in NDCs. Chemokines are well-established mediators of neuroimmune signaling, and their dysregulation during early development has been linked to altered neurodevelopmental trajectories in autism [68]. These findings suggest a potential mechanistic convergence between purinergic signaling and immune modulation in the context of NDCs.

Our findings also highlighted several other key cell-signaling pathways, most notably the PI3K-Akt and MAPK signaling. These pathways showed consistent associations across multiple data layers, involving proteins, metals, and genes carrying NDC-specific rare unique variants. Both pathways are well-documented regulators of neurodevelopmental processes, including neuronal survival, synaptic plasticity, and neuroimmune signaling [69, 70]. Genetic and functional studies have linked their dysregulation to a range of NDCs, with NDC-associated risk genes converging on these signaling cascades [11, 70–72], and perturbation correlating with symptom severity in idiopathic autism and cognitive impairments in intellectual disability [73, 74]. Among the key proteins we identified, EIF4EBP1 serves as a molecular marker of mTOR signaling [72], suggesting that downstream signals closely connected to PI3K-Akt, such as the mTOR axis, may also be implicated. Our findings add multi-layered evidence supporting PI3K-Akt/MAPK involvement in NDCs, though the underlying mechanisms warrant further investigation.

Among metabolites mapped to significantly enriched metabolic pathways, only thymidine, glutamine, and 2’-deoxyuridine, all involved in pyrimidine metabolism, were significantly lower in twins with NDCs than their unaffected co-twins in the within-pair CGEE model. This pattern suggests that alterations in specific components of pyrimidine metabolism may contribute to NDC liability independently of genetic or shared environmental influences. Glutamine, which is also modulated downstream of purine metabolism, has received considerable attention in NDC research due to its role in neurotransmitter balance. It cooperates with glutamate and GABA, which are crucial for maintaining excitatory–inhibitory homeostasis in the developing brain [75].

Among other differential findings, FMN levels were strongly negatively associated with NDC in the across-individual models, but not in the within-pair analysis, suggesting that shared genetic factors may mediate the relationship. While direct links between FMN and NDC are currently lacking, FMN serves as a critical cofactor for redox-active enzymes and ribonucleotide reductase [76] and may indirectly impact biological processes relevant to NDC physiology. In the within-pair model, the only metal that remained significantly associated with NDC was copper. Previous studies have reported elevated serum copper in autistic individuals, possibly contributing to oxidative stress [77]. However, our within-pair CGEE model employed a reduced sample size, especially for the discordant MZ twin subset, which diminished statistical power and increased the risk of false negatives.

Incorporating metallomics into the multi-omics framework offers valuable insights into environmental contributions to NDC mechanisms. In the differential metal profiles, we found that blood manganese levels were elevated across NDC individuals within the MZ subsample but were influenced by shared genetic or environmental factors. This may help explain part of the inconsistency in epidemiological studies of manganese exposure and NDC liability [78]. Indeed, polymorphisms in Mn-transporter genes have been shown to modulate manganese homeostasis and be associated with neurodevelopmental outcomes [79]. When genetic influence is not considered, blood manganese concentrations are generally not correlated with ingested manganese through drinking water and food [80]. Associations between blood manganese and NDC also converge with pathways highlighted earlier. In newborn rats, early-life oral manganese intake has been linked to attention deficits via dysregulation of gene expression in the prefrontal cortex, potentially involving mTOR signaling and neuroinflammatory mechanisms [81]. In addition, manganese overexposure may disrupt neurodevelopment through oxidative stress and neuroinflammation, which are also implicated in ADA function and purine metabolism [78]. Our findings underscore the complex and multilayered involvement of metal biology in pathways relevant to NDCs.

Our study has several limitations. First, our integrative multi-omics approach provides insights into NDC-associated candidate pathways. Still, it does not directly identify validated biomarkers or provide a predictive clinical model. Second, although substantial for a multi-omics twin design, our sample size limits our power to detect subtle cross-omics effects and to generalize to diverse populations, particularly within NDC diagnostic subgroups. Third, the scope and complexity of multi-omics outputs preclude exhaustive reporting, introducing potential selective emphasis in our discussion [82]. Fourth, WGS was limited to 14 MZ twin pairs, constraining variant-discovery power and generalizability, while proteomics was restricted to targeted Olink panels, potentially biasing findings toward represented pathways. Discordance status may also change over time, so some “discordant” pairs could reflect differences in NDC onset timing rather than stable biological discordance. Although no significant associations emerged between DIABLO variate scores and covariates after FDR correction, nominal associations were observed between age and variate 1, and between medication and variate 2; we cannot entirely rule out these as potential sources of residual confounding in the multi-omics signature. Future studies would benefit from untargeted proteome-wide screens and additional omics layers (*e.g*., transcriptomics, epigenomics) alongside richer phenotypic data.

In conclusion, this study presents a multi-omics framework embedded within a twin-cohort design to uncover potential coordinated molecular alterations underlying NDC. The convergence of signals across metabolomic, proteomic, blood metallomic, and genetic layers points to shared biological processes involving systemic metabolic dysregulation and immune-related signaling cascades. Notably, we observed several molecules that have not been well characterized in the NDC context (*e.g.,* the ADA protein and FMN), and emphasized that differential molecular levels may reflect underlying genetic or shared environmental influences rather than direct effects of NDCs. While limitations such as modest sample size constrain the generalizability of our conclusions, this study establishes a scalable framework for systems-level investigation and offers insights that may guide future biomarker discovery, liability stratification, and pathway-targeted interventions in NDC.

## Supporting information

Supplementary Materials (including supplementary methods, supplementary results, and supplementary figures)

## Acknowledgments

We would like to thank all twins and parents who have participated in this research. We would also like to thank the RATSS team, and coworkers at the KIND research center for their valuable contribution to the work presented in this study. We acknowledge the Swedish Twin Registry for data access. The Swedish Twin Registry is managed by Karolinska Institutet and receives funding through the Swedish Research Council under the grant no 2017-00641. The computations were performed using resources provided by the National Academic Infrastructure for Supercomputing in Sweden (NAISS) through Uppsala Multidisciplinary Center for Advanced Computational Science (UPPMAX). The project was supported by the Swedish Research Council (R.D.H., S.B. and K.T.), Swedish Foundation for Strategic Research (K.T.), the Swedish Brain Foundation – Hjärnfonden (K.T.), the Strategic Research Area Neuroscience StratNeuro (R.D.H., K.T.), The Committee for Research at KI (K.T.) and the China Scholarship Council (Y.Z.). Funding for RATSS was provided by the Swedish Research Council, Vinnova, Formas, FORTE, Hjärnfonden, Stockholm Brain Institute, Autism and Asperger Association Stockholm, Queen Silvia Jubilee Fund, Solstickan Foundation, PRIMA Child and Adult Psychiatry, the Pediatric Research Foundation at Astrid Lindgren Children’s Hospital, the Swedish Foundation for Strategic Research, Jerring Foundation, the Swedish Order of Freemasons, Kempe-Carlgrenska Foundation, Sunnderdahls Handikappsfond, the Jeansson Foundation, and EU-AIMS (European Autism Interventions), with support from the Innovative Medicines Initiative Joint Undertaking (grant agreement no. 115300), the resources of which are composed of financial contributions from the European Union’s Seventh Framework Programme (grant FP7/2007–2013), from the European Federation of Pharmaceutical Industries and Associations companies’ in-kind contributions, and from Autism Speaks. The results leading to this publication have also received funding from the Innovative Medicines Initiative 2 Joint Undertaking under grant agreement No 777394 for the project AIMS-2-TRIALS. This Joint Undertaking receives support from the European Union’s Horizon 2020 research and innovation programme and EFPIA and AUTISM SPEAKS, Autistica, SFARI. Any views expressed are those of the author(s) and not necessarily those of the funders (IHI-JU2).

## Author contributions

Y.Z.: Conceptualization, Methodology, Validation, Formal analysis, Data curation, Writing - original draft, Writing - review & editing, Visualization, Funding acquisition. B.v.W.: Formal analysis, Writing - original draft, Writing - review & editing. K.L.R.: Investigation, Resources, Data curation, Writing - review & editing. J.I.: Investigation, Resources, Data curation, Writing - review & editing. K.P.: Resources, Data curation, Writing - review & editing. A.G.: Resources, Data curation, Writing - review & editing. M.V.: Resources, Data curation, Writing - review & editing. T.B.: Resources, Data curation, Writing - review & editing, Funding acquisition. J.S.: Resources, Data curation, Writing - review & editing, Funding acquisition. M.K.: Resources, Data curation, Writing - review & editing, Funding acquisition. M.L.: Resources, Data curation, Writing - review & editing, Funding acquisition. R.D.H.: Resources, Data curation, Writing - review & editing, Funding acquisition. S.B.: Investigation, Resources, Data curation, Writing - review & editing, Supervision, Funding acquisition. K.T.: Conceptualization, Methodology, Validation, Investigation, Resources, Data curation, Writing - original draft, Writing - review & editing, Visualization, Supervision, Project administration, Funding acquisition.

## Data and Code availability

The data will be accessible, after necessary clearances, through the Swedish National Data Service’s (SND) research data catalogue. Custom code is available on GitHub (https://github.com/Tammimies-Lab/Multi-omics_RATSS.git).

## Supplementary Information

Supplementary Materials (including supplementary methods, supplementary results, and supplementary figures) and Supplementary Tables.

