## Supplementary Materials (including supplementary methods, supplementary results, and supplementary figures) for "Integrative Multi-Omics Analysis of Molecular Associations and Biological Pathways Underlying Neurodevelopmental Conditions in a Twin Cohort"

*Supplementary Materials include following parts:*

**Supplementary Methods**


S1 Study participants
S2 Behavioral Measures
S3 Biological sample processing
S3.1 Proteomics profiling of serum and CSF samples
S3.2 Metabolomics profiling of urine samples
S3.3 Metabolomic profiling of stool samples
S3.4 Metallomics profiling of blood samples
S4 Integrative omics analysis
S4.1 Data preprocessing and single-omics analysis
S4.2 Multi-omics analysis
S5 WGS rare variant identification


**Supplementary Results**


S1 Single omics analysis
S2 Integrative multi-omics analysis
S2.1 Multi-omics integration of serum-proteomics, CSF-proteomics, fecal-metabolomics, and blood-metallomics datasets
S2.2 Multi-omics integration of serum-proteomics, fecal-metabolomics, and blood-metallomics datasets
S2.3 Multi-omics integration of urine-metabolomics, fecal-metabolomics, and blood-metallomics datasets
S3 Differential analysis of molecules
S4 Metal exposure and NDC-related pathways
S5 Rare variants identified in WGS data


**Supplementary Figs (S1-S15)**

**Supplementary References**

*** The Supplementary Tables (S1-S17) are provided in a separate file (Supplementary Tables)**

### Supplementary Methods

#### S1 Study participants

The sample was drawn from the Roots of Autism and ADHD Twin Study in Sweden (RATSS) cohort [1, 2], a large collection of deeply phenotyped twin pairs with different NDC diagnoses, as well as neurotypically developing twin pairs, described in Supplementary Text and elsewhere in more detail [1]. Briefly, the twins were recruited from a population-based twin cohort as well as other sources with a specific recruitment towards autism and ADHD discordant twins. The exclusion criteria were profound intellectual disability (ID) with IQ < 35, serious psychiatric, neurological conditions, or known genetic syndromes. Within RATSS, a 2½-day comprehensive behavioral assessment using standardized instruments was conducted at a clinical research center. Twins were diagnostically assessed for NDCs and psychiatric disorders according to DSM-5 and ICD-10 criteria and consensus by a group of experienced clinicians [3]. The phenotypic information (*e.g.,* age, body mass index [BMI], medication usage) and biological samples (*e.g.,* blood, CSF, urine samples) were primarily collected from participants during the assessment at the unit, as well as via express service in some cases (fecal samples). The zygosity of twin pairs was determined by genotyping of saliva or whole-blood samples or using a panel of 48 single-nucleotide polymorphisms (SNPs) [4, 5]. Among the recruited twins, we selected 237 individuals with available one or several single omics data measured from the collected biological samples (Fig. S1, Table S1), including proteomics detected in serum and CSF [6], metabolomics detected in feces and urine [7], and metallomics detected in blood (Table S2). As part of the analysis, we also included available whole-genome sequencing (WGS) data from 14 MZ twin pairs who had also participated in the study "European Autism Interventions - A Multicenter Study for Developing New Medications" (EU-AIMS) [8].

#### S2 Behavioral Measures

To assess behavioral challenges, internationally widely used, age-appropriate instruments from the Achenbach System of Empirically Based Assessment collected in RATSS were used across all twins, including the parent-rated Child Behavior Checklist (CBCL) or Adult Behavior Checklist (ABCL), depending on the child's age, and the self-rated Youth Self-Report (YSR) or Adult Self-Report (ASR), depending on the participants' age. Adaptive functioning was assessed using the Adaptive Behavior Assessment System–Second Edition (ABAS-II), and autistic traits with the Social Responsiveness Scale 2nd Edition (SRS-2). Intellectual abilities were estimated using the Wechsler Intelligence Scales (WISC-IV for children, WAIS-IV for adults) or the Leiter scales. ADHD symptoms were measured using the attention subscales in CBCL or ABCL. For more detail regarding assessments, see Bölte et al [1]. The CBCL and ABCL total scores were standardized within their respective subsamples using z-score transformation and then merged into a unified outcome variable for subsequent analysis. The YSR and ASR scores were processed in a similar manner.

#### S3 Biological sample processing

##### S3.1 Proteomics profiling of serum and CSF samples

Full details of the serum and CSF proteomic profiling protocol are available in Smedler et al [6]. Briefly, blood samples were processed by coagulation and centrifugation to obtain serum, while CSF was collected by lumbar puncture performed by an experienced neurologist. Samples were aliquoted and stored at −80 °C at the Karolinska Institutet Biobank, Sweden, until analysis. Protein concentrations in serum and CSF were measured using three Olink multiplex immunoassay panels (Inflammation I, Cardiovascular I, and Oncology I; Olink Bioscience, Sweden), covering 203 different proteins across key biological pathways including inflammation, immune response, and cellular metabolism. The assays are based on proximity extension assay technology, which enables high-throughput and sensitive multiplex quantification of protein biomarkers.

##### S3.2 Metabolomics profiling of urine samples

Urine metabolite profiling was performed using untargeted 1H nuclear magnetic resonance (NMR) spectroscopy as previously described [7]. In brief, 112 urine samples were analyzed on a Bruker 600 MHz NMR spectrometer equipped with a SampleJet autosampler. Spectra were acquired using a standard one-dimensional solvent suppression pulse sequence and processed in Topspin 3.2 for phase and baseline correction, followed by manual alignment and probabilistic quotient normalization in Matlab (Version 2018a, MathWorks Inc.). Spectral regions corresponding to water, internal standard, and urea were excluded prior to normalization.

##### S3.3 Metabolomic profiling of stool samples

Untargeted metabolomic profiling of fecal samples from participants in the RATSS was conducted by Metabolon, Inc. (Durham, NC, USA). Stool aliquots were collected using sterile containers, immediately frozen at −80 °C, and shipped on dry ice. Sample preparation followed Metabolon's standardized automated MicroLab STAR® protocol (Hamilton Company). Recovery standards were added prior to protein precipitation with methanol to monitor extraction efficiency. After vigorous shaking for 2 min and centrifugation, the supernatant was divided into four aliquots, evaporated to dryness under nitrogen using a TurboVap® (Zymark), and stored overnight. Each aliquot was reconstituted in method-specific solvents containing internal standards to ensure chromatographic consistency.

Samples were analyzed using a Waters Acquity ultra-performance liquid chromatography (UPLC) system coupled to a Thermo Scientific Q-Exactive high-resolution mass spectrometer (MS) with a heated electrospray ionization source (HESI-II) and Orbitrap mass analyzer. Four complementary UPLC-MS/MS methods were applied: two reverse-phase (RP) methods under positive ionization optimized for hydrophilic or hydrophobic compounds, one RP method under basic negative ionization (6.5 mM ammonium bicarbonate, pH 8.0), and one hydrophilic-interaction liquid chromatography (HILIC) method under negative ionization (10 mM ammonium formate, pH 10.8). The mass spectrometer alternated between full-scan MS and data-dependent $MS^n$ acquisition across an m/z range of 70–1000. Raw data were processed using Metabolon's proprietary library of authenticated standards for retention-time, accurate-mass, and MS/MS spectral matching.

##### S3.4 Metallomics profiling of blood samples

The metallomics data comprise concentrations of multiple essential [calcium (Ca), copper (Cu), iron (Fe), magnesium (Mg), manganese (Mn), molybdenum (Mo), selenium (Se), and zinc (Zn)] and toxic metals [arsenic (As), cadmium (Cd), cobalt (Co), mercury (Hg), and lead (Pb)] in whole blood. Although selenium is not a metal, we included it in that concept for simplicity. All the essential metals as well as toxic metals derive from the diet, including drinking water. The major part of total arsenic concentrations in blood among Swedish individuals usually consists of arsenobetaine, originating almost exclusively from marine fish and shellfish.[9] Similarly, blood mercury is mainly methylmercury from seafood, as exposure to inorganic mercury is rare in Swedish adolescents [10]. The metal concentrations in blood were measured using inductively coupled plasma mass spectrometry (ICP-MS; Agilent 7900, Agilent Technologies, Tokyo, Japan) with an octopole reaction system. Blood samples were diluted 1:25 in an alkali solution containing 2% (w/v) 1-butanol, 0.05% (w/v) EDTA, 0.05% (w/v) Triton X-100, 1% (w/v) NH4OH, and 20 µg/L of internal standard [11]. As quality control (QC), two commercial whole blood reference materials were measured several times in each run [Seronorm whole blood L-1 (1103128) and L-2 (1406264)]. The limit of detection (LOD) was defined as three times the standard deviation of the blank concentrations. The obtained values in QC showed a good agreement with the reference values, except Fe (Table S15). The sensitivity for Fe was relatively poor, likely leading to underestimation of its levels. Since the reference materials provide only point value for Fe (as for some other elements) and the potential bias appeared consistent across samples, Fe was still retained in the analysis. Additionally, as our multi-omics models rely primarily on correlation structures rather than absolute concentrations, and no quantitative interpretation was included, this is unlikely to substantially affect the results.

#### S4 Integrative omics analysis

##### S4.1 Data preprocessing and single-omics analysis

We performed quality control and preprocessing for each omics dataset. Molecules with more than 50 percent missing values were removed, the remaining values were imputed using the K-nearest neighbors method (k=10), and all datasets were z-score normalized, respectively. After quality controls, the blood dataset contained 13 metals, the CSF dataset contained 86 proteins, the serum dataset contained 167 proteins, the fecal dataset retained 577 named metabolites, and the urine dataset contained 59 metabolites. We conducted principal component analysis (PCA) on the individuals with complete serum-proteomics, CSF-proteomics, fecal-metabolomics, and blood-metallomics data using prcomp() function in R.

We applied sparse Partial Least Squares Discriminant Analysis (sPLS-DA) to evaluate the number of key variables in each omics dataset [12]. The sPLS-DA model computes latent components that maximize covariance between datasets while fitting a supervised model that discriminates individuals with and without NDC. Through lasso penalization, it identifies a subset of key molecules that contribute most to this discrimination. To optimize model parameters, we employed the perf() function to select the optimal number of components, the type of distance measurements, and the calculation method of classification error rate that yielded the best predictive performance in models. Afterwards, the tune() function was used to select the optimal number of variables to retain based on the identified best-performing parameter combinations. Model performance was assessed using 10-fold cross-validation (CV) with 10 repeats. The parameters tested and selected are detailed in Table S3.

##### S4.2 Multi-omics analysis

The integrative multi-omics model construction pipeline is shown in Fig. S3. Multi-omics integration was performed using the DIABLO (Data Integration Analysis for Biomarker Discovery using Latent Variable Approaches for Omics Studies) framework implemented in the mixOmics R package [13]. DIABLO is a supervised N-integration method that combines multiple datasets from individuals with complete data across all included layers in relation to a categorical outcome variable. It applies a multiblock sPLS-DA to identify correlated molecules across multiple omics layers while optimizing discrimination between anyNDC and noNDC groups. For small sample sizes, the generalization, cross-validation strategies for parameter tuning, and feature selection minimize risk of overfitting. The model construction required a design matrix that defines the correlation structure between input datasets. We selected 0.1 to prioritize the discrimination power of models. For parameter tuning, we applied a 10-fold CV with 10 repeats, incorporating the optimized number of key molecules identified in single-omics analysis for each dataset. The best-performing number of components and number of variables to retain in each component for each omics dataset were used to construct the final multi-omics model. The tuning parameters and optimized parameters used for each multi-omics model are detailed in Table S3. To evaluate feature stability, we applied perf() function from the mixOmics package to assess feature selection frequency across 10 CV folds with 10 repeats, reported in Supplementary Table S16.

##### S5 WGS rare variant identification

Relatedness was confirmed using the --relatedness2 function in VCFtools (v0.1.16). During the quality control process, we filtered variants based on the GATK-suggested hard filters of germline short variants. Specifically, we excluded variants with QD < 2, FS > 60, SOR > 3, MQ < 40, MQRankSum < -12.5, or ReadPosRankSum < -8.0. Variants with missing genotype calls in any individual were also removed. We annotated variants using ANNOVAR (v2020.06.08) and VEP (v111). Exonic variants were identified based on Func.refGene annotations generated by ANNOVAR, including those labelled as "exonic," "splicing," or "ncRNA_exonic." The allele frequencies were referenced against gnomAD (v2.1.1) and the SweFreq database (SweGen Variant Frequency Dataset, accessed 2019-02-04) [14, 15]. The neurodevelopmentally-related gene list included SFARI genes (04-03-2023 release) [16] and genes listed in the Intellectual Disability - Microarray and Sequencing panel (Version 5.191) [17]. SFARI Gene is a curated, community-driven knowledgebase cataloguing genes with evidence of association with autism spectrum disorder, scored by strength of genetic evidence. We included SFARI genes categorized as syndromic or with gene scores of 1 or 2, representing the highest confidence autism-associated genes. For the analysis of genes with unique variants, we included genes harboring unique variants exclusively in individuals with any NDC and excluded those with any such variants in noNDC individuals. For the analysis of genes with shared variants, we selected genes containing variants shared within concordant and discordant NDC twin pairs, while excluding genes with variants shared within concordant noNDC twin pairs.

#### Supplementary Results

##### S1 Single omics analysis

After quality control procedures (described in method section), the blood dataset included 13 metals across 220 samples. The serum and CSF datasets included 167 and 111 proteins from 126 and 86 samples, respectively. The fecal and urine dataset retained 577 and 59 metabolites identified from 189 and 112 samples, respectively.

We performed sparse Partial Least Squares Discriminant Analysis (sPLS-DA) on each single-omics dataset to calculate the optimal number of features for NDC status discrimination. We found that the serum dataset required up to six components and 30 features per component to achieve the highest classification performance on NDC, indicating a more complex protein structure relevant to NDC in serum. The other four datasets achieved optimal discrimination with only one to three latent components. Despite the large feature space (577 metabolites), the fecal dataset reached its optimal performance with five and 50 key metabolites in two components, suggesting that most of the metabolites in fecal samples have limited combined discriminatory power for NDC status. Detailed results of parameter tuning and optimal model results are provided in Table S3.

##### S2 Integrative multi-omics analysis

We initially applied PCA using the molecular concentrations from integrated datasets (integrating serum-proteomics, CSF-proteomics, fecal-metabolomics, and blood-metallomics). However, this unsupervised approach failed to cluster individuals according to clinical characteristics (Fig. S14). We therefore adopted a supervised strategy, applying a multiblock sPLS-DA to investigate molecular relationships across different omics layers while optimizing the discrimination between anyNDC and noNDC groups. The missingness of key molecules identified in multiomics models before imputation was reported in Supplementary Table S17. Missingness was 0% for 60% of identified features, the 75th percentile was 3%, and the 90th percentile was 14%, with a maximum of 45%.

##### S2.1 Multi-omics integration of serum-proteomics, CSF-proteomics, fecal-metabolomics, and blood-metallomics datasets

In circos plot of *MultiOmic-SCFB* model, serum proteins showed limited connectivity with other omics layers, suggesting a more isolated molecular domain in this integrative model. Within serum features, only TNFRSF4 showed a strong correlation ($|r| \geq 0.6$) with other omics layers, showing positive correlation with Mn and Fe levels and negative correlation with Ca levels. More extensive correlations ($|r| \geq 0.6$) were observed between blood metals, fecal metabolites, and CSF proteins. Within the fecal features, cis-4-hydroxycyclohexylacetic acid showed positive correlations with several CSF proteins, including CCSL3, PSGL-1, FS, and DKK-1. These correlated molecules were mainly loaded on component 2 of the model, implying they may participate in a shared gut–central nervous system pathway relevant to NDC phenotypes. We also observed some cross-omics correlations involving three omics layers. Zn, Fe, and Mg were negatively correlated with fecal feature N-acetylglutamate and 3-methyladipate, and CSF feature GH. Here, 3-methyladipate and GH were also positively correlated with each other, reflecting a co-regulated metabolic module. Additionally, we identified a triangular correlation motif between 3-methyladipate, Ca, and CSF protein FUR, consistent with FUR's role as a calcium-dependent serine protease potentially responsive to metabolic or oxidative stress.

To evaluate whether the DIABLO multi-omics latent components capture NDC-related molecular variation independently of demographic and clinical covariates, we regressed variate scores from components 1 and 2 of *MultiOmic-SCFB* model against age, sex, BMI, medication use, and other psychiatric diagnoses using GEE models accounting for twin pair structure (Table. S4). *P*-values were FDR-corrected across the 10 covariate tests within each model. We observed nominally significant associations for age with variate 1 ($p$=0.020) and medication with variate 2 ($p$=0.026), but neither survived FDR correction (adjusted-$p$>0.05). No significant associations were observed after correction in two models, suggesting that the multi-omics variate scores reflect NDC-associated molecular patterns rather than covariate-driven variation.

##### S2.2 Multi-omics integration of serum-proteomics, fecal-metabolomics and blood-metallomics datasets

To include a larger sample size, we then integrated the blood, serum, and fecal datasets, resulting in a total of 109 individuals with complete data. Using parameters after tuning (Table S3), the multi-block sPLS-DA model constructed from this integration (*MultiOmic-SFB*) identified 86 key molecular features using three latent components, including 29 proteins, 44 metabolites, and 13 metals. However, compared to the previous *MultiOmic-SCFB* model, the absence of CSF data resulted in reduced discriminative power, despite the larger sample size (Figs. S5A-E). From circos plot we can see that more proteins exhibited strong correlations ($|r| \geq 0.6$) with molecules from other omics layers (Figs. S5F-I). Within the identified metabolites, myristoleate and 5-dodecenoate were positively correlated with multiple proteins, reflecting their roles as endpoints or intermediates in metabolic pathways regulated by diverse protein networks. Additionally, Co, which was also negatively associated with these two metabolites, showed positive correlations with several of the same proteins (MIP-1 alpha, HB-EGF, and HGF). Consistent with findings from the previous *MultiOmic-SCFB* model, TNFRSF4 again exhibited positive correlations with blood Mg and Fe levels, and a negative correlation with Ca level. The protein ADA displayed a similar correlation pattern with these metals as TNFRSF4 and was additionally positively associated with zinc.

##### S2.3 Multi-omics integration of urine-metabolomics, fecal-metabolomics, and blood-metallomics datasets

To address the limited availability of biosamples from the same individuals, we integrated urine with fecal and blood datasets, yielding a subsample of 89 individuals (29 anyNDC and 60 noNDC), which is the largest subset available for multiomics analysis including urine data. The resulting multi-block sPLS-DA model (*MultiOmic-UFB*), optimized after parameter tuning (Table S3), identified 58 key molecules across two latent components, including 35 fecal metabolites, 10 urinary metabolites, and 13 metals. Despite both being metabolomics layers, the strongest cross-block correlation was observed between urine and fecal datasets (r = 0.46), highlighting the complementary value of multi-tissue sampling (Figs. S6A-BB). When samples were projected to the latent component space, fecal block showed the greatest discriminative ability, while urine block captured a distinct subgroup of NDC cases, suggesting potential subtype-specific signals (Fig. S6C). The averaged projection across blocks still achieved better discrimination than any single omics layer (Fig. S6D). In circos plot (Figs. S6E-G), urine metabolites had more high correlations (|r| > 0.6) with other omics layers. Interestingly, branched-chain amino acid catabolites in urine (valine, HMB, and 3-hydroxyisovalerate) were strongly correlated with methionine-related metabolites in fecal samples (N-acetylmethioninesulfoxide and N-formylmethionine) on component 2, suggesting coordinated disruption of host BCAA catabolism and microbial metabolism in NDC. An additional unknown metabolite (d 6.70), highly correlated with Mn and inversely with fecal glycerol-3-phosphate, may also represent a novel marker of interest.

When comparing key molecules in three multi-omics models, valine was the only metabolite consistently identified in both fecal and urine samples. In *MultiOmic-UFB* model, only 20% of fecal key metabolites have also been identified in previous two models (including 2'-deoxyuridine, thymidine, pseudouridine, 2'-deoxyinosine, Flavin mononucleotide, 1-palmitoyl-2-oleoyl-GPC and 5-dodecenoate) and they are involved in diverse biological processes. The different integrated omics layers would provide their specific correlated molecular signatures, and each may contribute to a more comprehensive understanding of NDC etiology.

#### S3 Differential analysis of molecules

In addition to the molecules with levels associated with NDC diagnosis on the FDR-adjusted statistical significance, several molecules exhibited associations on nominal significance level. For the fecal metabolites in the whole samples, the across-individual GEE models revealed a nominal decrease of 2'-deoxyinosine level in the NDC group ($p$=0.014). In this within-pair model, only deoxycarnitine showed a nominally significant negative association with NDC ($p$=0.022). To further minimize genetic variability and other family-constant confounding, we performed same analyses restricted to MZ twins with fecal samples (Table S8). In GEE model including 134 individuals, 2'-deoxyinosine ($p$=0.007) showed results consistent with the trend observed in the whole individuals, while 2-aminoadipate was nominally elevated in NDC ($p$=0.009, Table 2). However, in within-pair models, these relations between three metabolites and NDC were not significant and only glutamine negatively associated with NDC at the nominal significance level ($p$=0.048). Urinary metabolites showed no significant differentiation across any of the tested models (Table S9). In the metallomic analysis, Se showed a nominal positive association with NDC diagnosis in the across-individual GEE models in all individuals ($p$=0.028) and MZ twin subset ($p$=0.028), though this was not maintained in within-pair comparisons (Table S10).

To explore whether the identified molecular associations were specific to particular NDC diagnoses, we conducted exploratory analyses using ADHD and autism diagnostic status as separate outcomes for molecules identified as significantly associated in the primary analysis. Individuals with comorbid ADHD and autism diagnoses were included in both subgroup analyses, reflecting clinical diagnostic overlap inherent to NDC populations. Available sample sizes varied across omics datasets and model types. We only applied the specific sample groups and model types (GEE or CGEE) in which each molecule showed a significant association with anyNDC in the primary analysis. Across molecules, the statistical significance of subgroup-specific associations varied, but effect directions were broadly consistent with the primary anyNDC analyses. For serum ADA, only ADHD showed nominally significantly higher levels in affected individuals across sample subgroups and GEE models, consistent with the primary anyNDC analysis (Table S11). For fecal metabolite FMN, the GEE model in MZ twins showed nominally significant associations with both ADHD (*p*=0.0003) and ASD (*p*=0.0038), though significance was lost in the whole individual group. For fecal metabolites thymidine, glutamine, and 2'-deoxyuridine, only glutamine showed a nominally significant negative association with ADHD (*p*=0.0011), while associations observed in primary anyNDC models were not retained for the other metabolites in subgroup analysis. For blood metals, Mn showed nominally significantly increased levels in ADHD in GEE model restricted to MZ twins (*p*=0.0029), and Cu showed nominally significantly elevated levels in ADHD in CGEE model for discordant MZ twins (*p*=0.038), consistent with the primary anyNDC findings. These heterogeneous patterns likely reflect both the genuine molecular heterogeneity across NDCs and especially insufficient statistical power for subgroup-specific detection, compounded by diagnostic overlap between subgroups given the high co-occurrence of ADHD and autism in this cohort.

#### S4 Metal exposure and NDC-related pathways

We found that seven pathways previously identified in our joint enrichment analysis of metabolites and proteins remained significantly enriched for these metal-associated genes (FDR-adjusted *p*<0.05), including PI3K-Akt signaling pathway, MAPK signaling pathway, HIF-1 signaling pathway, Proteoglycans in cancer, EGFR tyrosine kinase inhibitor resistance, Chagas disease, and Malaria. Additionally, the Lysine degradation pathway that was significantly enriched with metal-associated genes (FDR-adjusted *p*=0.01) was also nominally enriched in our prior overall *p*-value method-based joint enrichment analysis on metabolic pathways.

#### S5 Rare variants identified in WGS data

The divergent results observed between across-individuals GEE and within-pair CGEE models in molecular differentiation analyses indicate a genetic contribution to the molecular signatures of NDCs. To investigate genetic factors, we analyzed WGS data from 14 MZ twins, including 3 discordant anyNDC pairs, 6 concordant anyNDC pairs, and 5 concordant noNDC pairs. No significant differences were observed between three groups in the number of rare unique variants (AF<0.001; Kruskal–Wallis, *p*=0.76) or rare exonic shared variants (AF<0.01, restricted to NDC-associated genes; Kruskal–Wallis, *p*=0.072) (Fig. S15).

#### Supplementary Figs

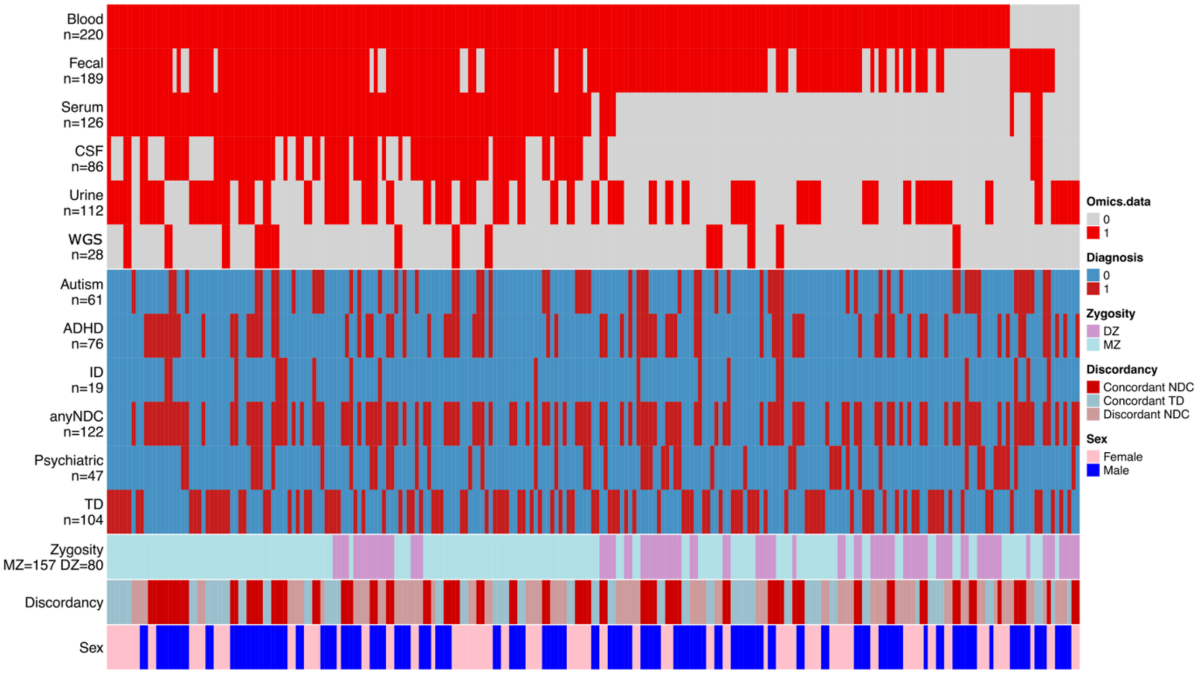


**Fig. S1 Overview of study population and data types.** Data modalities and phenotypic measures are organized by rows (n = number of individuals per dataset), with individual samples represented by columns. Grey bars indicate missing data. Omics layers include urine and fecal metabolomics, serum and CSF proteomics, blood metal ions, and whole genome sequencing data in the EU-AIMS study. NDC Diagnosis and medical data are aligned accordingly.

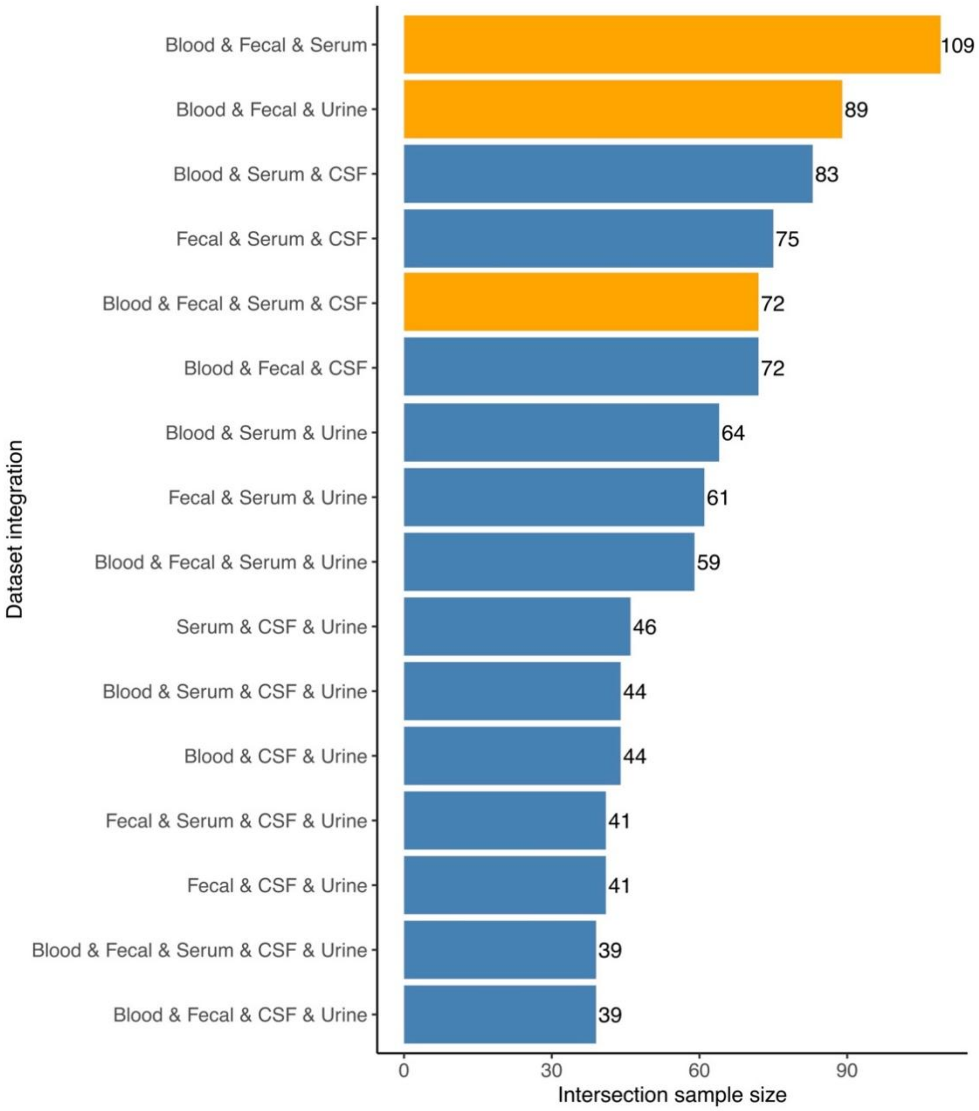


**Fig. S2 Overlapping samples among different combinations of omics datasets.**

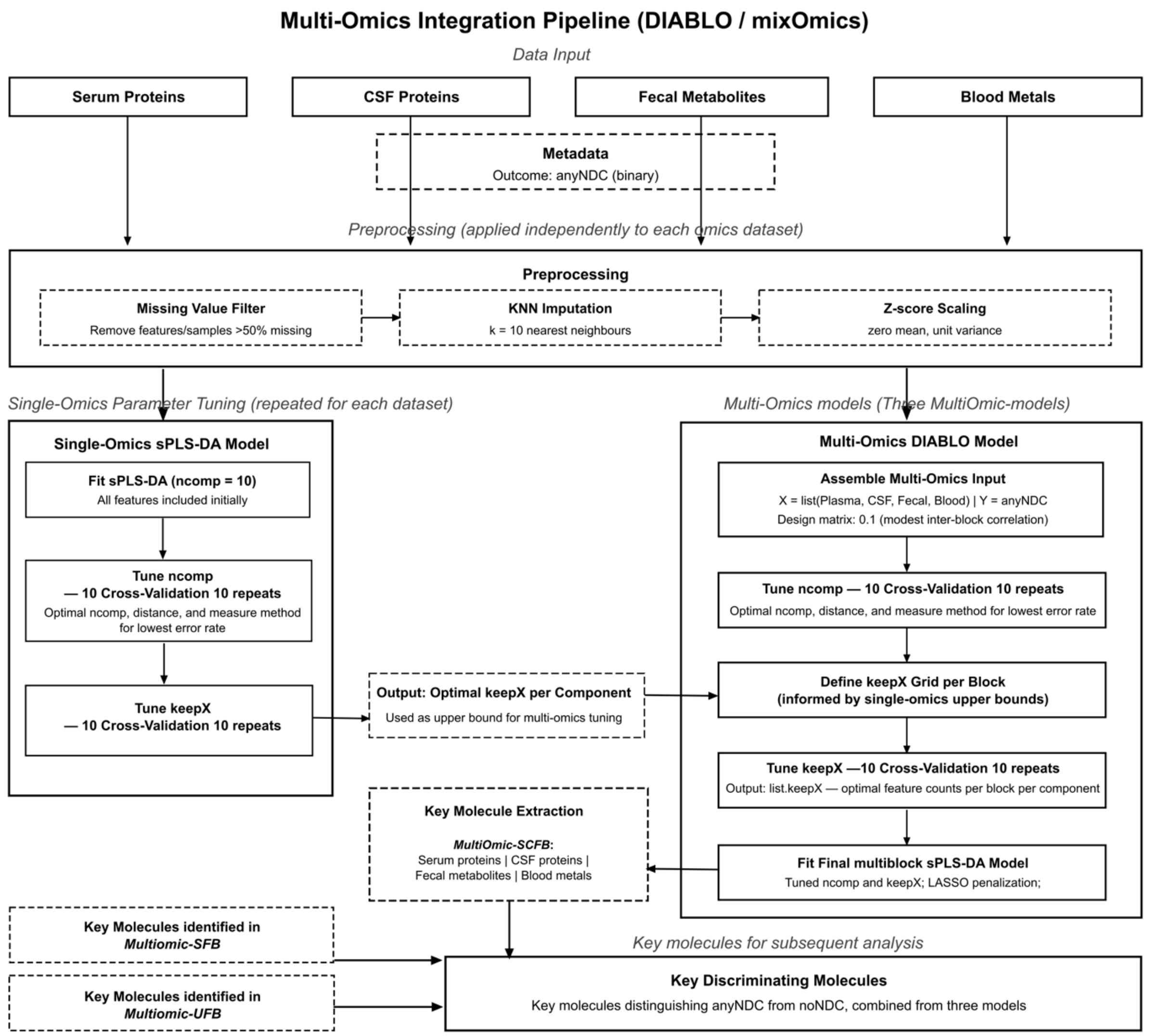


**Fig. S3 Schematic workflow of DIABLO multi-omics model construction.** For *MultiOmic-SCFB* model, four omics datasets (serum proteins, CSF proteins, fecal metabolites, blood metals) are preprocessed independently, including missing value filtering, KNN imputation, and z-score scaling. For each dataset, a single-omics sPLS-DA model is tuned separately (component number and keepX) via repeated cross-validation to determine optimal parameters, which inform tuning of the multi-omics DIABLO model. The multiblock sPLS-DA input is assembled from the omics blocks and outcome labels, with a design matrix specifying inter-omics correlation. Component number and keepX are tuned through repeated cross-validation, and the final multiblock model is fitted and key molecules are selected by LASSO penalization on the latent components. This process is applied to the other two multi-omics models *MultiOmic-SFB* (serum proteins, fecal metabolites, blood metals) and *MultiOmic-UFB* (urine metabolites, fecal metabolites, blood metals), yielding key discriminating molecules distinguishing anyNDC from noNDC across the combined models.

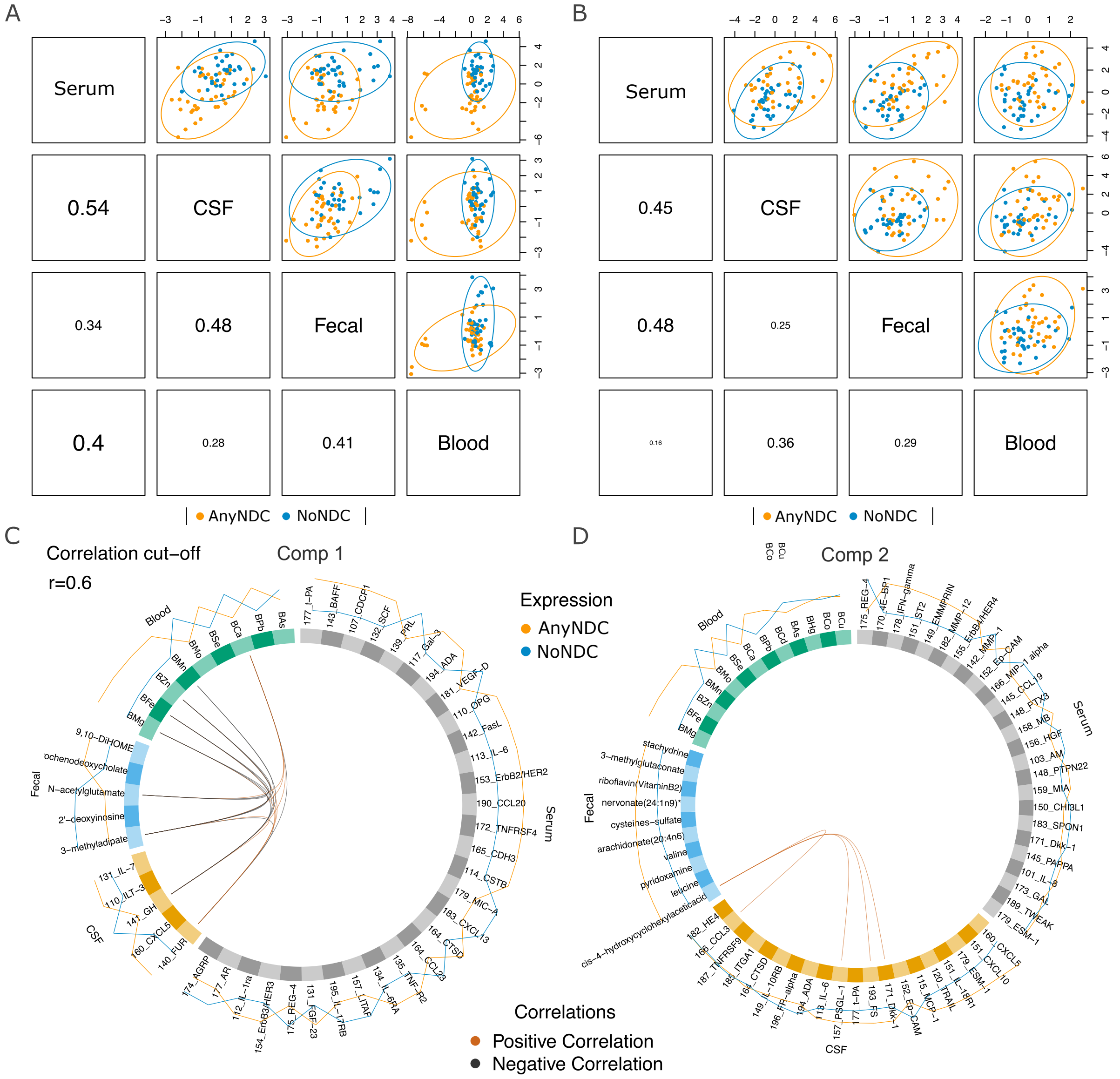


**Fig. S4 Overview of integrative multi-omics projections and cross-omics correlation structure in the *MultiOmic-SCFB* model.** Integration of serum-proteomics, CSF-proteomics, fecal-metabolomics, and blood-metallomics. (A-B) Diagnostic plots for the correlations between latent components from each data set, with the colours and ellipses representing the sample subgroups and indicating the discriminative power of each component to separate the different subgroups. (C-D) Circos plots displaying pairwise correlations between molecules across different omics datasets on each component. Edges represent cross-omics correlations with an absolute correlation coefficient ≥ 0.6; positive correlations are shown in orange, and negative correlations in grey. The outer rings around the circumference of the plot represent molecules abundance or expression level stratified by NDC status (anyNDC in orange, noNDC in blue).

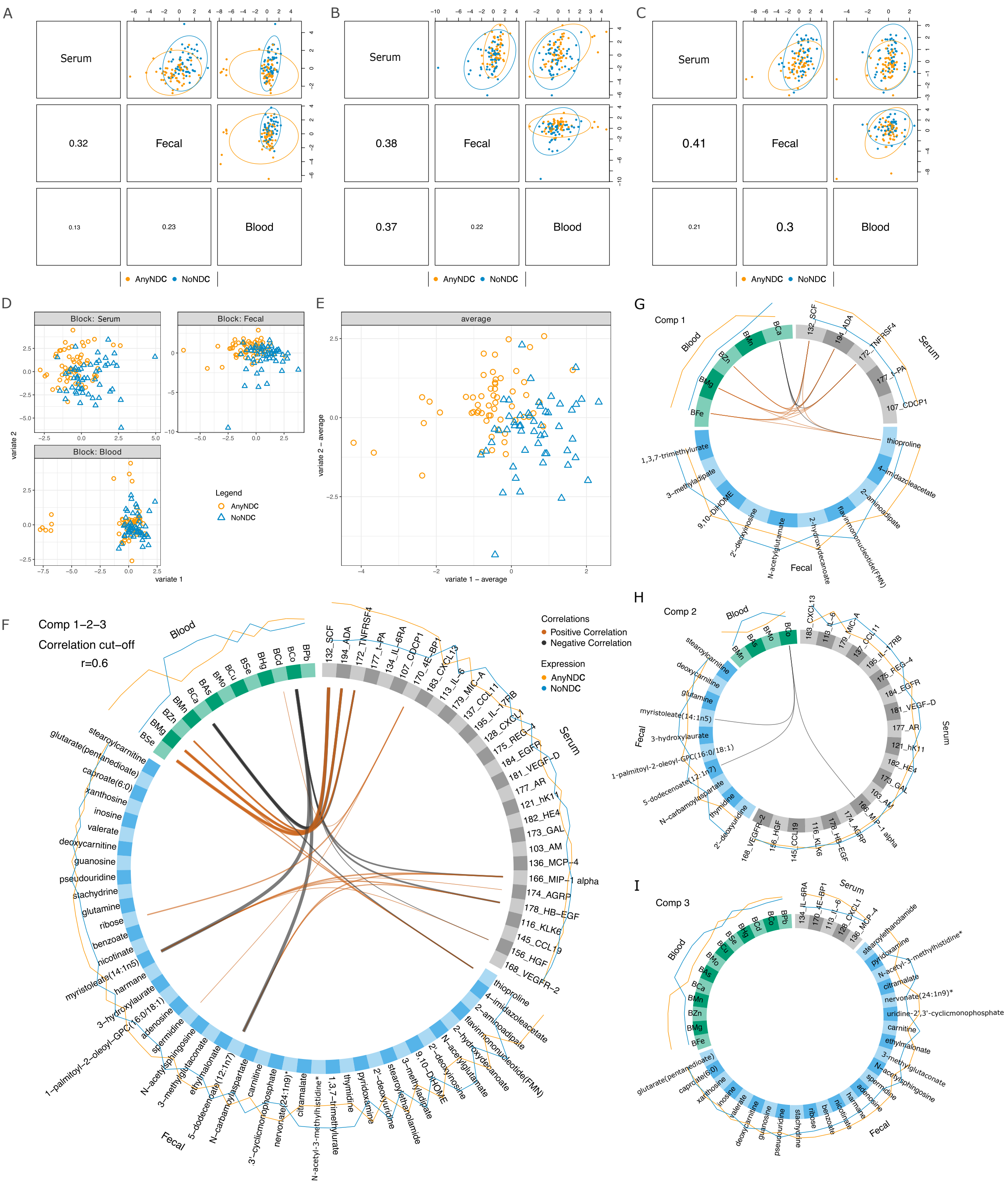


**Fig. S5 Overview of integrative multi-omics projections and inter-omics correlation structure in the *MultiOmic-SFB* model.** Integration of serum-proteomics, fecal-metabolomics, and blood-metallomics datasets**.** (A-C) Diagnostic plots for the correlations between latent components from each data set, with the colours and ellipses representing the sample subgroups and indicating the discriminative power of each component to separate the different subgroups. D. Sample projection plots for each omics block, displaying individual samples in the space defined by the first two components derived from the model. Each panel corresponds to a different omics dataset and shows the discriminative capacity of each omics layer, colored with NDC status (anyNDC in orange and noNDC in blue). E. Averaged component plot across all omics blocks, combining scores from components 1 and 2 into a single integrative sample-space representation. (F-I) Circos plots displaying pairwise correlations between molecules across different omics datasets on averaged (F) component and each component (G-I). Edges represent cross-omics correlations with an absolute correlation coefficient ≥ 0.6; positive correlations are shown in orange, and negative correlations in grey. The outer rings around the circumference of the plot represent molecule abundance or expression level stratified by NDC status (anyNDC in orange, noNDC in blue).

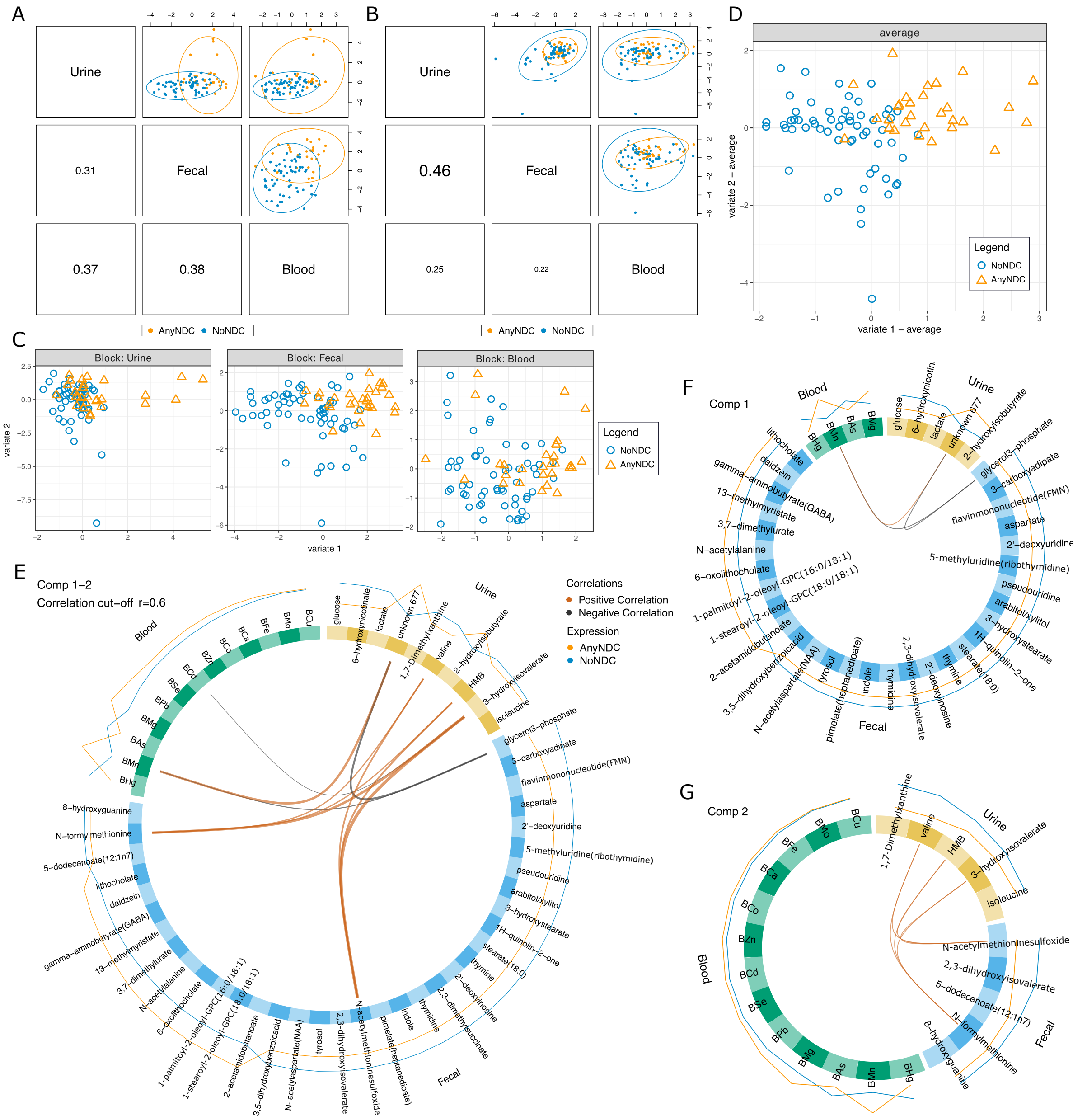


**Fig. S6 Overview of integrative multi-omics projections and inter-omics correlation structure in the *MultiOmic-UFB* model.** Integration of urine-metabolomics, fecal-metabolomics, and blood-metallomics datasets. (A-B) Diagnostic plots for the correlations between latent components from each data set, with the colours and ellipses representing the sample subgroups and indicating the discriminative power of each component to separate the different subgroups. C. Sample projection plots for each omics block, displaying individual samples in the space defined by the first two components derived from the model. Each panel corresponds to a different omics dataset and shows the discriminative capacity of each omics layer, colored with NDC status (anyNDC in orange and noNDC in blue). D. Averaged component plot across all omics blocks, combining scores from components 1 and 2 into a single integrative sample-space representation. (E-G) Circos plots displaying pairwise correlations between molecules across different omics datasets on averaged (E) component and each component (F-G). Edges represent cross-omics correlations with an absolute correlation coefficient ≥ 0.6; positive correlations are shown in orange, and negative correlations in grey. The outer rings around the circumference of the plot represent molecules abundance or expression level stratified by NDC status (anyNDC in orange, noNDC in blue)

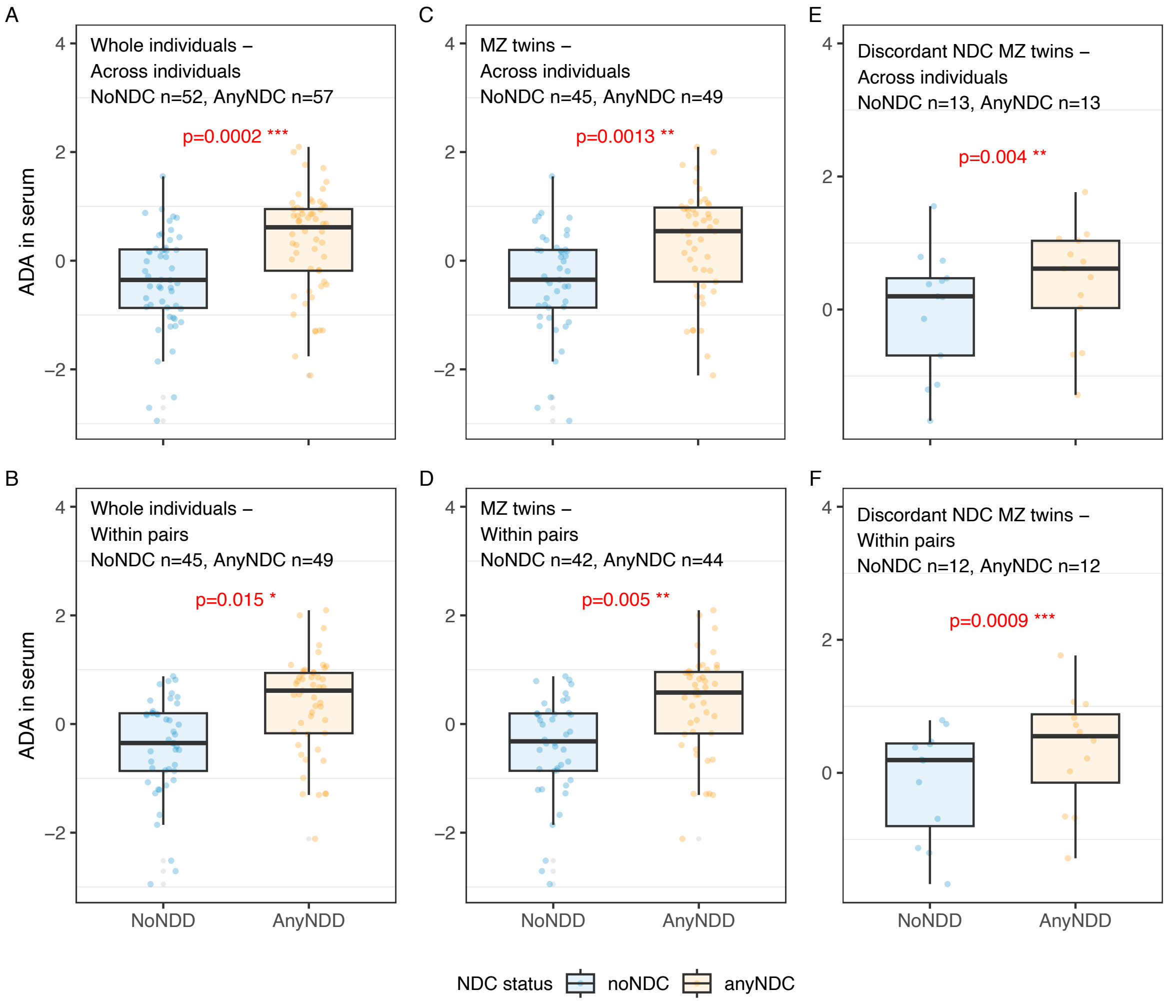


**Fig. S7 Distribution of serum ADA protein levels across diagnostic subgroups and subsamples.** Plots show ADA abundance in individuals stratified by NDC diagnosis and grouped into analytic subsets: (A) All individuals using a GEE model, (B) All individuals using a CGEE model, (C) MZ twins using GEE, (D) MZ twins using CGEE, (E) Discordant MZ twins using GEE, and (F) Discordant MZ twins using CGEE. Nominal *p*-values for group differences are shown for each comparison. The detailed estimates obtained from the models are presented in Supplementary Table S7.

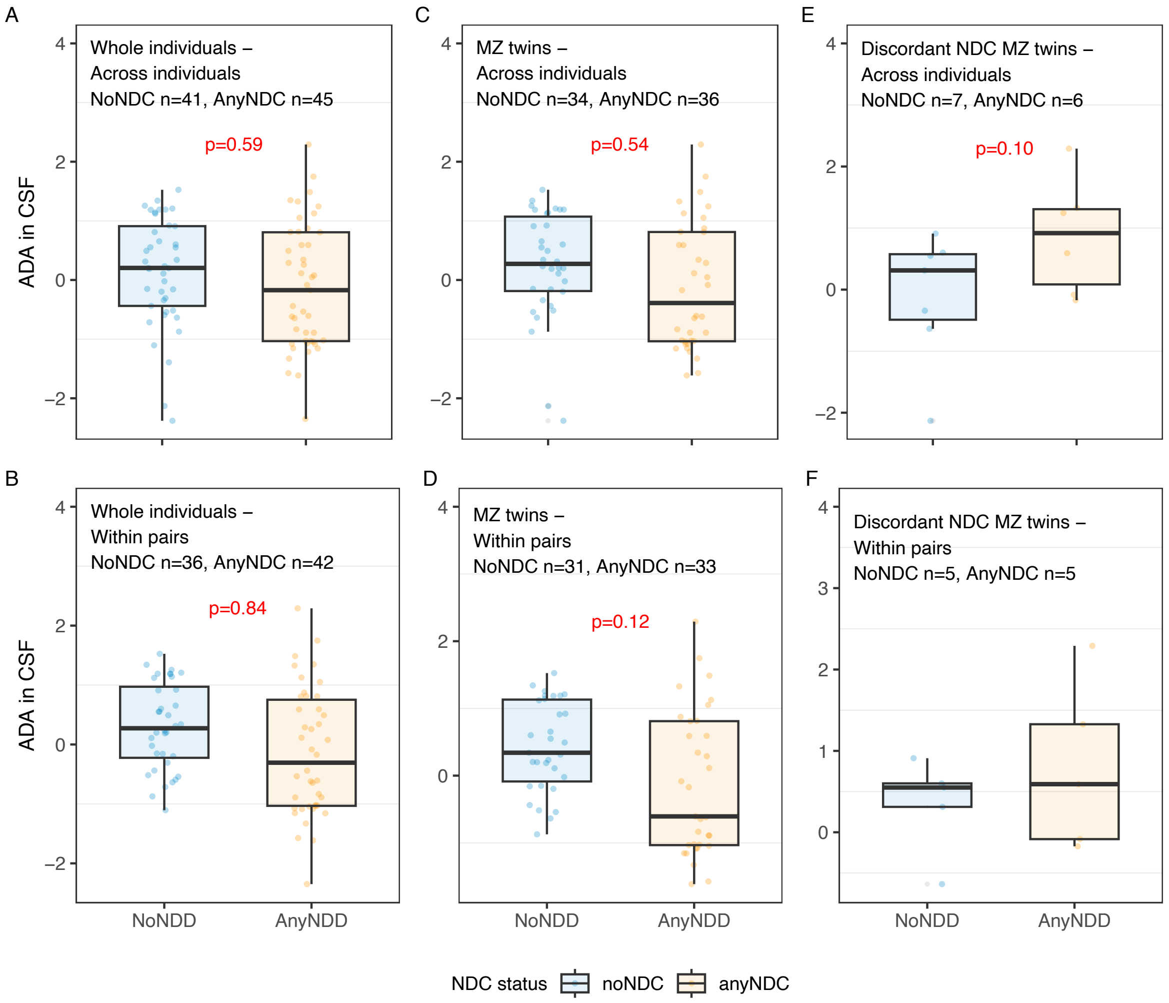


**Fig. S8 Distribution of CSF ADA protein levels across diagnostic subgroups and subsamples.** Plots show ADA abundance in individuals stratified by NDC diagnosis and grouped into analytic subsets: (A) All individuals using a GEE model, (B) All individuals using a CGEE model, (C) MZ twins using GEE, (D) MZ twins using CGEE, (E) Discordant MZ twins using GEE, and (F) Discordant MZ twins using CGEE. Nominal *p*-values for group differences are shown for each comparison. The detailed estimates obtained from the models are presented in Supplementary Table S7.

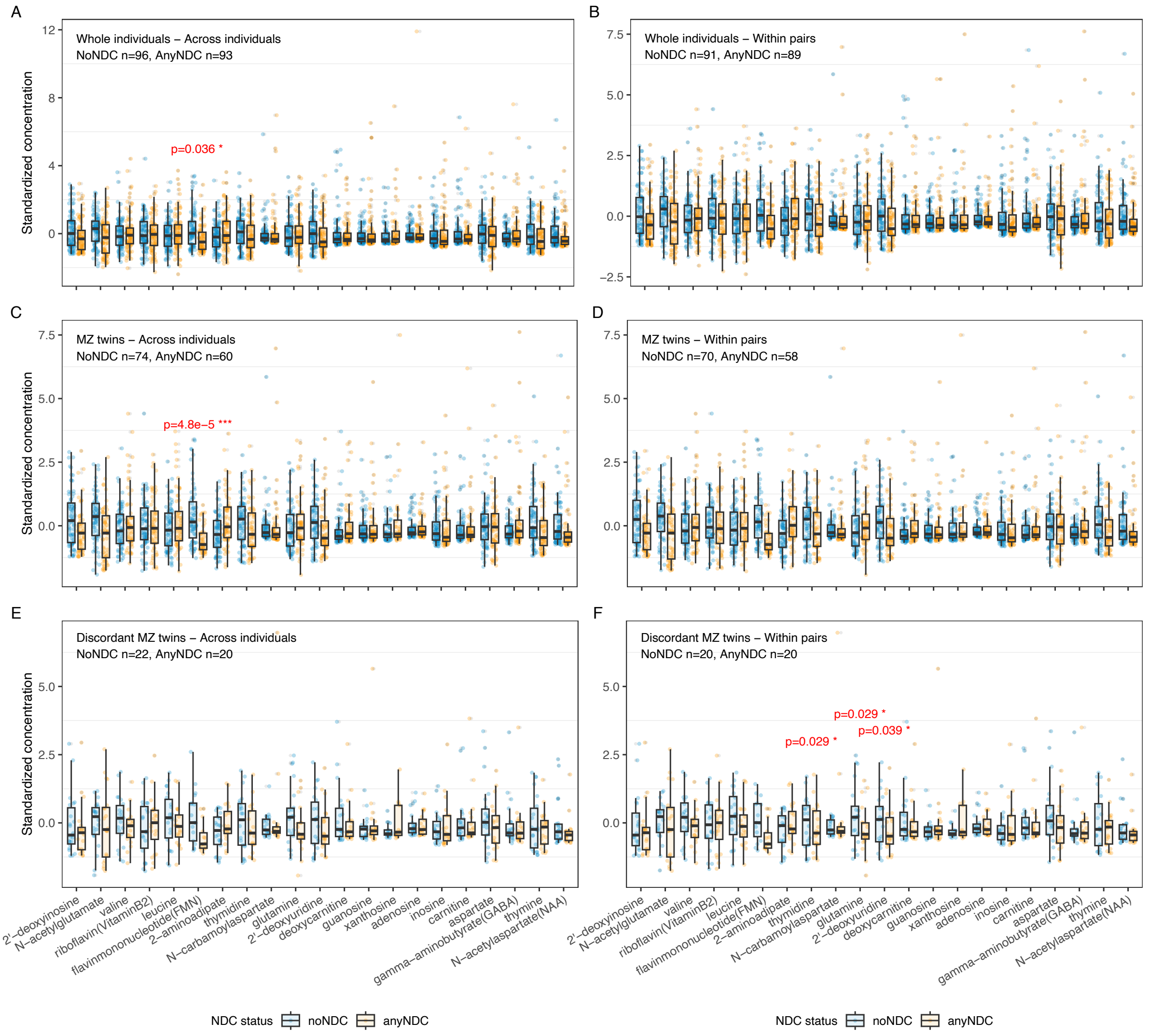


**Fig. S9 Group-wise distribution of selected fecal metabolites across subsamples used in differentiation analyses.** Plots show metabolite abundance across diagnostic groups (anyNDC vs. noNDC) in various analytic subsets: (A) All individuals using a cross individuals generalized estimating equation (GEE) model, (B) All individuals using a within-pair conditional GEE (CGEE) model, (C) Monozygotic (MZ) twins only with GEE, (D) MZ twins with CGEE, (E) Discordant MZ twins with GEE, and (F) Discordant MZ twins with CGEE. Only metabolites showing significant differences are labeled with FDR-adjusted *p*-values. The detailed estimates obtained from the models are presented in Supplementary Table S8.

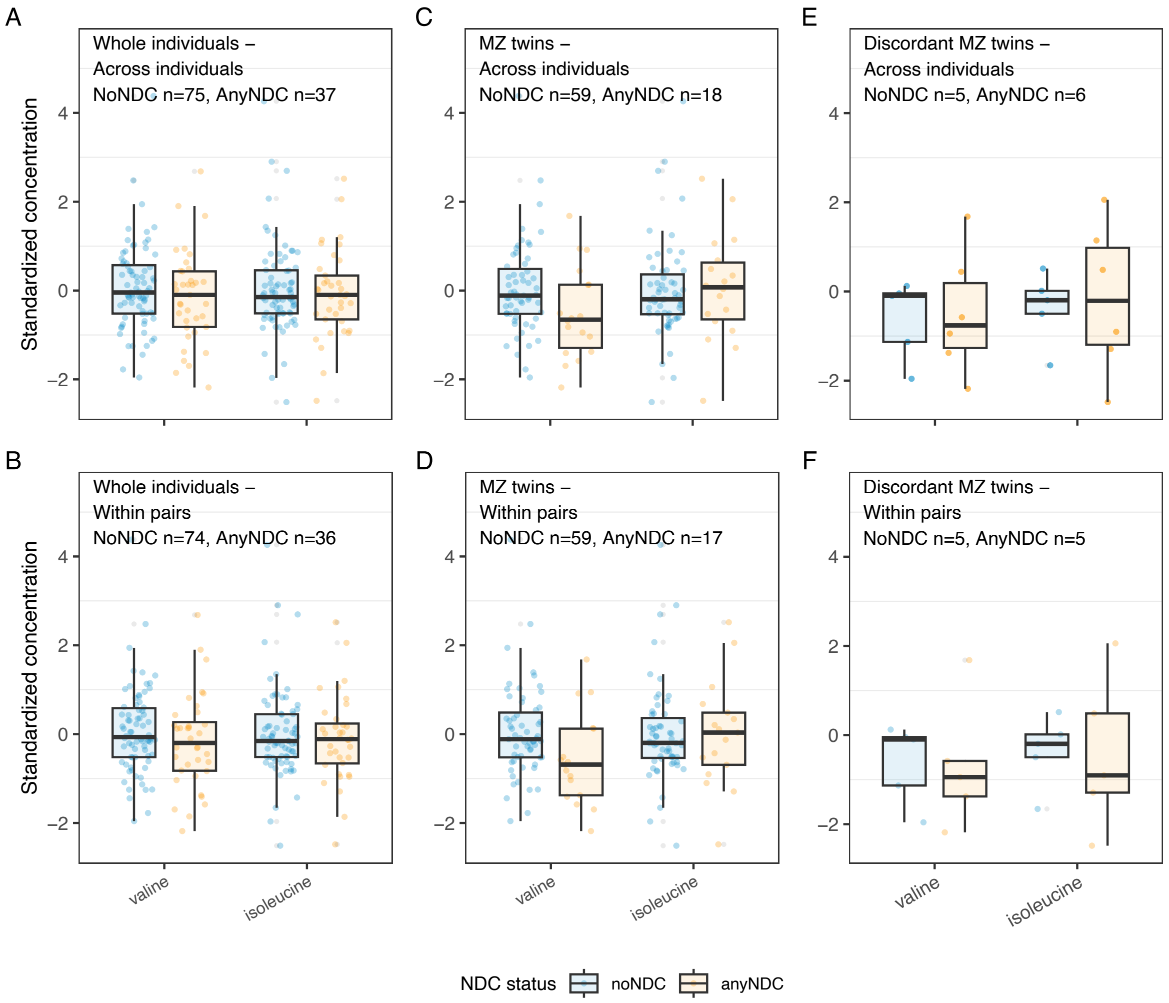


**Fig. S10 Group-wise distribution of selected urine metabolites across subsamples used in differentiation analyses.** Plots show metabolite abundance across diagnostic groups (anyNDC vs. noNDC) in various analytic subsets: (A) All individuals using a cross individuals generalized estimating equation (GEE) model, (B) All individuals using a within-pair conditional GEE (CGEE) model, (C) Monozygotic (MZ) twins only with GEE, (D) MZ twins with CGEE, (E) Discordant MZ twins with GEE, and (F) Discordant MZ twins with CGEE. No significant differences were observed between noNDC and anyNDC in any comparisons. The detailed estimates and *p*-values obtained from the models are presented in Supplementary Table S9.

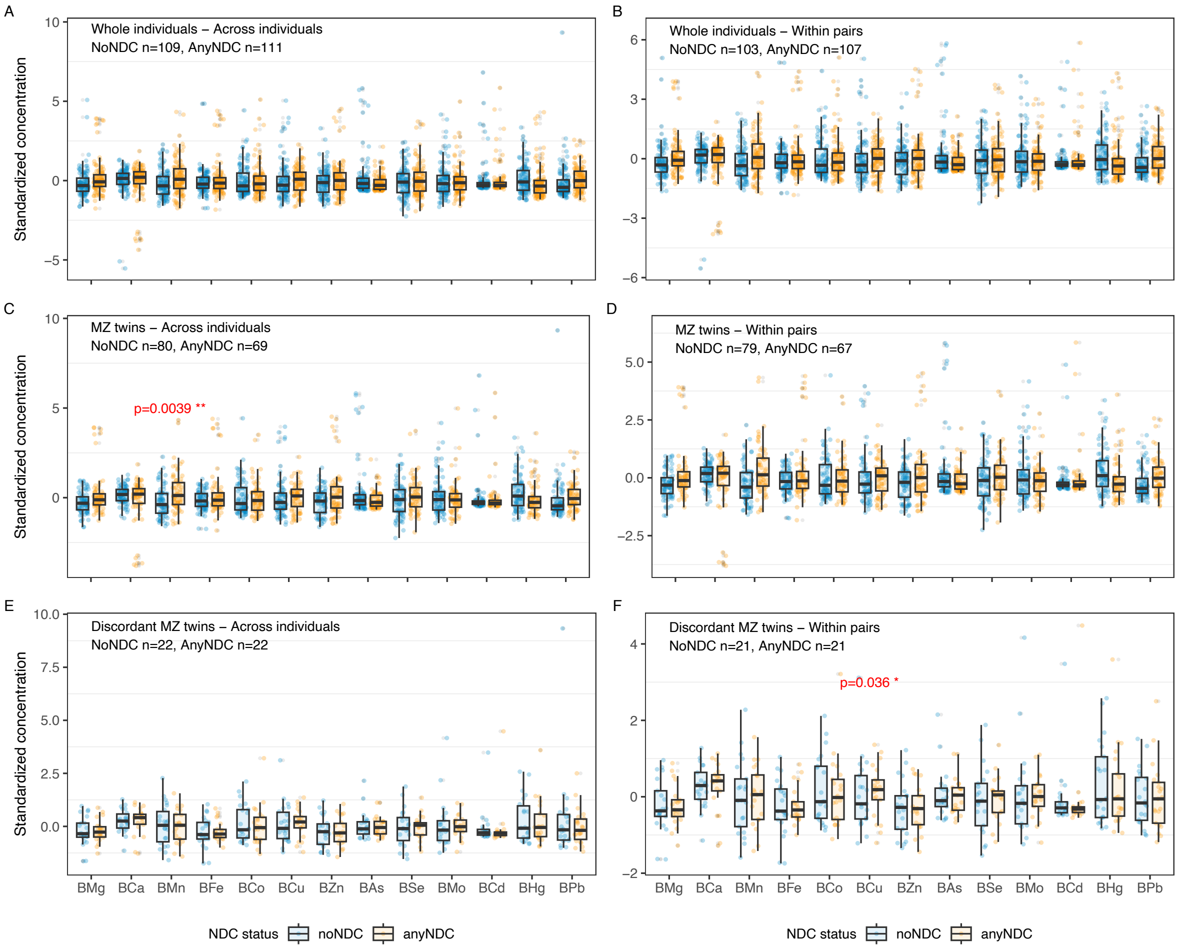


**Fig. S11 Group-wise distribution of metal ions across subsamples used in differentiation analyses.** Plots show metal ions abundance across diagnostic groups (anyNDC vs. noNDC) in various analytic subsets: (A) All individuals using a cross individuals generalized estimating equation (GEE) model, (B) All individuals using a within-pair conditional GEE (CGEE) model, (C) Monozygotic (MZ) twins only with GEE, (D) MZ twins with CGEE, (E) Discordant MZ twins with GEE, and (F) Discordant MZ twins with CGEE. Only metal ions showing significant differences are labeled with FDR-adjusted *p*-values. The detailed estimates obtained from the models are presented in Supplementary Table S10.

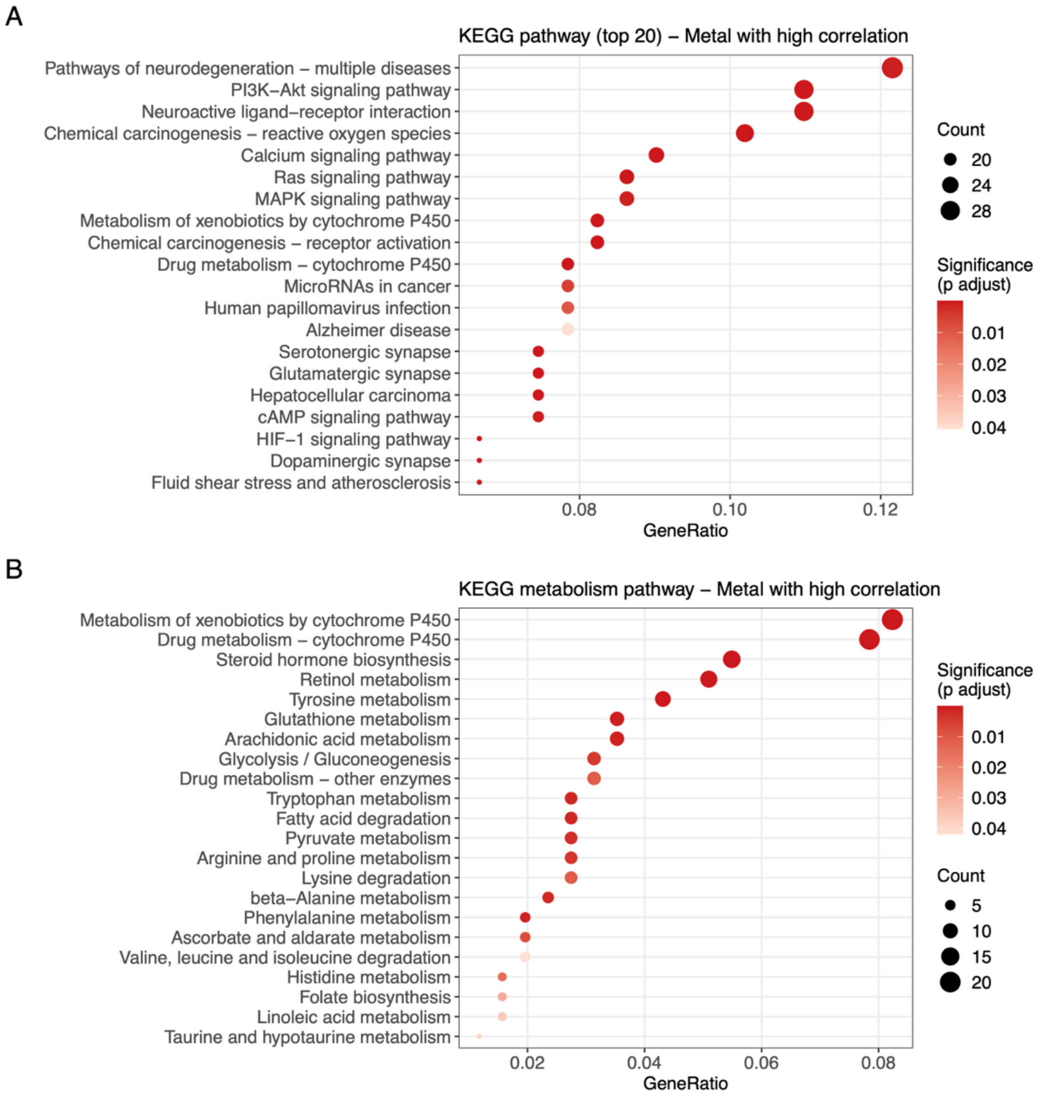


**Fig. S12 Dot plot of enriched KEGG pathways for metal- and NDC-associated gene sets.** A. The top 25 enriched pathways. B. The top enriched metabolism pathways. The full results of enriched pathways are listed in Supplementary Table S12.

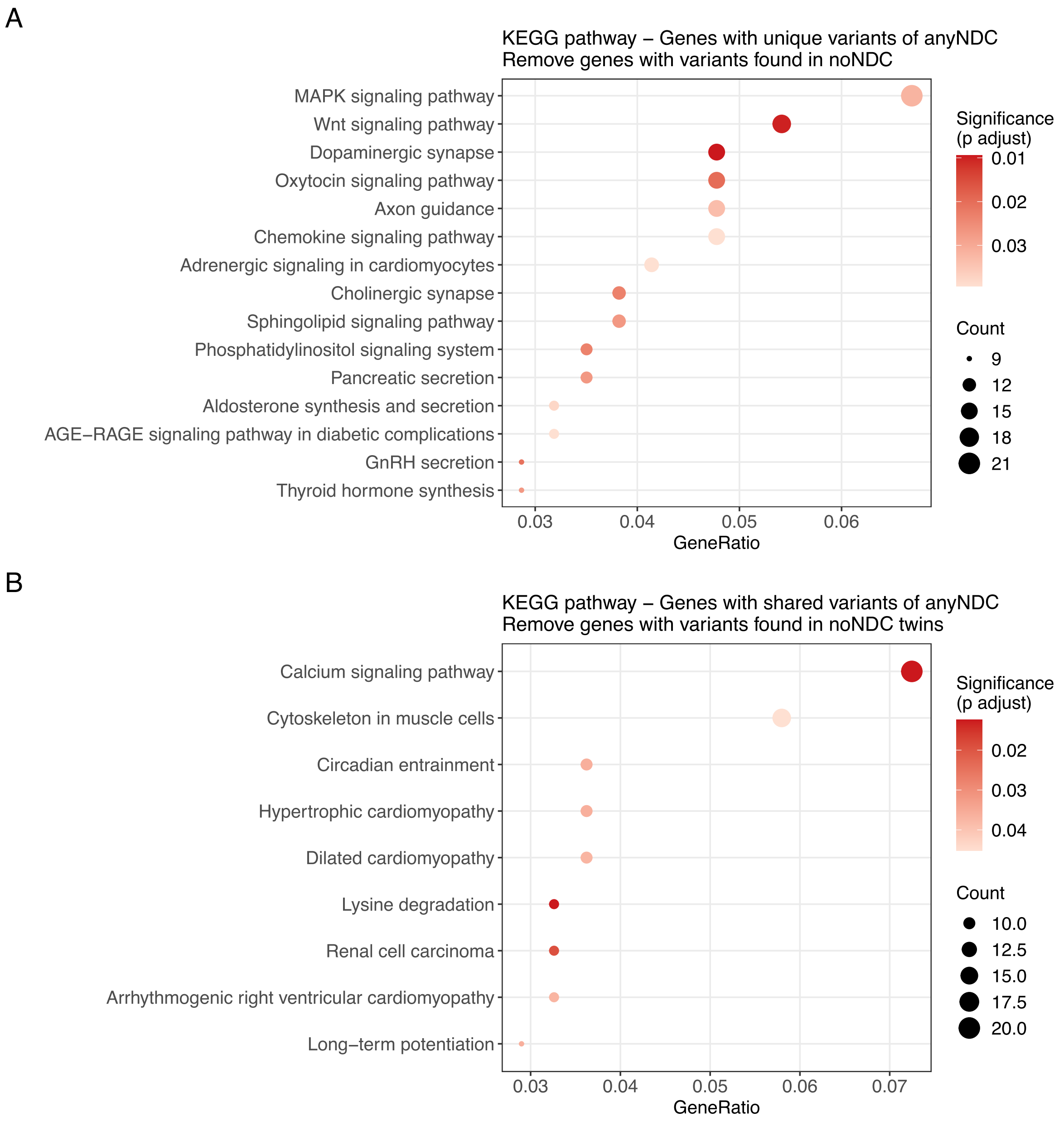


**Fig. S13 Dot plot of enriched KEGG pathways based on genes harboring rare variants.** A. Enriched pathways based on genes carrying rare variants uniquely present in NDC individuals, excluding any genes with unique variants observed in non-NDC individuals. B. Enriched pathways based on NDC genes harboring variants shared among concordant and discordant NDC twin pairs, excluding genes with any shared variants identified in concordant non-NDC twins. The full results of enriched pathways are listed in Supplementary Tables S13-14.

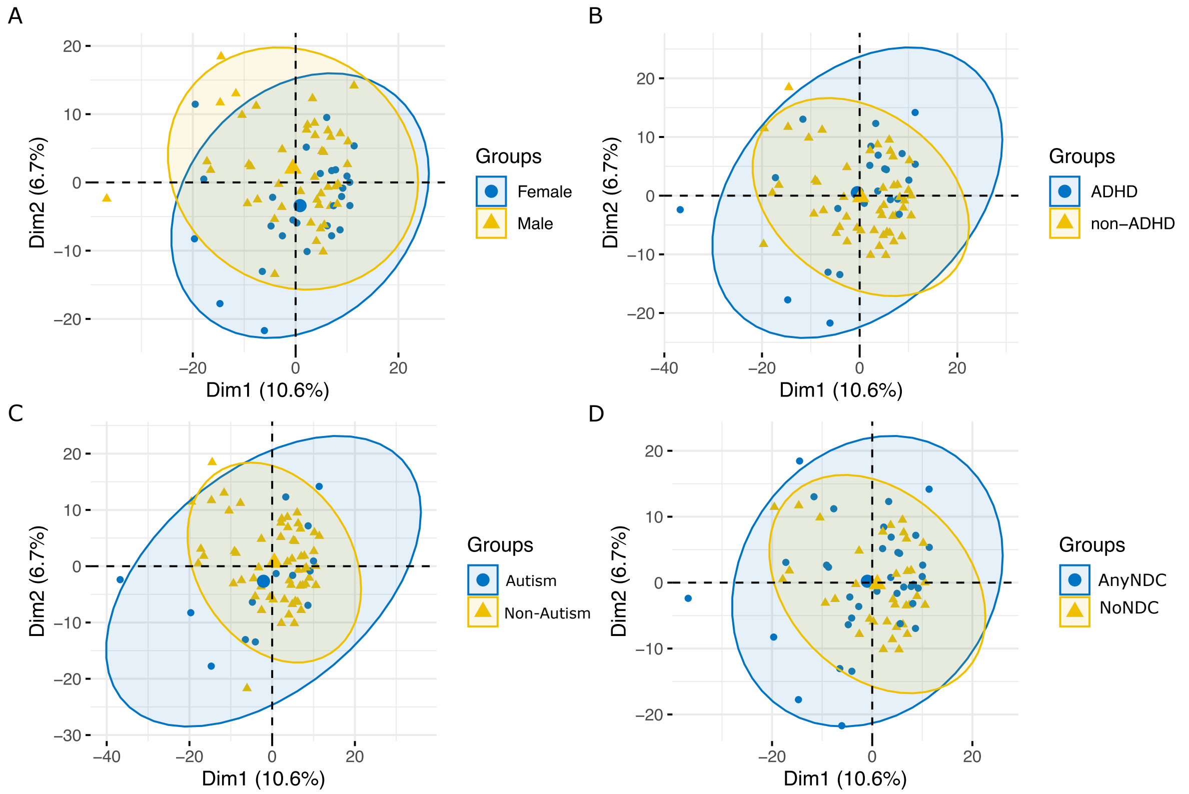


**Fig. S14 Principal component analysis of individuals overlapped in serum-proteomics, CSF-proteomics, fecal-metabolomics, and blood-metallomics datasets using concatenated omics features.** Scatter plots are grouped as per sex (A), ADHD diagnosis (B), autism diagnosis (C), and anyNDC diagnosis status (D).

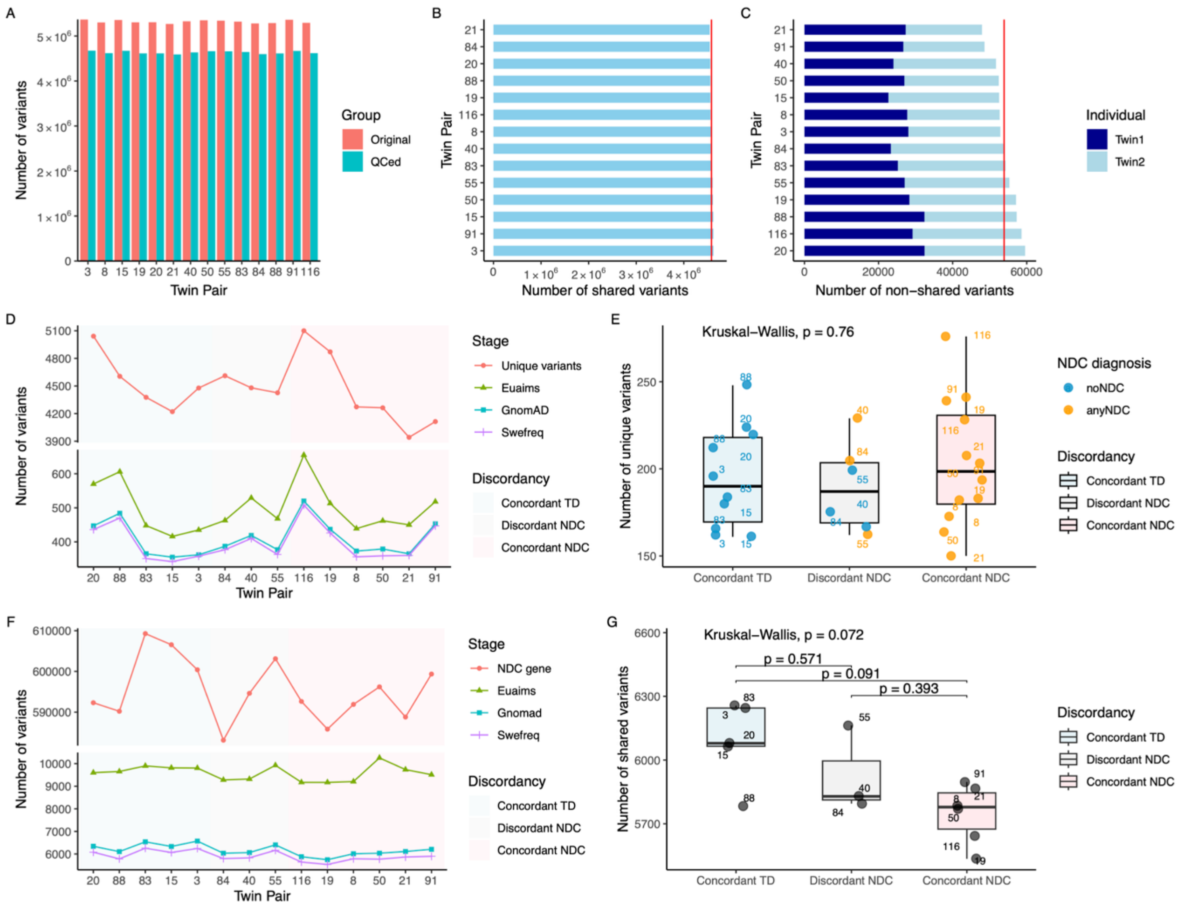


**Fig. S15 Number of unique and shared variants identified in monozygotic twins.** A. The number of variants in each twin pair before and after quality control. B. The number of shared variants in each twin pair after quality control. The red line indicates the average of all twin pairs. C. The number of unique variants in each individual within each twin pair. The red line indicates the average of all twin pairs. D. The number of unique variants retained after each round of population filtering (variants with AF > 0.001 were removed) using EU-AIMS AF, gnomAD MAX AF, and SweGen AF. E. The number of rare unique variants (AF < 0.001) in each individual. The significance of differences between three groups (concordant TD, discordant NDC, and concordant NDC) was assessed using the Kruskal-Wallis test. F. The number of shared variants in NDC genes retained in each twin pair after each round of population filtering (variants with AF > 0.01 were removed) using EU-AIMS AF, gnomAD MAX AF, and SweGen AF. G. The number of shared rare variants (AF < 0.01) in each twin pair. The significance of differences between three groups (concordant TD, discordant NDC, and concordant NDC) was assessed using the Kruskal-Wallis test, with pairwise comparisons performed using the Wilcoxon rank sum test and Benjamini–Hochberg false discovery rate correction.
