## Supplementary Tables for "Integrative Multi-Omics Analysis of Molecular Associations and Biological Pathways Underlying Neurodevelopmental Conditions in a Twin Cohort"

**This file includes Supplementary Table S1-S17:**

Table S1. Demographics of study population and data availability.
Table S2. Study population characteristics and descriptive statistics for each omics dataset.
Table S3. The parameters tunning in single omics and multi-omics models.
Table S4. GEE model estimates of covariate associations with DIABLO component scores of MultiOmic-SCFB model.
Table S5. Joint pathway enrichment analysis of key proteomic and metabolomic features in all pathways.
Table S6. Joint pathway enrichment analysis of key proteomic and metabolomic features in metabolic pathways.
Table S7. The association between ADA protein levels and anyNDC status evaluated by GEE model.
Table S8. The association between levels of key fecal metabolite and anyNDC status evaluated by GEE model.
Table S9. The association between levels of key urine metabolite and anyNDC status evaluated by GEE model.
Table S10. The association between metal levels and anyNDC status evaluated by GEE model.
Table S11. Differential analysis for NDC subgroups.
Table S12. Pathway enrichment analysis of metal-NDC-related genes.
Table S13. Pathway enrichment analysis of genes harboring rare unique NDC-specific variants.
Table S14. Pathway enrichment analysis of genes harboring rare shared variants in concordant and discordant NDC twin pairs, excluding genes with rare variants shared in concordant TD twin pairs.
Table S15. Quality control of toxic and essential metals in whole blood.
Table S16 The feature selection stability assessment for multiomics models.
Table S17. Missingness of key molecules identified in multiomics models.

**Table S1. Demographics of study population and data availability.** This table provides a individual-level overview of biospecimen availability, clinical diagnoses, and demographic metadata. Binary encoding is utilized where "1" denotes available data or the presence of a clinical condition, and "0" denotes missing data or the absence of the condition. Abbreviations: ADHD, Attention-Deficit/Hyperactivity Disorder; CSF, cerebrospinal fluid; DZ, dizygotic; ID, Intellectual Disability; MZ, monozygotic; NDC, neurodevelopmental condition; TD, typical development.

| Individual n=237 | Blood n=220 | Fecal n=189 | Serum n=126 | CSF n=86 | Urine n=112 | Euaims n=28 | Autism n=61 | ADHD n=76 | ID n=19 | anyNDC n=122 | Psychiatric n=47 | Typical development n=104 | Zygosity MZ=157 DZ=80 | Discordancy | Sex Male=133 Female=104 | Age range (years) | BMI | Medication n=83 | Family cod | Twin code | Complete twin n=116 |
|---|---|---|---|---|---|---|---|---|---|---|---|---|---|---|---|---|---|---|---|---|---|
| 1 | 1 | 1 | 1 | 1 | 1 | 0 | 0 | 0 | 0 | 0 | 0 | 1 | MZ | Concordant TD | Female | 18-22 | 19,26 | 0 | 1 | 1 | 1 |
| 2 | 1 | 1 | 1 | 0 | 1 | 0 | 0 | 0 | 0 | 0 | 0 | 1 | MZ | Concordant TD | Female | 18-22 | 21,5 | 0 | 1 | 1 | 1 |
| 3 | 1 | 1 | 1 | 0 | 1 | 0 | 0 | 0 | 0 | 0 | 0 | 1 | MZ | Concordant TD | Female | 13-17 | 19,84 | 0 | 2 | 2 | 1 |
| 4 | 1 | 1 | 1 | 0 | 1 | 0 | 0 | 0 | 0 | 0 | 0 | 1 | MZ | Concordant TD | Female | 13-17 | 19,86 | 0 | 2 | 2 | 1 |
| 5 | 1 | 1 | 1 | 1 | 1 | 1 | 0 | 0 | 0 | 0 | 0 | 1 | MZ | Concordant TD | Female | 13-17 | 18,98 | 0 | 3 | 3 | 1 |
| 6 | 1 | 1 | 1 | 1 | 1 | 1 | 0 | 0 | 0 | 0 | 0 | 1 | MZ | Concordant TD | Female | 13-17 | 19,77 | 1 | 3 | 3 | 1 |
| 7 | 1 | 1 | 1 | 0 | 0 | 0 | 1 | 0 | 0 | 1 | 0 | 0 | MZ | Discordant NDC | Female | 13-17 | 26,58 | 0 | 4 | 4 | 1 |
| 8 | 1 | 1 | 1 | 0 | 0 | 0 | 0 | 0 | 0 | 0 | 0 | 1 | MZ | Discordant NDC | Female | 13-17 | 27,14 | 0 | 4 | 4 | 1 |
| 9 | 1 | 1 | 1 | 1 | 1 | 0 | 0 | 0 | 0 | 0 | 0 | 1 | MZ | Discordant NDC | Male | 8-12 | 24,96 | 0 | 5 | 5 | 1 |
| 10 | 1 | 1 | 1 | 1 | 1 | 0 | 0 | 1 | 0 | 1 | 0 | 0 | MZ | Discordant NDC | Male | 8-12 | 23,91 | 0 | 5 | 5 | 1 |
| 11 | 1 | 1 | 1 | 0 | 1 | 0 | 0 | 1 | 0 | 1 | 0 | 0 | MZ | Concordant NDC | Female | 8-12 | 19,72 | 0 | 6 | 6 | 1 |
| 12 | 1 | 1 | 1 | 0 | 1 | 0 | 0 | 1 | 0 | 1 | 0 | 0 | MZ | Concordant NDC | Female | 8-12 | 21,13 | 0 | 6 | 6 | 1 |
| 13 | 1 | 1 | 1 | 0 | 1 | 0 | 0 | 1 | 0 | 1 | 0 | 0 | MZ | Concordant NDC | Male | 13-17 | 15,7 | 0 | 7 | 7 | 1 |
| 14 | 1 | 1 | 1 | 0 | 1 | 0 | 0 | 1 | 0 | 1 | 0 | 0 | MZ | Concordant NDC | Male | 13-17 | 15,09 | 0 | 7 | 7 | 1 |
| 15 | 1 | 1 | 1 | 1 | 0 | 1 | 0 | 1 | 1 | 1 | 0 | 0 | MZ | Concordant NDC | Male | 13-17 | 20,73 | 1 | 8 | 8 | 1 |
| 16 | 1 | 1 | 1 | 1 | 0 | 1 | 1 | 1 | 1 | 1 | 0 | 0 | MZ | Concordant NDC | Male | 13-17 | 23,53 | 1 | 8 | 8 | 1 |
| 17 | 1 | 0 | 1 | 1 | 0 | 0 | 1 | 1 | 0 | 1 | 0 | 0 | MZ | Concordant NDC | Male | 8-12 | 19,09 | 1 | 9 | 9 | 1 |
| 18 | 1 | 1 | 1 | 1 | 0 | 0 | 0 | 1 | 0 | 1 | 0 | 0 | MZ | Concordant NDC | Male | 8-12 | 18,86 | 0 | 9 | 9 | 1 |
| 19 | 1 | 0 | 1 | 1 | 0 | 0 | 0 | 0 | 0 | 1 | 1 | 0 | MZ | Concordant NDC | Male | 13-17 | 20,11 | 1 | 10 | 10 | 1 |
| 20 | 1 | 0 | 1 | 1 | 0 | 0 | 1 | 0 | 0 | 1 | 1 | 0 | MZ | Concordant NDC | Male | 13-17 | 18,96 | 1 | 10 | 10 | 1 |
| 21 | 1 | 1 | 1 | 0 | 1 | 0 | 0 | 0 | 0 | 0 | 0 | 1 | MZ | Concordant TD | Female | 13-17 | 27,32 | 0 | 11 | 11 | 1 |
| 22 | 1 | 1 | 1 | 0 | 1 | 0 | 0 | 0 | 0 | 0 | 0 | 1 | MZ | Concordant TD | Female | 13-17 | 23,11 | 0 | 11 | 11 | 1 |
| 23 | 1 | 1 | 1 | 0 | 1 | 0 | 0 | 0 | 0 | 0 | 0 | 1 | MZ | Discordant NDC | Female | 13-17 | 20,03 | 0 | 12 | 12 | 1 |
| 24 | 1 | 1 | 1 | 0 | 1 | 0 | 0 | 1 | 0 | 1 | 0 | 0 | MZ | Discordant NDC | Female | 13-17 | 18,76 | 1 | 12 | 12 | 1 |
| 25 | 1 | 1 | 1 | 0 | 1 | 0 | 0 | 0 | 0 | 0 | 0 | 1 | MZ | Concordant TD | Male | 8-12 | 26,23 | 0 | 13 | 13 | 1 |
| 26 | 1 | 1 | 1 | 0 | 1 | 0 | 0 | 0 | 0 | 0 | 0 | 1 | MZ | Concordant TD | Male | 8-12 | 25,33 | 0 | 13 | 13 | 1 |
| 27 | 1 | 0 | 1 | 1 | 1 | 0 | 0 | 0 | 0 | 0 | 0 | 1 | MZ | Concordant TD | Female | 13-17 | 17,82 | 0 | 14 | 14 | 1 |
| 28 | 1 | 1 | 1 | 1 | 1 | 0 | 0 | 0 | 0 | 0 | 0 | 1 | MZ | Concordant TD | Female | 13-17 | 18,29 | 0 | 14 | 14 | 1 |
| 29 | 1 | 1 | 1 | 1 | 1 | 1 | 0 | 0 | 0 | 0 | 0 | 1 | MZ | Concordant TD | Female | 13-17 | 21,54 | 0 | 15 | 15 | 1 |
| 30 | 1 | 1 | 1 | 1 | 1 | 1 | 0 | 0 | 0 | 0 | 0 | 1 | MZ | Concordant TD | Female | 13-17 | 20,5 | 0 | 15 | 15 | 1 |
| 31 | 1 | 1 | 1 | 1 | 0 | 0 | 0 | 1 | 0 | 1 | 0 | 0 | MZ | Concordant NDC | Male | 13-17 | 26,12 | 1 | 16 | 16 | 1 |
| 32 | 1 | 1 | 1 | 1 | 0 | 0 | 0 | 1 | 1 | 1 | 0 | 0 | MZ | Concordant NDC | Male | 13-17 | 28,89 | 1 | 16 | 16 | 1 |
| 33 | 1 | 1 | 1 | 1 | 1 | 0 | 0 | 0 | 0 | 0 | 0 | 1 | MZ | Concordant TD | Male | 18-22 | 21,59 | 0 | 17 | 17 | 1 |
| 34 | 1 | 1 | 1 | 1 | 1 | 0 | 0 | 0 | 0 | 0 | 0 | 1 | MZ | Concordant TD | Male | 18-22 | 20,87 | 0 | 17 | 17 | 1 |
| 35 | 1 | 1 | 1 | 1 | 1 | 0 | 0 | 1 | 0 | 1 | 0 | 0 | MZ | Concordant NDC | Male | 13-17 | 23,9 | 1 | 18 | 18 | 1 |
| 36 | 1 | 1 | 1 | 1 | 1 | 0 | 0 | 1 | 0 | 1 | 1 | 0 | MZ | Concordant NDC | Male | 13-17 | 20,74 | 1 | 18 | 18 | 1 |
| 37 | 1 | 1 | 1 | 1 | 0 | 1 | 1 | 1 | 0 | 1 | 1 | 0 | MZ | Concordant NDC | Male | 13-17 | 22,97 | 1 | 19 | 19 | 1 |
| 38 | 1 | 1 | 1 | 1 | 0 | 1 | 1 | 1 | 0 | 1 | 1 | 0 | MZ | Concordant NDC | Male | 13-17 | 21,87 | 1 | 19 | 19 | 1 |
| 39 | 1 | 1 | 1 | 1 | 1 | 1 | 0 | 0 | 0 | 0 | 0 | 1 | MZ | Concordant TD | Male | 13-17 | 19,6 | 0 | 20 | 20 | 1 |
| 40 | 1 | 1 | 1 | 1 | 1 | 1 | 0 | 0 | 0 | 0 | 0 | 1 | MZ | Concordant TD | Male | 13-17 | 19,24 | 0 | 20 | 20 | 1 |
| 41 | 1 | 1 | 1 | 1 | 0 | 1 | 1 | 1 | 0 | 1 | 1 | 0 | MZ | Concordant NDC | Male | 8-12 | 20,68 | 1 | 21 | 21 | 1 |
| 42 | 1 | 1 | 1 | 0 | 0 | 1 | 1 | 1 | 1 | 1 | 0 | 0 | MZ | Concordant NDC | Male | 8-12 | 22,92 | 1 | 21 | 21 | 1 |
| 43 | 1 | 1 | 1 | 0 | 0 | 0 | 0 | 0 | 1 | 1 | 0 | 0 | MZ | Concordant NDC | Male | 18-22 | 20,3 | 0 | 22 | 22 | 1 |
| 44 | 1 | 1 | 1 | 1 | 0 | 0 | 0 | 0 | 1 | 1 | 0 | 0 | MZ | Concordant NDC | Male | 18-22 | 19,26 | 0 | 22 | 22 | 1 |
| 45 | 1 | 1 | 1 | 0 | 0 | 0 | 0 | 0 | 0 | 0 | 0 | 1 | MZ | Discordant NDC | Female | 13-17 | 22,39 | 0 | 23 | 23 | 1 |
| 46 | 1 | 1 | 1 | 0 | 0 | 0 | 1 | 0 | 0 | 1 | 0 | 0 | MZ | Discordant NDC | Female | 13-17 | 16,37 | 0 | 23 | 23 | 1 |
| 47 | 1 | 1 | 1 | 1 | 0 | 0 | 0 | 0 | 0 | 0 | 0 | 1 | MZ | Discordant NDC | Male | 13-17 | 26,78 | 0 | 24 | 24 | 1 |
| 48 | 1 | 1 | 1 | 1 | 0 | 0 | 0 | 0 | 0 | 1 | 0 | 0 | MZ | Discordant NDC | Male | 13-17 | 21,82 | 0 | 24 | 24 | 1 |
| 49 | 1 | 1 | 1 | 0 | 1 | 0 | 0 | 0 | 0 | 0 | 0 | 1 | MZ | Concordant TD | Female | 13-17 | 21,11 | 0 | 25 | 25 | 1 |
| 50 | 1 | 1 | 1 | 0 | 1 | 0 | 0 | 0 | 0 | 0 | 0 | 1 | MZ | Concordant TD | Female | 13-17 | 21,08 | 0 | 25 | 25 | 1 |
| 51 | 1 | 1 | 1 | 1 | 0 | 0 | 1 | 0 | 1 | 1 | 0 | 0 | MZ | Concordant NDC | Female | 13-17 | 18,4 | 0 | 27 | 27 | 1 |
| 52 | 1 | 1 | 1 | 1 | 0 | 0 | 1 | 0 | 0 | 1 | 0 | 0 | MZ | Concordant NDC | Female | 13-17 | 18,87 | 0 | 27 | 27 | 1 |
| 53 | 1 | 1 | 1 | 0 | 0 | 0 | 1 | 0 | 0 | 1 | 1 | 0 | MZ | Discordant NDC | Male | 18-22 | 19,79 | 1 | 28 | 28 | 1 |
| 54 | 1 | 1 | 1 | 1 | 1 | 0 | 0 | 0 | 0 | 0 | 0 | 1 | MZ | Concordant TD | Male | 18-22 | 19,59 | 0 | 29 | 29 | 1 |
| 55 | 1 | 1 | 1 | 1 | 1 | 0 | 0 | 0 | 0 | 0 | 0 | 1 | MZ | Concordant TD | Male | 18-22 | 19,84 | 0 | 29 | 29 | 1 |
| 56 | 1 | 1 | 1 | 1 | 1 | 0 | 0 | 0 | 0 | 0 | 0 | 1 | DZ | Concordant TD | Male | 13-17 | 17,74 | 0 | 30 | 30 | 1 |
| 57 | 1 | 1 | 1 | 1 | 1 | 0 | 0 | 0 | 0 | 0 | 0 | 1 | DZ | Concordant TD | Female | 13-17 | 23,46 | 0 | 30 | 30 | 1 |
| 58 | 1 | 1 | 1 | 1 | 1 | 0 | 0 | 0 | 0 | 1 | 0 | 0 | DZ | Concordant NDC | Male | 18-22 | 21,87 | 1 | 31 | 31 | 1 |
| 59 | 1 | 1 | 1 | 1 | 1 | 0 | 0 | 1 | 0 | 1 | 0 | 0 | DZ | Concordant NDC | Male | 18-22 | 21,45 | 1 | 31 | 31 | 1 |
| 60 | 1 | 1 | 1 | 0 | 0 | 0 | 1 | 0 | 1 | 1 | 1 | 0 | MZ | Concordant NDC | Male | 8-12 | 22,27 | 1 | 32 | 32 | 1 |
| 61 | 1 | 1 | 1 | 1 | 0 | 0 | 0 | 0 | 0 | 0 | 0 | 1 | DZ | Discordant NDC | Male | 13-17 | 18,82 | 0 | 33 | 33 | 1 |
| 62 | 1 | 1 | 1 | 1 | 0 | 0 | 1 | 1 | 0 | 1 | 0 | 0 | DZ | Discordant NDC | Male | 13-17 | 18,31 | 1 | 33 | 33 | 1 |
| 63 | 1 | 1 | 1 | 1 | 1 | 0 | 0 | 0 | 0 | 0 | 0 | 1 | DZ | Discordant NDC | Female | 13-17 | 20,78 | 1 | 34 | 34 | 1 |
| 64 | 1 | 1 | 1 | 1 | 1 | 0 | 0 | 1 | 0 | 1 | 0 | 0 | DZ | Discordant NDC | Female | 13-17 | 18,02 | 0 | 34 | 34 | 1 |
| 65 | 1 | 0 | 1 | 1 | 1 | 0 | 0 | 1 | 0 | 1 | 1 | 0 | DZ | Concordant NDC | Male | 18-22 | 25,1 | 0 | 36 | 36 | 1 |
| 66 | 1 | 1 | 1 | 1 | 1 | 0 | 0 | 0 | 0 | 1 | 1 | 0 | DZ | Concordant NDC | Male | 18-22 | 26,27 | 0 | 36 | 36 | 1 |
| 67 | 1 | 0 | 1 | 0 | 0 | 0 | 1 | 0 | 1 | 1 | 0 | 0 | DZ | Discordant NDC | Male | 13-17 | NA | 1 | 37 | 37 | 1 |
| 68 | 1 | 0 | 1 | 1 | 0 | 0 | 0 | 0 | 0 | 0 | 0 | 1 | DZ | Discordant NDC | Male | 13-17 | 18,12 | 0 | 37 | 37 | 1 |
| 69 | 1 | 1 | 1 | 0 | 1 | 0 | 0 | 1 | 0 | 1 | 1 | 0 | DZ | Concordant NDC | Female | 8-12 | 15,16 | 1 | 39 | 39 | 1 |
| 70 | 1 | 1 | 1 | 0 | 1 | 0 | 0 | 1 | 0 | 1 | 1 | 0 | DZ | Concordant NDC | Female | 8-12 | 16,39 | 1 | 39 | 39 | 1 |
| 71 | 1 | 1 | 1 | 0 | 0 | 1 | 1 | 0 | 0 | 1 | 0 | 0 | MZ | Discordant NDC | Male | 8-12 | 17,09 | 0 | 40 | 40 | 1 |
| 72 | 1 | 1 | 1 | 1 | 0 | 1 | 0 | 0 | 0 | 0 | 0 | 1 | MZ | Discordant NDC | Male | 8-12 | 18,88 | 0 | 40 | 40 | 1 |
| 73 | 1 | 1 | 1 | 0 | 0 | 0 | 1 | 1 | 0 | 1 | 0 | 0 | MZ | Discordant NDC | Male | 8-12 | 19,2 | 1 | 41 | 41 | 1 |
| 74 | 1 | 1 | 1 | 0 | 0 | 0 | 0 | 0 | 0 | 0 | 0 | 1 | MZ | Discordant NDC | Male | 8-12 | 22,49 | 0 | 41 | 41 | 1 |
| 75 | 1 | 1 | 1 | 1 | 0 | 0 | 0 | 0 | 0 | 0 | 0 | 1 | DZ | Discordant NDC | Female | 13-17 | 21,7 | 0 | 42 | 42 | 1 |
| 76 | 1 | 1 | 1 | 1 | 0 | 0 | 1 | 0 | 0 | 1 | 1 | 0 | DZ | Discordant NDC | Female | 8-12 | 22,82 | 0 | 44 | 44 | 1 |
| 77 | 1 | 1 | 1 | 1 | 0 | 0 | 0 | 0 | 0 | 0 | 0 | 1 | DZ | Discordant NDC | Male | 8-12 | 17,08 | 0 | 44 | 44 | 1 |
| 78 | 1 | 1 | 1 | 1 | 1 | 0 | 0 | 0 | 0 | 1 | 0 | 0 | MZ | Concordant NDC | Male | 13-17 | 22,91 | 1 | 45 | 45 | 1 |
| 79 | 1 | 1 | 1 | 1 | 1 | 0 | 0 | 1 | 0 | 1 | 1 | 0 | MZ | Concordant NDC | Male | 13-17 | 21,56 | 1 | 45 | 45 | 1 |
| 80 | 1 | 1 | 1 | 1 | 0 | 0 | 0 | 0 | 0 | 0 | 0 | 1 | MZ | Discordant NDC | Female | 18-22 | 23,81 | 0 | 46 | 46 | 1 |
| 81 | 1 | 1 | 1 | 1 | 1 | 0 | 0 | 0 | 0 | 0 | 0 | 1 | MZ | Concordant TD | Male | 13-17 | 22,81 | 0 | 48 | 48 | 1 |
| 82 | 1 | 1 | 1 | 1 | 1 | 0 | 0 | 0 | 0 | 0 | 0 | 1 | MZ | Concordant TD | Male | 13-17 | 21,58 | 0 | 48 | 48 | 1 |
| 83 | 1 | 1 | 1 | 1 | 1 | 0 | 0 | 1 | 0 | 1 | 0 | 0 | MZ | Concordant NDC | Male | 8-12 | 16,19 | 1 | 49 | 49 | 1 |
| 84 | 1 | 1 | 1 | 1 | 1 | 0 | 0 | 1 | 0 | 1 | 0 | 0 | MZ | Concordant NDC | Male | 8-12 | 16,1 | 1 | 49 | 49 | 1 |
| 85 | 1 | 1 | 1 | 1 | 0 | 1 | 1 | 1 | 0 | 1 | 0 | 0 | MZ | Concordant NDC | Female | 8-12 | 17,93 | 1 | 50 | 50 | 1 |
| 86 | 1 | 1 | 1 | 1 | 0 | 1 | 1 | 1 | 0 | 1 | 0 | 0 | MZ | Concordant NDC | Female | 8-12 | 21,78 | 1 | 50 | 50 | 1 |
| 87 | 1 | 0 | 1 | 1 | 1 | 0 | 0 | 0 | 0 | 0 | 0 | 1 | MZ | Concordant TD | Female | 13-17 | 19,16 | 0 | 51 | 51 | 1 |
| 88 | 1 | 0 | 1 | 1 | 1 | 0 | 0 | 0 | 0 | 0 | 0 | 1 | MZ | Concordant TD | Female | 13-17 | 19,49 | 0 | 51 | 51 | 1 |
| 89 | 1 | 1 | 1 | 1 | 1 | 0 | 0 | 0 | 0 | 0 | 1 | 0 | MZ | Discordant NDC | Female | 8-12 | 15,04 | 0 | 53 | 53 | 1 |
| 90 | 1 | 1 | 1 | 1 | 1 | 0 | 0 | 1 | 0 | 1 | 0 | 0 | MZ | Discordant NDC | Female | 8-12 | 14,69 | 0 | 53 | 53 | 1 |
| 91 | 1 | 0 | 1 | 1 | 0 | 0 | 1 | 1 | 0 | 1 | 0 | 0 | MZ | Concordant NDC | Female | 23-28 | 28,38 | 1 | 54 | 54 | 1 |
| 92 | 1 | 0 | 1 | 1 | 0 | 0 | 1 | 1 | 0 | 1 | 1 | 0 | MZ | Concordant NDC | Female | 23-28 | 29,02 | 1 | 54 | 54 | 1 |
| 93 | 1 | 1 | 1 | 1 | 0 | 1 | 0 | 0 | 0 | 0 | 0 | 1 | MZ | Discordant NDC | Female | 8-12 | 14,53 | 1 | 55 | 55 | 1 |
| 94 | 1 | 1 | 1 | 0 | 0 | 1 | 1 | 1 | 0 | 1 | 0 | 0 | MZ | Discordant NDC | Female | 8-12 | 14,7 | 0 | 55 | 55 | 1 |
| 95 | 1 | 1 | 1 | 1 | 1 | 0 | 0 | 0 | 0 | 0 | 0 | 1 | MZ | Concordant TD | Male | 8-12 | 14,89 | 0 | 56 | 56 | 1 |
| 96 | 1 | 1 | 1 | 1 | 1 | 0 | 0 | 0 | 0 | 0 | 0 | 1 | MZ | Concordant TD | Male | 8-12 | 14,32 | 0 | 56 | 56 | 1 |
| 97 | 1 | 1 | 1 | 1 | 1 | 0 | 0 | 0 | 0 | 0 | 0 | 1 | MZ | Concordant TD | Female | 8-12 | 17,85 | 0 | 58 | 58 | 1 |
| 98 | 1 | 1 | 1 | 1 | 1 | 0 | 0 | 0 | 0 | 0 | 0 | 1 | MZ | Concordant TD | Female | 8-12 | 17,85 | 1 | 58 | 58 | 1 |
| 99 | 1 | 1 | 1 | 1 | 0 | 0 | 0 | 0 | 0 | 1 | 0 | 0 | MZ | Concordant NDC | Male | 8-12 | 20,65 | 0 | 59 | 59 | 1 |
| 100 | 1 | 1 | 1 | 1 | 0 | 0 | 0 | 0 | 0 | 1 | 0 | 0 | MZ | Concordant NDC | Male | 8-12 | 20,58 | 0 | 59 | 59 | 1 |
| 101 | 1 | 1 | 1 | 1 | 1 | 0 | 0 | 0 | 0 | 0 | 0 | 1 | MZ | Concordant TD | Male | 13-17 | 21,3 | 0 | 60 | 60 | 1 |
| 102 | 1 | 1 | 1 | 1 | 1 | 0 | 0 | 0 | 0 | 0 | 0 | 1 | MZ | Concordant TD | Male | 13-17 | 21,17 | 0 | 60 | 60 | 1 |
| 103 | 1 | 1 | 1 | 0 | 0 | 0 | 0 | 0 | 0 | 1 | 0 | 0 | MZ | Discordant NDC | Female | 13-17 | 17,59 | 0 | 61 | 61 | 1 |
| 104 | 1 | 1 | 1 | 0 | 0 | 0 | 0 | 0 | 0 | 0 | 0 | 1 | MZ | Discordant NDC | Female | 13-17 | 15,8079 | 0 | 61 | 61 | 1 |
| 105 | 1 | 1 | 1 | 0 | 0 | 0 | 0 | 0 | 1 | 1 | 1 | 0 | MZ | Discordant NDC | Female | 8-12 | 20,87 | 0 | 62 | 62 | 1 |
| 106 | 1 | 1 | 1 | 0 | 0 | 0 | 0 | 0 | 0 | 0 | 0 | 1 | MZ | Discordant NDC | Female | 8-12 | 21,23 | 0 | 62 | 62 | 1 |
| 107 | 1 | 1 | 1 | 1 | 0 | 0 | 1 | 0 | 0 | 1 | 0 | 0 | MZ | Concordant NDC | Male | 8-12 | 14,28 | 0 | 63 | 63 | 1 |
| 108 | 1 | 1 | 1 | 1 | 0 | 0 | 1 | 1 | 0 | 1 | 0 | 0 | MZ | Concordant NDC | Male | 8-12 | 14 | 0 | 63 | 63 | 1 |
| 109 | 1 | 1 | 1 | 0 | 1 | 0 | 0 | 0 | 0 | 0 | 0 | 1 | MZ | Discordant NDC | Male | 8-12 | 15,99 | 0 | 64 | 64 | 1 |
| 110 | 1 | 0 | 1 | 1 | 1 | 0 | 0 | 1 | 0 | 1 | 1 | 0 | MZ | Discordant NDC | Male | 8-12 | 15,52 | 0 | 64 | 64 | 1 |
| 111 | 1 | 1 | 1 | 1 | 0 | 0 | 0 | 0 | 0 | 0 | 0 | 1 | MZ | Discordant NDC | Male | 8-12 | 17,2 | 0 | 65 | 65 | 1 |
| 112 | 1 | 1 | 1 | 1 | 0 | 0 | 0 | 0 | 0 | 1 | 0 | 0 | MZ | Discordant NDC | Male | 8-12 | 17,1 | 0 | 65 | 65 | 1 |
| 113 | 1 | 1 | 1 | 1 | 1 | 0 | 0 | 0 | 0 | 0 | 0 | 1 | MZ | Concordant TD | Female | 8-12 | 18,07 | 0 | 66 | 66 | 1 |
| 114 | 1 | 1 | 1 | 1 | 1 | 0 | 0 | 0 | 0 | 0 | 0 | 1 | MZ | Concordant TD | Female | 8-12 | 17,94 | 0 | 66 | 66 | 1 |
| 115 | 1 | 1 | 1 | 1 | 0 | 0 | 1 | 0 | 0 | 1 | 0 | 0 | MZ | Concordant NDC | Female | 13-17 | 20,33 | 0 | 68 | 68 | 1 |
| 116 | 1 | 1 | 1 | 1 | 0 | 0 | 1 | 0 | 0 | 1 | 0 | 0 | MZ | Concordant NDC | Female | 13-17 | 18,1 | 0 | 68 | 68 | 1 |
| 117 | 1 | 0 | 1 | 0 | 0 | 0 | 1 | 0 | 0 | 1 | 1 | 0 | MZ | Concordant NDC | Female | 23-28 | 19,35 | 0 | 69 | 69 | 1 |
| 118 | 1 | 1 | 1 | 0 | 0 | 0 | 1 | 0 | 0 | 1 | 1 | 0 | MZ | Concordant NDC | Female | 23-28 | 19,18 | 0 | 69 | 69 | 1 |

| | | | | | | | | | | | | | | | | | | | | | |
|---|---|---|---|---|---|---|---|---|---|---|---|---|---|---|---|---|---|---|---|---|---|
| 119 | 1 | 1 | 0 | 0 | 1 | 0 | 0 | 0 | 0 | 0 | 0 | 1 | MZ | Concordant TD | Male | 13-17 | 18,78 | 0 | 70 | 70 | 1 |
| 120 | 1 | 1 | 0 | 0 | 1 | 0 | 0 | 0 | 0 | 0 | 0 | 1 | MZ | Concordant TD | Male | 13-17 | 26,26 | 0 | 70 | 70 | 1 |
| 121 | 1 | 1 | 1 | 1 | 0 | 0 | 0 | 1 | 0 | 1 | 0 | 0 | DZ | Concordant NDC | Female | 8-12 | 20,36 | 0 | 71 | 71 | 1 |
| 122 | 1 | 1 | 1 | 1 | 0 | 0 | 0 | 1 | 0 | 1 | 1 | 0 | DZ | Concordant NDC | Female | 8-12 | 22,15 | 0 | 71 | 71 | 1 |
| 123 | 1 | 1 | 1 | 0 | 1 | 0 | 0 | 0 | 0 | 0 | 0 | 1 | DZ | Concordant TD | Male | 8-12 | 19,73 | 0 | 73 | 73 | 1 |
| 124 | 1 | 1 | 1 | 0 | 1 | 0 | 0 | 0 | 0 | 0 | 0 | 1 | DZ | Concordant TD | Male | 8-12 | 15,76 | 0 | 73 | 73 | 1 |
| 125 | 1 | 1 | 0 | 0 | 1 | 0 | 0 | 0 | 0 | 0 | 0 | 1 | MZ | Discordant NDC | Male | 13-17 | 20,74 | 0 | 74 | 7401 | 1 |
| 126 | 1 | 1 | 0 | 0 | 1 | 0 | 0 | 1 | 0 | 1 | 0 | 0 | MZ | Discordant NDC | Male | 13-17 | 16,65 | 0 | 74 | 7401 | 1 |
| 127 | 1 | 1 | 0 | 0 | 0 | 0 | 0 | 0 | 0 | 0 | 0 | 1 | DZ | Discordant NDC | Male | 13-17 | 19,4392 | 0 | 74 | 7402 | 1 |
| 128 | 1 | 1 | 0 | 0 | 0 | 0 | 1 | 1 | 0 | 1 | 0 | 0 | DZ | Discordant NDC | Male | 13-17 | 22,6003 | 0 | 74 | 7402 | 1 |
| 129 | 1 | 1 | 0 | 0 | 0 | 0 | 0 | 0 | 0 | 0 | 0 | 1 | MZ | Discordant NDC | Female | 13-17 | 24,55 | 1 | 75 | 75 | 1 |
| 130 | 1 | 1 | 0 | 0 | 0 | 0 | 1 | 1 | 1 | 1 | 0 | 0 | MZ | Discordant NDC | Female | 13-17 | 21 | 1 | 75 | 75 | 1 |
| 131 | 1 | 1 | 0 | 0 | 0 | 0 | 1 | 1 | 0 | 1 | 1 | 0 | DZ | Concordant NDC | Male | 8-12 | 15,93 | 1 | 76 | 76 | 1 |
| 132 | 1 | 1 | 0 | 0 | 0 | 0 | 1 | 1 | 0 | 1 | 1 | 0 | DZ | Concordant NDC | Male | 8-12 | 16,84 | 1 | 76 | 76 | 1 |
| 133 | 1 | 1 | 0 | 0 | 1 | 0 | 0 | 1 | 0 | 1 | 1 | 0 | DZ | Concordant NDC | Male | 13-17 | 17,77 | 0 | 77 | 77 | 1 |
| 134 | 1 | 1 | 0 | 0 | 1 | 0 | 0 | 1 | 0 | 1 | 0 | 0 | DZ | Concordant NDC | Male | 13-17 | 23,02 | 0 | 77 | 77 | 1 |
| 135 | 1 | 1 | 0 | 0 | 0 | 0 | 0 | 0 | 0 | 0 | 0 | 1 | DZ | Concordant TD | Male | 18-22 | 22,55 | 0 | 78 | 7801 | 1 |
| 136 | 1 | 1 | 0 | 0 | 0 | 0 | 0 | 0 | 0 | 0 | 1 | 0 | DZ | Concordant TD | Female | 18-22 | 23,38 | 0 | 78 | 7801 | 1 |
| 137 | 1 | 1 | 0 | 0 | 1 | 0 | 0 | 1 | 0 | 1 | 1 | 0 | DZ | Concordant NDC | Female | 18-22 | 22,02 | 1 | 78 | 7802 | 1 |
| 138 | 1 | 1 | 0 | 0 | 1 | 0 | 0 | 1 | 0 | 1 | 0 | 0 | DZ | Concordant NDC | Female | 18-22 | 22,04 | 1 | 78 | 7802 | 1 |
| 139 | 1 | 1 | 0 | 0 | 0 | 0 | 0 | 1 | 0 | 1 | 1 | 0 | DZ | Concordant NDC | Male | 8-12 | 17,57 | 1 | 79 | 79 | 1 |
| 140 | 1 | 1 | 0 | 0 | 0 | 0 | 1 | 0 | 0 | 1 | 1 | 0 | DZ | Concordant NDC | Male | 8-12 | 19,42 | 0 | 79 | 79 | 1 |
| 141 | 1 | 1 | 0 | 0 | 1 | 0 | 0 | 0 | 0 | 0 | 0 | 1 | MZ | Concordant TD | Male | 13-17 | 20,69 | 0 | 80 | 80 | 1 |
| 142 | 1 | 1 | 0 | 0 | 1 | 0 | 0 | 0 | 0 | 0 | 0 | 1 | MZ | Concordant TD | Male | 13-17 | 20,63 | 0 | 80 | 80 | 1 |
| 143 | 1 | 1 | 0 | 0 | 0 | 0 | 0 | 0 | 0 | 0 | 0 | 1 | DZ | Discordant NDC | Male | 8-12 | 21,11 | 0 | 81 | 81 | 1 |
| 144 | 1 | 1 | 0 | 0 | 0 | 0 | 1 | 1 | 0 | 1 | 0 | 0 | DZ | Discordant NDC | Male | 8-12 | 18,83 | 1 | 81 | 81 | 1 |
| 145 | 1 | 1 | 0 | 0 | 0 | 0 | 1 | 1 | 1 | 1 | 0 | 0 | MZ | Discordant NDC | Male | 13-17 | 15,47 | 1 | 82 | 82 | 1 |
| 146 | 1 | 1 | 0 | 0 | 0 | 0 | 0 | 0 | 0 | 0 | 0 | 1 | MZ | Discordant NDC | Male | 13-17 | 15,31 | 0 | 82 | 82 | 1 |
| 147 | 1 | 1 | 0 | 0 | 0 | 1 | 0 | 0 | 0 | 0 | 0 | 1 | MZ | Concordant TD | Female | 18-22 | 22,29 | 0 | 83 | 83 | 1 |
| 148 | 1 | 1 | 0 | 0 | 0 | 1 | 0 | 0 | 0 | 0 | 1 | 0 | MZ | Concordant TD | Female | 18-22 | 25,72 | 0 | 83 | 83 | 1 |
| 149 | 1 | 1 | 0 | 0 | 0 | 1 | 1 | 0 | 0 | 1 | 0 | 0 | MZ | Discordant NDC | Male | 13-17 | 18,78 | 0 | 84 | 84 | 1 |
| 150 | 1 | 1 | 0 | 0 | 0 | 1 | 0 | 0 | 0 | 0 | 0 | 1 | MZ | Discordant NDC | Male | 13-17 | 23,57 | 0 | 84 | 84 | 1 |
| 151 | 1 | 1 | 0 | 0 | 0 | 0 | 0 | 0 | 0 | 1 | 0 | 0 | DZ | Concordant NDC | Female | 13-17 | 19,7 | 0 | 85 | 85 | 1 |
| 152 | 1 | 1 | 0 | 0 | 0 | 0 | 1 | 0 | 1 | 1 | 0 | 0 | DZ | Concordant NDC | Female | 13-17 | 21,04 | 1 | 85 | 85 | 1 |
| 153 | 1 | 1 | 0 | 0 | 1 | 0 | 0 | 0 | 0 | 0 | 0 | 1 | MZ | Concordant TD | Male | 18-22 | 19,41 | 0 | 86 | 86 | 1 |
| 154 | 1 | 1 | 0 | 0 | 1 | 0 | 0 | 0 | 0 | 0 | 0 | 1 | MZ | Concordant TD | Male | 18-22 | 19,14 | 0 | 86 | 86 | 1 |
| 155 | 1 | 1 | 0 | 0 | 1 | 0 | 0 | 0 | 0 | 0 | 0 | 1 | MZ | Concordant TD | Male | 18-22 | 23,91 | 1 | 87 | 87 | 1 |
| 156 | 1 | 1 | 0 | 0 | 1 | 0 | 0 | 0 | 0 | 0 | 1 | 0 | MZ | Concordant TD | Male | 18-22 | 23,22 | 1 | 87 | 87 | 1 |
| 157 | 1 | 1 | 0 | 0 | 1 | 1 | 0 | 0 | 0 | 0 | 0 | 1 | MZ | Concordant TD | Male | 13-17 | 24,63 | 0 | 88 | 88 | 1 |
| 158 | 1 | 1 | 0 | 0 | 1 | 1 | 0 | 0 | 0 | 0 | 0 | 1 | MZ | Concordant TD | Male | 13-17 | 23,25 | 0 | 88 | 88 | 1 |
| 159 | 1 | 1 | 0 | 0 | 0 | 0 | 0 | 0 | 0 | 0 | 0 | 1 | DZ | Discordant NDC | Male | 13-17 | 19,26 | 0 | 89 | 8901 | 1 |
| 160 | 1 | 1 | 0 | 0 | 0 | 0 | 1 | 1 | 0 | 1 | 0 | 0 | DZ | Discordant NDC | Male | 13-17 | 17,01 | 1 | 89 | 8901 | 1 |
| 161 | 1 | 1 | 0 | 0 | 0 | 0 | 0 | 0 | 0 | 0 | 0 | 1 | DZ | Discordant NDC | Female | 13-17 | 21,6 | 0 | 89 | 8902 | 1 |
| 162 | 1 | 0 | 0 | 0 | 0 | 0 | 1 | 0 | 0 | 1 | 0 | 0 | DZ | Concordant NDC | Male | 13-17 | 23,73 | 1 | 90 | 90 | 1 |
| 163 | 1 | 0 | 0 | 0 | 0 | 0 | 1 | 1 | 0 | 1 | 0 | 0 | DZ | Concordant NDC | Male | 13-17 | 26,14 | 0 | 90 | 90 | 1 |
| 164 | 1 | 1 | 0 | 0 | 0 | 1 | 1 | 1 | 1 | 1 | 0 | 0 | MZ | Concordant NDC | Female | 13-17 | 19,43 | 1 | 91 | 91 | 1 |
| 165 | 1 | 1 | 0 | 0 | 0 | 1 | 1 | 1 | 1 | 1 | 0 | 0 | MZ | Concordant NDC | Female | 13-17 | 18,66 | 1 | 91 | 91 | 1 |
| 166 | 1 | 1 | 0 | 0 | 0 | 0 | 0 | 0 | 0 | 0 | 0 | 1 | MZ | Concordant TD | Female | 13-17 | 24,93 | 0 | 92 | 92 | 1 |
| 167 | 1 | 0 | 0 | 0 | 0 | 0 | 0 | 0 | 0 | 0 | 1 | 0 | MZ | Concordant TD | Female | 13-17 | 22,85 | 0 | 92 | 92 | 1 |
| 168 | 1 | 0 | 0 | 0 | 0 | 0 | 0 | 1 | 0 | 1 | 1 | 0 | DZ | Concordant NDC | Female | 13-17 | 28,53 | 1 | 93 | 93 | 1 |
| 169 | 1 | 1 | 0 | 0 | 1 | 0 | 0 | 1 | 0 | 1 | 0 | 0 | MZ | Concordant NDC | Male | 13-17 | 17,51 | 0 | 94 | 94 | 1 |
| 170 | 1 | 1 | 0 | 0 | 1 | 0 | 0 | 1 | 0 | 1 | 0 | 0 | MZ | Concordant NDC | Male | 13-17 | 17,54 | 0 | 94 | 94 | 1 |
| 171 | 1 | 1 | 0 | 0 | 1 | 0 | 0 | 0 | 0 | 0 | 0 | 1 | MZ | Concordant TD | Female | 18-22 | 22,92 | 0 | 95 | 95 | 1 |
| 172 | 1 | 1 | 0 | 0 | 1 | 0 | 0 | 0 | 0 | 0 | 0 | 1 | MZ | Concordant TD | Female | 18-22 | 26,95 | 0 | 95 | 95 | 1 |
| 173 | 1 | 1 | 0 | 0 | 1 | 0 | 0 | 0 | 0 | 0 | 0 | 1 | MZ | Concordant TD | Female | 18-22 | 20,24 | 0 | 97 | 97 | 1 |
| 174 | 1 | 1 | 0 | 0 | 1 | 0 | 0 | 0 | 0 | 0 | 0 | 1 | MZ | Concordant TD | Female | 18-22 | 18,94 | 0 | 97 | 97 | 1 |
| 175 | 1 | 1 | 0 | 0 | 0 | 0 | 0 | 0 | 0 | 0 | 0 | 1 | MZ | Discordant NDC | Male | 13-17 | 22,59 | 0 | 98 | 98 | 1 |
| 176 | 1 | 1 | 0 | 0 | 0 | 0 | 0 | 0 | 0 | 1 | 0 | 0 | MZ | Discordant NDC | Male | 13-17 | 24,94 | 0 | 98 | 98 | 1 |
| 177 | 1 | 1 | 0 | 0 | 0 | 0 | 0 | 0 | 0 | 0 | 1 | 0 | MZ | Concordant TD | Female | 18-22 | 22,37 | 1 | 99 | 99 | 1 |
| 178 | 1 | 1 | 0 | 0 | 0 | 0 | 0 | 0 | 0 | 0 | 1 | 0 | MZ | Concordant TD | Female | 18-22 | 21,03 | 1 | 99 | 99 | 1 |
| 179 | 1 | 1 | 0 | 0 | 0 | 0 | 0 | 0 | 0 | 0 | 1 | 0 | DZ | Discordant NDC | Female | 23-28 | 21,41 | 1 | 100 | 100 | 1 |
| 180 | 1 | 1 | 0 | 0 | 0 | 0 | 0 | 0 | 0 | 1 | 0 | 0 | DZ | Discordant NDC | Female | 23-28 | 19,93 | 1 | 100 | 100 | 1 |
| 181 | 1 | 1 | 0 | 0 | 0 | 0 | 1 | 0 | 0 | 1 | 1 | 0 | MZ | Discordant NDC | Female | 18-22 | 31,25 | 1 | 101 | 101 | 1 |
| 182 | 1 | 1 | 0 | 0 | 0 | 0 | 0 | 0 | 0 | 0 | 0 | 1 | MZ | Discordant NDC | Female | 18-22 | 30,19 | 0 | 101 | 101 | 1 |
| 183 | 1 | 1 | 0 | 0 | 0 | 0 | 1 | 0 | 0 | 1 | 0 | 0 | DZ | Concordant NDC | Male | 8-12 | 14,94 | 1 | 102 | 102 | 1 |
| 184 | 1 | 1 | 0 | 0 | 0 | 0 | 0 | 1 | 0 | 1 | 1 | 0 | DZ | Concordant NDC | Male | 8-12 | 20,27 | 1 | 102 | 102 | 1 |
| 185 | 1 | 0 | 0 | 0 | 1 | 0 | 0 | 0 | 0 | 0 | 0 | 1 | MZ | Concordant TD | Male | 13-17 | 19,84 | 0 | 103 | 103 | 1 |
| 186 | 1 | 0 | 0 | 0 | 1 | 0 | 0 | 0 | 0 | 0 | 0 | 1 | MZ | Concordant TD | Male | 13-17 | 19,94 | 0 | 103 | 103 | 1 |
| 187 | 1 | 1 | 0 | 0 | 1 | 0 | 0 | 0 | 0 | 0 | 0 | 1 | DZ | Discordant NDC | Female | 13-17 | 23,23 | 0 | 105 | 105 | 1 |
| 188 | 1 | 0 | 0 | 0 | 1 | 0 | 0 | 1 | 0 | 1 | 0 | 0 | DZ | Discordant NDC | Female | 13-17 | 24,52 | 0 | 105 | 105 | 1 |
| 189 | 1 | 1 | 0 | 0 | 0 | 0 | 1 | 1 | 0 | 1 | 1 | 0 | DZ | Discordant NDC | Male | 13-17 | 15,85 | 1 | 106 | 106 | 1 |
| 190 | 1 | 1 | 0 | 0 | 0 | 0 | 0 | 0 | 0 | 0 | 0 | 1 | DZ | Discordant NDC | Male | 13-17 | 15,4502 | 1 | 106 | 106 | 1 |
| 191 | 1 | 0 | 0 | 0 | 0 | 0 | 1 | 1 | 0 | 1 | 0 | 0 | DZ | Concordant NDC | Male | 13-17 | 18,8 | 1 | 107 | 107 | 1 |
| 192 | 1 | 0 | 0 | 0 | 0 | 0 | 0 | 1 | 0 | 1 | 0 | 0 | DZ | Concordant NDC | Male | 13-17 | 21,97 | 1 | 107 | 107 | 1 |
| 193 | 1 | 1 | 0 | 0 | 0 | 0 | 0 | 0 | 0 | 0 | 0 | 1 | MZ | Discordant NDC | Male | 13-17 | 20,69 | 1 | 108 | 108 | 1 |
| 194 | 1 | 0 | 0 | 0 | 0 | 0 | 1 | 0 | 0 | 1 | 0 | 0 | MZ | Discordant NDC | Male | 13-17 | 20,56 | 0 | 108 | 108 | 1 |
| 195 | 1 | 1 | 0 | 0 | 1 | 0 | 0 | 0 | 0 | 0 | 0 | 1 | DZ | Discordant NDC | Female | 8-12 | 14,02 | 0 | 109 | 109 | 1 |
| 196 | 1 | 1 | 0 | 0 | 1 | 0 | 0 | 1 | 0 | 1 | 0 | 0 | DZ | Discordant NDC | Female | 8-12 | 13,43 | 1 | 109 | 109 | 1 |
| 197 | 1 | 0 | 0 | 0 | 0 | 0 | 0 | 0 | 0 | 0 | 0 | 1 | DZ | Discordant NDC | Female | 18-22 | 23,69 | 0 | 110 | 110 | 0 |
| 198 | 1 | 1 | 0 | 0 | 1 | 0 | 0 | 0 | 0 | 0 | 0 | 1 | DZ | Concordant TD | Female | 13-17 | 22,2 | 0 | 111 | 111 | 1 |
| 199 | 1 | 1 | 0 | 0 | 1 | 0 | 0 | 1 | 0 | 1 | 0 | 0 | DZ | Concordant NDC | Female | 13-17 | 17,56 | 1 | 112 | 112 | 1 |
| 200 | 1 | 1 | 0 | 0 | 1 | 0 | 0 | 1 | 0 | 1 | 0 | 0 | DZ | Concordant NDC | Male | 13-17 | 16,66 | 1 | 112 | 112 | 1 |
| 201 | 1 | 0 | 0 | 0 | 1 | 0 | 0 | 0 | 0 | 0 | 0 | 1 | MZ | Concordant TD | Female | 13-17 | 18,49 | 0 | 113 | 113 | 1 |
| 202 | 1 | 0 | 0 | 0 | 1 | 0 | 0 | 0 | 0 | 0 | 0 | 1 | MZ | Concordant TD | Female | 13-17 | 21,12 | 0 | 113 | 113 | 1 |
| 203 | 1 | 1 | 0 | 0 | 1 | 0 | 0 | 0 | 0 | 0 | 0 | 1 | DZ | Concordant TD | Male | 13-17 | 20,27 | 0 | 114 | 114 | 1 |
| 204 | 1 | 1 | 0 | 0 | 1 | 0 | 0 | 0 | 0 | 0 | 0 | 1 | DZ | Concordant TD | Male | 13-17 | 22,33 | 0 | 114 | 114 | 1 |
| 205 | 1 | 0 | 0 | 0 | 1 | 0 | 0 | 1 | 0 | 1 | 0 | 0 | DZ | Discordant NDC | Female | 8-12 | 16,7 | 0 | 115 | 115 | 1 |
| 206 | 1 | 0 | 0 | 0 | 1 | 0 | 0 | 0 | 0 | 0 | 0 | 1 | DZ | Discordant NDC | Female | 8-12 | 19,99 | 1 | 115 | 115 | 1 |
| 207 | 1 | 0 | 0 | 0 | 0 | 1 | 1 | 0 | 0 | 1 | 0 | 0 | MZ | Concordant NDC | Male | 8-12 | 12,87 | 1 | 116 | 116 | 1 |
| 208 | 1 | 0 | 0 | 0 | 0 | 1 | 1 | 0 | 0 | 1 | 0 | 0 | MZ | Concordant NDC | Male | 8-12 | 13,15 | 1 | 116 | 116 | 1 |
| 209 | 1 | 0 | 0 | 0 | 0 | 0 | 0 | 0 | 0 | 0 | 0 | 1 | DZ | Discordant NDC | Male | 18-22 | 24,02 | 0 | 117 | 117 | 1 |
| 210 | 1 | 0 | 0 | 0 | 0 | 0 | 1 | 0 | 1 | 1 | 1 | 0 | DZ | Discordant NDC | Male | 18-22 | 19,94 | 0 | 117 | 117 | 1 |
| 211 | 1 | 0 | 0 | 0 | 0 | 0 | 1 | 0 | 0 | 1 | 0 | 0 | MZ | Concordant NDC | Male | 13-17 | 17,12 | 0 | 118 | 118 | 1 |
| 212 | 1 | 0 | 0 | 0 | 0 | 0 | 1 | 0 | 0 | 1 | 0 | 0 | MZ | Concordant NDC | Male | 13-17 | 16,74 | 0 | 118 | 118 | 1 |
| 213 | 1 | 0 | 0 | 0 | 0 | 0 | 1 | 1 | 0 | 1 | 1 | 0 | DZ | Discordant NDC | Female | 8-12 | 17,9 | 1 | 119 | 119 | 1 |
| 214 | 1 | 0 | 0 | 0 | 0 | 0 | 0 | 0 | 0 | 0 | 1 | 0 | DZ | Discordant NDC | Female | 8-12 | 18,8 | 0 | 119 | 119 | 1 |
| 215 | 1 | 0 | 0 | 0 | 1 | 0 | 0 | 0 | 0 | 0 | 0 | 1 | DZ | Concordant TD | Female | 13-17 | 17,62 | 0 | 120 | 120 | 1 |
| 216 | 1 | 0 | 0 | 0 | 1 | 0 | 0 | 0 | 0 | 0 | 0 | 1 | DZ | Concordant TD | Male | 13-17 | 19,35 | 0 | 120 | 120 | 1 |
| 217 | 1 | 0 | 0 | 0 | 0 | 0 | 0 | 0 | 0 | 0 | 1 | 0 | DZ | Discordant NDC | Female | 13-17 | 20,75 | 1 | 121 | 121 | 0 |
| 218 | 1 | 0 | 0 | 0 | 0 | 0 | 0 | 1 | 0 | 1 | 1 | 0 | DZ | Concordant NDC | Female | 8-12 | 32,22 | 0 | 122 | 122 | 0 |
| 219 | 1 | 0 | 0 | 0 | 0 | 0 | 0 | 0 | 0 | 0 | 1 | 0 | MZ | Discordant NDC | Female | 18-22 | 20,87 | 0 | 123 | 123 | 1 |
| 220 | 1 | 0 | 0 | 0 | 0 | 0 | 0 | 1 | 0 | 1 | 1 | 0 | MZ | Discordant NDC | Female | 18-22 | 21,67 | 0 | 123 | 123 | 1 |
| 221 | 0 | 1 | 1 | 0 | 0 | 0 | 0 | 0 | 0 | 0 | 0 | 1 | MZ | Discordant NDC | Male | 18-22 | 26,97 | 1 | 28 | 28 | 1 |
| 222 | 0 | 1 | 0 | 0 | 0 | 0 | 1 | 0 | 1 | 1 | 1 | 0 | MZ | Concordant NDC | Male | 8-12 | 18,14 | 1 | 32 | 32 | 1 |
| 223 | 0 | 1 | 0 | 0 | 0 | 0 | 1 | 1 | 0 | 1 | 0 | 0 | MZ | Concordant NDC | Male | 13-17 | 28,8 | 1 | 38 | 38 | 1 |
| 224 | 0 | 1 | 0 | 0 | 0 | 0 | 1 | 1 | 0 | 1 | 0 | 0 | MZ | Concordant NDC | Male | 13-17 | 28,4 | 1 | 38 | 38 | 1 |
| 225 | 0 | 1 | 0 | 0 | 0 | 0 | 1 | 0 | 1 | 1 | 0 | 0 | DZ | Discordant NDC | Male | 13-17 | NA | 0 | 42 | 42 | 1 |
| 226 | 0 | 1 | 1 | 1 | 0 | 0 | 1 | 0 | 0 | 1 | 0 | 0 | MZ | Discordant NDC | Female | 18-22 | 21,53 | 0 | 46 | 46 | 1 |
| 227 | 0 | 1 | 1 | 1 | 1 | 0 | 0 | 0 | 0 | 0 | 0 | 1 | MZ | Concordant TD | Male | 18-22 | 22,46 | 1 | 47 | 47 | 1 |
| 228 | 0 | 1 | 1 | 1 | 1 | 0 | 0 | 0 | 0 | 0 | 0 | 1 | MZ | Concordant TD | Male | 18-22 | 21,8 | 1 | 47 | 47 | 1 |
| 229 | 0 | 1 | 0 | 0 | 0 | 0 | 1 | 1 | 0 | 1 | 0 | 0 | DZ | Discordant NDC | Male | 13-17 | 19,02 | 1 | 89 | 8902 | 1 |
| 230 | 0 | 1 | 0 | 0 | 0 | 0 | 1 | 1 | 1 | 1 | 0 | 0 | DZ | Concordant NDC | Female | 13-17 | 23,73 | 1 | 93 | 93 | 1 |
| 231 | 0 | 1 | 0 | 0 | 1 | 0 | 0 | 0 | 0 | 0 | 0 | 1 | DZ | Concordant TD | Female | 13-17 | 18,79 | 0 | 111 | 111 | 1 |
| 232 | 0 | 0 | 0 | 0 | 1 | 0 | 0 | 1 | 0 | 1 | 0 | 0 | MZ | Discordant NDC | Male | 8-12 | 21,22 | 0 | 132 | 132 | 0 |
| 233 | 0 | 0 | 0 | 0 | 1 | 0 | 0 | 0 | 0 | 0 | 0 | 1 | DZ | Discordant NDC | Male | 13-17 | 24,63 | 0 | 130 | 130 | 1 |
| 234 | 0 | 0 | 0 | 0 | 1 | 0 | 0 | 1 | 0 | 1 | 0 | 0 | DZ | Discordant NDC | Male | 13-17 | 23,18 | 1 | 130 | 130 | 1 |
| 235 | 0 | 0 | 0 | 0 | 1 | 0 | 0 | 0 | 0 | 0 | 0 | 1 | DZ | Concordant TD | Male | 8-12 | 17,85 | 1 | 128 | 128 | 0 |
| 236 | 0 | 0 | 0 | 0 | 1 | 0 | 0 | 0 | 0 | 1 | 1 | 0 | DZ | Concordant NDC | Female | 8-12 | 19,45 | 0 | 134 | 134 | 1 |
| 237 | 0 | 0 | 0 | 0 | 1 | 0 | 0 | 1 | 0 | 1 | 0 | 0 | DZ | Concordant NDC | Female | 8-12 | 19,85 | 1 | 134 | 134 | 1 |

**Table S2. Study population characteristics and descriptive statistics for each omics dataset.**

| **Dataset** | **Whole dataset n=237** | | **Blood n=220** | | **Fecal n=189** | | **Serum n=126** | | **CSF n=86** | | **Urine n=112** | | **Euaims n=28** | |
|---|---|---|---|---|---|---|---|---|---|---|---|---|---|---|
| NDC diagnosis | noNDC | anyNDC | noNDC | anyNDC | noNDC | anyNDC | noNDC | anyNDC | noNDC | anyNDC | noNDC | anyNDC | noNDC | anyNDC |
| Number (%) | 115 (48.5%) | 122 (51.5%) | 109 (49.5%) | 111 (50.5%) | 96 (50.8%) | 93 (49.2%) | 60 (47.6%) | 66 (52.4%) | 41 (47.7%) | 45 (52.3%) | 75 (67.0%) | 37 (33.0%) | 13 (46.4%) | 15 (53.6%) |
| Age, mean (SD) | 15.3 (3.3) | 14.1 (4.0) | 15.2 (3.3) | 14.2 (4.0) | 15.2 (3.3) | 13.9 (3.7) | 14.3 (3.0) | 14.1 (4.4) | 14.5 (3.1) | 14.2 (3.8) | 15.1 (3.0) | 13.8 (3.1) | 15.2 (3.1) | 12.8 (3.5) |
| Sex, Female N (%) | 58 (50.4%) | 46 (37.7%) | 57 (52.3%) | 42 (37.8%) | 46 (47.9%) | 35 (37.6%) | 31 (51.7%) | 26 (39.4%) | 19 (46.3%) | 14 (31.1%) | 36 (48.0%) | 15 (40.5%) | 7 (53.8%) | 5 (33.3%) |
| Concordant twin pairs, N (MZ:DZ) | 30:6 | 25:15 | 29:5 | 23:13 | 25:5 | 19:10 | 18:2 | 19:4 | 13:1 | 14:3 | 27:5 | 6:7 | 5:0 | 6:0 |
| Discordant NDC twin pairs, N (MZ:DZ) | 23:17 | | 21:14 | | 20:11 | | 15:4 | | 5:3 | | 5:5 | | 3:0 | |
| BMI, mean (SD) | 20.8 (3.1) | 20.2 (3.9) | 20.8 (3.1) | 20.0 (3.8) | 21.0 (3.2) | 19.9 (3.5) | 20.3 (3.2) | 20.2 (3.5) | 19.7 (2.7) | 20.8 (3.6) | 20.5 (2.8) | 19.4 (3.4) | 21.0 (3.0) | 19.1 (3.5) |
| Any medication, N (SD) | 18 (15.7%) | 65 (53.3%) | 14 (12.8%) | 58 (52.3%) | 15 (15.6%) | 50 (53.8%) | 7 (13.3%) | 31 (47.0%) | 6 (14.6%) | 23 (51.1%) | 9 (12.0%) | 18 (48.6%) | 2 (15.4%) | 12 (80.0%) |
| Psychiaric diagnosis, N (%) | 11 (9.6%) | 36 (29.5%) | 11 (10.1%) | 34 (30.6%) | 7 (7.3%) | 24 (25.8%) | 1 (1.7%) | 20 (30.3%) | 1 (2.4%) | 13 (28.9%) | 2 (2.7%) | 10 (27.0%) | 1 (7.7%) | 3 (20.0%) |

| | |
|---|---|
| Age, mean (SD) | 14.7 (3.7) |
| BMI, mean (SD) | 20.5 (3.5) |

**Table S3. The parameters tunning in single omics and multi-omics models.**

### Single omics

| | Distance measurement | Classification error rate | Tune.test.keepX | KeepX[1] | Component number |
|---|---|---|---|---|---|
| Serum | Mahalanobis distance | Balanced Error Rate | 5, 10, 15, 20, 25, 30, 35, 40, 50, 60, 70, 80, 90, 100, 110, 120, 130, 140, 150, 160 | 5, 5, 5, 5, 25, 30 | 6 |
| CSF | Maximum distance | Balanced Error Rate | 5, 10, 15, 20, 25, 30, 35, 40, 50, 60, 70, 80, 90, 100, 110 | 30, 50, 100 | 3 |
| Fecal | Mahalanobis distance | Balanced Error Rate | 5, 10, 15, 20, 25, 30, 35, 40, 50, 60, 70, 80, 90, 100, 200, 300, 400, 500 | 5,50 | 2 |
| Blood | Maximum distance | Overall Error Rate | 3, 4, 5, 6, 7, 8, 9, 10, 11, 12, 13 | 4, 5, 13 | 3 |
| Urine | Maximum distance | Overall Error Rate | 5, 10, 15, 20, 25, 30, 35, 40, 45, 50, 55 | 5 | 1 |

### Multi-omics

**MultiOmic-SCFB**

| | Distance measurement | Classification error rate | Test.tune | KeepX[1] | Component number |
|---|---|---|---|---|---|
| Serum | Mahalanobis distance | Balanced Error Rate | 5, 10, 15, 20, 25, 30 | 30, 25 | 2 |
| CSF | | | 5, 10, 15, 20, 25, 30, 50, 100 | 5, 20 | |
| Fecal | | | 5, 10, 15, 20, 25, 30, 40, 50 | 5, 10 | |
| Blood | | | 4, 5, 9, 13 | 9, 13 | |

**MultiOmic-SFB**

| | Distance measurement | Classification error rate | Test.tune | KeepX[1] | Component number |
|---|---|---|---|---|---|
| Serum | Mahalanobis distance | Balanced Error Rate | 5, 10, 15, 20, 25, 30 | 5, 20, 5 | 3 |
| Fecal | | | 5, 10, 15, 20, 25, 30, 40, 50 | 10, 10, 25 | |
| Blood | | | 4, 5, 9, 13 | 5, 4, 13 | |

**MultiOmic-UFB**

| | Distance measurement | Classification error rate | Test.tune | KeepX[1] | Component number |
|---|---|---|---|---|---|
| Urine | Maximum distance | Overall Error Rate | 5 | 5, 5 | 2 |
| Fecal | | | 5, 10, 15, 20, 25, 30, 40, 50 | 30, 5 | |
| Blood | | | 4, 5, 9, 13 | 4, 13 | |

1 KeepX = feature number selected after tuning

**Table S4. GEE model estimates of covariate associations with DIABLO component scores of *MultiOmic-SCFB* model.** Coefficients for associations between covariates and average component scores (variate 1 and variate 2) were evaluated in a GEE model with twin ID as the clustering variable.

| Covariate | Variate | Estimate | SE | p_raw | p_adj |
|---|---|---|---|---|---|
| Age | Variate 1 | 0,1265 | 0,0543 | 0,0199 | 0,1279 |
| Sex (Male) | Variate 1 | -0,2438 | 0,4726 | 0,6059 | 0,8408 |
| BMI | Variate 1 | -0,0108 | 0,0538 | 0,8408 | 0,8408 |
| Medication | Variate 1 | -0,6387 | 0,4725 | 0,1765 | 0,4412 |
| Other psychiatric | Variate 1 | -0,3813 | 0,3894 | 0,3274 | 0,5679 |
| Age | Variate 2 | 0,0507 | 0,0532 | 0,3408 | 0,5679 |
| Sex (Male) | Variate 2 | -0,0686 | 0,2774 | 0,8046 | 0,8408 |
| BMI | Variate 2 | 0,0893 | 0,0498 | 0,0726 | 0,2420 |
| Medication | Variate 2 | 0,6163 | 0,2761 | 0,0256 | 0,1279 |
| Other psychiatric | Variate 2 | 0,1022 | 0,4234 | 0,8092 | 0,8408 |

**Table S5. Joint pathway enrichment analysis of key proteomic and metabolomic features in all pathways.** This table details the functional enrichment of key molecular features identified through integrated multi-omic models. Analysis was performed using MetaboAnalyst 6.0 Joint Pathway Analysis based on the KEGG database with all pathway datasets and Combine Query method.

| | Total | Expected | Hits | Raw p | (-LOG10(p)) | Holm adjust | FDR | Impact |
|---|---|---|---|---|---|---|---|---|
| Cytokine-cytokine receptor interaction | 298 | 3,43 | 29 | 1,89E-19 | 18,72 | 6,64E-17 | 6,64E-17 | 0,149 |
| Viral protein interaction with cytokine and cytokine receptor | 100 | 1,15 | 18 | 4,36E-17 | 16,36 | 1,52E-14 | 7,64E-15 | 0,125 |
| ABC transporters | 184 | 2,12 | 18 | 2,52E-12 | 11,60 | 8,78E-10 | 2,94E-10 | 0,000 |
| IL-17 signaling pathway | 95 | 1,09 | 11 | 9,96E-09 | 8,00 | 3,47E-06 | 8,74E-07 | 0,430 |
| Rheumatoid arthritis | 98 | 1,13 | 10 | 1,64E-07 | 6,79 | 5,68E-05 | 1,15E-05 | 0,000 |
| Chemokine signaling pathway | 198 | 2,28 | 12 | 2,70E-06 | 5,57 | 0,00093 | 0,00016 | 0,015 |
| Central carbon metabolism in cancer | 108 | 1,24 | 9 | 3,89E-06 | 5,41 | 0,00134 | 0,00018 | 0,027 |
| PI3K-Akt signaling pathway | 366 | 4,21 | 16 | 4,08E-06 | 5,39 | 0,0014 | 0,00018 | 0,127 |
| EGFR tyrosine kinase inhibitor resistance | 84 | 0,97 | 8 | 5,06E-06 | 5,30 | 0,00174 | 0,0002 | 1,303 |
| TNF signaling pathway | 119 | 1,37 | 9 | 8,65E-06 | 5,06 | 0,00296 | 0,0003 | 0,038 |
| HIF-1 signaling pathway | 125 | 1,44 | 8 | 9,24E-05 | 4,03 | 0,03151 | 0,00291 | 0,120 |
| Bladder cancer | 41 | 0,47 | 5 | 9,96E-05 | 4,00 | 0,03386 | 0,00291 | 0,063 |
| Caffeine metabolism | 28 | 0,32 | 4 | 0,00028 | 3,56 | 0,09338 | 0,00744 | 0,200 |
| Malaria | 55 | 0,63 | 5 | 0,00041 | 3,39 | 0,1375 | 0,0102 | 0,024 |
| Pyrimidine metabolism | 124 | 1,43 | 7 | 0,00055 | 3,26 | 0,18623 | 0,01268 | 0,159 |
| ErbB signaling pathway | 90 | 1,04 | 6 | 0,00058 | 3,24 | 0,19427 | 0,01268 | 1,420 |
| Alanine, aspartate and glutamate metabolism | 65 | 0,75 | 5 | 0,00088 | 3,05 | 0,29539 | 0,01821 | 0,410 |
| Human cytomegalovirus infection | 232 | 2,67 | 9 | 0,00139 | 2,86 | 0,46553 | 0,02718 | 0,091 |
| Chagas disease | 109 | 1,25 | 6 | 0,00158 | 2,80 | 0,52502 | 0,02913 | 0,123 |
| MAPK signaling pathway | 305 | 3,51 | 10 | 0,00266 | 2,58 | 0,8827 | 0,04666 | 0,032 |
| Proteoglycans in cancer | 211 | 2,43 | 8 | 0,00296 | 2,53 | 0,98012 | 0,04949 | 0,115 |
| JAK-STAT signaling pathway | 168 | 1,93 | 7 | 0,00321 | 2,49 | 1 | 0,05117 | 0,159 |
| Valine, leucine and isoleucine biosynthesis | 27 | 0,31 | 3 | 0,00355 | 2,45 | 1 | 0,05423 | 0,154 |
| Mineral absorption | 90 | 1,04 | 5 | 0,00374 | 2,43 | 1 | 0,05468 | 0,000 |
| Influenza A | 176 | 2,03 | 7 | 0,00415 | 2,38 | 1 | 0,05821 | 0,096 |
| Lipid and atherosclerosis | 230 | 2,65 | 8 | 0,00499 | 2,30 | 1 | 0,06729 | 0,079 |
| Pathways in cancer | 564 | 6,49 | 14 | 0,00518 | 2,29 | 1 | 0,06729 | 0,130 |
| Hematopoietic cell lineage | 100 | 1,15 | 5 | 0,00585 | 2,23 | 1 | 0,07335 | 0,000 |
| Coronavirus disease - COVID-19 | 248 | 2,86 | 8 | 0,00776 | 2,11 | 1 | 0,09188 | 0,120 |
| NF-kappa B signaling pathway | 108 | 1,24 | 5 | 0,00806 | 2,09 | 1 | 0,09188 | 0,067 |
| Protein digestion and absorption | 152 | 1,75 | 6 | 0,00811 | 2,09 | 1 | 0,09188 | 0,000 |
| AGE-RAGE signaling pathway in diabetic complications | 110 | 1,27 | 5 | 0,00869 | 2,06 | 1 | 0,09533 | 0,118 |
| Amoebiasis | 116 | 1,34 | 5 | 0,01079 | 1,97 | 1 | 0,11477 | 0,000 |
| Calcium signaling pathway | 265 | 3,05 | 8 | 0,01133 | 1,95 | 1 | 0,11594 | 0,038 |
| Aminoacyl-tRNA biosynthesis | 118 | 1,36 | 5 | 0,01156 | 1,94 | 1 | 0,11594 | 0,052 |
| Epithelial cell signaling in Helicobacter pylori infection | 78 | 0,90 | 4 | 0,01237 | 1,91 | 1 | 0,12056 | 0,262 |
| African trypanosomiasis | 45 | 0,52 | 3 | 0,01485 | 1,83 | 1 | 0,14083 | 0,083 |
| Purine metabolism | 229 | 2,64 | 7 | 0,01643 | 1,78 | 1 | 0,14642 | 0,160 |
| Graft-versus-host disease | 47 | 0,54 | 3 | 0,01669 | 1,78 | 1 | 0,14642 | 0,087 |
| Arginine biosynthesis | 47 | 0,54 | 3 | 0,01669 | 1,78 | 1 | 0,14642 | 0,128 |
| Pertussis | 88 | 1,01 | 4 | 0,01853 | 1,73 | 1 | 0,15864 | 0,000 |
| Valine, leucine and isoleucine degradation | 90 | 1,04 | 4 | 0,01996 | 1,70 | 1 | 0,16677 | 0,074 |
| FoxO signaling pathway | 138 | 1,59 | 5 | 0,02141 | 1,67 | 1 | 0,17474 | 0,059 |
| Nicotinate and nicotinamide metabolism | 93 | 1,07 | 4 | 0,02222 | 1,65 | 1 | 0,17724 | 0,062 |
| Ras signaling pathway | 245 | 2,82 | 7 | 0,02286 | 1,64 | 1 | 0,17832 | 0,041 |
| Antifolate resistance | 58 | 0,67 | 3 | 0,029 | 1,54 | 1 | 0,21661 | 0,017 |
| Legionellosis | 58 | 0,67 | 3 | 0,029 | 1,54 | 1 | 0,21661 | 0,063 |
| Linoleic acid metabolism | 59 | 0,68 | 3 | 0,03031 | 1,52 | 1 | 0,22107 | 0,024 |
| Focal adhesion | 205 | 2,36 | 6 | 0,03086 | 1,51 | 1 | 0,22107 | 0,203 |
| Gastric cancer | 154 | 1,77 | 5 | 0,03237 | 1,49 | 1 | 0,22721 | 0,087 |
| beta-Alanine metabolism | 63 | 0,73 | 3 | 0,03583 | 1,45 | 1 | 0,23965 | 0,056 |
| Non-alcoholic fatty liver disease | 159 | 1,83 | 5 | 0,0364 | 1,44 | 1 | 0,23965 | 0,114 |
| Toll-like receptor signaling pathway | 109 | 1,25 | 4 | 0,03687 | 1,43 | 1 | 0,23965 | 0,144 |
| Prostate cancer | 109 | 1,25 | 4 | 0,03687 | 1,43 | 1 | 0,23965 | 0,258 |
| Choline metabolism in cancer | 110 | 1,27 | 4 | 0,03793 | 1,42 | 1 | 0,24207 | 0,075 |
| Rap1 signaling pathway | 217 | 2,50 | 6 | 0,03909 | 1,41 | 1 | 0,24502 | 0,043 |
| Inflammatory bowel disease | 66 | 0,76 | 3 | 0,04029 | 1,39 | 1 | 0,24809 | 0,161 |
| Vitamin digestion and absorption | 68 | 0,78 | 3 | 0,04341 | 1,36 | 1 | 0,2627 | 0,000 |
| Lysine degradation | 119 | 1,37 | 4 | 0,04829 | 1,32 | 1 | 0,28729 | 0,085 |
| Riboflavin metabolism | 32 | 0,37 | 2 | 0,05208 | 1,28 | 1 | 0,30468 | 0,250 |
| Melanoma | 74 | 0,85 | 3 | 0,05347 | 1,27 | 1 | 0,30768 | 0,091 |
| D-Amino acid metabolism | 75 | 0,86 | 3 | 0,05525 | 1,26 | 1 | 0,31278 | 0,000 |

| | | | | | | | | |
|---|---|---|---|---|---|---|---|---|
| Vitamin B6 metabolism | 35 | 0,40 | 2 | 0,06111 | 1,21 | 1 | 0,33312 | 0,100 |
| Herpes simplex virus 1 infection | 185 | 2,13 | 5 | 0,06221 | 1,21 | 1 | 0,33312 | 0,045 |
| Biosynthesis of unsaturated fatty acids | 79 | 0,91 | 3 | 0,06264 | 1,20 | 1 | 0,33312 | 0,015 |
| Non-small cell lung cancer | 79 | 0,91 | 3 | 0,06264 | 1,20 | 1 | 0,33312 | 0,186 |
| Natural killer cell mediated cytotoxicity | 138 | 1,59 | 4 | 0,07478 | 1,13 | 1 | 0,39176 | 0,092 |
| Allograft rejection | 40 | 0,46 | 2 | 0,07723 | 1,11 | 1 | 0,39866 | 0,033 |
| Estrogen signaling pathway | 147 | 1,69 | 4 | 0,08944 | 1,05 | 1 | 0,45497 | 0,128 |
| Fluid shear stress and atherosclerosis | 149 | 1,72 | 4 | 0,09287 | 1,03 | 1 | 0,46569 | 0,086 |
| Type I diabetes mellitus | 47 | 0,54 | 2 | 0,10171 | 0,99 | 1 | 0,50284 | 0,048 |
| Alcoholic liver disease | 155 | 1,78 | 4 | 0,10354 | 0,98 | 1 | 0,50478 | 0,037 |
| GnRH signaling pathway | 99 | 1,14 | 3 | 0,10576 | 0,98 | 1 | 0,50851 | 0,106 |
| Virion - Hepatitis viruses | 49 | 0,56 | 2 | 0,10906 | 0,96 | 1 | 0,5173 | 0,000 |
| Hepatitis C | 161 | 1,85 | 4 | 0,11475 | 0,94 | 1 | 0,53125 | 0,062 |
| Folate transport and metabolism | 51 | 0,59 | 2 | 0,11654 | 0,93 | 1 | 0,53125 | 0,000 |
| Pantothenate and CoA biosynthesis | 51 | 0,59 | 2 | 0,11654 | 0,93 | 1 | 0,53125 | 0,067 |
| Intestinal immune network for IgA production | 52 | 0,60 | 2 | 0,12033 | 0,92 | 1 | 0,54148 | 0,093 |
| Necroptosis | 169 | 1,95 | 4 | 0,13049 | 0,88 | 1 | 0,57345 | 0,033 |
| Endocrine resistance | 109 | 1,25 | 3 | 0,1307 | 0,88 | 1 | 0,57345 | 0,129 |
| Th17 cell differentiation | 113 | 1,30 | 3 | 0,14121 | 0,85 | 1 | 0,61192 | 0,057 |
| Endometrial cancer | 60 | 0,69 | 2 | 0,15164 | 0,82 | 1 | 0,64908 | 0,065 |
| Cysteine and methionine metabolism | 120 | 1,38 | 3 | 0,16026 | 0,80 | 1 | 0,67772 | 0,000 |
| Parathyroid hormone synthesis, secretion and action | 126 | 1,45 | 3 | 0,17717 | 0,75 | 1 | 0,74032 | 0,075 |
| Pentose phosphate pathway | 68 | 0,78 | 2 | 0,18433 | 0,73 | 1 | 0,76044 | 0,030 |
| NOD-like receptor signaling pathway | 195 | 2,25 | 4 | 0,18713 | 0,73 | 1 | 0,76044 | 0,045 |
| Histidine metabolism | 69 | 0,79 | 2 | 0,18848 | 0,72 | 1 | 0,76044 | 0,032 |
| Tuberculosis | 200 | 2,30 | 4 | 0,19884 | 0,70 | 1 | 0,78737 | 0,058 |
| Kaposi sarcoma-associated herpesvirus infection | 201 | 2,31 | 4 | 0,20121 | 0,70 | 1 | 0,78737 | 0,060 |
| Regulation of lipolysis in adipocytes | 73 | 0,84 | 2 | 0,20524 | 0,69 | 1 | 0,78737 | 0,032 |
| RIG-I-like receptor signaling pathway | 73 | 0,84 | 2 | 0,20524 | 0,69 | 1 | 0,78737 | 0,033 |
| Relaxin signaling pathway | 136 | 1,57 | 3 | 0,20638 | 0,69 | 1 | 0,78737 | 0,067 |
| Pathogenic Escherichia coli infection | 206 | 2,37 | 4 | 0,21318 | 0,67 | 1 | 0,8046 | 0,064 |
| Apoptosis | 140 | 1,61 | 3 | 0,21836 | 0,66 | 1 | 0,81535 | 0,071 |
| Platinum drug resistance | 78 | 0,90 | 2 | 0,2264 | 0,65 | 1 | 0,82299 | 0,060 |
| Adipocytokine signaling pathway | 78 | 0,90 | 2 | 0,2264 | 0,65 | 1 | 0,82299 | 0,067 |
| Yersinia infection | 143 | 1,65 | 3 | 0,22744 | 0,64 | 1 | 0,82299 | 0,048 |
| Pancreatic cancer | 79 | 0,91 | 2 | 0,23066 | 0,64 | 1 | 0,82613 | 0,140 |
| Osteoclast differentiation | 146 | 1,68 | 3 | 0,23659 | 0,63 | 1 | 0,83882 | 0,038 |
| Prolactin signaling pathway | 82 | 0,94 | 2 | 0,24345 | 0,61 | 1 | 0,8545 | 0,031 |
| Vascular smooth muscle contraction | 150 | 1,73 | 3 | 0,24889 | 0,60 | 1 | 0,86491 | 0,043 |
| Breast cancer | 151 | 1,74 | 3 | 0,25198 | 0,60 | 1 | 0,86491 | 0,117 |
| Leishmaniasis | 85 | 0,98 | 2 | 0,25627 | 0,59 | 1 | 0,86491 | 0,036 |
| Cytosolic DNA-sensing pathway | 85 | 0,98 | 2 | 0,25627 | 0,59 | 1 | 0,86491 | 0,057 |
| Phospholipase D signaling pathway | 160 | 1,84 | 3 | 0,28004 | 0,55 | 1 | 0,93614 | 0,155 |
| Cellular senescence | 162 | 1,87 | 3 | 0,28632 | 0,54 | 1 | 0,9481 | 0,009 |
| Adherens junction | 93 | 1,07 | 2 | 0,29049 | 0,54 | 1 | 0,94917 | 0,033 |
| Hepatitis B | 164 | 1,89 | 3 | 0,29261 | 0,53 | 1 | 0,94917 | 0,036 |
| PD-L1 expression and PD-1 checkpoint pathway in cancer | 94 | 1,08 | 2 | 0,29476 | 0,53 | 1 | 0,94917 | 0,050 |
| Retrograde endocannabinoid signaling | 168 | 1,93 | 3 | 0,30523 | 0,52 | 1 | 0,96851 | 0,000 |
| Biotin metabolism | 32 | 0,37 | 1 | 0,31006 | 0,51 | 1 | 0,96851 | 0,000 |
| Glycolysis or Gluconeogenesis | 98 | 1,13 | 2 | 0,3118 | 0,51 | 1 | 0,96851 | 0,031 |
| GABAergic synapse | 98 | 1,13 | 2 | 0,3118 | 0,51 | 1 | 0,96851 | 0,071 |
| Hypertrophic cardiomyopathy | 102 | 1,17 | 2 | 0,32876 | 0,48 | 1 | 1 | 0,000 |
| Morphine addiction | 103 | 1,19 | 2 | 0,33298 | 0,48 | 1 | 1 | 0,070 |
| Efferocytosis | 178 | 2,05 | 3 | 0,33684 | 0,47 | 1 | 1 | 0,008 |
| Asthma | 37 | 0,43 | 1 | 0,34901 | 0,46 | 1 | 1 | 0,000 |
| Nitrogen metabolism | 37 | 0,43 | 1 | 0,34901 | 0,46 | 1 | 1 | 0,021 |
| Primary immunodeficiency | 38 | 0,44 | 1 | 0,35653 | 0,45 | 1 | 1 | 0,000 |
| TGF-beta signaling pathway | 109 | 1,25 | 2 | 0,35815 | 0,45 | 1 | 1 | 0,021 |
| Neuroactive ligand-receptor interaction | 423 | 4,87 | 6 | 0,36053 | 0,44 | 1 | 1 | 0,041 |
| Shigellosis | 264 | 3,04 | 4 | 0,36173 | 0,44 | 1 | 1 | 0,037 |
| Proximal tubule bicarbonate reclamation | 40 | 0,46 | 1 | 0,37132 | 0,43 | 1 | 1 | 0,000 |
| Phenylalanine, tyrosine and tryptophan biosynthesis | 41 | 0,47 | 1 | 0,37859 | 0,42 | 1 | 1 | 0,000 |
| Toxoplasmosis | 115 | 1,32 | 2 | 0,38298 | 0,42 | 1 | 1 | 0,049 |
| Taste transduction | 119 | 1,37 | 2 | 0,39931 | 0,40 | 1 | 1 | 0,056 |
| Transcriptional misregulation in cancer | 198 | 2,28 | 3 | 0,39971 | 0,40 | 1 | 1 | 0,000 |
| Arginine and proline metabolism | 121 | 1,39 | 2 | 0,4074 | 0,39 | 1 | 1 | 0,101 |
| Proteasome | 46 | 0,53 | 1 | 0,4137 | 0,38 | 1 | 1 | 0,000 |
| Nicotine addiction | 48 | 0,55 | 1 | 0,42718 | 0,37 | 1 | 1 | 0,016 |

| | | | | | | | | |
|---|---|---|---|---|---|---|---|---|
| Insulin resistance | 129 | 1,49 | 2 | 0,43922 | 0,36 | 1 | 1 | 0,059 |
| Glucagon signaling pathway | 133 | 1,53 | 2 | 0,45479 | 0,34 | 1 | 1 | 0,000 |
| Type II diabetes mellitus | 53 | 0,61 | 1 | 0,45957 | 0,34 | 1 | 1 | 0,097 |
| Lysosome | 137 | 1,58 | 2 | 0,47011 | 0,33 | 1 | 1 | 0,000 |
| Sphingolipid signaling pathway | 137 | 1,58 | 2 | 0,47011 | 0,33 | 1 | 1 | 0,037 |
| Measles | 140 | 1,61 | 2 | 0,48143 | 0,32 | 1 | 1 | 0,034 |
| Insulin signaling pathway | 142 | 1,63 | 2 | 0,4889 | 0,31 | 1 | 1 | 0,060 |
| Arachidonic acid metabolism | 142 | 1,63 | 2 | 0,4889 | 0,31 | 1 | 1 | 0,147 |
| Autoimmune thyroid disease | 58 | 0,67 | 1 | 0,49014 | 0,31 | 1 | 1 | 0,024 |
| AMPK signaling pathway | 145 | 1,67 | 2 | 0,49998 | 0,30 | 1 | 1 | 0,010 |
| Regulation of actin cytoskeleton | 237 | 2,73 | 3 | 0,51655 | 0,29 | 1 | 1 | 0,103 |
| Viral life cycle - HIV-1 | 64 | 0,74 | 1 | 0,52457 | 0,28 | 1 | 1 | 0,000 |
| One carbon pool by folate | 65 | 0,75 | 1 | 0,53008 | 0,28 | 1 | 1 | 0,011 |
| VEGF signaling pathway | 66 | 0,76 | 1 | 0,53553 | 0,27 | 1 | 1 | 0,273 |
| Chemical carcinogenesis - receptor activation | 244 | 2,81 | 3 | 0,53628 | 0,27 | 1 | 1 | 0,044 |
| Human papillomavirus infection | 336 | 3,87 | 4 | 0,54448 | 0,26 | 1 | 1 | 0,050 |
| Cell adhesion molecules | 158 | 1,82 | 2 | 0,54618 | 0,26 | 1 | 1 | 0,019 |
| Glycerophospholipid metabolism | 159 | 1,83 | 2 | 0,54961 | 0,26 | 1 | 1 | 0,136 |
| Long-term depression | 69 | 0,79 | 1 | 0,5515 | 0,26 | 1 | 1 | 0,000 |
| Acute myeloid leukemia | 69 | 0,79 | 1 | 0,5515 | 0,26 | 1 | 1 | 0,021 |
| cAMP signaling pathway | 251 | 2,89 | 3 | 0,55555 | 0,26 | 1 | 1 | 0,031 |
| alpha-Linolenic acid metabolism | 70 | 0,81 | 1 | 0,5567 | 0,25 | 1 | 1 | 0,038 |
| mTOR signaling pathway | 162 | 1,87 | 2 | 0,55979 | 0,25 | 1 | 1 | 0,036 |
| Salmonella infection | 256 | 2,95 | 3 | 0,56903 | 0,24 | 1 | 1 | 0,041 |
| Ferroptosis | 73 | 0,84 | 1 | 0,57194 | 0,24 | 1 | 1 | 0,000 |
| Renal cell carcinoma | 73 | 0,84 | 1 | 0,57194 | 0,24 | 1 | 1 | 0,018 |
| Oxytocin signaling pathway | 167 | 1,92 | 2 | 0,5764 | 0,24 | 1 | 1 | 0,066 |
| Propanoate metabolism | 74 | 0,85 | 1 | 0,57691 | 0,24 | 1 | 1 | 0,000 |
| GnRH secretion | 74 | 0,85 | 1 | 0,57691 | 0,24 | 1 | 1 | 0,000 |
| Butanoate metabolism | 74 | 0,85 | 1 | 0,57691 | 0,24 | 1 | 1 | 0,050 |
| Fatty acid biosynthesis | 76 | 0,88 | 1 | 0,58667 | 0,23 | 1 | 1 | 0,000 |
| Ovarian steroidogenesis | 76 | 0,88 | 1 | 0,58667 | 0,23 | 1 | 1 | 0,000 |
| Starch and sucrose metabolism | 77 | 0,89 | 1 | 0,59147 | 0,23 | 1 | 1 | 0,171 |
| Galactose metabolism | 78 | 0,90 | 1 | 0,59621 | 0,22 | 1 | 1 | 0,042 |
| Carbohydrate digestion and absorption | 79 | 0,91 | 1 | 0,60089 | 0,22 | 1 | 1 | 0,000 |
| Pyruvate metabolism | 79 | 0,91 | 1 | 0,60089 | 0,22 | 1 | 1 | 0,036 |
| Fc epsilon RI signaling pathway | 80 | 0,92 | 1 | 0,60553 | 0,22 | 1 | 1 | 0,019 |
| Glioma | 80 | 0,92 | 1 | 0,60553 | 0,22 | 1 | 1 | 0,137 |
| Antigen processing and presentation | 81 | 0,93 | 1 | 0,6101 | 0,21 | 1 | 1 | 0,026 |
| PPAR signaling pathway | 81 | 0,93 | 1 | 0,6101 | 0,21 | 1 | 1 | 0,034 |
| Hepatocellular carcinoma | 184 | 2,12 | 2 | 0,62945 | 0,20 | 1 | 1 | 0,042 |
| Renin secretion | 86 | 0,99 | 1 | 0,63222 | 0,20 | 1 | 1 | 0,018 |
| Neomycin, kanamycin and gentamicin biosynthesis | 86 | 0,99 | 1 | 0,63222 | 0,20 | 1 | 1 | 0,024 |
| Folate biosynthesis | 86 | 0,99 | 1 | 0,63222 | 0,20 | 1 | 1 | 0,066 |
| Arrhythmogenic right ventricular cardiomyopathy | 88 | 1,01 | 1 | 0,64072 | 0,19 | 1 | 1 | 0,000 |
| Colorectal cancer | 88 | 1,01 | 1 | 0,64072 | 0,19 | 1 | 1 | 0,081 |
| Fructose and mannose metabolism | 89 | 1,02 | 1 | 0,64489 | 0,19 | 1 | 1 | 0,000 |
| Glycine, serine and threonine metabolism | 89 | 1,02 | 1 | 0,64489 | 0,19 | 1 | 1 | 0,000 |
| ECM-receptor interaction | 90 | 1,04 | 1 | 0,64902 | 0,19 | 1 | 1 | 0,014 |
| Synaptic vesicle cycle | 91 | 1,05 | 1 | 0,6531 | 0,19 | 1 | 1 | 0,000 |
| Fatty acid degradation | 93 | 1,07 | 1 | 0,66111 | 0,18 | 1 | 1 | 0,000 |
| Pentose and glucuronate interconversions | 95 | 1,09 | 1 | 0,66895 | 0,17 | 1 | 1 | 0,000 |
| Glyoxylate and dicarboxylate metabolism | 95 | 1,09 | 1 | 0,66895 | 0,17 | 1 | 1 | 0,011 |
| B cell receptor signaling pathway | 95 | 1,09 | 1 | 0,66895 | 0,17 | 1 | 1 | 0,019 |
| Th1 and Th2 cell differentiation | 96 | 1,11 | 1 | 0,6728 | 0,17 | 1 | 1 | 0,141 |
| Glutathione metabolism | 97 | 1,12 | 1 | 0,6766 | 0,17 | 1 | 1 | 0,029 |
| Insulin secretion | 98 | 1,13 | 1 | 0,68036 | 0,17 | 1 | 1 | 0,000 |
| Longevity regulating pathway | 98 | 1,13 | 1 | 0,68036 | 0,17 | 1 | 1 | 0,037 |
| Epstein-Barr virus infection | 207 | 2,38 | 2 | 0,6929 | 0,16 | 1 | 1 | 0,027 |
| Glycerolipid metabolism | 103 | 1,19 | 1 | 0,69853 | 0,16 | 1 | 1 | 0,035 |
| Gap junction | 103 | 1,19 | 1 | 0,69853 | 0,16 | 1 | 1 | 0,058 |
| Staphylococcus aureus infection | 104 | 1,20 | 1 | 0,70203 | 0,15 | 1 | 1 | 0,023 |
| MicroRNAs in cancer | 312 | 3,59 | 3 | 0,70208 | 0,15 | 1 | 1 | 0,016 |
| Melanogenesis | 107 | 1,23 | 1 | 0,71232 | 0,15 | 1 | 1 | 0,023 |
| Fc gamma R-mediated phagocytosis | 107 | 1,23 | 1 | 0,71232 | 0,15 | 1 | 1 | 0,036 |
| Dilated cardiomyopathy | 108 | 1,24 | 1 | 0,71567 | 0,15 | 1 | 1 | 0,019 |
| Human immunodeficiency virus 1 infection | 218 | 2,51 | 2 | 0,71998 | 0,14 | 1 | 1 | 0,045 |
| Tyrosine metabolism | 114 | 1,31 | 1 | 0,73496 | 0,13 | 1 | 1 | 0,000 |

| | | | | | | | | |
|---|---|---|---|---|---|---|---|---|
| C-type lectin receptor signaling pathway | 117 | 1,35 | 1 | 0,74412 | 0,13 | 1 | 1 | 0,053 |
| Complement and coagulation cascades | 119 | 1,37 | 1 | 0,75005 | 0,12 | 1 | 1 | 0,016 |
| Aldosterone synthesis and secretion | 120 | 1,38 | 1 | 0,75296 | 0,12 | 1 | 1 | 0,000 |
| Glutamatergic synapse | 124 | 1,43 | 1 | 0,76428 | 0,12 | 1 | 1 | 0,028 |
| Tryptophan metabolism | 125 | 1,44 | 1 | 0,76703 | 0,12 | 1 | 1 | 0,000 |
| Neurotrophin signaling pathway | 125 | 1,44 | 1 | 0,76703 | 0,12 | 1 | 1 | 0,012 |
| T cell receptor signaling pathway | 126 | 1,45 | 1 | 0,76975 | 0,11 | 1 | 1 | 0,042 |
| Diabetic cardiomyopathy | 244 | 2,81 | 2 | 0,77612 | 0,11 | 1 | 1 | 0,021 |
| Inflammatory mediator regulation of TRP channels | 134 | 1,54 | 1 | 0,79038 | 0,10 | 1 | 1 | 0,034 |
| Endocytosis | 258 | 2,97 | 2 | 0,80212 | 0,10 | 1 | 1 | 0,044 |
| Platelet activation | 140 | 1,61 | 1 | 0,80465 | 0,09 | 1 | 1 | 0,011 |
| Oocyte meiosis | 143 | 1,65 | 1 | 0,81142 | 0,09 | 1 | 1 | 0,024 |
| Bile secretion | 264 | 3,04 | 2 | 0,81243 | 0,09 | 1 | 1 | 0,000 |
| Systemic lupus erythematosus | 144 | 1,66 | 1 | 0,81362 | 0,09 | 1 | 1 | 0,000 |
| Apelin signaling pathway | 149 | 1,72 | 1 | 0,82426 | 0,08 | 1 | 1 | 0,027 |
| Serotonergic synapse | 157 | 1,81 | 1 | 0,84005 | 0,08 | 1 | 1 | 0,000 |
| Chemical carcinogenesis - reactive oxygen species | 284 | 3,27 | 2 | 0,84344 | 0,07 | 1 | 1 | 0,019 |
| Pathways of neurodegeneration - multiple diseases | 515 | 5,93 | 4 | 0,8512 | 0,07 | 1 | 1 | 0,020 |
| Steroid hormone biosynthesis | 164 | 1,89 | 1 | 0,8527 | 0,07 | 1 | 1 | 0,005 |
| Cushing syndrome | 168 | 1,93 | 1 | 0,85948 | 0,07 | 1 | 1 | 0,017 |
| Amino sugar and nucleotide sugar metabolism | 169 | 1,95 | 1 | 0,86113 | 0,06 | 1 | 1 | 0,000 |
| Tight junction | 171 | 1,97 | 1 | 0,86436 | 0,06 | 1 | 1 | 0,022 |
| Autophagy - animal | 175 | 2,01 | 1 | 0,87061 | 0,06 | 1 | 1 | 0,000 |
| Wnt signaling pathway | 175 | 2,01 | 1 | 0,87061 | 0,06 | 1 | 1 | 0,011 |
| cGMP-PKG signaling pathway | 176 | 2,03 | 1 | 0,87213 | 0,06 | 1 | 1 | 0,012 |
| Alcoholism | 198 | 2,28 | 1 | 0,90138 | 0,05 | 1 | 1 | 0,010 |
| Neutrophil extracellular trap formation | 206 | 2,37 | 1 | 0,91028 | 0,04 | 1 | 1 | 0,009 |
| Human T-cell leukemia virus 1 infection | 229 | 2,64 | 1 | 0,93168 | 0,03 | 1 | 1 | 0,015 |
| Cytoskeleton in muscle cells | 232 | 2,67 | 1 | 0,93407 | 0,03 | 1 | 1 | 0,000 |
| Amyotrophic lateral sclerosis | 385 | 4,43 | 2 | 0,94003 | 0,03 | 1 | 1 | 0,018 |
| Alzheimer disease | 410 | 4,72 | 2 | 0,95318 | 0,02 | 1 | 1 | 0,019 |
| Prion disease | 283 | 3,26 | 1 | 0,96406 | 0,02 | 1 | 1 | 0,000 |
| Parkinson disease | 297 | 3,42 | 1 | 0,96959 | 0,01 | 1 | 1 | 0,007 |

**Table S6. Joint pathway enrichment analysis of key proteomic and metabolomic features in metabolic pathways.** This table details the functional enrichment of key molecular features identified through integrated multi-omic models. Analysis was performed using MetaboAnalyst 6.0 Joint Pathway Analysis based on the KEGG database with metabolic pathway datasets and Overall Combine p values method.

| | Total | Hits.cmpd | Hits.gene | Raw p | (-LOG10(p)) | Holm adjust | FDR | Impact | Enrichment Ratio |
|---|---|---|---|---|---|---|---|---|---|
| Alanine, aspartate and glutamate metabolism | 61 | 5 | 0 | 0,0003 | 3,5078 | 0,0109 | 0,0099 | 0,4167 | 5,4379 |
| Valine, leucine and isoleucine biosynthesis | 12 | 3 | 0 | 0,0006 | 3,2494 | 0,0191 | 0,0099 | 0,3636 | 16,5856 |
| Pyrimidine metabolism | 103 | 5 | 0 | 0,0015 | 2,8192 | 0,0500 | 0,0177 | 0,2059 | 3,2204 |
| Riboflavin metabolism | 9 | 2 | 0 | 0,0029 | 2,5311 | 0,0942 | 0,0194 | 0,8750 | 14,7427 |
| Arginine biosynthesis | 28 | 3 | 0 | 0,0033 | 2,4775 | 0,1032 | 0,0194 | 0,1852 | 7,1082 |
| Purine metabolism | 170 | 6 | 1 | 0,0035 | 2,4564 | 0,1049 | 0,0194 | 0,1420 | 2,7317 |
| Aminoacyl-tRNA biosynthesis | 74 | 5 | 0 | 0,0039 | 2,4105 | 0,1127 | 0,0194 | 0,0685 | 4,4827 |
| Caffeine metabolism | 19 | 2 | 0 | 0,0203 | 1,6935 | 0,5671 | 0,0886 | 0,3889 | 6,9835 |
| Lysine degradation | 57 | 3 | 0 | 0,0289 | 1,5388 | 0,7809 | 0,1125 | 0,1250 | 3,4917 |
| Nicotinate and nicotinamide metabolism | 42 | 2 | 0 | 0,0440 | 1,3565 | 1 | 0,1429 | 0,1220 | 3,1592 |
| Neomycin, kanamycin and gentamicin biosynthesis | 4 | 1 | 0 | 0,0451 | 1,3462 | 1 | 0,1429 | 0,6667 | 16,5854 |
| Histidine metabolism | 32 | 2 | 0 | 0,0496 | 1,3047 | 1 | 0,1429 | 0,0645 | 4,1464 |
| Valine, leucine and isoleucine degradation | 87 | 3 | 0 | 0,0531 | 1,2750 | 1 | 0,1429 | 0,0814 | 2,2876 |
| Folate biosynthesis | 61 | 0 | 1 | 0,0710 | 1,1488 | 1 | 0,1731 | 0,0333 | 1,0876 |
| Pantothenate and CoA biosynthesis | 36 | 2 | 0 | 0,0742 | 1,1297 | 1 | 0,1731 | 0,0857 | 3,6857 |
| beta-Alanine metabolism | 44 | 2 | 0 | 0,0809 | 1,0923 | 1 | 0,1769 | 0,0698 | 3,0155 |
| Linoleic acid metabolism | 17 | 1 | 0 | 0,1090 | 0,9627 | 1 | 0,2244 | 0,0625 | 3,9024 |
| Nitrogen metabolism | 10 | 1 | 0 | 0,1294 | 0,8882 | 1 | 0,2515 | 0,1111 | 6,6344 |
| Vitamin B6 metabolism | 21 | 1 | 0 | 0,1878 | 0,7264 | 1 | 0,3132 | 0,2000 | 3,1592 |
| Biosynthesis of unsaturated fatty acids | 47 | 2 | 0 | 0,1969 | 0,7058 | 1 | 0,3132 | 0,0435 | 2,8231 |
| Arginine and proline metabolism | 74 | 2 | 0 | 0,1969 | 0,7058 | 1 | 0,3132 | 0,1507 | 1,7931 |
| Glycerophospholipid metabolism | 87 | 2 | 0 | 0,1969 | 0,7058 | 1 | 0,3132 | 0,1861 | 1,5251 |
| alpha-Linolenic acid metabolism | 22 | 1 | 0 | 0,2598 | 0,5854 | 1 | 0,3865 | 0,0952 | 3,0156 |
| Arachidonic acid metabolism | 95 | 2 | 0 | 0,2650 | 0,5767 | 1 | 0,3865 | 0,2234 | 1,3966 |
| Butanoate metabolism | 29 | 1 | 0 | 0,2934 | 0,5325 | 1 | 0,4108 | 0,1071 | 2,2876 |
| Glycerolipid metabolism | 34 | 1 | 0 | 0,3097 | 0,5091 | 1 | 0,4168 | 0,0606 | 1,9512 |
| Starch and sucrose metabolism | 43 | 1 | 0 | 0,3411 | 0,4671 | 1 | 0,4421 | 0,3333 | 1,5428 |
| Pentose phosphate pathway | 49 | 1 | 0 | 0,4137 | 0,3833 | 1 | 0,4993 | 0,0417 | 1,3539 |
| Pyruvate metabolism | 48 | 1 | 0 | 0,4137 | 0,3833 | 1 | 0,4993 | 0,0426 | 1,3821 |
| One carbon pool by folate | 72 | 1 | 0 | 0,4534 | 0,3435 | 1 | 0,5075 | 0,0141 | 0,9214 |
| Glycolysis or Gluconeogenesis | 60 | 1 | 0 | 0,4534 | 0,3435 | 1 | 0,5075 | 0,0339 | 1,1057 |
| Galactose metabolism | 51 | 1 | 0 | 0,4661 | 0,3315 | 1 | 0,5075 | 0,0600 | 1,3008 |
| Glutathione metabolism | 57 | 1 | 0 | 0,4785 | 0,3201 | 1 | 0,5075 | 0,0357 | 1,1639 |
| Glyoxylate and dicarboxylate metabolism | 56 | 1 | 0 | 0,5253 | 0,2796 | 1 | 0,5407 | 0,0182 | 1,1847 |
| Steroid hormone biosynthesis | 208 | 1 | 0 | 0,8729 | 0,0591 | 1 | 0,8729 | 0,0048 | 0,3189 |

**Table S7. The association between ADA protein levels and anyNDC status evaluated by GEE model.**

| | | GEE[1]: cond = F (across individuals) | | | | CGEE[2]: cond = T (within-twins) | | | |
|---|---|---|---|---|---|---|---|---|---|
| Sample | Population | Estimate | SE | p value | Sample size | Estimate | SE | p value | Sample size |
| Serum | All ind | 0,66 | 0,18 | **0,0002** | 109 individuals, 62 clusters | 0,37 | 0,15 | **0,0150** | 94 individuals, 47 clusters |
| | All MZ | 0,60 | 0,19 | **0,0013** | 94 individuals, 51 clusters | 0,45 | 0,17 | **0,0048** | 86 individuals, 43 clusters |
| | Discordant MZ | 0,49 | 0,17 | **0,0040** | 26 individuals, 14 clusters | 0,54 | 0,16 | **0,0009** | 24 individuals, 12 clusters |
| CSF | All ind | -0,14 | 0,25 | 0,59 | 86 individuals 47 cluster | -0,07 | 0,34 | 0,84 | 78 individuals, 39 cluster |
| | MZ | -0,18 | 0,29 | 0,54 | 70 individuals,38 clusters | 0,66 | 0,42 | 0,116 | 64 individuals, 32 clusters |
| | Discordant MZ | 0,96 | 0,59 | 0,10 | 13 individuals, 8 clusters | / | / | / | 10 individuals, 5 clusters |

1 GEE Model: Level(molecule) ~ AnyNDC + Age + Sex + Medication + BMI + Other_psychiatric

2 CGEE Model: Level(molecule) ~ AnyNDC + Medication + BMI + Other_psychiatric

**Table S8. The association between levels of key fecal metabolite and anyNDC status evaluated by GEE model.** P values were adjusted by FDR method within each subsamples.

**All individuals**

| | GEE[1]: cond = F (across individuals) 189 individuals, 99 clusters | | | | CGEE[2]: cond = T (within-twins) 180 individuals, 90 clusters | | | |
|---|---|---|---|---|---|---|---|---|
| **Metabolite** | **Estimate** | **SE** | **p value** | **Adj. p value** | **Estimate** | **SE** | **p value** | **Adj. p value** |
| 2'-deoxyinosine | -0,4479 | 0,1820 | **0,0138** | 0,1453 | -0,2399 | 0,2312 | 0,2994 | 0,6286 |
| N-acetylglutamate | -0,2888 | 0,1732 | 0,0953 | 0,3662 | 0,2536 | 0,2120 | 0,2315 | 0,6006 |
| valine | 0,2397 | 0,2058 | 0,2442 | 0,3662 | 0,0701 | 0,2286 | 0,7591 | 0,9377 |
| riboflavin(VitaminB2) | 0,1309 | 0,1670 | 0,4331 | 0,5052 | 0,2857 | 0,2118 | 0,1774 | 0,6006 |
| leucine | 0,2681 | 0,1995 | 0,1791 | 0,3662 | 0,1152 | 0,2589 | 0,6562 | 0,9377 |
| flavinmononucleotide(FMN) | -0,5607 | 0,1790 | **0,0017** | **0,0364** | 0,0167 | 0,2231 | 0,9403 | 0,9560 |
| 2-aminoadipate | 0,2912 | 0,1497 | 0,0517 | 0,3616 | 0,2984 | 0,2552 | 0,2422 | 0,6006 |
| thymidine | -0,2168 | 0,1714 | 0,2059 | 0,3662 | -0,0134 | 0,2432 | 0,9560 | 0,9560 |
| N-carbamoylaspartate | 0,2356 | 0,1749 | 0,1780 | 0,3662 | 0,4459 | 0,2513 | 0,0760 | 0,4992 |
| glutamine | 0,0995 | 0,1623 | 0,5398 | 0,5966 | -0,2038 | 0,2374 | 0,3905 | 0,6835 |
| 2'-deoxyuridine | -0,2086 | 0,1614 | 0,1963 | 0,3662 | 0,0784 | 0,2463 | 0,7504 | 0,9377 |
| deoxycarnitine | -0,1901 | 0,1912 | 0,3203 | 0,4484 | -0,8536 | 0,3726 | **0,0220** | 0,4617 |
| guanosine | 0,2552 | 0,1777 | 0,1509 | 0,3662 | 0,2837 | 0,3209 | 0,3766 | 0,6835 |
| xanthosine | 0,2877 | 0,2140 | 0,1789 | 0,3662 | 0,3937 | 0,2241 | 0,0790 | 0,4992 |
| adenosine | 0,2394 | 0,2054 | 0,2438 | 0,3662 | 0,0442 | 0,1058 | 0,6758 | 0,9377 |
| inosine | 0,0966 | 0,1949 | 0,6200 | 0,6510 | 0,1850 | 0,3415 | 0,5881 | 0,9377 |
| carnitine | -0,1508 | 0,1694 | 0,3732 | 0,4899 | -0,4230 | 0,2743 | 0,1231 | 0,5171 |
| aspartate | 0,0765 | 0,2004 | 0,7028 | 0,7028 | -0,0213 | 0,2362 | 0,9281 | 0,9560 |
| gamma-aminobutyrate(GABA) | 0,1789 | 0,1454 | 0,2184 | 0,3662 | -0,0279 | 0,1763 | 0,8745 | 0,9560 |
| thymine | -0,1648 | 0,2002 | 0,4105 | 0,5052 | 0,2204 | 0,1946 | 0,2574 | 0,6006 |
| N-acetylaspartate(NAA) | -0,2196 | 0,1694 | 0,1949 | 0,3662 | -0,2327 | 0,1394 | 0,0951 | 0,4992 |

**All MZ twins**

| | GEE[1]: cond = F (across individuals) 134 individuals, 70 clusters | | | | CGEE[2]: cond = T (within-twins) 128 individuals, 64 clusters | | | |
|---|---|---|---|---|---|---|---|---|
| **Metabolite** | **Estimate** | **SE** | **p value** | **Adj. p value** | **Estimate** | **SE** | **p value** | **Adj. p value** |
| 2'-deoxyinosine | -0,5588 | 0,2061 | **0,0067** | 0,0629 | -0,3990 | 0,2541 | 0,1163 | 0,4442 |
| N-acetylglutamate | -0,4089 | 0,2096 | 0,0511 | 0,2296 | -0,0713 | 0,2691 | 0,7911 | 0,9200 |
| valine | 0,2996 | 0,2492 | 0,2292 | 0,4376 | -0,1408 | 0,2685 | 0,6001 | 0,9200 |
| riboflavin(VitaminB2) | 0,1730 | 0,1973 | 0,3805 | 0,5167 | 0,0331 | 0,2346 | 0,8878 | 0,9200 |
| leucine | 0,3268 | 0,2360 | 0,1661 | 0,3876 | -0,1629 | 0,3002 | 0,5872 | 0,9200 |
| flavinmononucleotide(FMN) | -0,9165 | 0,1940 | **0,0000** | **0,0000** | -0,1757 | 0,2839 | 0,5360 | 0,9200 |
| 2-aminoadipate | 0,4005 | 0,1533 | **0,0090** | 0,0629 | 0,4501 | 0,2494 | 0,0711 | 0,4442 |
| thymidine | -0,2926 | 0,1844 | 0,1125 | 0,3876 | -0,2507 | 0,1752 | 0,1524 | 0,4442 |
| N-carbamoylaspartate | 0,1650 | 0,1946 | 0,3965 | 0,5167 | 0,5354 | 0,3547 | 0,1312 | 0,4442 |
| glutamine | 0,1770 | 0,1854 | 0,3399 | 0,5167 | -0,4988 | 0,2521 | **0,0479** | 0,4442 |
| 2'-deoxyuridine | -0,3436 | 0,1788 | 0,0547 | 0,2296 | -0,3097 | 0,2115 | 0,1430 | 0,4442 |
| deoxycarnitine | 0,1233 | 0,1564 | 0,4307 | 0,5167 | -0,0389 | 0,2471 | 0,8748 | 0,9200 |
| guanosine | 0,2488 | 0,1727 | 0,1498 | 0,3876 | 0,1330 | 0,2815 | 0,6366 | 0,9200 |
| xanthosine | 0,3510 | 0,2658 | 0,1867 | 0,3921 | 0,1959 | 0,2379 | 0,4102 | 0,8614 |
| adenosine | 0,0667 | 0,0883 | 0,4501 | 0,5167 | 0,0079 | 0,0782 | 0,9200 | 0,9200 |
| inosine | 0,1325 | 0,2095 | 0,5270 | 0,5270 | 0,0459 | 0,2450 | 0,8514 | 0,9200 |
| carnitine | 0,1254 | 0,1825 | 0,4921 | 0,5167 | 0,0305 | 0,2331 | 0,8958 | 0,9200 |
| aspartate | 0,1645 | 0,2335 | 0,4813 | 0,5167 | -0,3995 | 0,2906 | 0,1692 | 0,4442 |
| gamma-aminobutyrate(GABA) | 0,1782 | 0,1860 | 0,3379 | 0,5167 | -0,2101 | 0,2395 | 0,3804 | 0,8614 |
| thymine | -0,3380 | 0,2320 | 0,1451 | 0,3876 | -0,0398 | 0,2200 | 0,8565 | 0,9200 |
| N-acetylaspartate(NAA) | -0,1905 | 0,2159 | 0,3777 | 0,5167 | -0,3911 | 0,2238 | 0,0806 | 0,4442 |

**Discordant MZ twins**

| | GEE[1]: cond = F (across individuals) 42 individuals, 22 clusters | | | | CGEE[2]: cond = T (within-twins) 40 individuals, 20 clusters | | | |
|---|---|---|---|---|---|---|---|---|
| **Metabolite** | **Estimate** | **SE** | **p value** | **Adj. p value** | **Estimate** | **SE** | **p value** | **Adj. p value** |
| 2'-deoxyinosine | -0,0870 | 0,2689 | 0,7464 | 0,7464 | -0,3604 | 0,2766 | 0,1926 | 0,5779 |
| N-acetylglutamate | 0,1282 | 0,3039 | 0,6730 | 0,7439 | -0,3106 | 0,2690 | 0,2482 | 0,5790 |
| valine | -0,2715 | 0,2082 | 0,1923 | 0,5227 | -0,3687 | 0,2165 | 0,0885 | 0,3718 |
| riboflavin(VitaminB2) | 0,2439 | 0,2177 | 0,2626 | 0,6069 | 0,0142 | 0,1862 | 0,9390 | 0,9390 |
| leucine | -0,2591 | 0,2444 | 0,2890 | 0,6069 | -0,4046 | 0,2795 | 0,1477 | 0,5170 |
| flavinmononucleotide(FMN) | -0,5706 | 0,2717 | **0,0357** | 0,2502 | 0,1897 | 0,2942 | 0,5191 | 0,7482 |
| 2-aminoadipate | 0,2803 | 0,2004 | 0,1619 | 0,5227 | 0,1115 | 0,2394 | 0,6413 | 0,7482 |
| thymidine | -0,1794 | 0,2253 | 0,4258 | 0,7166 | -0,4065 | 0,1301 | **0,0018** | **0,0287** |
| N-carbamoylaspartate | 0,1873 | 0,2932 | 0,5230 | 0,7381 | 0,4532 | 0,3813 | 0,2346 | 0,5790 |
| glutamine | -0,5424 | 0,2094 | **0,0096** | 0,2016 | -0,7061 | 0,2356 | **0,0027** | **0,0287** |
| 2'-deoxyuridine | -0,1952 | 0,2548 | 0,4436 | 0,7166 | -0,4894 | 0,1765 | **0,0056** | **0,0390** |
| deoxycarnitine | -0,3554 | 0,2768 | 0,1991 | 0,5227 | -0,1189 | 0,2359 | 0,6141 | 0,7482 |
| guanosine | 0,2393 | 0,3100 | 0,4402 | 0,7166 | -0,0949 | 0,1922 | 0,6215 | 0,7482 |
| xanthosine | 0,2689 | 0,1708 | 0,1154 | 0,4848 | 0,1501 | 0,2149 | 0,4847 | 0,7482 |
| adenosine | 0,0533 | 0,1116 | 0,6326 | 0,7381 | -0,0542 | 0,0610 | 0,3743 | 0,7146 |
| inosine | 0,0862 | 0,2357 | 0,7147 | 0,7464 | -0,1552 | 0,1960 | 0,4283 | 0,7482 |
| carnitine | -0,1609 | 0,2781 | 0,5629 | 0,7381 | -0,0579 | 0,2304 | 0,8015 | 0,8567 |
| aspartate | -0,4859 | 0,2886 | 0,0923 | 0,4845 | -0,5936 | 0,2435 | **0,0148** | 0,0776 |
| gamma-aminobutyrate(GABA) | -0,1679 | 0,2923 | 0,5658 | 0,7381 | -0,1097 | 0,1905 | 0,5646 | 0,7482 |
| thymine | -0,1123 | 0,2307 | 0,6264 | 0,7381 | 0,0492 | 0,2114 | 0,8159 | 0,8567 |
| N-acetylaspartate(NAA) | -0,3684 | 0,1749 | **0,0351** | 0,2502 | -0,1023 | 0,1106 | 0,3551 | 0,7146 |

1 GEE Model: Level(molecule) ~ AnyNDC + Age + Sex + Medication + BMI + Other_psychiatric

2 CGEE Model: Level(molecule) ~ AnyNDC + Medication + BMI + Other_psychiatric

**Table S9. The association between levels of key urine metabolite and anyNDC status evaluated by GEE model.** P values were adjusted by FDR method within each subsamples.

**All individuals**

| | GEE[1]: cond = F (across individuals) 112 individuals, 57 clusters | | | | CGEE[2]: cond = T (within-twins) 110 individuals, 55 clusters | | | |
|---|---|---|---|---|---|---|---|---|
| **Metabolite** | **Estimate** | **SE** | **p value** | **Adj. p value** | **Estimate** | **SE** | **p value** | **Adj. p value** |
| valine | -0,2157 | 0,2101 | 0,3044 | 0,3519 | 0,1283 | 0,2991 | 0,6680 | 0,7502 |
| isoleucine | -0,2175 | 0,2336 | 0,3519 | 0,3519 | 0,1084 | 0,3405 | 0,7502 | 0,7502 |

**All MZ twins**

| | GEE[1]: cond = F (across individuals) 77 individuals, 39 clusters | | | | CGEE[2]: cond = T (within-twins) 76 individuals, 38 clusters | | | |
|---|---|---|---|---|---|---|---|---|
| **Metabolite** | **Estimate** | **SE** | **p value** | **Adj. p value** | **Estimate** | **SE** | **p value** | **Adj. p value** |
| valine | -0,3330 | 0,2909 | 0,2523 | 0,5045 | -0,0600 | 0,4698 | 0,8984 | 0,8984 |
| isoleucine | -0,0186 | 0,3354 | 0,9558 | 0,9558 | -0,1033 | 0,4452 | 0,8164 | 0,8984 |

1 GEE Model: Level(molecule) ~ AnyNDC + Age + Sex + Medication + BMI + Other_psychiatric
2 CGEE Model: Level(molecule) ~ AnyNDC + Medication + BMI + Other_psychiatric
* Discordant MZ twins (n = 5 twin pairs) here is too less to perform GEE model analysis.

**Table S10. The association between metal levels and anyNDC status evaluated by GEE model.** P values were adjusted by FDR method within each subsamples.

**All individuals**

| | GEE[1]: cond = F (across individuals) 220 individuals, 115 clusters | | | | CGEE[2]: cond = T (within-twins) 210 individuals, 105 clusters | | | |
|---|---|---|---|---|---|---|---|---|
| **Metal** | **Estimate** | **SE** | **p value** | **Adj. p value** | **Estimate** | **SE** | **p value** | **Adj. p value** |
| BMg | 0,3696 | 0,1998 | 0,0643 | 0,2787 | 0,0668 | 0,0820 | 0,4154 | 0,7593 |
| BCa | -0,1857 | 0,2174 | 0,3928 | 0,6383 | 0,0541 | 0,0827 | 0,5129 | 0,7593 |
| BMn | 0,4556 | 0,1628 | **0,0051** | 0,0668 | -0,1753 | 0,1448 | 0,2261 | 0,7593 |
| BFe | 0,2859 | 0,2168 | 0,1872 | 0,4056 | -0,0239 | 0,0842 | 0,7761 | 0,7761 |
| BCo | 0,0068 | 0,1389 | 0,9610 | 0,9610 | -0,0768 | 0,1910 | 0,6876 | 0,7593 |
| BCu | -0,0343 | 0,1636 | 0,8338 | 0,9610 | -0,1510 | 0,2188 | 0,4900 | 0,7593 |
| BZn | 0,3021 | 0,2058 | 0,1421 | 0,4056 | -0,0929 | 0,0833 | 0,2648 | 0,7593 |
| BAs | -0,1333 | 0,1347 | 0,3224 | 0,5988 | -0,1274 | 0,1379 | 0,3555 | 0,7593 |
| BSe | 0,3718 | 0,1691 | **0,0279** | 0,1815 | -0,1174 | 0,0990 | 0,2355 | 0,7593 |
| BMo | -0,0693 | 0,1587 | 0,6624 | 0,8611 | 0,0566 | 0,1474 | 0,7009 | 0,7593 |
| BCd | 0,0979 | 0,1554 | 0,5286 | 0,7635 | -0,0528 | 0,1198 | 0,6595 | 0,7593 |
| BHg | -0,0184 | 0,1662 | 0,9117 | 0,9610 | -0,0989 | 0,1567 | 0,5279 | 0,7593 |
| BPb | 0,2027 | 0,1462 | 0,1655 | 0,4056 | -0,1894 | 0,1826 | 0,2997 | 0,7593 |

**MZ twins**

| | GEE[1]: cond = F (across individuals) 149 individuals, 76 clusters | | | | CGEE[2]: cond = T (within-twins) 146 individuals, 73 clusters | | | |
|---|---|---|---|---|---|---|---|---|
| **Metal** | **Estimate** | **SE** | **p value** | **Adj. p value** | **Estimate** | **SE** | **p value** | **Adj. p value** |
| BMg | 0,4469 | 0,2366 | 0,0589 | 0,2239 | 0,0182 | 0,0695 | 0,7939 | 0,9755 |
| BCa | -0,3350 | 0,2539 | 0,1870 | 0,4052 | -0,0065 | 0,0913 | 0,9434 | 0,9755 |
| BMn | 0,7156 | 0,1981 | **0,0003** | **0,0039** | 0,0535 | 0,1086 | 0,6224 | 0,9755 |
| BFe | 0,4368 | 0,2599 | 0,0928 | 0,2412 | 0,0180 | 0,1157 | 0,8764 | 0,9755 |
| BCo | 0,1007 | 0,1692 | 0,5517 | 0,6156 | -0,0522 | 0,1586 | 0,7420 | 0,9755 |
| BCu | 0,0105 | 0,2046 | 0,9591 | 0,9591 | 0,2094 | 0,1470 | 0,1542 | 0,9755 |
| BZn | 0,4840 | 0,2661 | 0,0689 | 0,2239 | -0,0102 | 0,0869 | 0,9064 | 0,9755 |
| BAs | -0,1767 | 0,1877 | 0,3463 | 0,5211 | -0,0978 | 0,1369 | 0,4749 | 0,9755 |
| BSe | 0,4300 | 0,1955 | **0,0279** | 0,1811 | 0,0066 | 0,1122 | 0,9531 | 0,9755 |
| BMo | -0,1965 | 0,2150 | 0,3607 | 0,5211 | -0,0929 | 0,2084 | 0,6557 | 0,9755 |
| BCd | 0,1586 | 0,2047 | 0,4384 | 0,5699 | -0,0026 | 0,0857 | 0,9755 | 0,9755 |
| BHg | -0,1596 | 0,1705 | 0,3492 | 0,5211 | -0,1360 | 0,1163 | 0,2423 | 0,9755 |
| BPb | 0,1134 | 0,1988 | 0,5683 | 0,6156 | -0,4344 | 0,2848 | 0,1273 | 0,9755 |

**DIscordant MZ twins**

| | GEE[1]: cond = F (across individuals) 44 individuals, 23 clusters | | | | CGEE[2]: cond = T (within-twins) 42 individuals, 21 clusters | | | |
|---|---|---|---|---|---|---|---|---|
| **Metal** | **Estimate** | **SE** | **p value** | **Adj. p value** | **Estimate** | **SE** | **p value** | **Adj. p value** |
| BMg | 0,0031 | 0,0640 | 0,9613 | 0,9613 | 0,0029 | 0,0717 | 0,9682 | 0,9682 |
| BCa | -0,0055 | 0,0942 | 0,9533 | 0,9613 | -0,0153 | 0,1024 | 0,8815 | 0,9682 |
| BMn | 0,1234 | 0,1329 | 0,3533 | 0,9613 | 0,1456 | 0,1242 | 0,2410 | 0,9682 |
| BFe | 0,0959 | 0,1193 | 0,4217 | 0,9613 | -0,1083 | 0,1186 | 0,3612 | 0,9682 |
| BCo | -0,0770 | 0,1610 | 0,6325 | 0,9613 | -0,0385 | 0,1923 | 0,8414 | 0,9682 |
| BCu | 0,0403 | 0,1821 | 0,8247 | 0,9613 | 0,3668 | 0,1225 | **0,0028** | **0,0358** |
| BZn | -0,0552 | 0,0856 | 0,5192 | 0,9613 | -0,0548 | 0,1009 | 0,5871 | 0,9682 |
| BAs | 0,0054 | 0,1117 | 0,9613 | 0,9613 | 0,0456 | 0,0888 | 0,6076 | 0,9682 |
| BSe | 0,0595 | 0,1403 | 0,6718 | 0,9613 | -0,0119 | 0,1262 | 0,9249 | 0,9682 |
| BMo | -0,0598 | 0,2509 | 0,8116 | 0,9613 | -0,0088 | 0,1718 | 0,9589 | 0,9682 |
| BCd | -0,1186 | 0,1375 | 0,3884 | 0,9613 | 0,0119 | 0,0969 | 0,9020 | 0,9682 |
| BHg | -0,0113 | 0,1743 | 0,9483 | 0,9613 | -0,0636 | 0,1299 | 0,6242 | 0,9682 |
| BPb | -0,6055 | 0,4550 | 0,1832 | 0,9613 | -0,2886 | 0,2066 | 0,1625 | 0,9682 |

1 GEE Model: Level(molecule) ~ AnyNDC + Age + Sex + Medication + BMI + Other_psychiatric
2 CGEE Model: Level(molecule) ~ AnyNDC + Medication + BMI + Other_psychiatric

**Table S11. Differential analysis for NDC subgroups.** The association between molecular levels and NDC diagnosis status evaluated by GEE model.

| Molecular Type | Molecule | Sample Group | Model | Diagnosis | Number of diagnosed individuals | Estimate | Std. Error | p value |
|---|---|---|---|---|---|---|---|---|
| Serum protein | ADA | Whole individuals | GEE | Autism | 25 | 0,0475 | 0,2057 | 0,8174 |
| Serum protein | ADA | Whole individuals | GEE | ADHD | 35 | 0,7767 | 0,1940 | **0,0001** |
| Serum protein | ADA | Whole individuals | CGEE | Autism | 21 | 0,0163 | 0,1185 | 0,8905 |
| Serum protein | ADA | Whole individuals | CGEE | ADHD | 31 | 0,4927 | 0,1940 | **0,0111** |
| Serum protein | ADA | MZ | GEE | Autism | 24 | -0,0042 | 0,2094 | 0,9840 |
| Serum protein | ADA | MZ | GEE | ADHD | 29 | 0,8752 | 0,1970 | **0,0000** |
| Serum protein | ADA | MZ | CGEE | Autism | 20 | 0,0744 | 0,1314 | 0,5712 |
| Serum protein | ADA | MZ | CGEE | ADHD | 27 | 0,5052 | 0,1965 | **0,0102** |
| Serum protein | ADA | Discordant MZ | GEE | Autism | 6 | -0,0773 | 0,2665 | 0,7718 |
| Serum protein | ADA | Discordant MZ | GEE | ADHD | 6 | 0,8480 | 0,3432 | **0,0135** |
| Serum protein | ADA | Discordant MZ | CGEE | Autism | 6 | 0,2253 | 0,2527 | 0,3726 |
| Serum protein | ADA | Discordant MZ | CGEE | ADHD | 5 | 0,6330 | 0,2321 | **0,0064** |
| Fecal metabolites | Flavin mononucleotide (FMN) | Whole individuals | GEE | Autism | 40 | -0,2695 | 0,1797 | 0,1336 |
| Fecal metabolites | Flavin mononucleotide (FMN) | Whole individuals | GEE | ADHD | 52 | -0,4103 | 0,2135 | 0,0546 |
| Fecal metabolites | Flavin mononucleotide (FMN) | MZ | GEE | Autism | 28 | -0,6134 | 0,2119 | **0,0038** |
| Fecal metabolites | Flavin mononucleotide (FMN) | MZ | GEE | ADHD | 30 | -0,8497 | 0,2368 | **0,0003** |
| Fecal metabolites | Thymidine | Discordant MZ | CGEE | Autism | 11 | -0,1489 | 0,2081 | 0,4744 |
| Fecal metabolites | Glutamine | Discordant MZ | CGEE | Autism | 11 | -0,0126 | 0,5374 | 0,9812 |
| Fecal metabolites | 2'-Deoxyuridine | Discordant MZ | CGEE | Autism | 11 | 0,0952 | 0,3099 | 0,7586 |
| Fecal metabolites | Thymidine | Discordant MZ | CGEE | ADHD | 8 | -0,1655 | 0,2499 | 0,5079 |
| Fecal metabolites | Glutamine | Discordant MZ | CGEE | ADHD | 8 | -1,2264 | 0,3763 | **0,0011** |
| Fecal metabolites | 2'-Deoxyuridine | Discordant MZ | CGEE | ADHD | 8 | -0,4179 | 0,3850 | 0,2778 |
| Metal ion | Blood Mn (BMn) | MZ | GEE | Autism | 37 | 0,4869 | 0,2651 | 0,0663 |
| Metal ion | Blood Mn (BMn) | MZ | GEE | ADHD | 38 | 0,6473 | 0,2171 | **0,0029** |
| Metal ion | Blood Cu (BCu) | Discordant MZ | CGEE | Autism | 37 | 0,2084 | 0,2105 | 0,3222 |
| Metal ion | Blood Cu (BCu) | Discordant MZ | CGEE | ADHD | 38 | 0,4291 | 0,2073 | **0,0384** |

GEE Model: Level(molecule) ~ NDC_diagnosis + Age + Sex + Medication + BMI + Other_psychiatric
CGEE Model: Level(molecule) ~ NDC_diagnosis + Medication + BMI + Other_psychiatric

**Table S12. Pathway enrichment analysis of metal-NDC-related genes.** Analysis was performed using the R package ClusterProfiler and p-values were adjusted by the FDR method.

| | Description | GeneRatio | BgRatio | RichFactor | FoldEnrichment | zScore | pvalue | p.adjust | qvalue | Count |
|---|---|---|---|---|---|---|---|---|---|---|
| hsa00980 | Metabolism of xenobiotics by cytochrome P450 | 21/255 | 79/8538 | 0,2658 | 8,9004 | 12,3774 | 6,6616E-15 | 1,9518E-12 | 1,0939E-12 | 21 |
| hsa00982 | Drug metabolism - cytochrome P450 | 20/255 | 73/8538 | 0,2740 | 9,1732 | 12,3047 | 1,5878E-14 | 2,3262E-12 | 1,3037E-12 | 20 |
| hsa04726 | Serotonergic synapse | 19/255 | 115/8538 | 0,1652 | 5,5319 | 8,5846 | 1,0218E-09 | 8,7145E-08 | 4,8840E-08 | 19 |
| hsa04724 | Glutamatergic synapse | 19/255 | 116/8538 | 0,1638 | 5,4842 | 8,5317 | 1,1897E-09 | 8,7145E-08 | 4,8840E-08 | 19 |
| hsa05208 | Chemical carcinogenesis - reactive oxygen species | 26/255 | 227/8538 | 0,1145 | 3,8350 | 7,5957 | 3,1266E-09 | 1,5560E-07 | 8,7205E-08 | 26 |
| hsa00140 | Steroid hormone biosynthesis | 14/255 | 63/8538 | 0,2222 | 7,4405 | 9,0022 | 3,1863E-09 | 1,5560E-07 | 8,7205E-08 | 14 |
| hsa00350 | Tyrosine metabolism | 11/255 | 36/8538 | 0,3056 | 10,2307 | 9,7377 | 4,2197E-09 | 1,7662E-07 | 9,8988E-08 | 11 |
| hsa05204 | Chemical carcinogenesis - DNA adducts | 14/255 | 71/8538 | 0,1972 | 6,6022 | 8,3166 | 1,6568E-08 | 6,0681E-07 | 3,4009E-07 | 14 |
| hsa04066 | HIF-1 signaling pathway | 17/255 | 110/8538 | 0,1545 | 5,1745 | 7,7316 | 2,2603E-08 | 7,3584E-07 | 4,1240E-07 | 17 |
| hsa00830 | Retinol metabolism | 13/255 | 68/8538 | 0,1912 | 6,4010 | 7,8455 | 8,0426E-08 | 2,3565E-06 | 1,3207E-06 | 13 |
| hsa04728 | Dopaminergic synapse | 17/255 | 132/8538 | 0,1288 | 4,3121 | 6,7286 | 3,4924E-07 | 9,3025E-06 | 5,2136E-06 | 17 |
| hsa05225 | Hepatocellular carcinoma | 19/255 | 170/8538 | 0,1118 | 3,7421 | 6,3363 | 6,6837E-07 | 1,6319E-05 | 9,1461E-06 | 19 |
| hsa05418 | Fluid shear stress and atherosclerosis | 17/255 | 141/8538 | 0,1206 | 4,0369 | 6,3798 | 9,0484E-07 | 2,0394E-05 | 1,1430E-05 | 17 |
| hsa04960 | Aldosterone-regulated sodium reabsorption | 9/255 | 38/8538 | 0,2368 | 7,9300 | 7,5118 | 1,2539E-06 | 2,6243E-05 | 1,4708E-05 | 9 |
| hsa05207 | Chemical carcinogenesis - receptor activation | 21/255 | 215/8538 | 0,0977 | 3,2704 | 5,9157 | 1,6114E-06 | 3,1477E-05 | 1,7641E-05 | 21 |
| hsa04020 | Calcium signaling pathway | 23/255 | 254/8538 | 0,0906 | 3,0319 | 5,7679 | 1,9006E-06 | 3,4805E-05 | 1,9506E-05 | 23 |
| hsa04014 | Ras signaling pathway | 22/255 | 238/8538 | 0,0924 | 3,0950 | 5,7513 | 2,2818E-06 | 3,9328E-05 | 2,2041E-05 | 22 |
| hsa04913 | Ovarian steroidogenesis | 10/255 | 52/8538 | 0,1923 | 6,4389 | 6,9023 | 2,4792E-06 | 4,0355E-05 | 2,2617E-05 | 10 |
| hsa04151 | PI3K-Akt signaling pathway | 28/255 | 362/8538 | 0,0773 | 2,5898 | 5,4232 | 3,2962E-06 | 5,0831E-05 | 2,8488E-05 | 28 |
| hsa01521 | EGFR tyrosine kinase inhibitor resistance | 12/255 | 80/8538 | 0,1500 | 5,0224 | 6,3419 | 3,8530E-06 | 5,6446E-05 | 3,1635E-05 | 12 |
| hsa04080 | Neuroactive ligand-receptor interaction | 28/255 | 370/8538 | 0,0757 | 2,5338 | 5,2923 | 5,0275E-06 | 7,0146E-05 | 3,9313E-05 | 28 |
| hsa05030 | Cocaine addiction | 9/255 | 49/8538 | 0,1837 | 6,1498 | 6,3429 | 1,1855E-05 | 1,5788E-04 | 8,8485E-05 | 9 |
| hsa04919 | Thyroid hormone signaling pathway | 14/255 | 122/8538 | 0,1148 | 3,8422 | 5,5477 | 1,5100E-05 | 1,9237E-04 | 1,0781E-04 | 14 |
| hsa05022 | Pathways of neurodegeneration - multiple diseases | 31/255 | 483/8538 | 0,0642 | 2,1490 | 4,5612 | 4,1979E-05 | 5,1021E-04 | 2,8595E-04 | 31 |
| hsa04024 | cAMP signaling pathway | 19/255 | 226/8538 | 0,0841 | 2,8149 | 4,8515 | 4,3533E-05 | 5,1021E-04 | 2,8595E-04 | 19 |
| hsa05142 | Chagas disease | 12/255 | 103/8538 | 0,1165 | 3,9009 | 5,1967 | 5,3172E-05 | 5,9921E-04 | 3,3583E-04 | 12 |
| hsa00480 | Glutathione metabolism | 9/255 | 59/8538 | 0,1525 | 5,1075 | 5,5547 | 5,5865E-05 | 6,0624E-04 | 3,3976E-04 | 9 |
| hsa04730 | Long-term depression | 9/255 | 60/8538 | 0,1500 | 5,0224 | 5,4858 | 6,4034E-05 | 6,5033E-04 | 3,6447E-04 | 9 |
| hsa04625 | C-type lectin receptor signaling pathway | 12/255 | 105/8538 | 0,1143 | 3,8266 | 5,1132 | 6,4367E-05 | 6,5033E-04 | 3,6447E-04 | 12 |
| hsa00360 | Phenylalanine metabolism | 5/255 | 16/8538 | 0,3125 | 10,4632 | 6,6475 | 7,6185E-05 | 7,4407E-04 | 4,1701E-04 | 5 |
| hsa04010 | MAPK signaling pathway | 22/255 | 300/8538 | 0,0733 | 2,4554 | 4,5025 | 8,7270E-05 | 8,2484E-04 | 4,6228E-04 | 22 |
| hsa00590 | Arachidonic acid metabolism | 9/255 | 63/8538 | 0,1429 | 4,7832 | 5,2880 | 9,4817E-05 | 8,6817E-04 | 4,8656E-04 | 9 |
| hsa05144 | Malaria | 8/255 | 50/8538 | 0,1600 | 5,3572 | 5,4215 | 1,0266E-04 | 9,1151E-04 | 5,1085E-04 | 8 |
| hsa05140 | Leishmaniasis | 10/255 | 79/8538 | 0,1266 | 4,2383 | 5,0734 | 1,1113E-04 | 9,5772E-04 | 5,3675E-04 | 10 |
| hsa04713 | Circadian entrainment | 11/255 | 97/8538 | 0,1134 | 3,7970 | 4,8608 | 1,3938E-04 | 1,1668E-03 | 6,5392E-04 | 11 |
| hsa04630 | JAK-STAT signaling pathway | 15/255 | 168/8538 | 0,0893 | 2,9895 | 4,5694 | 1,4417E-04 | 1,1734E-03 | 6,5762E-04 | 15 |
| hsa05215 | Prostate cancer | 11/255 | 98/8538 | 0,1122 | 3,7582 | 4,8184 | 1,5292E-04 | 1,2110E-03 | 6,7869E-04 | 11 |
| hsa05033 | Nicotine addiction | 7/255 | 41/8538 | 0,1707 | 5,7165 | 5,3114 | 1,8398E-04 | 1,4186E-03 | 7,9505E-04 | 7 |
| hsa00380 | Tryptophan metabolism | 7/255 | 42/8538 | 0,1667 | 5,5804 | 5,2209 | 2,1523E-04 | 1,5766E-03 | 8,8358E-04 | 7 |
| hsa04216 | Ferroptosis | 7/255 | 42/8538 | 0,1667 | 5,5804 | 5,2209 | 2,1523E-04 | 1,5766E-03 | 8,8358E-04 | 7 |
| hsa00071 | Fatty acid degradation | 7/255 | 43/8538 | 0,1628 | 5,4506 | 5,1334 | 2,5062E-04 | 1,7910E-03 | 1,0038E-03 | 7 |
| hsa00410 | beta-Alanine metabolism | 6/255 | 31/8538 | 0,1935 | 6,4805 | 5,3634 | 2,6281E-04 | 1,8334E-03 | 1,0275E-03 | 6 |
| hsa04976 | Bile secretion | 10/255 | 90/8538 | 0,1111 | 3,7203 | 4,5518 | 3,3096E-04 | 2,2551E-03 | 1,2639E-03 | 10 |
| hsa04936 | Alcoholic liver disease | 13/255 | 144/8538 | 0,0903 | 3,0227 | 4,2950 | 3,5813E-04 | 2,3848E-03 | 1,3365E-03 | 13 |
| hsa04510 | Focal adhesion | 16/255 | 203/8538 | 0,0788 | 2,6390 | 4,1467 | 3,6715E-04 | 2,3906E-03 | 1,3398E-03 | 16 |
| hsa04540 | Gap junction | 10/255 | 92/8538 | 0,1087 | 3,6394 | 4,4658 | 3,9593E-04 | 2,5219E-03 | 1,4134E-03 | 10 |
| hsa00620 | Pyruvate metabolism | 7/255 | 47/8538 | 0,1489 | 4,9867 | 4,8086 | 4,4184E-04 | 2,7545E-03 | 1,5437E-03 | 7 |
| hsa04015 | Rap1 signaling pathway | 16/255 | 212/8538 | 0,0755 | 2,5270 | 3,9501 | 5,9375E-04 | 3,6243E-03 | 2,0312E-03 | 16 |
| hsa00330 | Arginine and proline metabolism | 7/255 | 50/8538 | 0,1400 | 4,6875 | 4,5882 | 6,5027E-04 | 3,7676E-03 | 2,1115E-03 | 7 |
| hsa04929 | GnRH secretion | 8/255 | 65/8538 | 0,1231 | 4,1209 | 4,4315 | 6,5610E-04 | 3,7676E-03 | 2,1115E-03 | 8 |
| hsa04925 | Aldosterone synthesis and secretion | 10/255 | 98/8538 | 0,1020 | 3,4166 | 4,2215 | 6,5756E-04 | 3,7676E-03 | 2,1115E-03 | 10 |
| hsa04725 | Cholinergic synapse | 11/255 | 116/8538 | 0,0948 | 3,1751 | 4,1383 | 6,6865E-04 | 3,7676E-03 | 2,1115E-03 | 11 |
| hsa04934 | Cushing syndrome | 13/255 | 155/8538 | 0,0839 | 2,8082 | 3,9860 | 7,2604E-04 | 4,0138E-03 | 2,2495E-03 | 13 |
| hsa00010 | Glycolysis / Gluconeogenesis | 8/255 | 67/8538 | 0,1194 | 3,9979 | 4,3223 | 8,0541E-04 | 4,2907E-03 | 2,4047E-03 | 8 |
| hsa04720 | Long-term potentiation | 8/255 | 67/8538 | 0,1194 | 3,9979 | 4,3223 | 8,0541E-04 | 4,2907E-03 | 2,4047E-03 | 8 |
| hsa04933 | AGE-RAGE signaling pathway in diabetic complications | 10/255 | 101/8538 | 0,0990 | 3,3151 | 4,1064 | 8,3415E-04 | 4,3644E-03 | 2,4460E-03 | 10 |
| hsa05031 | Amphetamine addiction | 8/255 | 69/8538 | 0,1159 | 3,8820 | 4,2173 | 9,8089E-04 | 5,0421E-03 | 2,8258E-03 | 8 |
| hsa05206 | MicroRNAs in cancer | 20/255 | 312/8538 | 0,0641 | 2,1463 | 3,6192 | 1,0380E-03 | 5,2439E-03 | 2,9389E-03 | 20 |
| hsa05205 | Proteoglycans in cancer | 15/255 | 204/8538 | 0,0735 | 2,4619 | 3,7081 | 1,1440E-03 | 5,6813E-03 | 3,1840E-03 | 15 |
| hsa05224 | Breast cancer | 12/255 | 148/8538 | 0,0811 | 2,7148 | 3,6922 | 1,5477E-03 | 7,5580E-03 | 4,2359E-03 | 12 |
| hsa01524 | Platinum drug resistance | 8/255 | 75/8538 | 0,1067 | 3,5715 | 3,9244 | 1,6973E-03 | 7,9712E-03 | 4,4674E-03 | 8 |
| hsa04918 | Thyroid hormone synthesis | 8/255 | 75/8538 | 0,1067 | 3,5715 | 3,9244 | 1,6973E-03 | 7,9712E-03 | 4,4674E-03 | 8 |
| hsa00053 | Ascorbate and aldarate metabolism | 5/255 | 30/8538 | 0,1667 | 5,5804 | 4,4094 | 1,7655E-03 | 7,9712E-03 | 4,4674E-03 | 5 |
| hsa01523 | Antifolate resistance | 5/255 | 30/8538 | 0,1667 | 5,5804 | 4,4094 | 1,7655E-03 | 7,9712E-03 | 4,4674E-03 | 5 |
| hsa05213 | Endometrial cancer | 7/255 | 59/8538 | 0,1186 | 3,9725 | 4,0198 | 1,7683E-03 | 7,9712E-03 | 4,4674E-03 | 7 |
| hsa04370 | VEGF signaling pathway | 7/255 | 60/8538 | 0,1167 | 3,9063 | 3,9636 | 1,9520E-03 | 8,6655E-03 | 4,8565E-03 | 7 |
| hsa04928 | Parathyroid hormone synthesis, secretion and action | 10/255 | 115/8538 | 0,0870 | 2,9115 | 3,6209 | 2,2483E-03 | 9,7255E-03 | 5,4506E-03 | 10 |
| hsa05165 | Human papillomavirus infection | 20/255 | 333/8538 | 0,0601 | 2,0110 | 3,3017 | 2,2669E-03 | 9,7255E-03 | 5,4506E-03 | 20 |
| hsa04921 | Oxytocin signaling pathway | 12/255 | 155/8538 | 0,0774 | 2,5922 | 3,5098 | 2,2903E-03 | 9,7255E-03 | 5,4506E-03 | 12 |
| hsa04721 | Synaptic vesicle cycle | 8/255 | 79/8538 | 0,1013 | 3,3906 | 3,7454 | 2,3695E-03 | 9,9181E-03 | 5,5585E-03 | 8 |
| hsa00310 | Lysine degradation | 7/255 | 63/8538 | 0,1111 | 3,7203 | 3,8022 | 2,5924E-03 | 1,0698E-02 | 5,9957E-03 | 7 |
| hsa01522 | Endocrine resistance | 9/255 | 99/8538 | 0,0909 | 3,0439 | 3,5888 | 2,7298E-03 | 1,1109E-02 | 6,2259E-03 | 9 |
| hsa00983 | Drug metabolism - other enzymes | 8/255 | 81/8538 | 0,0988 | 3,3069 | 3,6601 | 2,7759E-03 | 1,1142E-02 | 6,2442E-03 | 8 |
| hsa04927 | Cortisol synthesis and secretion | 7/255 | 65/8538 | 0,1077 | 3,6058 | 3,7000 | 3,1016E-03 | 1,2281E-02 | 6,8826E-03 | 7 |
| hsa05321 | Inflammatory bowel disease | 7/255 | 66/8538 | 0,1061 | 3,5512 | 3,6504 | 3,3833E-03 | 1,3217E-02 | 7,4075E-03 | 7 |
| hsa05146 | Amoebiasis | 9/255 | 103/8538 | 0,0874 | 2,9256 | 3,4497 | 3,5704E-03 | 1,3765E-02 | 7,7144E-03 | 9 |
| hsa00340 | Histidine metabolism | 4/255 | 22/8538 | 0,1818 | 6,0877 | 4,1922 | 3,7184E-03 | 1,4149E-02 | 7,9299E-03 | 4 |
| hsa04742 | Taste transduction | 8/255 | 86/8538 | 0,0930 | 3,1146 | 3,4581 | 4,0311E-03 | 1,5142E-02 | 8,4865E-03 | 8 |
| hsa05143 | African trypanosomiasis | 5/255 | 37/8538 | 0,1351 | 4,5246 | 3,7697 | 4,5572E-03 | 1,6902E-02 | 9,4726E-03 | 5 |
| hsa04723 | Retrograde endocannabinoid signaling | 11/255 | 149/8538 | 0,0738 | 2,4719 | 3,1800 | 4,9524E-03 | 1,8138E-02 | 1,0165E-02 | 11 |
| hsa05235 | PD-L1 expression and PD-1 checkpoint pathway in cancer | 8/255 | 90/8538 | 0,0889 | 2,9762 | 3,3068 | 5,3188E-03 | 1,9240E-02 | 1,0783E-02 | 8 |
| hsa05032 | Morphine addiction | 8/255 | 91/8538 | 0,0879 | 2,9435 | 3,2703 | 5,6850E-03 | 2,0314E-02 | 1,1385E-02 | 8 |
| hsa05218 | Melanoma | 7/255 | 73/8538 | 0,0959 | 3,2106 | 3,3281 | 5,9357E-03 | 2,0954E-02 | 1,1743E-02 | 7 |
| hsa05145 | Toxoplasmosis | 9/255 | 112/8538 | 0,0804 | 2,6905 | 3,1598 | 6,2028E-03 | 2,1636E-02 | 1,2126E-02 | 9 |
| hsa04068 | FoxO signaling pathway | 10/255 | 133/8538 | 0,0752 | 2,5175 | 3,0946 | 6,3833E-03 | 2,2004E-02 | 1,2332E-02 | 10 |
| hsa04658 | Th1 and Th2 cell differentiation | 8/255 | 93/8538 | 0,0860 | 2,8802 | 3,1987 | 6,4748E-03 | 2,2060E-02 | 1,2363E-02 | 8 |

| | | | | | | | | | | |
|---|---|---|---|---|---|---|---|---|---|---|
| hsa04115 | p53 signaling pathway | 7/255 | 75/8538 | 0,0933 | 3,1250 | 3,2431 | 6,8767E-03 | 2,3159E-02 | 1,2980E-02 | 7 |
| hsa04971 | Gastric acid secretion | 7/255 | 76/8538 | 0,0921 | 3,0839 | 3,2017 | 7,3867E-03 | 2,4595E-02 | 1,3784E-02 | 7 |
| hsa05133 | Pertussis | 7/255 | 78/8538 | 0,0897 | 3,0048 | 3,1208 | 8,4902E-03 | 2,7951E-02 | 1,5665E-02 | 7 |
| hsa00790 | Folate biosynthesis | 4/255 | 28/8538 | 0,1429 | 4,7832 | 3,5180 | 9,0482E-03 | 2,9457E-02 | 1,6509E-02 | 4 |
| hsa05034 | Alcoholism | 12/255 | 188/8538 | 0,0638 | 2,1372 | 2,7662 | 1,0587E-02 | 3,3941E-02 | 1,9022E-02 | 12 |
| hsa04935 | Growth hormone synthesis, secretion and action | 9/255 | 122/8538 | 0,0738 | 2,4700 | 2,8693 | 1,0657E-02 | 3,3941E-02 | 1,9022E-02 | 9 |
| hsa00591 | Linoleic acid metabolism | 4/255 | 30/8538 | 0,1333 | 4,4643 | 3,3350 | 1,1561E-02 | 3,6422E-02 | 2,0413E-02 | 4 |
| hsa05010 | Alzheimer disease | 20/255 | 391/8538 | 0,0512 | 1,7127 | 2,5310 | 1,3056E-02 | 4,0493E-02 | 2,2694E-02 | 20 |
| hsa00430 | Taurine and hypotaurine metabolism | 3/255 | 17/8538 | 0,1765 | 5,9087 | 3,5544 | 1,3129E-02 | 4,0493E-02 | 2,2694E-02 | 3 |
| hsa00280 | Valine, leucine and isoleucine degradation | 5/255 | 48/8538 | 0,1042 | 3,4877 | 3,0325 | 1,3736E-02 | 4,1924E-02 | 2,3496E-02 | 5 |
| hsa05226 | Gastric cancer | 10/255 | 150/8538 | 0,0667 | 2,2322 | 2,6712 | 1,4272E-02 | 4,3111E-02 | 2,4161E-02 | 10 |
| hsa05210 | Colorectal cancer | 7/255 | 87/8538 | 0,0805 | 2,6940 | 2,7864 | 1,4996E-02 | 4,4836E-02 | 2,5128E-02 | 7 |
| hsa04659 | Th17 cell differentiation | 8/255 | 109/8538 | 0,0734 | 2,4574 | 2,6868 | 1,6107E-02 | 4,7671E-02 | 2,6717E-02 | 8 |
| hsa04727 | GABAergic synapse | 7/255 | 89/8538 | 0,0787 | 2,6334 | 2,7178 | 1,6824E-02 | 4,9293E-02 | 2,7626E-02 | 7 |
| hsa05211 | Renal cell carcinoma | 6/255 | 70/8538 | 0,0857 | 2,8699 | 2,7562 | 1,7814E-02 | 5,1252E-02 | 2,8724E-02 | 6 |
| hsa05163 | Human cytomegalovirus infection | 13/255 | 226/8538 | 0,0575 | 1,9260 | 2,4753 | 1,7842E-02 | 5,1252E-02 | 2,8724E-02 | 13 |
| hsa04218 | Cellular senescence | 10/255 | 157/8538 | 0,0637 | 2,1326 | 2,5132 | 1,9078E-02 | 5,4270E-02 | 3,0416E-02 | 10 |
| hsa04961 | Endocrine and other factor-regulated calcium reabsorption | 5/255 | 53/8538 | 0,0943 | 3,1587 | 2,7659 | 2,0425E-02 | 5,7341E-02 | 3,2137E-02 | 5 |
| hsa00100 | Steroid biosynthesis | 3/255 | 20/8538 | 0,1500 | 5,0224 | 3,1598 | 2,0611E-02 | 5,7341E-02 | 3,2137E-02 | 3 |
| hsa04520 | Adherens junction | 7/255 | 93/8538 | 0,0753 | 2,5202 | 2,5862 | 2,0940E-02 | 5,7341E-02 | 3,2137E-02 | 7 |
| hsa04912 | GnRH signaling pathway | 7/255 | 93/8538 | 0,0753 | 2,5202 | 2,5862 | 2,0940E-02 | 5,7341E-02 | 3,2137E-02 | 7 |
| hsa05223 | Non-small cell lung cancer | 6/255 | 73/8538 | 0,0822 | 2,7520 | 2,6376 | 2,1493E-02 | 5,8139E-02 | 3,2584E-02 | 6 |
| hsa00040 | Pentose and glucuronate interconversions | 4/255 | 36/8538 | 0,1111 | 3,7203 | 2,8697 | 2,1629E-02 | 5,8139E-02 | 3,2584E-02 | 4 |
| hsa04915 | Estrogen signaling pathway | 9/255 | 139/8538 | 0,0647 | 2,1679 | 2,4358 | 2,3213E-02 | 6,1349E-02 | 3,4383E-02 | 9 |
| hsa04657 | IL-17 signaling pathway | 7/255 | 95/8538 | 0,0737 | 2,4671 | 2,5229 | 2,3241E-02 | 6,1349E-02 | 3,4383E-02 | 7 |
| hsa05216 | Thyroid cancer | 4/255 | 37/8538 | 0,1081 | 3,6197 | 2,8019 | 2,3698E-02 | 6,1995E-02 | 3,4745E-02 | 4 |
| hsa04371 | Apelin signaling pathway | 9/255 | 140/8538 | 0,0643 | 2,1524 | 2,4122 | 2,4186E-02 | 6,2711E-02 | 3,5146E-02 | 9 |
| hsa05214 | Glioma | 6/255 | 76/8538 | 0,0789 | 2,6433 | 2,5248 | 2,5654E-02 | 6,5508E-02 | 3,6714E-02 | 6 |
| hsa04970 | Salivary secretion | 7/255 | 97/8538 | 0,0722 | 2,4163 | 2,4613 | 2,5711E-02 | 6,5508E-02 | 3,6714E-02 | 7 |
| hsa04722 | Neurotrophin signaling pathway | 8/255 | 120/8538 | 0,0667 | 2,2322 | 2,3850 | 2,7009E-02 | 6,8220E-02 | 3,8234E-02 | 8 |
| hsa05330 | Allograft rejection | 4/255 | 39/8538 | 0,1026 | 3,4341 | 2,6731 | 2,8186E-02 | 7,0407E-02 | 3,9460E-02 | 4 |
| hsa05231 | Choline metabolism in cancer | 7/255 | 99/8538 | 0,0707 | 2,3674 | 2,4011 | 2,8355E-02 | 7,0407E-02 | 3,9460E-02 | 7 |
| hsa04660 | T cell receptor signaling pathway | 8/255 | 122/8538 | 0,0656 | 2,1956 | 2,3336 | 2,9435E-02 | 7,2475E-02 | 4,0618E-02 | 8 |
| hsa00220 | Arginine biosynthesis | 3/255 | 23/8538 | 0,1304 | 4,3673 | 2,8371 | 2,9994E-02 | 7,2630E-02 | 4,0705E-02 | 3 |
| hsa04964 | Proximal tubule bicarbonate reclamation | 3/255 | 23/8538 | 0,1304 | 4,3673 | 2,8371 | 2,9994E-02 | 7,2630E-02 | 4,0705E-02 | 3 |
| hsa04916 | Melanogenesis | 7/255 | 101/8538 | 0,0693 | 2,3206 | 2,3424 | 3,1178E-02 | 7,4878E-02 | 4,1965E-02 | 7 |
| hsa05219 | Bladder cancer | 4/255 | 41/8538 | 0,0976 | 3,2666 | 2,5525 | 3,3146E-02 | 7,8958E-02 | 4,4252E-02 | 4 |
| hsa04072 | Phospholipase D signaling pathway | 9/255 | 149/8538 | 0,0604 | 2,0224 | 2,2090 | 3,4278E-02 | 8,0995E-02 | 4,5393E-02 | 9 |
| hsa04611 | Platelet activation | 8/255 | 126/8538 | 0,0635 | 2,1259 | 2,2338 | 3,4731E-02 | 8,1402E-02 | 4,5621E-02 | 8 |
| hsa04978 | Mineral absorption | 5/255 | 61/8538 | 0,0820 | 2,7445 | 2,3990 | 3,5006E-02 | 8,1402E-02 | 4,5621E-02 | 5 |
| hsa04213 | Longevity regulating pathway - multiple species | 5/255 | 62/8538 | 0,0806 | 2,7002 | 2,3574 | 3,7185E-02 | 8,5118E-02 | 4,7704E-02 | 5 |
| hsa04330 | Notch signaling pathway | 5/255 | 62/8538 | 0,0806 | 2,7002 | 2,3574 | 3,7185E-02 | 8,5118E-02 | 4,7704E-02 | 5 |
| hsa04972 | Pancreatic secretion | 7/255 | 106/8538 | 0,0660 | 2,2111 | 2,2014 | 3,9043E-02 | 8,8680E-02 | 4,9700E-02 | 7 |
| hsa04926 | Relaxin signaling pathway | 8/255 | 130/8538 | 0,0615 | 2,0605 | 2,1377 | 4,0639E-02 | 9,1594E-02 | 5,1333E-02 | 8 |
| hsa04940 | Type I diabetes mellitus | 4/255 | 44/8538 | 0,0909 | 3,0439 | 2,3848 | 4,1484E-02 | 9,2786E-02 | 5,2001E-02 | 4 |
| hsa05412 | Arrhythmogenic right ventricular cardiomyopathy | 6/255 | 86/8538 | 0,0698 | 2,3360 | 2,1847 | 4,3276E-02 | 9,6059E-02 | 5,3836E-02 | 6 |
| hsa04931 | Insulin resistance | 7/255 | 109/8538 | 0,0642 | 2,1502 | 2,1205 | 4,4336E-02 | 9,7311E-02 | 5,4538E-02 | 7 |
| hsa05332 | Graft-versus-host disease | 4/255 | 45/8538 | 0,0889 | 2,9762 | 2,3320 | 4,4504E-02 | 9,7311E-02 | 5,4538E-02 | 4 |
| hsa04150 | mTOR signaling pathway | 9/255 | 158/8538 | 0,0570 | 1,9072 | 2,0195 | 4,6961E-02 | 1,0165E-01 | 5,6969E-02 | 9 |
| hsa04270 | Vascular smooth muscle contraction | 8/255 | 134/8538 | 0,0597 | 1,9989 | 2,0449 | 4,7182E-02 | 1,0165E-01 | 5,6969E-02 | 8 |
| hsa00860 | Porphyrin metabolism | 4/255 | 46/8538 | 0,0870 | 2,9115 | 2,2808 | 4,7644E-02 | 1,0190E-01 | 5,7107E-02 | 4 |
| hsa04930 | Type II diabetes mellitus | 4/255 | 47/8538 | 0,0851 | 2,8496 | 2,2308 | 5,0903E-02 | 1,0808E-01 | 6,0572E-02 | 4 |
| hsa04211 | Longevity regulating pathway | 6/255 | 90/8538 | 0,0667 | 2,2322 | 2,0618 | 5,2029E-02 | 1,0967E-01 | 6,1466E-02 | 6 |
| hsa04924 | Renin secretion | 5/255 | 69/8538 | 0,0725 | 2,4263 | 2,0871 | 5,4744E-02 | 1,1457E-01 | 6,4211E-02 | 5 |
| hsa05161 | Hepatitis B | 9/255 | 163/8538 | 0,0552 | 1,8487 | 1,9195 | 5,5205E-02 | 1,1472E-01 | 6,4293E-02 | 9 |
| hsa05417 | Lipid and atherosclerosis | 11/255 | 216/8538 | 0,0509 | 1,7051 | 1,8416 | 5,8817E-02 | 1,2136E-01 | 6,8016E-02 | 11 |
| hsa05222 | Small cell lung cancer | 6/255 | 93/8538 | 0,0645 | 2,1602 | 1,9737 | 5,9255E-02 | 1,2141E-01 | 6,8044E-02 | 6 |
| hsa04917 | Prolactin signaling pathway | 5/255 | 71/8538 | 0,0704 | 2,3579 | 2,0159 | 6,0509E-02 | 1,2241E-01 | 6,8604E-02 | 5 |
| hsa04022 | cGMP-PKG signaling pathway | 9/255 | 166/8538 | 0,0542 | 1,8153 | 1,8612 | 6,0578E-02 | 1,2241E-01 | 6,8604E-02 | 9 |
| hsa04672 | Intestinal immune network for IgA production | 4/255 | 50/8538 | 0,0800 | 2,6786 | 2,0886 | 6,1395E-02 | 1,2321E-01 | 6,9053E-02 | 4 |
| hsa05323 | Rheumatoid arthritis | 6/255 | 95/8538 | 0,0632 | 2,1147 | 1,9169 | 6,4390E-02 | 1,2834E-01 | 7,1928E-02 | 6 |
| hsa05167 | Kaposi sarcoma-associated herpesvirus infection | 10/255 | 196/8538 | 0,0510 | 1,7083 | 1,7601 | 6,8886E-02 | 1,3513E-01 | 7,5734E-02 | 10 |
| hsa00270 | Cysteine and methionine metabolism | 4/255 | 52/8538 | 0,0769 | 2,5756 | 1,9995 | 6,8976E-02 | 1,3513E-01 | 7,5734E-02 | 4 |
| hsa05310 | Asthma | 3/255 | 32/8538 | 0,0938 | 3,1390 | 2,1269 | 6,9180E-02 | 1,3513E-01 | 7,5734E-02 | 3 |
| hsa04071 | Sphingolipid signaling pathway | 7/255 | 122/8538 | 0,0574 | 1,9211 | 1,7979 | 7,2464E-02 | 1,4061E-01 | 7,8803E-02 | 7 |
| hsa04750 | Inflammatory mediator regulation of TRP channels | 6/255 | 99/8538 | 0,0606 | 2,0292 | 1,8072 | 7,5419E-02 | 1,4443E-01 | 8,0945E-02 | 6 |
| hsa05410 | Hypertrophic cardiomyopathy | 6/255 | 99/8538 | 0,0606 | 2,0292 | 1,8072 | 7,5419E-02 | 1,4443E-01 | 8,0945E-02 | 6 |
| hsa05134 | Legionellosis | 4/255 | 56/8538 | 0,0714 | 2,3916 | 1,8331 | 8,5505E-02 | 1,6268E-01 | 9,1174E-02 | 4 |
| hsa04261 | Adrenergic signaling in cardiomyocytes | 8/255 | 154/8538 | 0,0519 | 1,7393 | 1,6245 | 8,9932E-02 | 1,6807E-01 | 9,4194E-02 | 8 |
| hsa00120 | Primary bile acid biosynthesis | 2/255 | 17/8538 | 0,1176 | 3,9391 | 2,1282 | 9,0057E-02 | 1,6807E-01 | 9,4194E-02 | 2 |
| hsa03273 | Virion - Lassa virus and SFTS virus | 2/255 | 17/8538 | 0,1176 | 3,9391 | 2,1282 | 9,0057E-02 | 1,6807E-01 | 9,4194E-02 | 2 |
| hsa04064 | NF-kappa B signaling pathway | 6/255 | 105/8538 | 0,0571 | 1,9133 | 1,6521 | 9,3850E-02 | 1,7404E-01 | 9,7539E-02 | 6 |
| hsa04148 | Efferocytosis | 8/255 | 157/8538 | 0,0510 | 1,7061 | 1,5668 | 9,7814E-02 | 1,8025E-01 | 1,0102E-01 | 8 |
| hsa04923 | Regulation of lipolysis in adipocytes | 4/255 | 59/8538 | 0,0678 | 2,2700 | 1,7174 | 9,9051E-02 | 1,8139E-01 | 1,0166E-01 | 4 |
| hsa04350 | TGF-beta signaling pathway | 6/255 | 108/8538 | 0,0556 | 1,8601 | 1,5783 | 1,0390E-01 | 1,8908E-01 | 1,0597E-01 | 6 |
| hsa04911 | Insulin secretion | 5/255 | 86/8538 | 0,0581 | 1,9466 | 1,5481 | 1,1410E-01 | 2,0636E-01 | 1,1565E-01 | 5 |
| hsa05135 | Yersinia infection | 7/255 | 138/8538 | 0,0507 | 1,6984 | 1,4512 | 1,1885E-01 | 2,1363E-01 | 1,1973E-01 | 7 |
| hsa00260 | Glycine, serine and threonine metabolism | 3/255 | 41/8538 | 0,0732 | 2,4499 | 1,6328 | 1,2282E-01 | 2,1942E-01 | 1,2297E-01 | 3 |
| hsa00770 | Pantothenate and CoA biosynthesis | 2/255 | 21/8538 | 0,0952 | 3,1888 | 1,7620 | 1,2882E-01 | 2,2747E-01 | 1,2749E-01 | 2 |
| hsa00561 | Glycerolipid metabolism | 4/255 | 65/8538 | 0,0615 | 2,0605 | 1,5058 | 1,2888E-01 | 2,2747E-01 | 1,2749E-01 | 4 |
| hsa04380 | Osteoclast differentiation | 7/255 | 143/8538 | 0,0490 | 1,6390 | 1,3520 | 1,3590E-01 | 2,3844E-01 | 1,3363E-01 | 7 |
| hsa05202 | Transcriptional misregulation in cancer | 9/255 | 198/8538 | 0,0455 | 1,5219 | 1,3037 | 1,3828E-01 | 2,4035E-01 | 1,3470E-01 | 9 |
| hsa04550 | Signaling pathways regulating pluripotency of stem cells | 7/255 | 144/8538 | 0,0486 | 1,6276 | 1,3327 | 1,3945E-01 | 2,4035E-01 | 1,3470E-01 | 7 |
| hsa05017 | Spinocerebellar ataxia | 7/255 | 144/8538 | 0,0486 | 1,6276 | 1,3327 | 1,3945E-01 | 2,4035E-01 | 1,3470E-01 | 7 |
| hsa05014 | Amyotrophic lateral sclerosis | 15/255 | 371/8538 | 0,0404 | 1,3537 | 1,2223 | 1,4367E-01 | 2,4618E-01 | 1,3797E-01 | 15 |
| hsa05221 | Acute myeloid leukemia | 4/255 | 68/8538 | 0,0588 | 1,9696 | 1,4084 | 1,4504E-01 | 2,4707E-01 | 1,3847E-01 | 4 |
| hsa00515 | Mannose type O-glycan biosynthesis | 2/255 | 23/8538 | 0,0870 | 2,9115 | 1,6106 | 1,4942E-01 | 2,5214E-01 | 1,4131E-01 | 2 |
| hsa02010 | ABC transporters | 3/255 | 45/8538 | 0,0667 | 2,2322 | 1,4540 | 1,5032E-01 | 2,5214E-01 | 1,4131E-01 | 3 |

| | | | | | | | | | | |
|---|---|---|---|---|---|---|---|---|---|---|
| hsa04664 | Fc epsilon RI signaling pathway | 4/255 | 69/8538 | 0,0580 | 1,9410 | 1,3770 | 1,5059E-01 | 2,5214E-01 | 1,4131E-01 | 4 |
| hsa04920 | Adipocytokine signaling pathway | 4/255 | 70/8538 | 0,0571 | 1,9133 | 1,3461 | 1,5623E-01 | 2,6009E-01 | 1,4577E-01 | 4 |
| hsa05120 | Epithelial cell signaling in Helicobacter pylori infection | 4/255 | 71/8538 | 0,0563 | 1,8863 | 1,3158 | 1,6195E-01 | 2,6657E-01 | 1,4940E-01 | 4 |
| hsa05230 | Central carbon metabolism in cancer | 4/255 | 71/8538 | 0,0563 | 1,8863 | 1,3158 | 1,6195E-01 | 2,6657E-01 | 1,4940E-01 | 4 |
| hsa04070 | Phosphatidylinositol signaling system | 5/255 | 98/8538 | 0,0510 | 1,7083 | 1,2373 | 1,6886E-01 | 2,7641E-01 | 1,5491E-01 | 5 |
| hsa05171 | Coronavirus disease - COVID-19 | 10/255 | 238/8538 | 0,0420 | 1,4068 | 1,1168 | 1,7476E-01 | 2,8447E-01 | 1,5943E-01 | 10 |
| hsa05152 | Tuberculosis | 8/255 | 182/8538 | 0,0440 | 1,4718 | 1,1287 | 1,7764E-01 | 2,8757E-01 | 1,6117E-01 | 8 |
| hsa04060 | Cytokine-cytokine receptor interaction | 12/255 | 298/8538 | 0,0403 | 1,3483 | 1,0738 | 1,8075E-01 | 2,9098E-01 | 1,6308E-01 | 12 |
| hsa05170 | Human immunodeficiency virus 1 infection | 9/255 | 213/8538 | 0,0423 | 1,4147 | 1,0755 | 1,8692E-01 | 2,9928E-01 | 1,6773E-01 | 9 |
| hsa04979 | Cholesterol metabolism | 3/255 | 51/8538 | 0,0588 | 1,9696 | 1,2184 | 1,9474E-01 | 3,0985E-01 | 1,7365E-01 | 3 |
| hsa05212 | Pancreatic cancer | 4/255 | 77/8538 | 0,0519 | 1,7393 | 1,1434 | 1,9772E-01 | 3,0985E-01 | 1,7365E-01 | 4 |
| hsa05220 | Chronic myeloid leukemia | 4/255 | 77/8538 | 0,0519 | 1,7393 | 1,1434 | 1,9772E-01 | 3,0985E-01 | 1,7365E-01 | 4 |
| hsa04217 | Necroptosis | 7/255 | 159/8538 | 0,0440 | 1,4741 | 1,0587 | 1,9775E-01 | 3,0985E-01 | 1,7365E-01 | 7 |
| hsa04973 | Carbohydrate digestion and absorption | 3/255 | 52/8538 | 0,0577 | 1,9317 | 1,1823 | 2,0244E-01 | 3,1468E-01 | 1,7636E-01 | 3 |
| hsa05100 | Bacterial invasion of epithelial cells | 4/255 | 78/8538 | 0,0513 | 1,7170 | 1,1162 | 2,0391E-01 | 3,1468E-01 | 1,7636E-01 | 4 |
| hsa04621 | NOD-like receptor signaling pathway | 8/255 | 189/8538 | 0,0423 | 1,4172 | 1,0177 | 2,0406E-01 | 3,1468E-01 | 1,7636E-01 | 8 |
| hsa05132 | Salmonella infection | 10/255 | 251/8538 | 0,0398 | 1,3340 | 0,9422 | 2,1790E-01 | 3,3275E-01 | 1,8649E-01 | 10 |
| hsa05320 | Autoimmune thyroid disease | 3/255 | 54/8538 | 0,0556 | 1,8601 | 1,1125 | 2,1805E-01 | 3,3275E-01 | 1,8649E-01 | 3 |
| hsa04210 | Apoptosis | 6/255 | 136/8538 | 0,0441 | 1,4772 | 0,9842 | 2,2107E-01 | 3,3562E-01 | 1,8809E-01 | 6 |
| hsa04620 | Toll-like receptor signaling pathway | 5/255 | 109/8538 | 0,0459 | 1,5359 | 0,9879 | 2,2635E-01 | 3,4186E-01 | 1,9159E-01 | 5 |
| hsa04146 | Peroxisome | 4/255 | 83/8538 | 0,0482 | 1,6136 | 0,9856 | 2,3563E-01 | 3,5171E-01 | 1,9712E-01 | 4 |
| hsa05162 | Measles | 6/255 | 139/8538 | 0,0432 | 1,4453 | 0,9287 | 2,3567E-01 | 3,5171E-01 | 1,9712E-01 | 6 |
| hsa04981 | Folate transport and metabolism | 2/255 | 31/8538 | 0,0645 | 2,1602 | 1,1354 | 2,3648E-01 | 3,5171E-01 | 1,9712E-01 | 2 |
| hsa04140 | Autophagy - animal | 7/255 | 169/8538 | 0,0414 | 1,3868 | 0,8912 | 2,4115E-01 | 3,5685E-01 | 2,0000E-01 | 7 |
| hsa04012 | ErbB signaling pathway | 4/255 | 86/8538 | 0,0465 | 1,5573 | 0,9114 | 2,5520E-01 | 3,7575E-01 | 2,1059E-01 | 4 |
| hsa05164 | Influenza A | 7/255 | 173/8538 | 0,0405 | 1,3548 | 0,8271 | 2,5932E-01 | 3,7974E-01 | 2,1283E-01 | 7 |
| hsa05130 | Pathogenic Escherichia coli infection | 8/255 | 203/8538 | 0,0394 | 1,3195 | 0,8083 | 2,6107E-01 | 3,7974E-01 | 2,1283E-01 | 8 |
| hsa04260 | Cardiac muscle contraction | 4/255 | 87/8538 | 0,0460 | 1,5394 | 0,8873 | 2,6180E-01 | 3,7974E-01 | 2,1283E-01 | 4 |
| hsa04610 | Complement and coagulation cascades | 4/255 | 88/8538 | 0,0455 | 1,5219 | 0,8635 | 2,6843E-01 | 3,8745E-01 | 2,1714E-01 | 4 |
| hsa04710 | Circadian rhythm | 2/255 | 34/8538 | 0,0588 | 1,9696 | 0,9939 | 2,6998E-01 | 3,8776E-01 | 2,1732E-01 | 2 |
| hsa04668 | TNF signaling pathway | 5/255 | 119/8538 | 0,0420 | 1,4068 | 0,7841 | 2,8285E-01 | 4,0427E-01 | 2,2657E-01 | 5 |
| hsa04662 | B cell receptor signaling pathway | 4/255 | 91/8538 | 0,0440 | 1,4718 | 0,7938 | 2,8850E-01 | 4,1034E-01 | 2,2997E-01 | 4 |
| hsa05217 | Basal cell carcinoma | 3/255 | 63/8538 | 0,0476 | 1,5944 | 0,8308 | 2,9065E-01 | 4,1047E-01 | 2,3005E-01 | 3 |
| hsa05012 | Parkinson disease | 10/255 | 271/8538 | 0,0369 | 1,2355 | 0,6913 | 2,9139E-01 | 4,1047E-01 | 2,3005E-01 | 10 |
| hsa04152 | AMPK signaling pathway | 5/255 | 122/8538 | 0,0410 | 1,3722 | 0,7265 | 3,0033E-01 | 4,1919E-01 | 2,3493E-01 | 5 |
| hsa05168 | Herpes simplex virus 1 infection | 7/255 | 182/8538 | 0,0385 | 1,2878 | 0,6885 | 3,0151E-01 | 4,1919E-01 | 2,3493E-01 | 7 |
| hsa01240 | Biosynthesis of cofactors | 6/255 | 152/8538 | 0,0395 | 1,3217 | 0,7021 | 3,0187E-01 | 4,1919E-01 | 2,3493E-01 | 6 |
| hsa00250 | Alanine, aspartate and glutamate metabolism | 2/255 | 37/8538 | 0,0541 | 1,8099 | 0,8662 | 3,0345E-01 | 4,1938E-01 | 2,3504E-01 | 2 |
| hsa00130 | Ubiquinone and other terpenoid-quinone biosynthesis | 1/255 | Dec-38 | 0,0833 | 2,7902 | 1,0888 | 3,0518E-01 | 4,1980E-01 | 2,3527E-01 | 1 |
| hsa05020 | Prion disease | 10/255 | 278/8538 | 0,0360 | 1,2044 | 0,6079 | 3,1864E-01 | 4,3626E-01 | 2,4450E-01 | 10 |
| hsa05016 | Huntington disease | 11/255 | 311/8538 | 0,0354 | 1,1843 | 0,5808 | 3,2565E-01 | 4,4380E-01 | 2,4872E-01 | 11 |
| hsa04390 | Hippo signaling pathway | 6/255 | 157/8538 | 0,0382 | 1,2796 | 0,6204 | 3,2825E-01 | 4,4527E-01 | 2,4955E-01 | 6 |
| hsa05131 | Shigellosis | 9/255 | 250/8538 | 0,0360 | 1,2054 | 0,5782 | 3,3115E-01 | 4,4712E-01 | 2,5059E-01 | 9 |
| hsa05160 | Hepatitis C | 6/255 | 159/8538 | 0,0377 | 1,2635 | 0,5884 | 3,3889E-01 | 4,5549E-01 | 2,5528E-01 | 6 |
| hsa00533 | Glycosaminoglycan biosynthesis - keratan sulfate | 1/255 | 14/8538 | 0,0714 | 2,3916 | 0,9143 | 3,4612E-01 | 4,6307E-01 | 2,5953E-01 | 1 |
| hsa04640 | Hematopoietic cell lineage | 4/255 | 100/8538 | 0,0400 | 1,3393 | 0,5988 | 3,4965E-01 | 4,6567E-01 | 2,6099E-01 | 4 |
| hsa04650 | Natural killer cell mediated cytotoxicity | 5/255 | 134/8538 | 0,0373 | 1,2493 | 0,5104 | 3,7152E-01 | 4,9214E-01 | 2,7582E-01 | 5 |
| hsa00562 | Inositol phosphate metabolism | 3/255 | 73/8538 | 0,0411 | 1,3760 | 0,5660 | 3,7288E-01 | 4,9214E-01 | 2,7582E-01 | 3 |
| hsa04974 | Protein digestion and absorption | 4/255 | 105/8538 | 0,0381 | 1,2755 | 0,4984 | 3,8380E-01 | 5,0202E-01 | 2,8136E-01 | 4 |
| hsa05414 | Dilated cardiomyopathy | 4/255 | 105/8538 | 0,0381 | 1,2755 | 0,4984 | 3,8380E-01 | 5,0202E-01 | 2,8136E-01 | 4 |
| hsa04810 | Regulation of actin cytoskeleton | 8/255 | 232/8538 | 0,0345 | 1,1546 | 0,4188 | 3,9032E-01 | 5,0828E-01 | 2,8486E-01 | 8 |
| hsa04910 | Insulin signaling pathway | 5/255 | 138/8538 | 0,0362 | 1,2131 | 0,4429 | 3,9539E-01 | 5,1261E-01 | 2,8729E-01 | 5 |
| hsa04114 | Oocyte meiosis | 5/255 | 139/8538 | 0,0360 | 1,2044 | 0,4263 | 4,0135E-01 | 5,1572E-01 | 2,8903E-01 | 5 |
| hsa00450 | Selenocompound metabolism | 1/255 | 17/8538 | 0,0588 | 1,9696 | 0,7021 | 4,0307E-01 | 5,1572E-01 | 2,8903E-01 | 1 |
| hsa00910 | Nitrogen metabolism | 1/255 | 17/8538 | 0,0588 | 1,9696 | 0,7021 | 4,0307E-01 | 5,1572E-01 | 2,8903E-01 | 1 |
| hsa05322 | Systemic lupus erythematosus | 5/255 | 141/8538 | 0,0355 | 1,1873 | 0,3935 | 4,1325E-01 | 5,2645E-01 | 2,9505E-01 | 5 |
| hsa00565 | Ether lipid metabolism | 2/255 | 50/8538 | 0,0400 | 1,3393 | 0,4222 | 4,4291E-01 | 5,6178E-01 | 3,1485E-01 | 2 |
| hsa05110 | Vibrio cholerae infection | 2/255 | 51/8538 | 0,0392 | 1,3130 | 0,3934 | 4,5304E-01 | 5,6991E-01 | 3,1941E-01 | 2 |
| hsa04623 | Cytosolic DNA-sensing pathway | 3/255 | 83/8538 | 0,0361 | 1,2102 | 0,3376 | 4,5321E-01 | 5,6991E-01 | 3,1941E-01 | 3 |
| hsa04670 | Leukocyte transendothelial migration | 4/255 | 116/8538 | 0,0345 | 1,1546 | 0,2941 | 4,5787E-01 | 5,7331E-01 | 3,2131E-01 | 4 |
| hsa00532 | Glycosaminoglycan biosynthesis - chondroitin sulfate / dermatan sulfate | 1/255 | 21/8538 | 0,0476 | 1,5944 | 0,4785 | 4,7139E-01 | 5,8774E-01 | 3,2939E-01 | 1 |
| hsa04512 | ECM-receptor interaction | 3/255 | 89/8538 | 0,0337 | 1,1286 | 0,2140 | 4,9936E-01 | 6,1997E-01 | 3,4746E-01 | 3 |
| hsa04614 | Renin-angiotensin system | 1/255 | 23/8538 | 0,0435 | 1,4558 | 0,3840 | 5,0257E-01 | 6,2133E-01 | 3,4822E-01 | 1 |
| hsa05166 | Human T-cell leukemia virus 1 infection | 7/255 | 224/8538 | 0,0313 | 1,0463 | 0,1233 | 5,0617E-01 | 6,2314E-01 | 3,4924E-01 | 7 |
| hsa00534 | Glycosaminoglycan biosynthesis - heparan sulfate / heparin | 1/255 | 24/8538 | 0,0417 | 1,3951 | 0,3401 | 5,1747E-01 | 6,3439E-01 | 3,5554E-01 | 1 |
| hsa00230 | Purine metabolism | 4/255 | 128/8538 | 0,0313 | 1,0463 | 0,0926 | 5,3497E-01 | 6,5311E-01 | 3,6603E-01 | 4 |
| hsa00592 | alpha-Linolenic acid metabolism | 1/255 | 26/8538 | 0,0385 | 1,2878 | 0,2578 | 5,4594E-01 | 6,6100E-01 | 3,7045E-01 | 1 |
| hsa04950 | Maturity onset diabetes of the young | 1/255 | 26/8538 | 0,0385 | 1,2878 | 0,2578 | 5,4594E-01 | 6,6100E-01 | 3,7045E-01 | 1 |
| hsa00650 | Butanoate metabolism | 1/255 | 27/8538 | 0,0370 | 1,2401 | 0,2192 | 5,5955E-01 | 6,7468E-01 | 3,7812E-01 | 1 |
| hsa04666 | Fc gamma R-mediated phagocytosis | 3/255 | 99/8538 | 0,0303 | 1,0146 | 0,0257 | 5,7160E-01 | 6,8360E-01 | 3,8312E-01 | 3 |
| hsa04966 | Collecting duct acid secretion | 1/255 | 28/8538 | 0,0357 | 1,1958 | 0,1821 | 5,7274E-01 | 6,8360E-01 | 3,8312E-01 | 1 |
| hsa04530 | Tight junction | 5/255 | 170/8538 | 0,0294 | 0,9848 | -0,0352 | 5,7764E-01 | 6,8360E-01 | 3,8312E-01 | 5 |
| hsa04061 | Viral protein interaction with cytokine and cytokine receptor | 3/255 | 100/8538 | 0,0300 | 1,0045 | 0,0079 | 5,7847E-01 | 6,8360E-01 | 3,8312E-01 | 3 |
| hsa05415 | Diabetic cardiomyopathy | 6/255 | 205/8538 | 0,0293 | 0,9800 | -0,0509 | 5,7861E-01 | 6,8360E-01 | 3,8312E-01 | 6 |
| hsa04310 | Wnt signaling pathway | 5/255 | 174/8538 | 0,0287 | 0,9621 | -0,0885 | 5,9844E-01 | 7,0161E-01 | 3,9322E-01 | 5 |
| hsa00564 | Glycerophospholipid metabolism | 3/255 | 103/8538 | 0,0291 | 0,9752 | -0,0444 | 5,9865E-01 | 7,0161E-01 | 3,9322E-01 | 3 |
| hsa00630 | Glyoxylate and dicarboxylate metabolism | 1/255 | 31/8538 | 0,0323 | 1,0801 | 0,0784 | 6,1002E-01 | 7,1127E-01 | 3,9863E-01 | 1 |
| hsa04137 | Mitophagy - animal | 3/255 | 105/8538 | 0,0286 | 0,9566 | -0,0784 | 6,1174E-01 | 7,1127E-01 | 3,9863E-01 | 3 |
| hsa00052 | Galactose metabolism | 1/255 | 32/8538 | 0,0313 | 1,0463 | 0,0461 | 6,2171E-01 | 7,1435E-01 | 4,0036E-01 | 1 |
| hsa00640 | Propanoate metabolism | 1/255 | 32/8538 | 0,0313 | 1,0463 | 0,0461 | 6,2171E-01 | 7,1435E-01 | 4,0036E-01 | 1 |
| hsa04215 | Apoptosis - multiple species | 1/255 | 32/8538 | 0,0313 | 1,0463 | 0,0461 | 6,2171E-01 | 7,1435E-01 | 4,0036E-01 | 1 |
| hsa04622 | RIG-I-like receptor signaling pathway | 2/255 | 72/8538 | 0,0278 | 0,9301 | -0,1046 | 6,3874E-01 | 7,3106E-01 | 4,0972E-01 | 2 |
| hsa00051 | Fructose and mannose metabolism | 1/255 | 34/8538 | 0,0294 | 0,9848 | -0,0156 | 6,4405E-01 | 7,3427E-01 | 4,1152E-01 | 1 |
| hsa04360 | Axon guidance | 5/255 | 184/8538 | 0,0272 | 0,9098 | -0,2169 | 6,4784E-01 | 7,3572E-01 | 4,1233E-01 | 5 |
| hsa01230 | Biosynthesis of amino acids | 2/255 | 75/8538 | 0,0267 | 0,8929 | -0,1635 | 6,6084E-01 | 7,4759E-01 | 4,1898E-01 | 2 |
| hsa03320 | PPAR signaling pathway | 2/255 | 76/8538 | 0,0263 | 0,8811 | -0,1827 | 6,6796E-01 | 7,5274E-01 | 4,2187E-01 | 2 |
| hsa01200 | Carbon metabolism | 3/255 | 116/8538 | 0,0259 | 0,8659 | -0,2551 | 6,7857E-01 | 7,6177E-01 | 4,2693E-01 | 3 |
| hsa00760 | Nicotinate and nicotinamide metabolism | 1/255 | 38/8538 | 0,0263 | 0,8811 | -0,1289 | 6,8487E-01 | 7,6299E-01 | 4,2761E-01 | 1 |

| | | | | | | | | | | |
|---|---|---|---|---|---|---|---|---|---|---|
| hsa05340 | Primary immunodeficiency | 1/255 | 38/8538 | 0,0263 | 0,8811 | -0,1289 | 6,8487E-01 | 7,6299E-01 | 4,2761E-01 | 1 |
| hsa04062 | Chemokine signaling pathway | 5/255 | 193/8538 | 0,0259 | 0,8674 | -0,3269 | 6,8891E-01 | 7,6458E-01 | 4,2851E-01 | 5 |
| hsa00670 | One carbon pool by folate | 1/255 | 39/8538 | 0,0256 | 0,8585 | -0,1554 | 6,9432E-01 | 7,6565E-01 | 4,2911E-01 | 1 |
| hsa04932 | Non-alcoholic fatty liver disease | 4/255 | 157/8538 | 0,0255 | 0,8531 | -0,3261 | 6,9510E-01 | 7,6565E-01 | 4,2911E-01 | 4 |
| hsa00513 | Various types of N-glycan biosynthesis | 1/255 | 42/8538 | 0,0238 | 0,7972 | -0,2312 | 7,2102E-01 | 7,9123E-01 | 4,4344E-01 | 1 |
| hsa01232 | Nucleotide metabolism | 2/255 | 85/8538 | 0,0235 | 0,7878 | -0,3449 | 7,2674E-01 | 7,9447E-01 | 4,4526E-01 | 2 |
| hsa04975 | Fat digestion and absorption | 1/255 | 43/8538 | 0,0233 | 0,7787 | -0,2553 | 7,2939E-01 | 7,9447E-01 | 4,4526E-01 | 1 |
| hsa03410 | Base excision repair | 1/255 | 44/8538 | 0,0227 | 0,7610 | -0,2789 | 7,3752E-01 | 7,9739E-01 | 4,4689E-01 | 1 |
| hsa04962 | Vasopressin-regulated water reabsorption | 1/255 | 44/8538 | 0,0227 | 0,7610 | -0,2789 | 7,3752E-01 | 7,9739E-01 | 4,4689E-01 | 1 |
| hsa03050 | Proteasome | 1/255 | 46/8538 | 0,0217 | 0,7279 | -0,3247 | 7,5304E-01 | 8,1118E-01 | 4,5462E-01 | 1 |
| hsa00600 | Sphingolipid metabolism | 1/255 | 54/8538 | 0,0185 | 0,6200 | -0,4914 | 8,0651E-01 | 8,6560E-01 | 4,8512E-01 | 1 |
| hsa05150 | Staphylococcus aureus infection | 2/255 | 102/8538 | 0,0196 | 0,6565 | -0,6123 | 8,1390E-01 | 8,7034E-01 | 4,8778E-01 | 2 |
| hsa04340 | Hedgehog signaling pathway | 1/255 | 56/8538 | 0,0179 | 0,5979 | -0,5297 | 8,1797E-01 | 8,7151E-01 | 4,8843E-01 | 1 |
| hsa01212 | Fatty acid metabolism | 1/255 | 57/8538 | 0,0175 | 0,5874 | -0,5484 | 8,2344E-01 | 8,7416E-01 | 4,8992E-01 | 1 |
| hsa04613 | Neutrophil extracellular trap formation | 4/255 | 193/8538 | 0,0207 | 0,6939 | -0,7546 | 8,3379E-01 | 8,7937E-01 | 4,9284E-01 | 4 |
| hsa04922 | Glucagon signaling pathway | 2/255 | 107/8538 | 0,0187 | 0,6258 | -0,6833 | 8,3435E-01 | 8,7937E-01 | 4,9284E-01 | 2 |
| hsa04914 | Progesterone-mediated oocyte maturation | 2/255 | 111/8538 | 0,0180 | 0,6033 | -0,7381 | 8,4923E-01 | 8,9185E-01 | 4,9983E-01 | 2 |
| hsa04145 | Phagosome | 3/255 | 159/8538 | 0,0189 | 0,6317 | -0,8224 | 8,5906E-01 | 8,9894E-01 | 5,0381E-01 | 3 |
| hsa05169 | Epstein-Barr virus infection | 4/255 | 204/8538 | 0,0196 | 0,6565 | -0,8712 | 8,6398E-01 | 9,0028E-01 | 5,0456E-01 | 4 |
| hsa05203 | Viral carcinogenesis | 4/255 | 205/8538 | 0,0195 | 0,6533 | -0,8815 | 8,6648E-01 | 9,0028E-01 | 5,0456E-01 | 4 |
| hsa04144 | Endocytosis | 5/255 | 252/8538 | 0,0198 | 0,6643 | -0,9490 | 8,7741E-01 | 9,0841E-01 | 5,0911E-01 | 5 |
| hsa05416 | Viral myocarditis | 1/255 | 70/8538 | 0,0143 | 0,4783 | -0,7689 | 8,8131E-01 | 9,0924E-01 | 5,0958E-01 | 1 |
| hsa04612 | Antigen processing and presentation | 1/255 | 81/8538 | 0,0123 | 0,4134 | -0,9308 | 9,1523E-01 | 9,3975E-01 | 5,2668E-01 | 1 |
| hsa03083 | Polycomb repressive complex | 1/255 | 83/8538 | 0,0120 | 0,4034 | -0,9583 | 9,2026E-01 | 9,3975E-01 | 5,2668E-01 | 1 |
| hsa04820 | Cytoskeleton in muscle cells | 4/255 | 232/8538 | 0,0172 | 0,5773 | -1,1453 | 9,2051E-01 | 9,3975E-01 | 5,2668E-01 | 4 |
| hsa04120 | Ubiquitin mediated proteolysis | 2/255 | 142/8538 | 0,0141 | 0,4716 | -1,1141 | 9,2914E-01 | 9,4527E-01 | 5,2977E-01 | 2 |
| hsa04514 | Cell adhesion molecules | 2/255 | 158/8538 | 0,0127 | 0,4238 | -1,2826 | 9,5270E-01 | 9,6589E-01 | 5,4133E-01 | 2 |
| hsa03013 | Nucleocytoplasmic transport | 1/255 | 108/8538 | 0,0093 | 0,3100 | -1,2661 | 9,6296E-01 | 9,7189E-01 | 5,4469E-01 | 1 |
| hsa04141 | Protein processing in endoplasmic reticulum | 2/255 | 170/8538 | 0,0118 | 0,3939 | -1,4005 | 9,6526E-01 | 9,7189E-01 | 5,4469E-01 | 2 |
| hsa04714 | Thermogenesis | 3/255 | 235/8538 | 0,0128 | 0,4274 | -1,5616 | 9,7388E-01 | 9,7721E-01 | 5,4768E-01 | 3 |
| hsa04814 | Motor proteins | 1/255 | 197/8538 | 0,0051 | 0,1700 | -2,0680 | 9,9763E-01 | 9,9763E-01 | 5,5912E-01 | 1 |

**Table S13. Pathway enrichment analysis of genes harboring rare unique NDC-specific variants**. Analysis was performed using the R package ClusterProfiler and p-values were adjusted by the FDR method.

| | Description | GeneRatio | BgRatio | RichFactor | FoldEnrichment | zScore | pvalue | p.adjust | qvalue | Count |
|---|---|---|---|---|---|---|---|---|---|---|
| hsa04728 | Dopaminergic synapse | 15/314 | 132/9392 | 0,1136 | 3,3990 | 5,1621 | 0,0000 | 0,0095 | 0,0076 | 15 |
| hsa04310 | Wnt signaling pathway | 17/314 | 174/9392 | 0,0977 | 2,9223 | 4,7600 | 0,0001 | 0,0101 | 0,0082 | 17 |
| hsa04921 | Oxytocin signaling pathway | 15/314 | 155/9392 | 0,0968 | 2,8946 | 4,4233 | 0,0002 | 0,0199 | 0,0161 | 15 |
| hsa04929 | GnRH secretion | 9/314 | 65/9392 | 0,1385 | 4,1415 | 4,7266 | 0,0003 | 0,0207 | 0,0167 | 9 |
| hsa04070 | Phosphatidylinositol signaling system | 11/314 | 98/9392 | 0,1122 | 3,3573 | 4,3627 | 0,0004 | 0,0232 | 0,0188 | 11 |
| hsa04725 | Cholinergic synapse | 12/314 | 116/9392 | 0,1034 | 3,0942 | 4,2208 | 0,0005 | 0,0232 | 0,0188 | 12 |
| hsa04071 | Sphingolipid signaling pathway | 12/314 | 122/9392 | 0,0984 | 2,9420 | 4,0154 | 0,0008 | 0,0269 | 0,0218 | 12 |
| hsa04972 | Pancreatic secretion | 11/314 | 106/9392 | 0,1038 | 3,1040 | 4,0514 | 0,0008 | 0,0269 | 0,0218 | 11 |
| hsa04918 | Thyroid hormone synthesis | 9/314 | 75/9392 | 0,1200 | 3,5893 | 4,1870 | 0,0008 | 0,0269 | 0,0218 | 9 |
| hsa04010 | MAPK signaling pathway | 21/314 | 300/9392 | 0,0700 | 2,0938 | 3,5808 | 0,0011 | 0,0317 | 0,0257 | 21 |
| hsa04360 | Axon guidance | 15/314 | 184/9392 | 0,0815 | 2,4384 | 3,6646 | 0,0013 | 0,0333 | 0,0269 | 15 |
| hsa04925 | Aldosterone synthesis and secretion | 10/314 | 98/9392 | 0,1020 | 3,0521 | 3,7979 | 0,0016 | 0,0377 | 0,0305 | 10 |
| hsa04261 | Adrenergic signaling in cardiomyocytes | 13/314 | 154/9392 | 0,0844 | 2,5249 | 3,5485 | 0,0019 | 0,0393 | 0,0318 | 13 |
| hsa04933 | AGE-RAGE signaling pathway in diabetic complications | 10/314 | 101/9392 | 0,0990 | 2,9615 | 3,6858 | 0,0020 | 0,0393 | 0,0318 | 10 |
| hsa04062 | Chemokine signaling pathway | 15/314 | 193/9392 | 0,0777 | 2,3247 | 3,4581 | 0,0020 | 0,0393 | 0,0318 | 15 |
| hsa04120 | Ubiquitin mediated proteolysis | 12/314 | 142/9392 | 0,0845 | 2,5277 | 3,4114 | 0,0028 | 0,0510 | 0,0412 | 12 |
| hsa04022 | cGMP-PKG signaling pathway | 13/314 | 166/9392 | 0,0783 | 2,3424 | 3,2453 | 0,0037 | 0,0611 | 0,0494 | 13 |
| hsa04520 | Adherens junction | 9/314 | 93/9392 | 0,0968 | 2,8946 | 3,4148 | 0,0038 | 0,0611 | 0,0494 | 9 |
| hsa04713 | Circadian entrainment | 9/314 | 97/9392 | 0,0928 | 2,7752 | 3,2684 | 0,0050 | 0,0715 | 0,0578 | 9 |
| hsa04020 | Calcium signaling pathway | 17/314 | 254/9392 | 0,0669 | 2,0019 | 3,0105 | 0,0050 | 0,0715 | 0,0578 | 17 |
| hsa04270 | Vascular smooth muscle contraction | 11/314 | 134/9392 | 0,0821 | 2,4554 | 3,1557 | 0,0052 | 0,0715 | 0,0578 | 11 |
| hsa04934 | Cushing syndrome | 12/314 | 155/9392 | 0,0774 | 2,3157 | 3,0717 | 0,0057 | 0,0725 | 0,0586 | 12 |
| hsa04927 | Cortisol synthesis and secretion | 7/314 | 65/9392 | 0,1077 | 3,2212 | 3,3419 | 0,0058 | 0,0725 | 0,0586 | 7 |
| hsa04814 | Motor proteins | 14/314 | 197/9392 | 0,0711 | 2,1256 | 2,9695 | 0,0063 | 0,0735 | 0,0595 | 14 |
| hsa00512 | Mucin type O-glycan biosynthesis | 5/314 | 36/9392 | 0,1389 | 4,1543 | 3,5264 | 0,0065 | 0,0735 | 0,0595 | 5 |
| hsa04720 | Long-term potentiation | 7/314 | 67/9392 | 0,1045 | 3,1250 | 3,2464 | 0,0068 | 0,0735 | 0,0595 | 7 |
| hsa04082 | Neuroactive ligand signaling | 14/314 | 199/9392 | 0,0704 | 2,1043 | 2,9282 | 0,0069 | 0,0735 | 0,0595 | 14 |
| hsa04973 | Carbohydrate digestion and absorption | 6/314 | 52/9392 | 0,1154 | 3,4512 | 3,2964 | 0,0074 | 0,0735 | 0,0595 | 6 |
| hsa04152 | AMPK signaling pathway | 10/314 | 122/9392 | 0,0820 | 2,4517 | 3,0015 | 0,0076 | 0,0735 | 0,0595 | 10 |
| hsa04660 | T cell receptor signaling pathway | 10/314 | 122/9392 | 0,0820 | 2,4517 | 3,0015 | 0,0076 | 0,0735 | 0,0595 | 10 |
| hsa04960 | Aldosterone-regulated sodium reabsorption | 5/314 | 38/9392 | 0,1316 | 3,9356 | 3,3723 | 0,0082 | 0,0765 | 0,0619 | 5 |
| hsa04611 | Platelet activation | 10/314 | 126/9392 | 0,0794 | 2,3739 | 2,8874 | 0,0095 | 0,0858 | 0,0694 | 10 |
| hsa04211 | Longevity regulating pathway | 8/314 | 90/9392 | 0,0889 | 2,6587 | 2,9406 | 0,0103 | 0,0900 | 0,0728 | 8 |
| hsa00562 | Inositol phosphate metabolism | 7/314 | 73/9392 | 0,0959 | 2,8682 | 2,9800 | 0,0108 | 0,0916 | 0,0741 | 7 |
| hsa04971 | Gastric acid secretion | 7/314 | 76/9392 | 0,0921 | 2,7549 | 2,8568 | 0,0133 | 0,1097 | 0,0887 | 7 |
| hsa05167 | Kaposi sarcoma-associated herpesvirus infection | 13/314 | 196/9392 | 0,0663 | 1,9839 | 2,5888 | 0,0143 | 0,1122 | 0,0907 | 13 |
| hsa04730 | Long-term depression | 6/314 | 60/9392 | 0,1000 | 2,9911 | 2,8774 | 0,0146 | 0,1122 | 0,0907 | 6 |
| hsa04928 | Parathyroid hormone synthesis, secretion and action | 9/314 | 115/9392 | 0,0783 | 2,3408 | 2,6906 | 0,0147 | 0,1122 | 0,0907 | 9 |
| hsa04724 | Glutamatergic synapse | 9/314 | 116/9392 | 0,0776 | 2,3207 | 2,6618 | 0,0155 | 0,1151 | 0,0931 | 9 |
| hsa04148 | Efferocytosis | 11/314 | 157/9392 | 0,0701 | 2,0957 | 2,5747 | 0,0161 | 0,1166 | 0,0943 | 11 |
| hsa05135 | Yersinia infection | 10/314 | 138/9392 | 0,0725 | 2,1675 | 2,5694 | 0,0172 | 0,1181 | 0,0955 | 10 |
| hsa04750 | Inflammatory mediator regulation of TRP channels | 8/314 | 99/9392 | 0,0808 | 2,4170 | 2,6360 | 0,0176 | 0,1181 | 0,0955 | 8 |
| hsa05231 | Choline metabolism in cancer | 8/314 | 99/9392 | 0,0808 | 2,4170 | 2,6360 | 0,0176 | 0,1181 | 0,0955 | 8 |
| hsa04722 | Neurotrophin signaling pathway | 9/314 | 120/9392 | 0,0750 | 2,2433 | 2,5492 | 0,0190 | 0,1248 | 0,1009 | 9 |
| hsa00514 | Other types of O-glycan biosynthesis | 5/314 | 47/9392 | 0,1064 | 3,1820 | 2,7889 | 0,0197 | 0,1264 | 0,1022 | 5 |
| hsa00280 | Valine, leucine and isoleucine degradation | 5/314 | 48/9392 | 0,1042 | 3,1157 | 2,7330 | 0,0214 | 0,1312 | 0,1061 | 5 |
| hsa04380 | Osteoclast differentiation | 10/314 | 143/9392 | 0,0699 | 2,0917 | 2,4464 | 0,0215 | 0,1312 | 0,1061 | 10 |
| hsa05146 | Amoebiasis | 8/314 | 103/9392 | 0,0777 | 2,3232 | 2,5112 | 0,0218 | 0,1312 | 0,1061 | 8 |
| hsa04911 | Insulin secretion | 7/314 | 86/9392 | 0,0814 | 2,4346 | 2,4856 | 0,0247 | 0,1456 | 0,1177 | 7 |
| hsa04924 | Renin secretion | 6/314 | 69/9392 | 0,0870 | 2,6009 | 2,4823 | 0,0274 | 0,1538 | 0,1243 | 6 |
| hsa00230 | Purine metabolism | 9/314 | 128/9392 | 0,0703 | 2,1031 | 2,3369 | 0,0275 | 0,1538 | 0,1243 | 9 |
| hsa04072 | Phospholipase D signaling pathway | 10/314 | 149/9392 | 0,0671 | 2,0074 | 2,3053 | 0,0277 | 0,1538 | 0,1243 | 10 |
| hsa05417 | Lipid and atherosclerosis | 13/314 | 216/9392 | 0,0602 | 1,8002 | 2,2127 | 0,0292 | 0,1594 | 0,1289 | 13 |
| hsa04926 | Relaxin signaling pathway | 9/314 | 130/9392 | 0,0692 | 2,0707 | 2,2863 | 0,0301 | 0,1608 | 0,1300 | 9 |
| hsa00532 | Glycosaminoglycan biosynthesis - chondroitin sulfate / dermatan sulfate | 3/314 | 21/9392 | 0,1429 | 4,2730 | 2,7924 | 0,0315 | 0,1612 | 0,1303 | 3 |
| hsa00770 | Pantothenate and CoA biosynthesis | 3/314 | 21/9392 | 0,1429 | 4,2730 | 2,7924 | 0,0315 | 0,1612 | 0,1303 | 3 |
| hsa04081 | Hormone signaling | 13/314 | 219/9392 | 0,0594 | 1,7755 | 2,1597 | 0,0322 | 0,1612 | 0,1303 | 13 |
| hsa05032 | Morphine addiction | 7/314 | 91/9392 | 0,0769 | 2,3008 | 2,3190 | 0,0323 | 0,1612 | 0,1303 | 7 |
| hsa05143 | African trypanosomiasis | 4/314 | 37/9392 | 0,1081 | 3,2336 | 2,5317 | 0,0340 | 0,1661 | 0,1343 | 4 |
| hsa04151 | PI3K-Akt signaling pathway | 19/314 | 362/9392 | 0,0525 | 1,5699 | 2,0565 | 0,0345 | 0,1661 | 0,1343 | 19 |
| hsa04650 | Natural killer cell mediated cytotoxicity | 9/314 | 134/9392 | 0,0672 | 2,0089 | 2,1877 | 0,0355 | 0,1683 | 0,1361 | 9 |
| hsa04340 | Hedgehog signaling pathway | 5/314 | 56/9392 | 0,0893 | 2,6706 | 2,3319 | 0,0385 | 0,1796 | 0,1452 | 5 |
| hsa05163 | Human cytomegalovirus infection | 13/314 | 226/9392 | 0,0575 | 1,7205 | 2,0391 | 0,0399 | 0,1805 | 0,1459 | 13 |
| hsa00515 | Mannose type O-glycan biosynthesis | 3/314 | 23/9392 | 0,1304 | 3,9014 | 2,5909 | 0,0400 | 0,1805 | 0,1459 | 3 |
| hsa04970 | Salivary secretion | 7/314 | 97/9392 | 0,0722 | 2,1585 | 2,1330 | 0,0435 | 0,1932 | 0,1563 | 7 |
| hsa04371 | Apelin signaling pathway | 9/314 | 140/9392 | 0,0643 | 1,9228 | 2,0460 | 0,0450 | 0,1969 | 0,1592 | 9 |
| hsa04666 | Fc gamma R-mediated phagocytosis | 7/314 | 99/9392 | 0,0707 | 2,1149 | 2,0740 | 0,0477 | 0,2056 | 0,1662 | 7 |
| hsa04935 | Growth hormone synthesis, secretion and action | 8/314 | 122/9392 | 0,0656 | 1,9614 | 1,9877 | 0,0517 | 0,2187 | 0,1768 | 8 |
| hsa04015 | Rap1 signaling pathway | 12/314 | 212/9392 | 0,0566 | 1,6931 | 1,8982 | 0,0522 | 0,2187 | 0,1768 | 12 |
| hsa00513 | Various types of N-glycan biosynthesis | 4/314 | 43/9392 | 0,0930 | 2,7824 | 2,1786 | 0,0545 | 0,2218 | 0,1793 | 4 |
| hsa04014 | Ras signaling pathway | 13/314 | 238/9392 | 0,0546 | 1,6338 | 1,8418 | 0,0561 | 0,2218 | 0,1793 | 13 |

| | | | | | | | | | | |
|---|---|---|---|---|---|---|---|---|---|---|
| hsa00564 | Glycerophospholipid metabolism | 7/314 | 103/9392 | 0,0680 | 2,0328 | 1,9600 | 0,0568 | 0,2218 | 0,1793 | 7 |
| hsa03015 | mRNA surveillance pathway | 7/314 | 103/9392 | 0,0680 | 2,0328 | 1,9600 | 0,0568 | 0,2218 | 0,1793 | 7 |
| hsa05142 | Chagas disease | 7/314 | 103/9392 | 0,0680 | 2,0328 | 1,9600 | 0,0568 | 0,2218 | 0,1793 | 7 |
| hsa04530 | Tight junction | 10/314 | 170/9392 | 0,0588 | 1,7595 | 1,8584 | 0,0589 | 0,2269 | 0,1835 | 10 |
| hsa02010 | ABC transporters | 4/314 | 45/9392 | 0,0889 | 2,6587 | 2,0743 | 0,0625 | 0,2378 | 0,1923 | 4 |
| hsa01232 | Nucleotide metabolism | 6/314 | 85/9392 | 0,0706 | 2,1114 | 1,9142 | 0,0646 | 0,2426 | 0,1961 | 6 |
| hsa04922 | Glucagon signaling pathway | 7/314 | 107/9392 | 0,0654 | 1,9568 | 1,8511 | 0,0669 | 0,2443 | 0,1975 | 7 |
| hsa04742 | Taste transduction | 6/314 | 86/9392 | 0,0698 | 2,0868 | 1,8830 | 0,0676 | 0,2443 | 0,1975 | 6 |
| hsa05412 | Arrhythmogenic right ventricular cardiomyopathy | 6/314 | 86/9392 | 0,0698 | 2,0868 | 1,8830 | 0,0676 | 0,2443 | 0,1975 | 6 |
| hsa05210 | Colorectal cancer | 6/314 | 87/9392 | 0,0690 | 2,0628 | 1,8522 | 0,0707 | 0,2507 | 0,2027 | 6 |
| hsa04930 | Type II diabetes mellitus | 4/314 | 47/9392 | 0,0851 | 2,5456 | 1,9755 | 0,0711 | 0,2507 | 0,2027 | 4 |
| hsa04066 | HIF-1 signaling pathway | 7/314 | 110/9392 | 0,0636 | 1,9034 | 1,7725 | 0,0752 | 0,2619 | 0,2118 | 7 |
| hsa00533 | Glycosaminoglycan biosynthesis - keratan sulfate | 2/314 | 14/9392 | 0,1429 | 4,2730 | 2,2792 | 0,0778 | 0,2642 | 0,2136 | 2 |
| hsa04144 | Endocytosis | 13/314 | 252/9392 | 0,0516 | 1,5430 | 1,6251 | 0,0801 | 0,2642 | 0,2136 | 13 |
| hsa04390 | Hippo signaling pathway | 9/314 | 157/9392 | 0,0573 | 1,7146 | 1,6793 | 0,0803 | 0,2642 | 0,2136 | 9 |
| hsa05031 | Amphetamine addiction | 5/314 | 69/9392 | 0,0725 | 2,1675 | 1,8101 | 0,0804 | 0,2642 | 0,2136 | 5 |
| hsa05205 | Proteoglycans in cancer | 11/314 | 204/9392 | 0,0539 | 1,6128 | 1,6458 | 0,0804 | 0,2642 | 0,2136 | 11 |
| hsa05203 | Viral carcinogenesis | 11/314 | 205/9392 | 0,0537 | 1,6050 | 1,6287 | 0,0826 | 0,2659 | 0,2150 | 11 |
| hsa04150 | mTOR signaling pathway | 9/314 | 158/9392 | 0,0570 | 1,7038 | 1,6592 | 0,0828 | 0,2659 | 0,2150 | 9 |
| hsa04662 | B cell receptor signaling pathway | 6/314 | 91/9392 | 0,0659 | 1,9721 | 1,7331 | 0,0838 | 0,2661 | 0,2152 | 6 |
| hsa04540 | Gap junction | 6/314 | 92/9392 | 0,0652 | 1,9507 | 1,7042 | 0,0873 | 0,2731 | 0,2208 | 6 |
| hsa00730 | Thiamine metabolism | 2/314 | 15/9392 | 0,1333 | 3,9881 | 2,1540 | 0,0879 | 0,2731 | 0,2208 | 2 |
| hsa04810 | Regulation of actin cytoskeleton | 12/314 | 232/9392 | 0,0517 | 1,5471 | 1,5693 | 0,0889 | 0,2733 | 0,2210 | 12 |
| hsa04726 | Serotonergic synapse | 7/314 | 115/9392 | 0,0609 | 1,8207 | 1,6468 | 0,0903 | 0,2734 | 0,2211 | 7 |
| hsa04912 | GnRH signaling pathway | 6/314 | 93/9392 | 0,0645 | 1,9297 | 1,6757 | 0,0908 | 0,2734 | 0,2211 | 6 |
| hsa05161 | Hepatitis B | 9/314 | 163/9392 | 0,0552 | 1,6515 | 1,5605 | 0,0959 | 0,2859 | 0,2312 | 9 |
| hsa00603 | Glycosphingolipid biosynthesis - globo and isoglobo series | 2/314 | 16/9392 | 0,1250 | 3,7389 | 2,0391 | 0,0983 | 0,2898 | 0,2344 | 2 |
| hsa05017 | Spinocerebellar ataxia | 8/314 | 144/9392 | 0,0556 | 1,6617 | 1,4882 | 0,1095 | 0,3182 | 0,2573 | 8 |
| hsa05214 | Glioma | 5/314 | 76/9392 | 0,0658 | 1,9678 | 1,5755 | 0,1101 | 0,3182 | 0,2573 | 5 |
| hsa05410 | Hypertrophic cardiomyopathy | 6/314 | 99/9392 | 0,0606 | 1,8128 | 1,5119 | 0,1137 | 0,3233 | 0,2614 | 6 |
| hsa04919 | Thyroid hormone signaling pathway | 7/314 | 122/9392 | 0,0574 | 1,7162 | 1,4808 | 0,1141 | 0,3233 | 0,2614 | 7 |
| hsa05225 | Hepatocellular carcinoma | 9/314 | 170/9392 | 0,0529 | 1,5835 | 1,4279 | 0,1163 | 0,3262 | 0,2638 | 9 |
| hsa05100 | Bacterial invasion of epithelial cells | 5/314 | 78/9392 | 0,0641 | 1,9174 | 1,5130 | 0,1194 | 0,3319 | 0,2684 | 5 |
| hsa04916 | Melanogenesis | 6/314 | 101/9392 | 0,0594 | 1,7769 | 1,4599 | 0,1219 | 0,3346 | 0,2705 | 6 |
| hsa05224 | Breast cancer | 8/314 | 148/9392 | 0,0541 | 1,6168 | 1,4066 | 0,1227 | 0,3346 | 0,2705 | 8 |
| hsa04723 | Retrograde endocannabinoid signaling | 8/314 | 149/9392 | 0,0537 | 1,6060 | 1,3866 | 0,1262 | 0,3408 | 0,2755 | 8 |
| hsa01521 | EGFR tyrosine kinase inhibitor resistance | 5/314 | 80/9392 | 0,0625 | 1,8694 | 1,4524 | 0,1291 | 0,3456 | 0,2794 | 5 |
| hsa05213 | Endometrial cancer | 4/314 | 59/9392 | 0,0678 | 2,0279 | 1,4729 | 0,1342 | 0,3558 | 0,2877 | 4 |
| hsa04024 | cAMP signaling pathway | 11/314 | 226/9392 | 0,0487 | 1,4558 | 1,2900 | 0,1363 | 0,3581 | 0,2896 | 11 |
| hsa04625 | C-type lectin receptor signaling pathway | 6/314 | 105/9392 | 0,0571 | 1,7092 | 1,3591 | 0,1392 | 0,3587 | 0,2900 | 6 |
| hsa05414 | Dilated cardiomyopathy | 6/314 | 105/9392 | 0,0571 | 1,7092 | 1,3591 | 0,1392 | 0,3587 | 0,2900 | 6 |
| hsa04370 | VEGF signaling pathway | 4/314 | 60/9392 | 0,0667 | 1,9941 | 1,4366 | 0,1403 | 0,3587 | 0,2900 | 4 |
| hsa04510 | Focal adhesion | 10/314 | 203/9392 | 0,0493 | 1,4734 | 1,2682 | 0,1426 | 0,3616 | 0,2924 | 10 |
| hsa05165 | Human papillomavirus infection | 15/314 | 333/9392 | 0,0450 | 1,3473 | 1,2002 | 0,1482 | 0,3714 | 0,3003 | 15 |
| hsa05415 | Diabetic cardiomyopathy | 10/314 | 205/9392 | 0,0488 | 1,4591 | 1,2359 | 0,1491 | 0,3714 | 0,3003 | 10 |
| hsa04213 | Longevity regulating pathway - multiple species | 4/314 | 62/9392 | 0,0645 | 1,9297 | 1,3660 | 0,1527 | 0,3771 | 0,3049 | 4 |
| hsa00590 | Arachidonic acid metabolism | 4/314 | 63/9392 | 0,0635 | 1,8991 | 1,3316 | 0,1590 | 0,3862 | 0,3123 | 4 |
| hsa05217 | Basal cell carcinoma | 4/314 | 63/9392 | 0,0635 | 1,8991 | 1,3316 | 0,1590 | 0,3862 | 0,3123 | 4 |
| hsa05170 | Human immunodeficiency virus 1 infection | 10/314 | 213/9392 | 0,0469 | 1,4043 | 1,1099 | 0,1763 | 0,4246 | 0,3433 | 10 |
| hsa04114 | Oocyte meiosis | 7/314 | 139/9392 | 0,0504 | 1,5063 | 1,1184 | 0,1837 | 0,4388 | 0,3548 | 7 |
| hsa05022 | Pathways of neurodegeneration - multiple diseases | 20/314 | 483/9392 | 0,0414 | 1,2385 | 1,0010 | 0,1892 | 0,4479 | 0,3622 | 20 |
| hsa04670 | Leukocyte transendothelial migration | 6/314 | 116/9392 | 0,0517 | 1,5471 | 1,1027 | 0,1921 | 0,4479 | 0,3622 | 6 |
| hsa05221 | Acute myeloid leukemia | 4/314 | 68/9392 | 0,0588 | 1,7595 | 1,1689 | 0,1922 | 0,4479 | 0,3622 | 4 |
| hsa05211 | Renal cell carcinoma | 4/314 | 70/9392 | 0,0571 | 1,7092 | 1,1076 | 0,2060 | 0,4763 | 0,3851 | 4 |
| hsa04977 | Vitamin digestion and absorption | 2/314 | 26/9392 | 0,0769 | 2,3008 | 1,2353 | 0,2153 | 0,4939 | 0,3994 | 2 |
| hsa05218 | Melanoma | 4/314 | 73/9392 | 0,0548 | 1,6389 | 1,0192 | 0,2273 | 0,5171 | 0,4182 | 4 |
| hsa00565 | Ether lipid metabolism | 3/314 | 50/9392 | 0,0600 | 1,7946 | 1,0478 | 0,2335 | 0,5273 | 0,4264 | 3 |
| hsa05226 | Gastric cancer | 7/314 | 150/9392 | 0,0467 | 1,3958 | 0,9089 | 0,2363 | 0,5294 | 0,4280 | 7 |
| hsa00601 | Glycosphingolipid biosynthesis - lacto and neolacto series | 2/314 | 28/9392 | 0,0714 | 2,1365 | 1,1201 | 0,2403 | 0,5333 | 0,4312 | 2 |
| hsa04115 | p53 signaling pathway | 4/314 | 75/9392 | 0,0533 | 1,5952 | 0,9625 | 0,2417 | 0,5333 | 0,4312 | 4 |
| hsa04744 | Phototransduction | 2/314 | 29/9392 | 0,0690 | 2,0628 | 1,0660 | 0,2528 | 0,5534 | 0,4475 | 2 |
| hsa05212 | Pancreatic cancer | 4/314 | 77/9392 | 0,0519 | 1,5538 | 0,9075 | 0,2564 | 0,5571 | 0,4505 | 4 |
| hsa04961 | Endocrine and other factor-regulated calcium reabsorption | 3/314 | 53/9392 | 0,0566 | 1,6931 | 0,9410 | 0,2606 | 0,5620 | 0,4545 | 3 |
| hsa01523 | Antifolate resistance | 2/314 | 30/9392 | 0,0667 | 1,9941 | 1,0142 | 0,2653 | 0,5679 | 0,4592 | 2 |
| hsa04110 | Cell cycle | 7/314 | 158/9392 | 0,0443 | 1,3252 | 0,7666 | 0,2773 | 0,5861 | 0,4739 | 7 |
| hsa04981 | Folate transport and metabolism | 2/314 | 31/9392 | 0,0645 | 1,9297 | 0,9643 | 0,2778 | 0,5861 | 0,4739 | 2 |
| hsa00052 | Galactose metabolism | 2/314 | 32/9392 | 0,0625 | 1,8694 | 0,9162 | 0,2903 | 0,5994 | 0,4846 | 2 |
| hsa00640 | Propanoate metabolism | 2/314 | 32/9392 | 0,0625 | 1,8694 | 0,9162 | 0,2903 | 0,5994 | 0,4846 | 2 |
| hsa04215 | Apoptosis - multiple species | 2/314 | 32/9392 | 0,0625 | 1,8694 | 0,9162 | 0,2903 | 0,5994 | 0,4846 | 2 |
| hsa03013 | Nucleocytoplasmic transport | 5/314 | 108/9392 | 0,0463 | 1,3848 | 0,7479 | 0,2938 | 0,6023 | 0,4870 | 5 |
| hsa04620 | Toll-like receptor signaling pathway | 5/314 | 109/9392 | 0,0459 | 1,3721 | 0,7266 | 0,3004 | 0,6071 | 0,4909 | 5 |
| hsa04931 | Insulin resistance | 5/314 | 109/9392 | 0,0459 | 1,3721 | 0,7266 | 0,3004 | 0,6071 | 0,4909 | 5 |
| hsa04130 | SNARE interactions in vesicular transport | 2/314 | 33/9392 | 0,0606 | 1,8128 | 0,8698 | 0,3028 | 0,6078 | 0,4914 | 2 |

| | | | | | | | | | | |
|---|---|---|---|---|---|---|---|---|---|---|
| hsa04923 | Regulation of lipolysis in adipocytes | 3/314 | 59/9392 | 0,0508 | 1,5209 | 0,7464 | 0,3157 | 0,6292 | 0,5088 | 3 |
| hsa04915 | Estrogen signaling pathway | 6/314 | 139/9392 | 0,0432 | 1,2911 | 0,6431 | 0,3208 | 0,6340 | 0,5126 | 6 |
| hsa04012 | ErbB signaling pathway | 4/314 | 86/9392 | 0,0465 | 1,3912 | 0,6778 | 0,3242 | 0,6340 | 0,5126 | 4 |
| hsa04630 | JAK-STAT signaling pathway | 7/314 | 168/9392 | 0,0417 | 1,2463 | 0,5990 | 0,3307 | 0,6340 | 0,5126 | 7 |
| hsa04260 | Cardiac muscle contraction | 4/314 | 87/9392 | 0,0460 | 1,3752 | 0,6539 | 0,3318 | 0,6340 | 0,5126 | 4 |
| hsa04613 | Neutrophil extracellular trap formation | 8/314 | 196/9392 | 0,0408 | 1,2209 | 0,5811 | 0,3331 | 0,6340 | 0,5126 | 8 |
| hsa05166 | Human T-cell leukemia virus 1 infection | 9/314 | 224/9392 | 0,0402 | 1,2018 | 0,5684 | 0,3344 | 0,6340 | 0,5126 | 9 |
| hsa03265 | Virion - Ebolavirus, Lyssavirus and Morbillivirus | 1/314 | Dec-92 | 0,0833 | 2,4926 | 0,9622 | 0,3352 | 0,6340 | 0,5126 | 1 |
| hsa05131 | Shigellosis | 10/314 | 253/9392 | 0,0395 | 1,1822 | 0,5465 | 0,3394 | 0,6340 | 0,5126 | 10 |
| hsa04610 | Complement and coagulation cascades | 4/314 | 88/9392 | 0,0455 | 1,3596 | 0,6303 | 0,3394 | 0,6340 | 0,5126 | 4 |
| hsa03030 | DNA replication | 2/314 | 36/9392 | 0,0556 | 1,6617 | 0,7398 | 0,3400 | 0,6340 | 0,5126 | 2 |
| hsa04330 | Notch signaling pathway | 3/314 | 62/9392 | 0,0484 | 1,4473 | 0,6572 | 0,3434 | 0,6361 | 0,5144 | 3 |
| hsa04727 | GABAergic synapse | 4/314 | 89/9392 | 0,0449 | 1,3443 | 0,6070 | 0,3471 | 0,6389 | 0,5166 | 4 |
| hsa05216 | Thyroid cancer | 2/314 | 37/9392 | 0,0541 | 1,6168 | 0,6991 | 0,3523 | 0,6444 | 0,5210 | 2 |
| hsa05235 | PD-L1 expression and PD-1 checkpoint pathway in cancer | 4/314 | 90/9392 | 0,0444 | 1,3294 | 0,5839 | 0,3548 | 0,6448 | 0,5214 | 4 |
| hsa05164 | Influenza A | 7/314 | 173/9392 | 0,0405 | 1,2103 | 0,5191 | 0,3580 | 0,6467 | 0,5229 | 7 |
| hsa00561 | Glycerolipid metabolism | 3/314 | 65/9392 | 0,0462 | 1,3805 | 0,5725 | 0,3710 | 0,6659 | 0,5385 | 3 |
| hsa04658 | Th1 and Th2 cell differentiation | 4/314 | 93/9392 | 0,0430 | 1,2865 | 0,5164 | 0,3777 | 0,6738 | 0,5448 | 4 |
| hsa00604 | Glycosphingolipid biosynthesis - ganglio series | 1/314 | 15/9392 | 0,0667 | 1,9941 | 0,7166 | 0,3998 | 0,7057 | 0,5706 | 1 |
| hsa05033 | Nicotine addiction | 2/314 | 41/9392 | 0,0488 | 1,4591 | 0,5479 | 0,4005 | 0,7057 | 0,5706 | 2 |
| hsa04664 | Fc epsilon RI signaling pathway | 3/314 | 69/9392 | 0,0435 | 1,3005 | 0,4659 | 0,4074 | 0,7136 | 0,5770 | 3 |
| hsa01240 | Biosynthesis of cofactors | 6/314 | 154/9392 | 0,0390 | 1,1654 | 0,3848 | 0,4107 | 0,7150 | 0,5782 | 6 |
| hsa05215 | Prostate cancer | 4/314 | 98/9392 | 0,0408 | 1,2209 | 0,4087 | 0,4158 | 0,7164 | 0,5793 | 4 |
| hsa05416 | Viral myocarditis | 3/314 | 70/9392 | 0,0429 | 1,2819 | 0,4403 | 0,4164 | 0,7164 | 0,5793 | 3 |
| hsa01522 | Endocrine resistance | 4/314 | 99/9392 | 0,0404 | 1,2085 | 0,3879 | 0,4233 | 0,7232 | 0,5848 | 4 |
| hsa04917 | Prolactin signaling pathway | 3/314 | 71/9392 | 0,0423 | 1,2638 | 0,4150 | 0,4254 | 0,7232 | 0,5848 | 3 |
| hsa05010 | Alzheimer disease | 14/314 | 391/9392 | 0,0358 | 1,0710 | 0,2666 | 0,4351 | 0,7235 | 0,5850 | 14 |
| hsa03410 | Base excision repair | 2/314 | 44/9392 | 0,0455 | 1,3596 | 0,4446 | 0,4355 | 0,7235 | 0,5850 | 2 |
| hsa04962 | Vasopressin-regulated water reabsorption | 2/314 | 44/9392 | 0,0455 | 1,3596 | 0,4446 | 0,4355 | 0,7235 | 0,5850 | 2 |
| hsa00910 | Nitrogen metabolism | 1/314 | 17/9392 | 0,0588 | 1,7595 | 0,5829 | 0,4393 | 0,7235 | 0,5850 | 1 |
| hsa05207 | Chemical carcinogenesis - receptor activation | 8/314 | 217/9392 | 0,0369 | 1,1027 | 0,2847 | 0,4402 | 0,7235 | 0,5850 | 8 |
| hsa05160 | Hepatitis C | 6/314 | 159/9392 | 0,0377 | 1,1287 | 0,3044 | 0,4406 | 0,7235 | 0,5850 | 6 |
| hsa05223 | Non-small cell lung cancer | 3/314 | 73/9392 | 0,0411 | 1,2292 | 0,3656 | 0,4432 | 0,7237 | 0,5852 | 3 |
| hsa04514 | Cell adhesion molecules | 6/314 | 160/9392 | 0,0375 | 1,1217 | 0,2886 | 0,4466 | 0,7251 | 0,5863 | 6 |
| hsa04980 | Cobalamin transport and metabolism | 1/314 | 18/9392 | 0,0556 | 1,6617 | 0,5226 | 0,4581 | 0,7381 | 0,5969 | 1 |
| hsa04142 | Lysosome | 5/314 | 133/9392 | 0,0376 | 1,1245 | 0,2689 | 0,4597 | 0,7381 | 0,5969 | 5 |
| hsa04064 | NF-kappa B signaling pathway | 4/314 | 105/9392 | 0,0381 | 1,1395 | 0,2673 | 0,4681 | 0,7474 | 0,6043 | 4 |
| hsa00531 | Glycosaminoglycan degradation | 1/314 | 19/9392 | 0,0526 | 1,5743 | 0,4660 | 0,4762 | 0,7562 | 0,6115 | 1 |
| hsa05030 | Cocaine addiction | 2/314 | 49/9392 | 0,0408 | 1,2209 | 0,2883 | 0,4913 | 0,7684 | 0,6213 | 2 |
| hsa00100 | Steroid biosynthesis | 1/314 | 20/9392 | 0,0500 | 1,4955 | 0,4126 | 0,4938 | 0,7684 | 0,6213 | 1 |
| hsa00541 | Biosynthesis of various nucleotide sugars | 1/314 | 20/9392 | 0,0500 | 1,4955 | 0,4126 | 0,4938 | 0,7684 | 0,6213 | 1 |
| hsa04721 | Synaptic vesicle cycle | 3/314 | 79/9392 | 0,0380 | 1,1359 | 0,2255 | 0,4952 | 0,7684 | 0,6213 | 3 |
| hsa04659 | Th17 cell differentiation | 4/314 | 109/9392 | 0,0367 | 1,0976 | 0,1907 | 0,4972 | 0,7684 | 0,6213 | 4 |
| hsa05418 | Fluid shear stress and atherosclerosis | 5/314 | 142/9392 | 0,0352 | 1,0532 | 0,1188 | 0,5175 | 0,7914 | 0,6400 | 5 |
| hsa05130 | Pathogenic Escherichia coli infection | 7/314 | 203/9392 | 0,0345 | 1,0314 | 0,0841 | 0,5214 | 0,7914 | 0,6400 | 7 |
| hsa00270 | Cysteine and methionine metabolism | 2/314 | 52/9392 | 0,0385 | 1,1504 | 0,2023 | 0,5231 | 0,7914 | 0,6400 | 2 |
| hsa04913 | Ovarian steroidogenesis | 2/314 | 52/9392 | 0,0385 | 1,1504 | 0,2023 | 0,5231 | 0,7914 | 0,6400 | 2 |
| hsa03083 | Polycomb repressive complex | 3/314 | 83/9392 | 0,0361 | 1,0811 | 0,1380 | 0,5285 | 0,7936 | 0,6417 | 3 |
| hsa04550 | Signaling pathways regulating pluripotency of stem cells | 5/314 | 144/9392 | 0,0347 | 1,0386 | 0,0867 | 0,5300 | 0,7936 | 0,6417 | 5 |
| hsa03430 | Mismatch repair | 1/314 | 23/9392 | 0,0435 | 1,3005 | 0,2683 | 0,5430 | 0,8089 | 0,6541 | 1 |
| hsa03082 | ATP-dependent chromatin remodeling | 4/314 | 117/9392 | 0,0342 | 1,0226 | 0,0457 | 0,5533 | 0,8161 | 0,6599 | 4 |
| hsa00510 | N-Glycan biosynthesis | 2/314 | 55/9392 | 0,0364 | 1,0877 | 0,1213 | 0,5535 | 0,8161 | 0,6599 | 2 |
| hsa05152 | Tuberculosis | 6/314 | 182/9392 | 0,0330 | 0,9861 | -0,0353 | 0,5724 | 0,8397 | 0,6790 | 6 |
| hsa05020 | Prion disease | 9/314 | 278/9392 | 0,0324 | 0,9683 | -0,0997 | 0,5877 | 0,8578 | 0,6936 | 9 |
| hsa00650 | Butanoate metabolism | 1/314 | 27/9392 | 0,0370 | 1,1078 | 0,1043 | 0,6012 | 0,8639 | 0,6986 | 1 |
| hsa01040 | Biosynthesis of unsaturated fatty acids | 1/314 | 27/9392 | 0,0370 | 1,1078 | 0,1043 | 0,6012 | 0,8639 | 0,6986 | 1 |
| hsa05222 | Small cell lung cancer | 3/314 | 93/9392 | 0,0323 | 0,9649 | -0,0633 | 0,6061 | 0,8639 | 0,6986 | 3 |
| hsa04218 | Cellular senescence | 5/314 | 157/9392 | 0,0318 | 0,9526 | -0,1114 | 0,6073 | 0,8639 | 0,6986 | 5 |
| hsa04932 | Non-alcoholic fatty liver disease | 5/314 | 157/9392 | 0,0318 | 0,9526 | -0,1114 | 0,6073 | 0,8639 | 0,6986 | 5 |
| hsa00062 | Fatty acid elongation | 1/314 | 28/9392 | 0,0357 | 1,0682 | 0,0673 | 0,6146 | 0,8639 | 0,6986 | 1 |
| hsa00790 | Folate biosynthesis | 1/314 | 28/9392 | 0,0357 | 1,0682 | 0,0673 | 0,6146 | 0,8639 | 0,6986 | 1 |
| hsa05034 | Alcoholism | 6/314 | 191/9392 | 0,0314 | 0,9396 | -0,1568 | 0,6197 | 0,8639 | 0,6986 | 6 |
| hsa04392 | Hippo signaling pathway - multiple species | 1/314 | 29/9392 | 0,0345 | 1,0314 | 0,0315 | 0,6275 | 0,8639 | 0,6986 | 1 |
| hsa00140 | Steroid hormone biosynthesis | 2/314 | 63/9392 | 0,0317 | 0,9496 | -0,0747 | 0,6278 | 0,8639 | 0,6986 | 2 |
| hsa00310 | Lysine degradation | 2/314 | 63/9392 | 0,0317 | 0,9496 | -0,0747 | 0,6278 | 0,8639 | 0,6986 | 2 |
| hsa03420 | Nucleotide excision repair | 2/314 | 63/9392 | 0,0317 | 0,9496 | -0,0747 | 0,6278 | 0,8639 | 0,6986 | 2 |
| hsa03250 | Viral life cycle - HIV-1 | 2/314 | 64/9392 | 0,0313 | 0,9347 | -0,0975 | 0,6363 | 0,8684 | 0,7022 | 2 |
| hsa00020 | Citrate cycle (TCA cycle) | 1/314 | 30/9392 | 0,0333 | 0,9970 | -0,0030 | 0,6400 | 0,8684 | 0,7022 | 1 |
| hsa00563 | Glycosylphosphatidylinositol (GPI)-anchor biosynthesis | 1/314 | 30/9392 | 0,0333 | 0,9970 | -0,0030 | 0,6400 | 0,8684 | 0,7022 | 1 |
| hsa00030 | Pentose phosphate pathway | 1/314 | 31/9392 | 0,0323 | 0,9649 | -0,0364 | 0,6521 | 0,8758 | 0,7082 | 1 |
| hsa00970 | Aminoacyl-tRNA biosynthesis | 2/314 | 66/9392 | 0,0303 | 0,9064 | -0,1419 | 0,6530 | 0,8758 | 0,7082 | 2 |
| hsa04068 | FoxO signaling pathway | 4/314 | 133/9392 | 0,0301 | 0,8996 | -0,2169 | 0,6546 | 0,8758 | 0,7082 | 4 |
| hsa04820 | Cytoskeleton in muscle cells | 7/314 | 232/9392 | 0,0302 | 0,9025 | -0,2797 | 0,6627 | 0,8790 | 0,7107 | 7 |
| hsa00630 | Glyoxylate and dicarboxylate metabolism | 1/314 | 32/9392 | 0,0313 | 0,9347 | -0,0688 | 0,6638 | 0,8790 | 0,7107 | 1 |
| hsa05202 | Transcriptional misregulation in cancer | 6/314 | 201/9392 | 0,0299 | 0,8929 | -0,2856 | 0,6687 | 0,8790 | 0,7107 | 6 |
| hsa00830 | Retinol metabolism | 2/314 | 68/9392 | 0,0294 | 0,8797 | -0,1851 | 0,6691 | 0,8790 | 0,7107 | 2 |

| | | | | | | | | | | |
|---|---|---|---|---|---|---|---|---|---|---|
| hsa01210 | 2-Oxocarboxylic acid metabolism | 1/314 | 33/9392 | 0,0303 | 0,9064 | -0,1002 | 0,6751 | 0,8811 | 0,7125 | 1 |
| hsa04210 | Apoptosis | 4/314 | 137/9392 | 0,0292 | 0,8733 | -0,2778 | 0,6774 | 0,8811 | 0,7125 | 4 |
| hsa00051 | Fructose and mannose metabolism | 1/314 | 34/9392 | 0,0294 | 0,8797 | -0,1307 | 0,6860 | 0,8811 | 0,7125 | 1 |
| hsa04710 | Circadian rhythm | 1/314 | 34/9392 | 0,0294 | 0,8797 | -0,1307 | 0,6860 | 0,8811 | 0,7125 | 1 |
| hsa04137 | Mitophagy - animal | 3/314 | 105/9392 | 0,0286 | 0,8546 | -0,2787 | 0,6874 | 0,8811 | 0,7125 | 3 |
| hsa05204 | Chemical carcinogenesis - DNA adducts | 2/314 | 71/9392 | 0,0282 | 0,8426 | -0,2477 | 0,6921 | 0,8811 | 0,7125 | 2 |
| hsa05230 | Central carbon metabolism in cancer | 2/314 | 71/9392 | 0,0282 | 0,8426 | -0,2477 | 0,6921 | 0,8811 | 0,7125 | 2 |
| hsa04080 | Neuroactive ligand-receptor interaction | 11/314 | 370/9392 | 0,0297 | 0,8892 | -0,4043 | 0,6990 | 0,8811 | 0,7125 | 11 |
| hsa05014 | Amyotrophic lateral sclerosis | 11/314 | 371/9392 | 0,0296 | 0,8868 | -0,4136 | 0,7023 | 0,8811 | 0,7125 | 11 |
| hsa00982 | Drug metabolism - cytochrome P450 | 2/314 | 73/9392 | 0,0274 | 0,8195 | -0,2880 | 0,7067 | 0,8811 | 0,7125 | 2 |
| hsa05016 | Huntington disease | 9/314 | 311/9392 | 0,0289 | 0,8656 | -0,4483 | 0,7185 | 0,8811 | 0,7125 | 9 |
| hsa01230 | Biosynthesis of amino acids | 2/314 | 75/9392 | 0,0267 | 0,7976 | -0,3273 | 0,7207 | 0,8811 | 0,7125 | 2 |
| hsa01524 | Platinum drug resistance | 2/314 | 75/9392 | 0,0267 | 0,7976 | -0,3273 | 0,7207 | 0,8811 | 0,7125 | 2 |
| hsa04914 | Progesterone-mediated oocyte maturation | 3/314 | 111/9392 | 0,0270 | 0,8084 | -0,3776 | 0,7230 | 0,8811 | 0,7125 | 3 |
| hsa00760 | Nicotinate and nicotinamide metabolism | 1/314 | 38/9392 | 0,0263 | 0,7871 | -0,2445 | 0,7260 | 0,8811 | 0,7125 | 1 |
| hsa05340 | Primary immunodeficiency | 1/314 | 38/9392 | 0,0263 | 0,7871 | -0,2445 | 0,7260 | 0,8811 | 0,7125 | 1 |
| hsa03320 | PPAR signaling pathway | 2/314 | 76/9392 | 0,0263 | 0,7871 | -0,3465 | 0,7275 | 0,8811 | 0,7125 | 2 |
| hsa05168 | Herpes simplex virus 1 infection | 5/314 | 182/9392 | 0,0275 | 0,8217 | -0,4517 | 0,7334 | 0,8811 | 0,7125 | 5 |
| hsa05220 | Chronic myeloid leukemia | 2/314 | 77/9392 | 0,0260 | 0,7769 | -0,3656 | 0,7341 | 0,8811 | 0,7125 | 2 |
| hsa00670 | One carbon pool by folate | 1/314 | 39/9392 | 0,0256 | 0,7669 | -0,2712 | 0,7352 | 0,8811 | 0,7125 | 1 |
| hsa04382 | Cornified envelope formation | 6/314 | 217/9392 | 0,0276 | 0,8270 | -0,4794 | 0,7383 | 0,8811 | 0,7125 | 6 |
| hsa05132 | Salmonella infection | 7/314 | 251/9392 | 0,0279 | 0,8342 | -0,4953 | 0,7401 | 0,8811 | 0,7125 | 7 |
| hsa00500 | Starch and sucrose metabolism | 1/314 | 40/9392 | 0,0250 | 0,7478 | -0,2973 | 0,7441 | 0,8811 | 0,7125 | 1 |
| hsa00980 | Metabolism of xenobiotics by cytochrome P450 | 2/314 | 79/9392 | 0,0253 | 0,7572 | -0,4030 | 0,7470 | 0,8811 | 0,7125 | 2 |
| hsa05140 | Leishmaniasis | 2/314 | 79/9392 | 0,0253 | 0,7572 | -0,4030 | 0,7470 | 0,8811 | 0,7125 | 2 |
| hsa04612 | Antigen processing and presentation | 2/314 | 81/9392 | 0,0247 | 0,7385 | -0,4395 | 0,7593 | 0,8890 | 0,7188 | 2 |
| hsa00380 | Tryptophan metabolism | 1/314 | 42/9392 | 0,0238 | 0,7122 | -0,3477 | 0,7610 | 0,8890 | 0,7188 | 1 |
| hsa04621 | NOD-like receptor signaling pathway | 5/314 | 189/9392 | 0,0265 | 0,7913 | -0,5391 | 0,7629 | 0,8890 | 0,7188 | 5 |
| hsa04217 | Necroptosis | 4/314 | 159/9392 | 0,0252 | 0,7525 | -0,5854 | 0,7838 | 0,9066 | 0,7331 | 4 |
| hsa05332 | Graft-versus-host disease | 1/314 | 45/9392 | 0,0222 | 0,6647 | -0,4193 | 0,7843 | 0,9066 | 0,7331 | 1 |
| hsa04512 | ECM-receptor interaction | 2/314 | 89/9392 | 0,0225 | 0,6722 | -0,5779 | 0,8036 | 0,9237 | 0,7469 | 2 |
| hsa05012 | Parkinson disease | 7/314 | 271/9392 | 0,0258 | 0,7726 | -0,7064 | 0,8073 | 0,9237 | 0,7469 | 7 |
| hsa04976 | Bile secretion | 2/314 | 90/9392 | 0,0222 | 0,6647 | -0,5944 | 0,8086 | 0,9237 | 0,7469 | 2 |
| hsa05169 | Epstein-Barr virus infection | 5/314 | 204/9392 | 0,0245 | 0,7331 | -0,7167 | 0,8177 | 0,9304 | 0,7523 | 5 |
| hsa05110 | Vibrio cholerae infection | 1/314 | 51/9392 | 0,0196 | 0,5865 | -0,5507 | 0,8243 | 0,9342 | 0,7554 | 1 |
| hsa04910 | Insulin signaling pathway | 3/314 | 138/9392 | 0,0217 | 0,6502 | -0,7698 | 0,8455 | 0,9541 | 0,7715 | 3 |
| hsa05162 | Measles | 3/314 | 139/9392 | 0,0216 | 0,6456 | -0,7830 | 0,8489 | 0,9541 | 0,7715 | 3 |
| hsa04640 | Hematopoietic cell lineage | 2/314 | 100/9392 | 0,0200 | 0,5982 | -0,7512 | 0,8527 | 0,9541 | 0,7715 | 2 |
| hsa01212 | Fatty acid metabolism | 1/314 | 57/9392 | 0,0175 | 0,5248 | -0,6693 | 0,8569 | 0,9541 | 0,7715 | 1 |
| hsa05150 | Staphylococcus aureus infection | 2/314 | 102/9392 | 0,0196 | 0,5865 | -0,7809 | 0,8603 | 0,9541 | 0,7715 | 2 |
| hsa00240 | Pyrimidine metabolism | 1/314 | 58/9392 | 0,0172 | 0,5157 | -0,6880 | 0,8617 | 0,9541 | 0,7715 | 1 |
| hsa00480 | Glutathione metabolism | 1/314 | 59/9392 | 0,0169 | 0,5070 | -0,7065 | 0,8664 | 0,9556 | 0,7727 | 1 |
| hsa04974 | Protein digestion and absorption | 2/314 | 105/9392 | 0,0190 | 0,5697 | -0,8246 | 0,8711 | 0,9572 | 0,7740 | 2 |
| hsa05208 | Chemical carcinogenesis - reactive oxygen species | 5/314 | 227/9392 | 0,0220 | 0,6588 | -0,9677 | 0,8816 | 0,9650 | 0,7803 | 5 |
| hsa04350 | TGF-beta signaling pathway | 2/314 | 110/9392 | 0,0182 | 0,5438 | -0,8950 | 0,8874 | 0,9654 | 0,7806 | 2 |
| hsa05145 | Toxoplasmosis | 2/314 | 112/9392 | 0,0179 | 0,5341 | -0,9224 | 0,8933 | 0,9654 | 0,7806 | 2 |
| hsa05321 | Inflammatory bowel disease | 1/314 | 66/9392 | 0,0152 | 0,4532 | -0,8291 | 0,8948 | 0,9654 | 0,7806 | 1 |
| hsa00010 | Glycolysis / Gluconeogenesis | 1/314 | 67/9392 | 0,0149 | 0,4464 | -0,8457 | 0,8984 | 0,9654 | 0,7806 | 1 |
| hsa05171 | Coronavirus disease - COVID-19 | 5/314 | 238/9392 | 0,0210 | 0,6284 | -1,0800 | 0,9047 | 0,9654 | 0,7806 | 5 |
| hsa01200 | Carbon metabolism | 2/314 | 117/9392 | 0,0171 | 0,5113 | -0,9893 | 0,9069 | 0,9654 | 0,7806 | 2 |
| hsa04920 | Adipocytokine signaling pathway | 1/314 | 70/9392 | 0,0143 | 0,4273 | -0,8944 | 0,9083 | 0,9654 | 0,7806 | 1 |
| hsa05120 | Epithelial cell signaling in Helicobacter pylori infection | 1/314 | 71/9392 | 0,0141 | 0,4213 | -0,9103 | 0,9114 | 0,9654 | 0,7806 | 1 |
| hsa04668 | TNF signaling pathway | 2/314 | 119/9392 | 0,0168 | 0,5027 | -1,0153 | 0,9119 | 0,9654 | 0,7806 | 2 |
| hsa04140 | Autophagy - animal | 3/314 | 169/9392 | 0,0178 | 0,5310 | -1,1443 | 0,9257 | 0,9706 | 0,7848 | 3 |
| hsa04141 | Protein processing in endoplasmic reticulum | 3/314 | 171/9392 | 0,0175 | 0,5248 | -1,1664 | 0,9293 | 0,9706 | 0,7848 | 3 |
| hsa03018 | RNA degradation | 1/314 | 78/9392 | 0,0128 | 0,3835 | -1,0169 | 0,9303 | 0,9706 | 0,7848 | 1 |
| hsa05133 | Pertussis | 1/314 | 78/9392 | 0,0128 | 0,3835 | -1,0169 | 0,9303 | 0,9706 | 0,7848 | 1 |
| hsa00983 | Drug metabolism - other enzymes | 1/314 | 81/9392 | 0,0123 | 0,3693 | -1,0603 | 0,9371 | 0,9742 | 0,7877 | 1 |
| hsa04146 | Peroxisome | 1/314 | 83/9392 | 0,0120 | 0,3604 | -1,0885 | 0,9413 | 0,9750 | 0,7884 | 1 |
| hsa04714 | Thermogenesis | 4/314 | 235/9392 | 0,0170 | 0,5091 | -1,4173 | 0,9577 | 0,9885 | 0,7993 | 4 |
| hsa05323 | Rheumatoid arthritis | 1/314 | 95/9392 | 0,0105 | 0,3149 | -1,2482 | 0,9611 | 0,9885 | 0,7993 | 1 |
| hsa04145 | Phagosome | 2/314 | 159/9392 | 0,0126 | 0,3762 | -1,4753 | 0,9718 | 0,9959 | 0,8053 | 2 |
| hsa00190 | Oxidative phosphorylation | 1/314 | 138/9392 | 0,0072 | 0,2167 | -1,7239 | 0,9912 | 0,9999 | 0,8085 | 1 |
| hsa04936 | Alcoholic liver disease | 1/314 | 144/9392 | 0,0069 | 0,2077 | -1,7818 | 0,9928 | 0,9999 | 0,8085 | 1 |
| hsa05322 | Systemic lupus erythematosus | 1/314 | 144/9392 | 0,0069 | 0,2077 | -1,7818 | 0,9928 | 0,9999 | 0,8085 | 1 |
| hsa05206 | MicroRNAs in cancer | 4/314 | 314/9392 | 0,0127 | 0,3810 | -2,0747 | 0,9941 | 0,9999 | 0,8085 | 4 |
| hsa03040 | Spliceosome | 2/314 | 235/9392 | 0,0085 | 0,2546 | -2,1523 | 0,9971 | 0,9999 | 0,8085 | 2 |
| hsa04060 | Cytokine-cytokine receptor interaction | 3/314 | 298/9392 | 0,0101 | 0,3011 | -2,2801 | 0,9977 | 0,9999 | 0,8085 | 3 |
| hsa04740 | Olfactory transduction | 4/314 | 453/9392 | 0,0088 | 0,2641 | -2,9857 | 0,9999 | 0,9999 | 0,8085 | 4 |

**Table S14. Pathway enrichment analysis of genes harboring rare shared variants in concordant and discordant NDC twin pairs, excluding genes with rare variants shared in concordant TD twin pairs.** Analysis was performed using the R package ClusterProfiler and p-values were adjusted by the FDR method.

| | Description | GeneRatio | BgRatio | RichFactor | FoldEnrichment | zScore | pvalue | p.adjust | qvalue | Count |
|---|---|---|---|---|---|---|---|---|---|---|
| hsa04020 | Calcium signaling pathway | 20/276 | 254/9436 | 0,0787 | 2,6920 | 4,7449 | 0,0001 | 0,0125 | 0,0106 | 20 |
| hsa00310 | Lysine degradation | 9/276 | 63/9436 | 0,1429 | 4,8841 | 5,3690 | 0,0001 | 0,0125 | 0,0106 | 9 |
| hsa05211 | Renal cell carcinoma | 9/276 | 70/9436 | 0,1286 | 4,3957 | 4,9496 | 0,0002 | 0,0191 | 0,0163 | 9 |
| hsa04713 | Circadian entrainment | 10/276 | 97/9436 | 0,1031 | 3,5246 | 4,3381 | 0,0005 | 0,0361 | 0,0308 | 10 |
| hsa05410 | Hypertrophic cardiomyopathy | 10/276 | 99/9436 | 0,1010 | 3,4534 | 4,2595 | 0,0006 | 0,0361 | 0,0308 | 10 |
| hsa04720 | Long-term potentiation | 8/276 | 67/9436 | 0,1194 | 4,0822 | 4,3947 | 0,0007 | 0,0361 | 0,0308 | 8 |
| hsa05412 | Arrhythmogenic right ventricular cardiomyopathy | 9/276 | 86/9436 | 0,1047 | 3,5779 | 4,1685 | 0,0009 | 0,0372 | 0,0317 | 9 |
| hsa05414 | Dilated cardiomyopathy | 10/276 | 105/9436 | 0,0952 | 3,2560 | 4,0351 | 0,0010 | 0,0372 | 0,0317 | 10 |
| hsa04820 | Cytoskeleton in muscle cells | 16/276 | 233/9436 | 0,0687 | 2,3477 | 3,6156 | 0,0013 | 0,0452 | 0,0386 | 16 |
| hsa04110 | Cell cycle | 12/276 | 158/9436 | 0,0759 | 2,5966 | 3,5129 | 0,0023 | 0,0699 | 0,0596 | 12 |
| hsa00340 | Histidine metabolism | 4/276 | 22/9436 | 0,1818 | 6,2161 | 4,2515 | 0,0035 | 0,0961 | 0,0820 | 4 |
| hsa05165 | Human papillomavirus infection | 19/276 | 333/9436 | 0,0571 | 1,9507 | 3,0658 | 0,0041 | 0,0981 | 0,0836 | 19 |
| hsa05225 | Hepatocellular carcinoma | 12/276 | 170/9436 | 0,0706 | 2,4133 | 3,2277 | 0,0042 | 0,0981 | 0,0836 | 12 |
| hsa00510 | N-Glycan biosynthesis | 6/276 | 55/9436 | 0,1091 | 3,7296 | 3,5240 | 0,0052 | 0,1129 | 0,0963 | 6 |
| hsa04068 | FoxO signaling pathway | 10/276 | 133/9436 | 0,0752 | 2,5706 | 3,1663 | 0,0056 | 0,1136 | 0,0969 | 10 |
| hsa05205 | Proteoglycans in cancer | 13/276 | 204/9436 | 0,0637 | 2,1787 | 2,9542 | 0,0069 | 0,1315 | 0,1122 | 13 |
| hsa04722 | Neurotrophin signaling pathway | 9/276 | 120/9436 | 0,0750 | 2,5641 | 2,9931 | 0,0085 | 0,1370 | 0,1168 | 9 |
| hsa04330 | Notch signaling pathway | 6/276 | 62/9436 | 0,0968 | 3,3086 | 3,1656 | 0,0093 | 0,1370 | 0,1168 | 6 |
| hsa04916 | Melanogenesis | 8/276 | 101/9436 | 0,0792 | 2,7080 | 2,9955 | 0,0094 | 0,1370 | 0,1168 | 8 |
| hsa04919 | Thyroid hormone signaling pathway | 9/276 | 122/9436 | 0,0738 | 2,5221 | 2,9372 | 0,0094 | 0,1370 | 0,1168 | 9 |
| hsa02010 | ABC transporters | 5/276 | 45/9436 | 0,1111 | 3,7987 | 3,2665 | 0,0097 | 0,1370 | 0,1168 | 5 |
| hsa04151 | PI3K-Akt signaling pathway | 19/276 | 362/9436 | 0,0525 | 1,7944 | 2,6754 | 0,0098 | 0,1370 | 0,1168 | 19 |
| hsa00563 | Glycosylphosphatidylinositol (GPI)-anchor biosynthesis | 4/276 | 30/9436 | 0,1333 | 4,5585 | 3,3884 | 0,0108 | 0,1415 | 0,1207 | 4 |
| hsa05224 | Breast cancer | 10/276 | 148/9436 | 0,0676 | 2,3100 | 2,7882 | 0,0115 | 0,1415 | 0,1207 | 10 |
| hsa04929 | GnRH secretion | 6/276 | 65/9436 | 0,0923 | 3,1559 | 3,0273 | 0,0116 | 0,1415 | 0,1207 | 6 |
| hsa00410 | beta-Alanine metabolism | 4/276 | 31/9436 | 0,1290 | 4,4114 | 3,3023 | 0,0121 | 0,1424 | 0,1215 | 4 |
| hsa04310 | Wnt signaling pathway | 11/276 | 174/9436 | 0,0632 | 2,1613 | 2,6838 | 0,0131 | 0,1490 | 0,1271 | 11 |
| hsa04010 | MAPK signaling pathway | 16/276 | 300/9436 | 0,0533 | 1,8234 | 2,5157 | 0,0149 | 0,1569 | 0,1338 | 16 |
| hsa05031 | Amphetamine addiction | 6/276 | 69/9436 | 0,0870 | 2,9729 | 2,8550 | 0,0152 | 0,1569 | 0,1338 | 6 |
| hsa04024 | cAMP signaling pathway | 13/276 | 226/9436 | 0,0575 | 1,9666 | 2,5530 | 0,0154 | 0,1569 | 0,1338 | 13 |
| hsa04390 | Hippo signaling pathway | 10/276 | 157/9436 | 0,0637 | 2,1776 | 2,5827 | 0,0168 | 0,1660 | 0,1416 | 10 |
| hsa05230 | Central carbon metabolism in cancer | 6/276 | 71/9436 | 0,0845 | 2,8892 | 2,7735 | 0,0174 | 0,1660 | 0,1416 | 6 |
| hsa04520 | Adherens junction | 7/276 | 93/9436 | 0,0753 | 2,5733 | 2,6466 | 0,0190 | 0,1757 | 0,1499 | 7 |
| hsa00785 | Lipoic acid metabolism | 3/276 | 20/9436 | 0,1500 | 5,1283 | 3,2079 | 0,0195 | 0,1757 | 0,1499 | 3 |
| hsa03082 | ATP-dependent chromatin remodeling | 8/276 | 117/9436 | 0,0684 | 2,3377 | 2,5272 | 0,0212 | 0,1853 | 0,1581 | 8 |
| hsa04015 | Rap1 signaling pathway | 12/276 | 212/9436 | 0,0566 | 1,9352 | 2,3905 | 0,0218 | 0,1853 | 0,1581 | 12 |
| hsa01522 | Endocrine resistance | 7/276 | 99/9436 | 0,0707 | 2,4174 | 2,4608 | 0,0258 | 0,2074 | 0,1769 | 7 |
| hsa05231 | Choline metabolism in cancer | 7/276 | 99/9436 | 0,0707 | 2,4174 | 2,4608 | 0,0258 | 0,2074 | 0,1769 | 7 |
| hsa04933 | AGE-RAGE signaling pathway in diabetic complications | 7/276 | 101/9436 | 0,0693 | 2,3695 | 2,4018 | 0,0283 | 0,2224 | 0,1897 | 7 |
| hsa00260 | Glycine, serine and threonine metabolism | 4/276 | 41/9436 | 0,0976 | 3,3355 | 2,6013 | 0,0311 | 0,2362 | 0,2015 | 4 |
| hsa05166 | Human T-cell leukemia virus 1 infection | 12/276 | 224/9436 | 0,0536 | 1,8315 | 2,1862 | 0,0316 | 0,2362 | 0,2015 | 12 |
| hsa00380 | Tryptophan metabolism | 4/276 | 42/9436 | 0,0952 | 3,2560 | 2,5435 | 0,0336 | 0,2423 | 0,2067 | 4 |
| hsa04974 | Protein digestion and absorption | 7/276 | 105/9436 | 0,0667 | 2,2792 | 2,2880 | 0,0340 | 0,2423 | 0,2067 | 7 |
| hsa05215 | Prostate cancer | 7/276 | 106/9436 | 0,0660 | 2,2577 | 2,2603 | 0,0356 | 0,2424 | 0,2068 | 7 |
| hsa00513 | Various types of N-glycan biosynthesis | 4/276 | 43/9436 | 0,0930 | 3,1803 | 2,4873 | 0,0362 | 0,2424 | 0,2068 | 4 |
| hsa04921 | Oxytocin signaling pathway | 9/276 | 155/9436 | 0,0581 | 1,9851 | 2,1466 | 0,0381 | 0,2424 | 0,2068 | 9 |
| hsa04934 | Cushing syndrome | 9/276 | 155/9436 | 0,0581 | 1,9851 | 2,1466 | 0,0381 | 0,2424 | 0,2068 | 9 |
| hsa05415 | Diabetic cardiomyopathy | 11/276 | 205/9436 | 0,0537 | 1,8345 | 2,0968 | 0,0383 | 0,2424 | 0,2068 | 11 |
| hsa03022 | Basal transcription factors | 4/276 | 44/9436 | 0,0909 | 3,1080 | 2,4328 | 0,0389 | 0,2424 | 0,2068 | 4 |
| hsa04911 | Insulin secretion | 6/276 | 86/9436 | 0,0698 | 2,3852 | 2,2400 | 0,0398 | 0,2424 | 0,2068 | 6 |
| hsa04927 | Cortisol synthesis and secretion | 5/276 | 65/9436 | 0,0769 | 2,6299 | 2,2887 | 0,0411 | 0,2424 | 0,2068 | 5 |
| hsa04142 | Lysosome | 8/276 | 133/9436 | 0,0602 | 2,0564 | 2,1298 | 0,0412 | 0,2424 | 0,2068 | 8 |
| hsa04150 | mTOR signaling pathway | 9/276 | 158/9436 | 0,0570 | 1,9474 | 2,0846 | 0,0422 | 0,2437 | 0,2079 | 9 |
| hsa04512 | ECM-receptor interaction | 6/276 | 89/9436 | 0,0674 | 2,3048 | 2,1468 | 0,0458 | 0,2596 | 0,2215 | 6 |
| hsa04014 | Ras signaling pathway | 12/276 | 238/9436 | 0,0504 | 1,7238 | 1,9630 | 0,0468 | 0,2596 | 0,2215 | 12 |
| hsa04930 | Type II diabetes mellitus | 4/276 | 47/9436 | 0,0851 | 2,9097 | 2,2781 | 0,0478 | 0,2596 | 0,2215 | 4 |
| hsa05221 | Acute myeloid leukemia | 5/276 | 68/9436 | 0,0735 | 2,5139 | 2,1747 | 0,0484 | 0,2596 | 0,2215 | 5 |
| hsa05161 | Hepatitis B | 9/276 | 163/9436 | 0,0552 | 1,8877 | 1,9844 | 0,0497 | 0,2624 | 0,2239 | 9 |
| hsa04371 | Apelin signaling pathway | 8/276 | 140/9436 | 0,0571 | 1,9536 | 1,9732 | 0,0529 | 0,2727 | 0,2327 | 8 |
| hsa04724 | Glutamatergic synapse | 7/276 | 116/9436 | 0,0603 | 2,0631 | 1,9997 | 0,0535 | 0,2727 | 0,2327 | 7 |
| hsa00330 | Arginine and proline metabolism | 4/276 | 50/9436 | 0,0800 | 2,7351 | 2,1352 | 0,0578 | 0,2898 | 0,2472 | 4 |
| hsa04140 | Autophagy - animal | 9/276 | 169/9436 | 0,0533 | 1,8207 | 1,8686 | 0,0599 | 0,2955 | 0,2521 | 9 |
| hsa04530 | Tight junction | 9/276 | 170/9436 | 0,0529 | 1,8100 | 1,8498 | 0,0617 | 0,2969 | 0,2533 | 9 |
| hsa00562 | Inositol phosphate metabolism | 5/276 | 73/9436 | 0,0685 | 2,3417 | 1,9975 | 0,0621 | 0,2969 | 0,2533 | 5 |
| hsa04814 | Motor proteins | 10/276 | 197/9436 | 0,0508 | 1,7355 | 1,8107 | 0,0636 | 0,2993 | 0,2553 | 10 |
| hsa04925 | Aldosterone synthesis and secretion | 6/276 | 98/9436 | 0,0612 | 2,0932 | 1,8882 | 0,0671 | 0,3113 | 0,2655 | 6 |
| hsa04115 | p53 signaling pathway | 5/276 | 75/9436 | 0,0667 | 2,2792 | 1,9306 | 0,0682 | 0,3113 | 0,2655 | 5 |

| | | | | | | | | | | |
|---|---|---|---|---|---|---|---|---|---|---|
| hsa04072 | Phospholipase D signaling pathway | 8/276 | 149/9436 | 0,0537 | 1,8356 | 1,7846 | 0,0708 | 0,3187 | 0,2719 | 8 |
| hsa05226 | Gastric cancer | 8/276 | 150/9436 | 0,0533 | 1,8234 | 1,7645 | 0,0730 | 0,3238 | 0,2762 | 8 |
| hsa04510 | Focal adhesion | 10/276 | 203/9436 | 0,0493 | 1,6842 | 1,7105 | 0,0745 | 0,3255 | 0,2777 | 10 |
| hsa00603 | Glycosphingolipid biosynthesis - globo and isoglobo series | 2/276 | 16/9436 | 0,1250 | 4,2736 | 2,2747 | 0,0781 | 0,3332 | 0,2842 | 2 |
| hsa04810 | Regulation of actin cytoskeleton | 11/276 | 232/9436 | 0,0474 | 1,6210 | 1,6624 | 0,0784 | 0,3332 | 0,2842 | 11 |
| hsa01240 | Biosynthesis of cofactors | 8/276 | 154/9436 | 0,0519 | 1,7760 | 1,6854 | 0,0822 | 0,3446 | 0,2940 | 8 |
| hsa01521 | EGFR tyrosine kinase inhibitor resistance | 5/276 | 80/9436 | 0,0625 | 2,1368 | 1,7724 | 0,0847 | 0,3499 | 0,2985 | 5 |
| hsa00450 | Selenocompound metabolism | 2/276 | 17/9436 | 0,1176 | 4,0222 | 2,1648 | 0,0869 | 0,3499 | 0,2985 | 2 |
| hsa03273 | Virion - Lassa virus and SFTS virus | 2/276 | 17/9436 | 0,1176 | 4,0222 | 2,1648 | 0,0869 | 0,3499 | 0,2985 | 2 |
| hsa04148 | Efferocytosis | 8/276 | 157/9436 | 0,0510 | 1,7421 | 1,6275 | 0,0895 | 0,3512 | 0,2996 | 8 |
| hsa04218 | Cellular senescence | 8/276 | 157/9436 | 0,0510 | 1,7421 | 1,6275 | 0,0895 | 0,3512 | 0,2996 | 8 |
| hsa04728 | Dopaminergic synapse | 7/276 | 132/9436 | 0,0530 | 1,8130 | 1,6328 | 0,0921 | 0,3569 | 0,3045 | 7 |
| hsa04922 | Glucagon signaling pathway | 6/276 | 107/9436 | 0,0561 | 1,9171 | 1,6560 | 0,0933 | 0,3570 | 0,3046 | 6 |
| hsa03083 | Polycomb repressive complex | 5/276 | 83/9436 | 0,0602 | 2,0595 | 1,6829 | 0,0955 | 0,3607 | 0,3077 | 5 |
| hsa04066 | HIF-1 signaling pathway | 6/276 | 110/9436 | 0,0545 | 1,8648 | 1,5836 | 0,1031 | 0,3789 | 0,3232 | 6 |
| hsa04350 | TGF-beta signaling pathway | 6/276 | 110/9436 | 0,0545 | 1,8648 | 1,5836 | 0,1031 | 0,3789 | 0,3232 | 6 |
| hsa00670 | One carbon pool by folate | 3/276 | 39/9436 | 0,0769 | 2,6299 | 1,7704 | 0,1049 | 0,3789 | 0,3232 | 3 |
| hsa00531 | Glycosaminoglycan degradation | 2/276 | 19/9436 | 0,1053 | 3,5988 | 1,9682 | 0,1052 | 0,3789 | 0,3232 | 2 |
| hsa00500 | Starch and sucrose metabolism | 3/276 | 40/9436 | 0,0750 | 2,5641 | 1,7207 | 0,1110 | 0,3951 | 0,3370 | 3 |
| hsa05022 | Pathways of neurodegeneration - multiple diseases | 19/276 | 483/9436 | 0,0393 | 1,3449 | 1,3506 | 0,1154 | 0,4058 | 0,3462 | 19 |
| hsa03250 | Viral life cycle - HIV-1 | 4/276 | 64/9436 | 0,0625 | 2,1368 | 1,5839 | 0,1170 | 0,4070 | 0,3472 | 4 |
| hsa00561 | Glycerolipid metabolism | 4/276 | 65/9436 | 0,0615 | 2,1039 | 1,5501 | 0,1220 | 0,4141 | 0,3533 | 4 |
| hsa04120 | Ubiquitin mediated proteolysis | 7/276 | 142/9436 | 0,0493 | 1,6853 | 1,4283 | 0,1226 | 0,4141 | 0,3533 | 7 |
| hsa04725 | Cholinergic synapse | 6/276 | 116/9436 | 0,0517 | 1,7684 | 1,4453 | 0,1243 | 0,4141 | 0,3533 | 6 |
| hsa00770 | Pantothenate and CoA biosynthesis | 2/276 | 21/9436 | 0,0952 | 3,2560 | 1,7965 | 0,1245 | 0,4141 | 0,3533 | 2 |
| hsa00071 | Fatty acid degradation | 3/276 | 43/9436 | 0,0698 | 2,3852 | 1,5803 | 0,1304 | 0,4289 | 0,3659 | 3 |
| hsa05202 | Transcriptional misregulation in cancer | 9/276 | 201/9436 | 0,0448 | 1,5308 | 1,3204 | 0,1351 | 0,4389 | 0,3744 | 9 |
| hsa04912 | GnRH signaling pathway | 5/276 | 93/9436 | 0,0538 | 1,8381 | 1,4098 | 0,1363 | 0,4389 | 0,3744 | 5 |
| hsa00515 | Mannose type O-glycan biosynthesis | 2/276 | 23/9436 | 0,0870 | 2,9729 | 1,6443 | 0,1445 | 0,4536 | 0,3869 | 2 |
| hsa04964 | Proximal tubule bicarbonate reclamation | 2/276 | 23/9436 | 0,0870 | 2,9729 | 1,6443 | 0,1445 | 0,4536 | 0,3869 | 2 |
| hsa04152 | AMPK signaling pathway | 6/276 | 122/9436 | 0,0492 | 1,6814 | 1,3149 | 0,1474 | 0,4536 | 0,3869 | 6 |
| hsa04935 | Growth hormone synthesis, secretion and action | 6/276 | 122/9436 | 0,0492 | 1,6814 | 1,3149 | 0,1474 | 0,4536 | 0,3869 | 6 |
| hsa05416 | Viral myocarditis | 4/276 | 70/9436 | 0,0571 | 1,9536 | 1,3900 | 0,1482 | 0,4536 | 0,3869 | 4 |
| hsa00620 | Pyruvate metabolism | 3/276 | 47/9436 | 0,0638 | 2,1822 | 1,4103 | 0,1578 | 0,4774 | 0,4072 | 3 |
| hsa04070 | Phosphatidylinositol signaling system | 5/276 | 98/9436 | 0,0510 | 1,7443 | 1,2856 | 0,1591 | 0,4774 | 0,4072 | 5 |
| hsa05218 | Melanoma | 4/276 | 73/9436 | 0,0548 | 1,8733 | 1,3002 | 0,1649 | 0,4856 | 0,4142 | 4 |
| hsa04261 | Adrenergic signaling in cardiomyocytes | 7/276 | 154/9436 | 0,0455 | 1,5540 | 1,2032 | 0,1650 | 0,4856 | 0,4142 | 7 |
| hsa04360 | Axon guidance | 8/276 | 184/9436 | 0,0435 | 1,4865 | 1,1567 | 0,1713 | 0,4968 | 0,4239 | 8 |
| hsa05030 | Cocaine addiction | 3/276 | 49/9436 | 0,0612 | 2,0932 | 1,3317 | 0,1721 | 0,4968 | 0,4239 | 3 |
| hsa04977 | Vitamin digestion and absorption | 2/276 | 26/9436 | 0,0769 | 2,6299 | 1,4445 | 0,1756 | 0,5023 | 0,4285 | 2 |
| hsa05144 | Malaria | 3/276 | 50/9436 | 0,0600 | 2,0513 | 1,2938 | 0,1794 | 0,5057 | 0,4314 | 3 |
| hsa05214 | Glioma | 4/276 | 76/9436 | 0,0526 | 1,7994 | 1,2145 | 0,1822 | 0,5057 | 0,4314 | 4 |
| hsa05142 | Chagas disease | 5/276 | 103/9436 | 0,0485 | 1,6596 | 1,1684 | 0,1834 | 0,5057 | 0,4314 | 5 |
| hsa05146 | Amoebiasis | 5/276 | 103/9436 | 0,0485 | 1,6596 | 1,1684 | 0,1834 | 0,5057 | 0,4314 | 5 |
| hsa05220 | Chronic myeloid leukemia | 4/276 | 77/9436 | 0,0519 | 1,7760 | 1,1868 | 0,1881 | 0,5139 | 0,4385 | 4 |
| hsa05100 | Bacterial invasion of epithelial cells | 4/276 | 78/9436 | 0,0513 | 1,7533 | 1,1595 | 0,1941 | 0,5255 | 0,4483 | 4 |
| hsa00062 | Fatty acid elongation | 2/276 | 28/9436 | 0,0714 | 2,4420 | 1,3264 | 0,1969 | 0,5285 | 0,4509 | 2 |
| hsa04972 | Pancreatic secretion | 5/276 | 106/9436 | 0,0472 | 1,6127 | 1,1011 | 0,1986 | 0,5285 | 0,4509 | 5 |
| hsa05016 | Huntington disease | 12/276 | 311/9436 | 0,0386 | 1,3192 | 0,9935 | 0,2004 | 0,5286 | 0,4510 | 12 |
| hsa03013 | Nucleocytoplasmic transport | 5/276 | 108/9436 | 0,0463 | 1,5828 | 1,0573 | 0,2090 | 0,5466 | 0,4663 | 5 |
| hsa04931 | Insulin resistance | 5/276 | 109/9436 | 0,0459 | 1,5683 | 1,0358 | 0,2142 | 0,5556 | 0,4740 | 5 |
| hsa03460 | Fanconi anemia pathway | 3/276 | 55/9436 | 0,0545 | 1,8648 | 1,1165 | 0,2170 | 0,5572 | 0,4753 | 3 |
| hsa00053 | Ascorbate and aldarate metabolism | 2/276 | 30/9436 | 0,0667 | 2,2792 | 1,2181 | 0,2185 | 0,5572 | 0,4753 | 2 |
| hsa01212 | Fatty acid metabolism | 3/276 | 57/9436 | 0,0526 | 1,7994 | 1,0507 | 0,2325 | 0,5881 | 0,5017 | 3 |
| hsa00630 | Glyoxylate and dicarboxylate metabolism | 2/276 | 32/9436 | 0,0625 | 2,1368 | 1,1181 | 0,2403 | 0,5980 | 0,5101 | 2 |
| hsa00240 | Pyrimidine metabolism | 3/276 | 58/9436 | 0,0517 | 1,7684 | 1,0188 | 0,2404 | 0,5980 | 0,5101 | 3 |
| hsa04550 | Signaling pathways regulating pluripotency of stem cells | 6/276 | 144/9436 | 0,0417 | 1,4245 | 0,8910 | 0,2458 | 0,6020 | 0,5136 | 6 |
| hsa04260 | Cardiac muscle contraction | 4/276 | 87/9436 | 0,0460 | 1,5719 | 0,9302 | 0,2500 | 0,6020 | 0,5136 | 4 |
| hsa05210 | Colorectal cancer | 4/276 | 87/9436 | 0,0460 | 1,5719 | 0,9302 | 0,2500 | 0,6020 | 0,5136 | 4 |
| hsa01210 | 2-Oxocarboxylic acid metabolism | 2/276 | 33/9436 | 0,0606 | 2,0720 | 1,0708 | 0,2512 | 0,6020 | 0,5136 | 2 |
| hsa05203 | Viral carcinogenesis | 8/276 | 205/9436 | 0,0390 | 1,3342 | 0,8397 | 0,2518 | 0,6020 | 0,5136 | 8 |
| hsa04730 | Long-term depression | 3/276 | 60/9436 | 0,0500 | 1,7094 | 0,9569 | 0,2561 | 0,6048 | 0,5160 | 3 |
| hsa00920 | Sulfur metabolism | 1/276 | Oct-36 | 0,1000 | 3,4188 | 1,3284 | 0,2570 | 0,6048 | 0,5160 | 1 |
| hsa04723 | Retrograde endocannabinoid signaling | 6/276 | 149/9436 | 0,0403 | 1,3767 | 0,8045 | 0,2705 | 0,6319 | 0,5391 | 6 |
| hsa05217 | Basal cell carcinoma | 3/276 | 63/9436 | 0,0476 | 1,6280 | 0,8681 | 0,2800 | 0,6457 | 0,5508 | 3 |

| | | | | | | | | | | |
|---|---|---|---|---|---|---|---|---|---|---|
| hsa04540 | Gap junction | 4/276 | 92/9436 | 0,0435 | 1,4865 | 0,8139 | 0,2825 | 0,6457 | 0,5508 | 4 |
| hsa00350 | Tyrosine metabolism | 2/276 | 36/9436 | 0,0556 | 1,8994 | 0,9384 | 0,2840 | 0,6457 | 0,5508 | 2 |
| hsa04071 | Sphingolipid signaling pathway | 5/276 | 122/9436 | 0,0410 | 1,4012 | 0,7741 | 0,2857 | 0,6457 | 0,5508 | 5 |
| hsa04658 | Th1 and Th2 cell differentiation | 4/276 | 93/9436 | 0,0430 | 1,4705 | 0,7914 | 0,2891 | 0,6457 | 0,5508 | 4 |
| hsa05222 | Small cell lung cancer | 4/276 | 93/9436 | 0,0430 | 1,4705 | 0,7914 | 0,2891 | 0,6457 | 0,5508 | 4 |
| hsa00130 | Ubiquinone and other terpenoid-quinone biosynthesis | 1/276 | Dec-36 | 0,0833 | 2,8490 | 1,1125 | 0,2998 | 0,6601 | 0,5631 | 1 |
| hsa03265 | Virion - Ebolavirus, Lyssavirus and Morbillivirus | 1/276 | Dec-36 | 0,0833 | 2,8490 | 1,1125 | 0,2998 | 0,6601 | 0,5631 | 1 |
| hsa00970 | Aminoacyl-tRNA biosynthesis | 3/276 | 66/9436 | 0,0455 | 1,5540 | 0,7840 | 0,3041 | 0,6647 | 0,5671 | 3 |
| hsa04144 | Endocytosis | 9/276 | 252/9436 | 0,0357 | 1,2210 | 0,6173 | 0,3179 | 0,6898 | 0,5885 | 9 |
| hsa03450 | Non-homologous end-joining | 1/276 | 13/9436 | 0,0769 | 2,6299 | 1,0207 | 0,3203 | 0,6903 | 0,5889 | 1 |
| hsa04924 | Renin secretion | 3/276 | 69/9436 | 0,0435 | 1,4865 | 0,7040 | 0,3283 | 0,6989 | 0,5962 | 3 |
| hsa04666 | Fc gamma R-mediated phagocytosis | 4/276 | 99/9436 | 0,0404 | 1,3813 | 0,6621 | 0,3289 | 0,6989 | 0,5962 | 4 |
| hsa05033 | Nicotine addiction | 2/276 | 41/9436 | 0,0488 | 1,6677 | 0,7437 | 0,3383 | 0,7131 | 0,6084 | 2 |
| hsa00533 | Glycosaminoglycan biosynthesis - keratan sulfate | 1/276 | 14/9436 | 0,0714 | 2,4420 | 0,9372 | 0,3403 | 0,7131 | 0,6084 | 1 |
| hsa05120 | Epithelial cell signaling in Helicobacter pylori infection | 3/276 | 71/9436 | 0,0423 | 1,4446 | 0,6527 | 0,3444 | 0,7170 | 0,6116 | 3 |
| hsa00564 | Glycerophospholipid metabolism | 4/276 | 103/9436 | 0,0388 | 1,3277 | 0,5805 | 0,3556 | 0,7306 | 0,6233 | 4 |
| hsa00604 | Glycosphingolipid biosynthesis - ganglio series | 1/276 | 15/9436 | 0,0667 | 2,2792 | 0,8606 | 0,3596 | 0,7306 | 0,6233 | 1 |
| hsa00730 | Thiamine metabolism | 1/276 | 15/9436 | 0,0667 | 2,2792 | 0,8606 | 0,3596 | 0,7306 | 0,6233 | 1 |
| hsa05223 | Non-small cell lung cancer | 3/276 | 73/9436 | 0,0411 | 1,4050 | 0,6030 | 0,3605 | 0,7306 | 0,6233 | 3 |
| hsa04082 | Neuroactive ligand signaling | 7/276 | 199/9436 | 0,0352 | 1,2026 | 0,5014 | 0,3641 | 0,7328 | 0,6252 | 7 |
| hsa03410 | Base excision repair | 2/276 | 44/9436 | 0,0455 | 1,5540 | 0,6394 | 0,3703 | 0,7328 | 0,6252 | 2 |
| hsa04210 | Apoptosis | 5/276 | 137/9436 | 0,0365 | 1,2478 | 0,5070 | 0,3725 | 0,7328 | 0,6252 | 5 |
| hsa04918 | Thyroid hormone synthesis | 3/276 | 75/9436 | 0,0400 | 1,3675 | 0,5547 | 0,3765 | 0,7328 | 0,6252 | 3 |
| hsa00360 | Phenylalanine metabolism | 1/276 | 16/9436 | 0,0625 | 2,1368 | 0,7899 | 0,3783 | 0,7328 | 0,6252 | 1 |
| hsa04910 | Insulin signaling pathway | 5/276 | 138/9436 | 0,0362 | 1,2387 | 0,4903 | 0,3784 | 0,7328 | 0,6252 | 5 |
| hsa05135 | Yersinia infection | 5/276 | 138/9436 | 0,0362 | 1,2387 | 0,4903 | 0,3784 | 0,7328 | 0,6252 | 5 |
| hsa04714 | Thermogenesis | 8/276 | 235/9436 | 0,0340 | 1,1639 | 0,4415 | 0,3819 | 0,7349 | 0,6270 | 8 |
| hsa04971 | Gastric acid secretion | 3/276 | 76/9436 | 0,0395 | 1,3495 | 0,5311 | 0,3845 | 0,7354 | 0,6274 | 3 |
| hsa05212 | Pancreatic cancer | 3/276 | 77/9436 | 0,0390 | 1,3320 | 0,5078 | 0,3925 | 0,7460 | 0,6364 | 3 |
| hsa00910 | Nitrogen metabolism | 1/276 | 17/9436 | 0,0588 | 2,0111 | 0,7242 | 0,3966 | 0,7490 | 0,6390 | 1 |
| hsa00514 | Other types of O-glycan biosynthesis | 2/276 | 47/9436 | 0,0426 | 1,4548 | 0,5426 | 0,4016 | 0,7506 | 0,6404 | 2 |
| hsa05014 | Amyotrophic lateral sclerosis | 12/276 | 371/9436 | 0,0323 | 1,1058 | 0,3610 | 0,4023 | 0,7506 | 0,6404 | 12 |
| hsa04914 | Progesterone-mediated oocyte maturation | 4/276 | 111/9436 | 0,0360 | 1,2320 | 0,4268 | 0,4090 | 0,7525 | 0,6419 | 4 |
| hsa00280 | Valine, leucine and isoleucine degradation | 2/276 | 48/9436 | 0,0417 | 1,4245 | 0,5118 | 0,4119 | 0,7525 | 0,6419 | 2 |
| hsa05017 | Spinocerebellar ataxia | 5/276 | 144/9436 | 0,0347 | 1,1871 | 0,3927 | 0,4134 | 0,7525 | 0,6419 | 5 |
| hsa00511 | Other glycan degradation | 1/276 | 18/9436 | 0,0556 | 1,8994 | 0,6629 | 0,4142 | 0,7525 | 0,6419 | 1 |
| hsa05145 | Toxoplasmosis | 4/276 | 112/9436 | 0,0357 | 1,2210 | 0,4084 | 0,4156 | 0,7525 | 0,6419 | 4 |
| hsa05020 | Prion disease | 9/276 | 278/9436 | 0,0324 | 1,1068 | 0,3138 | 0,4263 | 0,7673 | 0,6546 | 9 |
| hsa04726 | Serotonergic synapse | 4/276 | 115/9436 | 0,0348 | 1,1892 | 0,3543 | 0,4353 | 0,7790 | 0,6646 | 4 |
| hsa04670 | Leukocyte transendothelial migration | 4/276 | 116/9436 | 0,0345 | 1,1789 | 0,3365 | 0,4419 | 0,7823 | 0,6674 | 4 |
| hsa04979 | Cholesterol metabolism | 2/276 | 51/9436 | 0,0392 | 1,3407 | 0,4235 | 0,4423 | 0,7823 | 0,6674 | 2 |
| hsa05207 | Chemical carcinogenesis - receptor activation | 7/276 | 217/9436 | 0,0323 | 1,1029 | 0,2661 | 0,4507 | 0,7884 | 0,6726 | 7 |
| hsa04973 | Carbohydrate digestion and absorption | 2/276 | 52/9436 | 0,0385 | 1,3149 | 0,3953 | 0,4522 | 0,7884 | 0,6726 | 2 |
| hsa05132 | Salmonella infection | 8/276 | 251/9436 | 0,0319 | 1,0897 | 0,2499 | 0,4534 | 0,7884 | 0,6726 | 8 |
| hsa04742 | Taste transduction | 3/276 | 86/9436 | 0,0349 | 1,1926 | 0,3115 | 0,4627 | 0,7999 | 0,6824 | 3 |
| hsa05320 | Autoimmune thyroid disease | 2/276 | 54/9436 | 0,0370 | 1,2662 | 0,3406 | 0,4717 | 0,8110 | 0,6918 | 2 |
| hsa00600 | Sphingolipid metabolism | 2/276 | 55/9436 | 0,0364 | 1,2432 | 0,3140 | 0,4813 | 0,8147 | 0,6950 | 2 |
| hsa04727 | GABAergic synapse | 3/276 | 89/9436 | 0,0337 | 1,1524 | 0,2508 | 0,4853 | 0,8147 | 0,6950 | 3 |
| hsa04932 | Non-alcoholic fatty liver disease | 5/276 | 157/9436 | 0,0318 | 1,0888 | 0,1948 | 0,4879 | 0,8147 | 0,6950 | 5 |
| hsa04340 | Hedgehog signaling pathway | 2/276 | 56/9436 | 0,0357 | 1,2210 | 0,2879 | 0,4909 | 0,8147 | 0,6950 | 2 |
| hsa04211 | Longevity regulating pathway | 3/276 | 90/9436 | 0,0333 | 1,1396 | 0,2310 | 0,4928 | 0,8147 | 0,6950 | 3 |
| hsa04976 | Bile secretion | 3/276 | 90/9436 | 0,0333 | 1,1396 | 0,2310 | 0,4928 | 0,8147 | 0,6950 | 3 |
| hsa05235 | PD-L1 expression and PD-1 checkpoint pathway in cancer | 3/276 | 90/9436 | 0,0333 | 1,1396 | 0,2310 | 0,4928 | 0,8147 | 0,6950 | 3 |
| hsa00220 | Arginine biosynthesis | 1/276 | 23/9436 | 0,0435 | 1,4865 | 0,4054 | 0,4952 | 0,8147 | 0,6950 | 1 |
| hsa05208 | Chemical carcinogenesis - reactive oxygen species | 7/276 | 227/9436 | 0,0308 | 1,0543 | 0,1437 | 0,4980 | 0,8149 | 0,6952 | 7 |
| hsa04514 | Cell adhesion molecules | 5/276 | 160/9436 | 0,0313 | 1,0684 | 0,1514 | 0,5046 | 0,8190 | 0,6987 | 5 |
| hsa04611 | Platelet activation | 4/276 | 126/9436 | 0,0317 | 1,0853 | 0,1674 | 0,5059 | 0,8190 | 0,6987 | 4 |
| hsa00534 | Glycosaminoglycan biosynthesis - heparan sulfate / heparin | 1/276 | 24/9436 | 0,0417 | 1,4245 | 0,3614 | 0,5100 | 0,8214 | 0,7007 | 1 |
| hsa04923 | Regulation of lipolysis in adipocytes | 2/276 | 59/9436 | 0,0339 | 1,1589 | 0,2126 | 0,5187 | 0,8233 | 0,7024 | 2 |
| hsa05213 | Endometrial cancer | 2/276 | 59/9436 | 0,0339 | 1,1589 | 0,2126 | 0,5187 | 0,8233 | 0,7024 | 2 |
| hsa04370 | VEGF signaling pathway | 2/276 | 60/9436 | 0,0333 | 1,1396 | 0,1883 | 0,5278 | 0,8233 | 0,7024 | 2 |
| hsa04926 | Relaxin signaling pathway | 4/276 | 130/9436 | 0,0308 | 1,0520 | 0,1035 | 0,5306 | 0,8233 | 0,7024 | 4 |
| hsa04978 | Mineral absorption | 2/276 | 61/9436 | 0,0328 | 1,1209 | 0,1645 | 0,5367 | 0,8233 | 0,7024 | 2 |
| hsa04022 | cGMP-PKG signaling pathway | 5/276 | 166/9436 | 0,0301 | 1,0298 | 0,0672 | 0,5374 | 0,8233 | 0,7024 | 5 |
| hsa05012 | Parkinson disease | 8/276 | 271/9436 | 0,0295 | 1,0093 | 0,0268 | 0,5406 | 0,8233 | 0,7024 | 8 |

| | | | | | | | | | | |
|---|---|---|---|---|---|---|---|---|---|---|
| hsa04970 | Salivary secretion | 3/276 | 97/9436 | 0,0309 | 1,0574 | 0,0986 | 0,5433 | 0,8233 | 0,7024 | 3 |
| hsa04630 | JAK-STAT signaling pathway | 5/276 | 168/9436 | 0,0298 | 1,0175 | 0,0398 | 0,5481 | 0,8233 | 0,7024 | 5 |
| hsa05130 | Pathogenic Escherichia coli infection | 6/276 | 203/9436 | 0,0296 | 1,0105 | 0,0262 | 0,5482 | 0,8233 | 0,7024 | 6 |
| hsa01040 | Biosynthesis of unsaturated fatty acids | 1/276 | 27/9436 | 0,0370 | 1,2662 | 0,2405 | 0,5519 | 0,8233 | 0,7024 | 1 |
| hsa00140 | Steroid hormone biosynthesis | 2/276 | 63/9436 | 0,0317 | 1,0853 | 0,1180 | 0,5542 | 0,8233 | 0,7024 | 2 |
| hsa03420 | Nucleotide excision repair | 2/276 | 63/9436 | 0,0317 | 1,0853 | 0,1180 | 0,5542 | 0,8233 | 0,7024 | 2 |
| hsa04270 | Vascular smooth muscle contraction | 4/276 | 134/9436 | 0,0299 | 1,0205 | 0,0416 | 0,5546 | 0,8233 | 0,7024 | 4 |
| hsa04750 | Inflammatory mediator regulation of TRP channels | 3/276 | 99/9436 | 0,0303 | 1,0360 | 0,0625 | 0,5572 | 0,8233 | 0,7024 | 3 |
| hsa04141 | Protein processing in endoplasmic reticulum | 5/276 | 171/9436 | 0,0292 | 0,9997 | -0,0008 | 0,5640 | 0,8233 | 0,7024 | 5 |
| hsa04640 | Hematopoietic cell lineage | 3/276 | 100/9436 | 0,0300 | 1,0257 | 0,0448 | 0,5641 | 0,8233 | 0,7024 | 3 |
| hsa00601 | Glycosphingolipid biosynthesis - lacto and neolacto series | 1/276 | 28/9436 | 0,0357 | 1,2210 | 0,2033 | 0,5650 | 0,8233 | 0,7024 | 1 |
| hsa00790 | Folate biosynthesis | 1/276 | 28/9436 | 0,0357 | 1,2210 | 0,2033 | 0,5650 | 0,8233 | 0,7024 | 1 |
| hsa04966 | Collecting duct acid secretion | 1/276 | 28/9436 | 0,0357 | 1,2210 | 0,2033 | 0,5650 | 0,8233 | 0,7024 | 1 |
| hsa04392 | Hippo signaling pathway - multiple species | 1/276 | 29/9436 | 0,0345 | 1,1789 | 0,1675 | 0,5778 | 0,8275 | 0,7059 | 1 |
| hsa05321 | Inflammatory bowel disease | 2/276 | 66/9436 | 0,0303 | 1,0360 | 0,0510 | 0,5796 | 0,8275 | 0,7059 | 2 |
| hsa04114 | Oocyte meiosis | 4/276 | 139/9436 | 0,0288 | 0,9838 | -0,0333 | 0,5838 | 0,8275 | 0,7059 | 4 |
| hsa00010 | Glycolysis / Gluconeogenesis | 2/276 | 67/9436 | 0,0299 | 1,0205 | 0,0293 | 0,5878 | 0,8275 | 0,7059 | 2 |
| hsa00020 | Citrate cycle (TCA cycle) | 1/276 | 30/9436 | 0,0333 | 1,1396 | 0,1329 | 0,5902 | 0,8275 | 0,7059 | 1 |
| hsa01523 | Antifolate resistance | 1/276 | 30/9436 | 0,0333 | 1,1396 | 0,1329 | 0,5902 | 0,8275 | 0,7059 | 1 |
| hsa05206 | MicroRNAs in cancer | 9/276 | 320/9436 | 0,0281 | 0,9615 | -0,1215 | 0,5962 | 0,8275 | 0,7059 | 9 |
| hsa05010 | Alzheimer disease | 11/276 | 391/9436 | 0,0281 | 0,9618 | -0,1338 | 0,5970 | 0,8275 | 0,7059 | 11 |
| hsa00030 | Pentose phosphate pathway | 1/276 | 31/9436 | 0,0323 | 1,1029 | 0,0996 | 0,6022 | 0,8275 | 0,7059 | 1 |
| hsa03060 | Protein export | 1/276 | 31/9436 | 0,0323 | 1,1029 | 0,0996 | 0,6022 | 0,8275 | 0,7059 | 1 |
| hsa04981 | Folate transport and metabolism | 1/276 | 31/9436 | 0,0323 | 1,1029 | 0,0996 | 0,6022 | 0,8275 | 0,7059 | 1 |
| hsa04664 | Fc epsilon RI signaling pathway | 2/276 | 69/9436 | 0,0290 | 0,9910 | -0,0131 | 0,6039 | 0,8275 | 0,7059 | 2 |
| hsa04936 | Alcoholic liver disease | 4/276 | 144/9436 | 0,0278 | 0,9497 | -0,1056 | 0,6118 | 0,8275 | 0,7059 | 4 |
| hsa00052 | Galactose metabolism | 1/276 | 32/9436 | 0,0313 | 1,0684 | 0,0673 | 0,6139 | 0,8275 | 0,7059 | 1 |
| hsa00640 | Propanoate metabolism | 1/276 | 32/9436 | 0,0313 | 1,0684 | 0,0673 | 0,6139 | 0,8275 | 0,7059 | 1 |
| hsa04136 | Autophagy - other | 1/276 | 32/9436 | 0,0313 | 1,0684 | 0,0673 | 0,6139 | 0,8275 | 0,7059 | 1 |
| hsa04215 | Apoptosis - multiple species | 1/276 | 32/9436 | 0,0313 | 1,0684 | 0,0673 | 0,6139 | 0,8275 | 0,7059 | 1 |
| hsa05152 | Tuberculosis | 5/276 | 182/9436 | 0,0275 | 0,9392 | -0,1437 | 0,6196 | 0,8316 | 0,7094 | 5 |
| hsa04130 | SNARE interactions in vesicular transport | 1/276 | 33/9436 | 0,0303 | 1,0360 | 0,0360 | 0,6252 | 0,8354 | 0,7127 | 1 |
| hsa03020 | RNA polymerase | 1/276 | 34/9436 | 0,0294 | 1,0055 | 0,0056 | 0,6362 | 0,8427 | 0,7190 | 1 |
| hsa04710 | Circadian rhythm | 1/276 | 34/9436 | 0,0294 | 1,0055 | 0,0056 | 0,6362 | 0,8427 | 0,7190 | 1 |
| hsa01230 | Biosynthesis of amino acids | 2/276 | 75/9436 | 0,0267 | 0,9117 | -0,1333 | 0,6494 | 0,8460 | 0,7217 | 2 |
| hsa00512 | Mucin type O-glycan biosynthesis | 1/276 | 36/9436 | 0,0278 | 0,9497 | -0,0525 | 0,6572 | 0,8460 | 0,7217 | 1 |
| hsa04928 | Parathyroid hormone synthesis, secretion and action | 3/276 | 115/9436 | 0,0261 | 0,8919 | -0,2025 | 0,6588 | 0,8460 | 0,7217 | 3 |
| hsa05034 | Alcoholism | 5/276 | 191/9436 | 0,0262 | 0,8950 | -0,2545 | 0,6619 | 0,8460 | 0,7217 | 5 |
| hsa00250 | Alanine, aspartate and glutamate metabolism | 1/276 | 37/9436 | 0,0270 | 0,9240 | -0,0804 | 0,6673 | 0,8460 | 0,7217 | 1 |
| hsa05143 | African trypanosomiasis | 1/276 | 37/9436 | 0,0270 | 0,9240 | -0,0804 | 0,6673 | 0,8460 | 0,7217 | 1 |
| hsa05216 | Thyroid cancer | 1/276 | 37/9436 | 0,0270 | 0,9240 | -0,0804 | 0,6673 | 0,8460 | 0,7217 | 1 |
| hsa01200 | Carbon metabolism | 3/276 | 117/9436 | 0,0256 | 0,8766 | -0,2331 | 0,6702 | 0,8460 | 0,7217 | 3 |
| hsa04062 | Chemokine signaling pathway | 5/276 | 193/9436 | 0,0259 | 0,8857 | -0,2785 | 0,6708 | 0,8460 | 0,7217 | 5 |
| hsa00520 | Amino sugar and nucleotide sugar metabolism | 1/276 | 38/9436 | 0,0263 | 0,8997 | -0,1075 | 0,6771 | 0,8460 | 0,7217 | 1 |
| hsa04960 | Aldosterone-regulated sodium reabsorption | 1/276 | 38/9436 | 0,0263 | 0,8997 | -0,1075 | 0,6771 | 0,8460 | 0,7217 | 1 |
| hsa05340 | Primary immunodeficiency | 1/276 | 38/9436 | 0,0263 | 0,8997 | -0,1075 | 0,6771 | 0,8460 | 0,7217 | 1 |
| hsa04721 | Synaptic vesicle cycle | 2/276 | 79/9436 | 0,0253 | 0,8655 | -0,2083 | 0,6773 | 0,8460 | 0,7217 | 2 |
| hsa05140 | Leishmaniasis | 2/276 | 79/9436 | 0,0253 | 0,8655 | -0,2083 | 0,6773 | 0,8460 | 0,7217 | 2 |
| hsa04613 | Neutrophil extracellular trap formation | 5/276 | 196/9436 | 0,0255 | 0,8722 | -0,3139 | 0,6840 | 0,8474 | 0,7229 | 5 |
| hsa05167 | Kaposi sarcoma-associated herpesvirus infection | 5/276 | 196/9436 | 0,0255 | 0,8722 | -0,3139 | 0,6840 | 0,8474 | 0,7229 | 5 |
| hsa00983 | Drug metabolism - other enzymes | 2/276 | 81/9436 | 0,0247 | 0,8442 | -0,2445 | 0,6906 | 0,8521 | 0,7269 | 2 |
| hsa04660 | T cell receptor signaling pathway | 3/276 | 122/9436 | 0,0246 | 0,8407 | -0,3074 | 0,6976 | 0,8573 | 0,7314 | 3 |
| hsa03440 | Homologous recombination | 1/276 | 41/9436 | 0,0244 | 0,8339 | -0,1850 | 0,7047 | 0,8591 | 0,7329 | 1 |
| hsa05219 | Bladder cancer | 1/276 | 41/9436 | 0,0244 | 0,8339 | -0,1850 | 0,7047 | 0,8591 | 0,7329 | 1 |
| hsa04216 | Ferroptosis | 1/276 | 42/9436 | 0,0238 | 0,8140 | -0,2097 | 0,7134 | 0,8657 | 0,7386 | 1 |
| hsa01232 | Nucleotide metabolism | 2/276 | 85/9436 | 0,0235 | 0,8044 | -0,3144 | 0,7158 | 0,8657 | 0,7386 | 2 |
| hsa04975 | Fat digestion and absorption | 1/276 | 43/9436 | 0,0233 | 0,7951 | -0,2338 | 0,7218 | 0,8662 | 0,7390 | 1 |
| hsa04012 | ErbB signaling pathway | 2/276 | 86/9436 | 0,0233 | 0,7951 | -0,3314 | 0,7218 | 0,8662 | 0,7390 | 2 |
| hsa00230 | Purine metabolism | 3/276 | 128/9436 | 0,0234 | 0,8013 | -0,3929 | 0,7282 | 0,8691 | 0,7415 | 3 |
| hsa04962 | Vasopressin-regulated water reabsorption | 1/276 | 44/9436 | 0,0227 | 0,7770 | -0,2573 | 0,7300 | 0,8691 | 0,7415 | 1 |
| hsa04610 | Complement and coagulation cascades | 2/276 | 88/9436 | 0,0227 | 0,7770 | -0,3648 | 0,7336 | 0,8700 | 0,7422 | 2 |
| hsa00860 | Porphyrin metabolism | 1/276 | 46/9436 | 0,0217 | 0,7432 | -0,3030 | 0,7456 | 0,8764 | 0,7476 | 1 |
| hsa05164 | Influenza A | 4/276 | 173/9436 | 0,0231 | 0,7905 | -0,4828 | 0,7501 | 0,8764 | 0,7476 | 4 |
| hsa04662 | B cell receptor signaling pathway | 2/276 | 91/9436 | 0,0220 | 0,7514 | -0,4136 | 0,7504 | 0,8764 | 0,7476 | 2 |
| hsa05032 | Morphine addiction | 2/276 | 91/9436 | 0,0220 | 0,7514 | -0,4136 | 0,7504 | 0,8764 | 0,7476 | 2 |

| | | | | | | | | | | |
|---|---|---|---|---|---|---|---|---|---|---|
| hsa04650 | Natural killer cell mediated cytotoxicity | 3/276 | 134/9436 | 0,0224 | 0,7654 | -0,4747 | 0,7563 | 0,8787 | 0,7497 | 3 |
| hsa03272 | Virion - Hepatitis viruses | 1/276 | 48/9436 | 0,0208 | 0,7123 | -0,3469 | 0,7603 | 0,8787 | 0,7497 | 1 |
| hsa04080 | Neuroactive ligand-receptor interaction | 9/276 | 370/9436 | 0,0243 | 0,8316 | -0,5736 | 0,7610 | 0,8787 | 0,7497 | 9 |
| hsa05323 | Rheumatoid arthritis | 2/276 | 95/9436 | 0,0211 | 0,7198 | -0,4765 | 0,7714 | 0,8816 | 0,7521 | 2 |
| hsa04081 | Hormone signaling | 5/276 | 219/9436 | 0,0228 | 0,7806 | -0,5703 | 0,7730 | 0,8816 | 0,7521 | 5 |
| hsa04672 | Intestinal immune network for IgA production | 1/276 | 50/9436 | 0,0200 | 0,6838 | -0,3892 | 0,7742 | 0,8816 | 0,7521 | 1 |
| hsa04915 | Estrogen signaling pathway | 3/276 | 139/9436 | 0,0216 | 0,7379 | -0,5404 | 0,7779 | 0,8816 | 0,7521 | 3 |
| hsa05162 | Measles | 3/276 | 139/9436 | 0,0216 | 0,7379 | -0,5404 | 0,7779 | 0,8816 | 0,7521 | 3 |
| hsa04913 | Ovarian steroidogenesis | 1/276 | 52/9436 | 0,0192 | 0,6575 | -0,4299 | 0,7873 | 0,8890 | 0,7584 | 1 |
| hsa04961 | Endocrine and other factor-regulated calcium reabsorption | 1/276 | 53/9436 | 0,0189 | 0,6451 | -0,4498 | 0,7936 | 0,8900 | 0,7592 | 1 |
| hsa04380 | Osteoclast differentiation | 3/276 | 143/9436 | 0,0210 | 0,7172 | -0,5914 | 0,7940 | 0,8900 | 0,7592 | 3 |
| hsa03015 | mRNA surveillance pathway | 2/276 | 103/9436 | 0,0194 | 0,6639 | -0,5954 | 0,8088 | 0,9032 | 0,7705 | 2 |
| hsa00480 | Glutathione metabolism | 1/276 | 59/9436 | 0,0169 | 0,5795 | -0,5624 | 0,8274 | 0,9204 | 0,7852 | 1 |
| hsa04620 | Toll-like receptor signaling pathway | 2/276 | 109/9436 | 0,0183 | 0,6273 | -0,6793 | 0,8331 | 0,9204 | 0,7852 | 2 |
| hsa04659 | Th17 cell differentiation | 2/276 | 109/9436 | 0,0183 | 0,6273 | -0,6793 | 0,8331 | 0,9204 | 0,7852 | 2 |
| hsa04213 | Longevity regulating pathway - multiple species | 1/276 | 62/9436 | 0,0161 | 0,5514 | -0,6151 | 0,8422 | 0,9245 | 0,7887 | 1 |
| hsa00590 | Arachidonic acid metabolism | 1/276 | 63/9436 | 0,0159 | 0,5427 | -0,6322 | 0,8469 | 0,9245 | 0,7887 | 1 |
| hsa04145 | Phagosome | 3/276 | 159/9436 | 0,0189 | 0,6451 | -0,7835 | 0,8490 | 0,9245 | 0,7887 | 3 |
| hsa04217 | Necroptosis | 3/276 | 159/9436 | 0,0189 | 0,6451 | -0,7835 | 0,8490 | 0,9245 | 0,7887 | 3 |
| hsa05169 | Epstein-Barr virus infection | 4/276 | 204/9436 | 0,0196 | 0,6704 | -0,8262 | 0,8527 | 0,9253 | 0,7894 | 4 |
| hsa04668 | TNF signaling pathway | 2/276 | 119/9436 | 0,0168 | 0,5746 | -0,8106 | 0,8676 | 0,9350 | 0,7977 | 2 |
| hsa05131 | Shigellosis | 5/276 | 253/9436 | 0,0198 | 0,6757 | -0,9077 | 0,8678 | 0,9350 | 0,7977 | 5 |
| hsa04920 | Adipocytokine signaling pathway | 1/276 | 70/9436 | 0,0143 | 0,4884 | -0,7457 | 0,8758 | 0,9354 | 0,7980 | 1 |
| hsa04917 | Prolactin signaling pathway | 1/276 | 71/9436 | 0,0141 | 0,4815 | -0,7612 | 0,8794 | 0,9354 | 0,7980 | 1 |
| hsa05417 | Lipid and atherosclerosis | 4/276 | 216/9436 | 0,0185 | 0,6331 | -0,9468 | 0,8816 | 0,9354 | 0,7980 | 4 |
| hsa04622 | RIG-I-like receptor signaling pathway | 1/276 | 72/9436 | 0,0139 | 0,4748 | -0,7764 | 0,8830 | 0,9354 | 0,7980 | 1 |
| hsa04382 | Cornified envelope formation | 4/276 | 217/9436 | 0,0184 | 0,6302 | -0,9566 | 0,8837 | 0,9354 | 0,7980 | 4 |
| hsa00982 | Drug metabolism - cytochrome P450 | 1/276 | 73/9436 | 0,0137 | 0,4683 | -0,7915 | 0,8865 | 0,9354 | 0,7980 | 1 |
| hsa01524 | Platinum drug resistance | 1/276 | 75/9436 | 0,0133 | 0,4558 | -0,8212 | 0,8930 | 0,9391 | 0,8011 | 1 |
| hsa03320 | PPAR signaling pathway | 1/276 | 76/9436 | 0,0132 | 0,4498 | -0,8359 | 0,8962 | 0,9392 | 0,8012 | 1 |
| hsa05163 | Human cytomegalovirus infection | 4/276 | 226/9436 | 0,0177 | 0,6051 | -1,0430 | 0,9017 | 0,9418 | 0,8034 | 4 |
| hsa05168 | Herpes simplex virus 1 infection | 3/276 | 182/9436 | 0,0165 | 0,5635 | -1,0320 | 0,9056 | 0,9426 | 0,8041 | 3 |
| hsa00190 | Oxidative phosphorylation | 2/276 | 138/9436 | 0,0145 | 0,4955 | -1,0363 | 0,9158 | 0,9468 | 0,8077 | 2 |
| hsa04146 | Peroxisome | 1/276 | 83/9436 | 0,0120 | 0,4119 | -0,9341 | 0,9158 | 0,9468 | 0,8077 | 1 |
| hsa05160 | Hepatitis C | 2/276 | 159/9436 | 0,0126 | 0,4300 | -1,2581 | 0,9497 | 0,9756 | 0,8323 | 2 |
| hsa05170 | Human immunodeficiency virus 1 infection | 3/276 | 213/9436 | 0,0141 | 0,4815 | -1,3285 | 0,9517 | 0,9756 | 0,8323 | 3 |
| hsa04064 | NF-kappa B signaling pathway | 1/276 | 105/9436 | 0,0095 | 0,3256 | -1,2062 | 0,9565 | 0,9756 | 0,8323 | 1 |
| hsa04625 | C-type lectin receptor signaling pathway | 1/276 | 105/9436 | 0,0095 | 0,3256 | -1,2062 | 0,9565 | 0,9756 | 0,8323 | 1 |
| hsa04060 | Cytokine-cytokine receptor interaction | 4/276 | 298/9436 | 0,0134 | 0,4589 | -1,6475 | 0,9772 | 0,9934 | 0,8475 | 4 |
| hsa05418 | Fluid shear stress and atherosclerosis | 1/276 | 142/9436 | 0,0070 | 0,2408 | -1,5823 | 0,9857 | 0,9988 | 0,8521 | 1 |
| hsa04621 | NOD-like receptor signaling pathway | 1/276 | 189/9436 | 0,0053 | 0,1809 | -1,9745 | 0,9965 | 1,0000 | 0,8531 | 1 |
| hsa03040 | Spliceosome | 1/276 | 235/9436 | 0,0043 | 0,1455 | -2,3026 | 0,9991 | 1,0000 | 0,8531 | 1 |
| hsa05171 | Coronavirus disease - COVID-19 | 1/276 | 238/9436 | 0,0042 | 0,1436 | -2,3226 | 0,9992 | 1,0000 | 0,8531 | 1 |
| hsa04740 | Olfactory transduction | 1/276 | 453/9436 | 0,0022 | 0,0755 | -3,5006 | 1,0000 | 1,0000 | 0,8531 | 1 |

**Table S15. Quality control of toxic and essential metals in whole blood.**

| | Seronorm whole blod-1103128 L-1 | | Seronorm whole blod-1406264 L-2 | | |
|---|---|---|---|---|---|
| Metals | Reference value/interval | Obtained value | Reference value/interval | Obtained value | LOD1 |
| As (µg/L) | 2.4±0.5 | 2.6±0.6 | 12.7±1.6 | 14.1±2.8 | 0,14 |
| Ca (mg/L) | 15 | 12.0±0.86 | 15 | 13.8±0.70 | 0,31 |
| Cd (µg/L) | 0.36±0.02 | 0.32±0.04 | 5.01±1.01 | 4.78±0.25 | 0,004 |
| Co (µg/L) | 0.16±0.03 | 0.15±0.02 | 5.18±1.04 | 4.56±0.20 | 0,003 |
| Cu (mg/L) | 0.68±0.14 | 0.56±0.029 | 1.34±0.27 | 1.13±0.049 | 0,09 |
| Fe (mg/L) | 331 | 254±12.5 | 332 | 258±14.1 | 1,65 |
| Hg (µg/L) | 1.5±0.3 | 1.6±0.3 | 17±3.4 | 18±3.2 | 0,01 |
| Mg (mg/L) | 16,2 | 13.9±0.62 | 14,4 | 14.3±0.51 | 0,07 |
| Mn (µg/L) | 20.7±4.2 | 16.7±0.7 | 31.4±6.3 | 26.9±1.1 | 0,002 |
| Mo (µg/L) | 0.94±0.22 | 0.94±0.09 | 5.31±1.07 | 4.63±0.23 | 0,004 |
| Pb (µg/L) | 10.2±2.1 | 9.33±0.7 | 337±68 | 291±15 | 0,006 |
| Se (µg/L) | 59±12 | 56±2.8 | 161±32 | 152±7 | 0,03 |
| Zn (mg/L) | 4.4±0.2 | 3.7±0.2 | 7.1±1.4 | 6.5±0.3 | 2,22 |

**Table S16. The feature selection stability assessment for multiomics models.**

| Model | feature | loading | stability | block | component |
|---|---|---|---|---|---|
| Multiomic-SCFB | 177_t-PA | -0,35 | 1 | Serum | Comp1 |
| Multiomic-SCFB | 139_PRL | 0,31 | 1 | Serum | Comp1 |
| Multiomic-SCFB | 117_Gal-3 | -0,30 | 1 | Serum | Comp1 |
| Multiomic-SCFB | 142_FasL | -0,23 | 1 | Serum | Comp1 |
| Multiomic-SCFB | 143_BAFF | -0,22 | 1 | Serum | Comp1 |
| Multiomic-SCFB | 132_SCF | -0,20 | 1 | Serum | Comp1 |
| Multiomic-SCFB | 181_VEGF-D | 0,30 | 0,99 | Serum | Comp1 |
| Multiomic-SCFB | 107_CDCP1 | -0,27 | 0,99 | Serum | Comp1 |
| Multiomic-SCFB | 114_CSTB | -0,25 | 0,99 | Serum | Comp1 |
| Multiomic-SCFB | 190_CCL20 | -0,25 | 0,99 | Serum | Comp1 |
| Multiomic-SCFB | 110_OPG | -0,20 | 0,99 | Serum | Comp1 |
| Multiomic-SCFB | 164_CTSD | -0,14 | 0,97 | Serum | Comp1 |
| Multiomic-SCFB | 194_ADA | -0,12 | 0,95 | Serum | Comp1 |
| Multiomic-SCFB | 113_IL-6 | -0,17 | 0,94 | Serum | Comp1 |
| Multiomic-SCFB | 179_MIC-A | 0,18 | 0,92 | Serum | Comp1 |
| Multiomic-SCFB | 164_CCL23 | 0,17 | 0,92 | Serum | Comp1 |
| Multiomic-SCFB | 183_CXCL13 | 0,15 | 0,92 | Serum | Comp1 |
| Multiomic-SCFB | 172_TNFRSF4 | -0,11 | 0,9 | Serum | Comp1 |
| Multiomic-SCFB | 157_LITAF | 0,11 | 0,84 | Serum | Comp1 |
| Multiomic-SCFB | 134_IL-6RA | -0,11 | 0,83 | Serum | Comp1 |
| Multiomic-SCFB | 153_ErbB2/HER2 | -0,11 | 0,82 | Serum | Comp1 |
| Multiomic-SCFB | 112_IL-1ra | -0,11 | 0,75 | Serum | Comp1 |
| Multiomic-SCFB | 195_IL-17RB | 0,09 | 0,73 | Serum | Comp1 |
| Multiomic-SCFB | 135_TNF-R2 | -0,08 | 0,72 | Serum | Comp1 |
| Multiomic-SCFB | 165_CDH3 | -0,08 | 0,69 | Serum | Comp1 |
| Multiomic-SCFB | 154_ErbB3/HER3 | -0,04 | 0,49 | Serum | Comp1 |
| Multiomic-SCFB | 174_AGRP | 0,02 | 0,44 | Serum | Comp1 |
| Multiomic-SCFB | 131_FGF-23 | -0,03 | 0,43 | Serum | Comp1 |
| Multiomic-SCFB | 175_REG-4 | 0,00 | 0,35 | Serum | Comp1 |
| Multiomic-SCFB | 177_AR | 0,02 | 0,35 | Serum | Comp1 |
| Multiomic-SCFB | 175_REG-4 | -0,47 | 0,92 | Serum | Comp2 |
| Multiomic-SCFB | 150_CHI3L1 | -0,34 | 0,91 | Serum | Comp2 |
| Multiomic-SCFB | 145_CCL19 | -0,29 | 0,82 | Serum | Comp2 |
| Multiomic-SCFB | 166_MIP-1 alpha | -0,26 | 0,79 | Serum | Comp2 |
| Multiomic-SCFB | 183_SPON1 | -0,42 | 0,74 | Serum | Comp2 |
| Multiomic-SCFB | 171_Dkk-1 | -0,17 | 0,7 | Serum | Comp2 |
| Multiomic-SCFB | 155_ErbB4/HER4 | 0,20 | 0,67 | Serum | Comp2 |
| Multiomic-SCFB | 152_Ep-CAM | 0,16 | 0,66 | Serum | Comp2 |
| Multiomic-SCFB | 182_MMP-12 | -0,26 | 0,64 | Serum | Comp2 |
| Multiomic-SCFB | 189_TWEAK | -0,22 | 0,63 | Serum | Comp2 |
| Multiomic-SCFB | 103_AM | -0,08 | 0,61 | Serum | Comp2 |
| Multiomic-SCFB | 101_IL-8 | -0,08 | 0,6 | Serum | Comp2 |
| Multiomic-SCFB | 151_ST2 | 0,03 | 0,58 | Serum | Comp2 |
| Multiomic-SCFB | 179_ESM-1 | -0,30 | 0,54 | Serum | Comp2 |
| Multiomic-SCFB | 158_MB | 0,02 | 0,52 | Serum | Comp2 |
| Multiomic-SCFB | 148_PTPN22 | 0,07 | 0,46 | Serum | Comp2 |
| Multiomic-SCFB | 159_MIA | -0,04 | 0,41 | Serum | Comp2 |
| Multiomic-SCFB | 156_HGF | -0,10 | 0,39 | Serum | Comp2 |
| Multiomic-SCFB | 148_PTX3 | -0,07 | 0,39 | Serum | Comp2 |
| Multiomic-SCFB | 170_4E-BP1 | 0,00 | 0,29 | Serum | Comp2 |
| Multiomic-SCFB | 142_MMP-1 | -0,04 | 0,28 | Serum | Comp2 |
| Multiomic-SCFB | 178_IFN-gamma | 0,03 | 0,27 | Serum | Comp2 |
| Multiomic-SCFB | 149_EMMPRIN | -0,02 | 0,27 | Serum | Comp2 |
| Multiomic-SCFB | 145_PAPPA | -0,06 | 0,25 | Serum | Comp2 |
| Multiomic-SCFB | 173_GAL | 0,04 | 0,1 | Serum | Comp2 |
| Multiomic-SCFB | 140_FUR | -0,55 | 1 | CSF | Comp1 |
| Multiomic-SCFB | 141_GH | 0,77 | 0,99 | CSF | Comp1 |
| Multiomic-SCFB | 160_CXCL5 | -0,22 | 0,67 | CSF | Comp1 |
| Multiomic-SCFB | 131_IL-7 | -0,24 | 0,65 | CSF | Comp1 |
| Multiomic-SCFB | 110_ILT-3 | -0,05 | 0,18 | CSF | Comp1 |
| Multiomic-SCFB | 193_FS | 0,39 | 0,79 | CSF | Comp2 |
| Multiomic-SCFB | 171_Dkk-1 | 0,31 | 0,77 | CSF | Comp2 |
| Multiomic-SCFB | 151_IL-18R1 | 0,28 | 0,75 | CSF | Comp2 |
| Multiomic-SCFB | 157_PSGL-1 | 0,35 | 0,72 | CSF | Comp2 |
| Multiomic-SCFB | 120_TRAIL | 0,26 | 0,7 | CSF | Comp2 |
| Multiomic-SCFB | 160_CXCL5 | 0,26 | 0,69 | CSF | Comp2 |

| | | | | | |
|---|---|---|---|---|---|
| Multiomic-SCFB | 151_CXCL10 | 0,16 | 0,69 | CSF | Comp2 |
| Multiomic-SCFB | 179_ESM-1 | 0,14 | 0,67 | CSF | Comp2 |
| Multiomic-SCFB | 177_t-PA | 0,11 | 0,66 | CSF | Comp2 |
| Multiomic-SCFB | 185_ITGA1 | -0,29 | 0,5 | CSF | Comp2 |
| Multiomic-SCFB | 113_IL-6 | 0,21 | 0,49 | CSF | Comp2 |
| Multiomic-SCFB | 164_CTSD | 0,14 | 0,45 | CSF | Comp2 |
| Multiomic-SCFB | 182_HE4 | -0,27 | 0,4 | CSF | Comp2 |
| Multiomic-SCFB | 187_TNFRSF9 | -0,30 | 0,38 | CSF | Comp2 |
| Multiomic-SCFB | 194_ADA | -0,13 | 0,37 | CSF | Comp2 |
| Multiomic-SCFB | 152_Ep-CAM | -0,11 | 0,36 | CSF | Comp2 |
| Multiomic-SCFB | 196_FR-alpha | -0,02 | 0,36 | CSF | Comp2 |
| Multiomic-SCFB | 149_IL-10RB | -0,11 | 0,35 | CSF | Comp2 |
| Multiomic-SCFB | 166_CCL3 | 0,07 | 0,28 | CSF | Comp2 |
| Multiomic-SCFB | 115_MCP-1 | 0,01 | 0,28 | CSF | Comp2 |
| Multiomic-SCFB | BMg | -0,43 | 1 | Blood | Comp1 |
| Multiomic-SCFB | Bfe | -0,41 | 1 | Blood | Comp1 |
| Multiomic-SCFB | BZn | -0,38 | 1 | Blood | Comp1 |
| Multiomic-SCFB | BMn | -0,37 | 1 | Blood | Comp1 |
| Multiomic-SCFB | Bmo | 0,33 | 1 | Blood | Comp1 |
| Multiomic-SCFB | Bca | 0,31 | 1 | Blood | Comp1 |
| Multiomic-SCFB | BPb | -0,29 | 1 | Blood | Comp1 |
| Multiomic-SCFB | Bse | -0,26 | 1 | Blood | Comp1 |
| Multiomic-SCFB | Bas | 0,04 | 0,72 | Blood | Comp1 |
| Multiomic-SCFB | BCd | 0,57 | 1 | Blood | Comp2 |
| Multiomic-SCFB | Bmo | -0,39 | 1 | Blood | Comp2 |
| Multiomic-SCFB | Bcu | 0,31 | 1 | Blood | Comp2 |
| Multiomic-SCFB | Bca | 0,29 | 1 | Blood | Comp2 |
| Multiomic-SCFB | Bse | 0,29 | 1 | Blood | Comp2 |
| Multiomic-SCFB | BPb | 0,26 | 1 | Blood | Comp2 |
| Multiomic-SCFB | Bco | 0,25 | 1 | Blood | Comp2 |
| Multiomic-SCFB | BMg | -0,18 | 1 | Blood | Comp2 |
| Multiomic-SCFB | BMn | -0,18 | 1 | Blood | Comp2 |
| Multiomic-SCFB | Bas | -0,17 | 1 | Blood | Comp2 |
| Multiomic-SCFB | Bfe | -0,14 | 1 | Blood | Comp2 |
| Multiomic-SCFB | BHg | -0,08 | 1 | Blood | Comp2 |
| Multiomic-SCFB | BZn | 0,02 | 1 | Blood | Comp2 |
| Multiomic-SCFB | 3-methyladipate | 0,83 | 0,77 | Fecal | Comp1 |
| Multiomic-SCFB | 2'-deoxyinosine | 0,38 | 0,52 | Fecal | Comp1 |
| Multiomic-SCFB | N-acetylglutamate | 0,34 | 0,51 | Fecal | Comp1 |
| Multiomic-SCFB | taurochenodeoxycholate | -0,19 | 0,46 | Fecal | Comp1 |
| Multiomic-SCFB | 9,10-DiHOME | -0,14 | 0,42 | Fecal | Comp1 |
| Multiomic-SCFB | cis-4-hydroxycyclohexylaceticacid | 0,87 | 0,61 | Fecal | Comp2 |
| Multiomic-SCFB | pyridoxamine | 0,34 | 0,38 | Fecal | Comp2 |
| Multiomic-SCFB | stachydrine | 0,21 | 0,25 | Fecal | Comp2 |
| Multiomic-SCFB | arachidonate(20:4n6) | 0,04 | 0,24 | Fecal | Comp2 |
| Multiomic-SCFB | cysteines-sulfate | 0,16 | 0,23 | Fecal | Comp2 |
| Multiomic-SCFB | nervonate(24:1n9)* | 0,19 | 0,22 | Fecal | Comp2 |
| Multiomic-SCFB | 3-methylglutaconate | 0,09 | 0,22 | Fecal | Comp2 |
| Multiomic-SCFB | valine | 0,08 | 0,21 | Fecal | Comp2 |
| Multiomic-SCFB | leucine | 0,00 | 0,2 | Fecal | Comp2 |
| Multiomic-SCFB | riboflavin(VitaminB2) | 0,07 | 0,07 | Fecal | Comp2 |
| Multiomic-SFB | 132_SCF | -0,77 | 1 | Serum | Comp1 |
| Multiomic-SFB | 194_ADA | -0,63 | 0,99 | Serum | Comp1 |
| Multiomic-SFB | 172_TNFRSF4 | -0,08 | 0,53 | Serum | Comp1 |
| Multiomic-SFB | 107_CDCP1 | -0,02 | 0,39 | Serum | Comp1 |
| Multiomic-SFB | 177_t-PA | -0,04 | 0,38 | Serum | Comp1 |
| Multiomic-SFB | 179_MIC-A | -0,47 | 1 | Serum | Comp2 |
| Multiomic-SFB | 166_MIP-1 alpha | -0,46 | 1 | Serum | Comp2 |
| Multiomic-SFB | 156_HGF | -0,25 | 1 | Serum | Comp2 |
| Multiomic-SFB | 183_CXCL13 | -0,29 | 0,99 | Serum | Comp2 |
| Multiomic-SFB | 174_AGRP | -0,24 | 0,99 | Serum | Comp2 |
| Multiomic-SFB | 181_VEGF-D | -0,30 | 0,97 | Serum | Comp2 |
| Multiomic-SFB | 178_HB-EGF | -0,20 | 0,96 | Serum | Comp2 |
| Multiomic-SFB | 182_HE4 | -0,25 | 0,95 | Serum | Comp2 |
| Multiomic-SFB | 168_VEGFR-2 | -0,17 | 0,94 | Serum | Comp2 |
| Multiomic-SFB | 137_CCL11 | -0,15 | 0,92 | Serum | Comp2 |
| Multiomic-SFB | 175_REG-4 | -0,22 | 0,91 | Serum | Comp2 |
| Multiomic-SFB | 103_AM | -0,11 | 0,81 | Serum | Comp2 |
| Multiomic-SFB | 116_KLK6 | -0,11 | 0,79 | Serum | Comp2 |
| Multiomic-SFB | 145_CCL19 | -0,11 | 0,73 | Serum | Comp2 |
| Multiomic-SFB | 184_EGFR | -0,06 | 0,58 | Serum | Comp2 |

| | | | | | |
|---|---|---|---|---|---|
| Multiomic-SFB | 195_IL-17RB | -0,10 | 0,56 | Serum | Comp2 |
| Multiomic-SFB | 113_IL-6 | 0,13 | 0,5 | Serum | Comp2 |
| Multiomic-SFB | 173_GAL | -0,07 | 0,4 | Serum | Comp2 |
| Multiomic-SFB | 121_hK11 | -0,01 | 0,36 | Serum | Comp2 |
| Multiomic-SFB | 177_AR | 0,00 | 0,29 | Serum | Comp2 |
| Multiomic-SFB | 170_4E-BP1 | -0,89 | 0,91 | Serum | Comp3 |
| Multiomic-SFB | 113_IL-6 | -0,41 | 0,77 | Serum | Comp3 |
| Multiomic-SFB | 128_CXCL1 | -0,13 | 0,39 | Serum | Comp3 |
| Multiomic-SFB | 136_MCP-4 | -0,09 | 0,36 | Serum | Comp3 |
| Multiomic-SFB | 134_IL-6RA | -0,15 | 0,26 | Serum | Comp3 |
| Multiomic-SFB | thioproline | -0,51 | 1 | Fecal | Comp1 |
| Multiomic-SFB | 2-aminoadipate | -0,44 | 1 | Fecal | Comp1 |
| Multiomic-SFB | flavinmononucleotide(FMN) | 0,50 | 0,99 | Fecal | Comp1 |
| Multiomic-SFB | 4-imidazoleacetate | -0,31 | 0,93 | Fecal | Comp1 |
| Multiomic-SFB | 2-hydroxydecanoate | -0,31 | 0,9 | Fecal | Comp1 |
| Multiomic-SFB | N-acetylglutamate | 0,25 | 0,76 | Fecal | Comp1 |
| Multiomic-SFB | 9,10-DiHOME | -0,12 | 0,63 | Fecal | Comp1 |
| Multiomic-SFB | 3-methyladipate | 0,11 | 0,48 | Fecal | Comp1 |
| Multiomic-SFB | 2'-deoxyinosine | 0,08 | 0,37 | Fecal | Comp1 |
| Multiomic-SFB | 1,3,7-trimethylurate | 0,01 | 0,12 | Fecal | Comp1 |
| Multiomic-SFB | 5-dodecenoate(12:1n7) | -0,63 | 0,98 | Fecal | Comp2 |
| Multiomic-SFB | myristoleate(14:1n5) | -0,57 | 0,95 | Fecal | Comp2 |
| Multiomic-SFB | 3-hydroxylaurate | -0,30 | 0,77 | Fecal | Comp2 |
| Multiomic-SFB | thymidine | -0,22 | 0,75 | Fecal | Comp2 |
| Multiomic-SFB | 1-palmitoyl-2-oleoyl-GPC(16:0/18:1) | 0,24 | 0,72 | Fecal | Comp2 |
| Multiomic-SFB | N-carbamoylaspartate | 0,21 | 0,7 | Fecal | Comp2 |
| Multiomic-SFB | glutamine | -0,15 | 0,62 | Fecal | Comp2 |
| Multiomic-SFB | 2'-deoxyuridine | -0,10 | 0,53 | Fecal | Comp2 |
| Multiomic-SFB | stearoylcarnitine | -0,09 | 0,45 | Fecal | Comp2 |
| Multiomic-SFB | deoxycarnitine | -0,01 | 0,38 | Fecal | Comp2 |
| Multiomic-SFB | uridine-2',3'-cyclicmonophosphate | -0,53 | 0,98 | Fecal | Comp3 |
| Multiomic-SFB | 3-methylglutaconate | -0,40 | 0,93 | Fecal | Comp3 |
| Multiomic-SFB | citramalate | 0,31 | 0,82 | Fecal | Comp3 |
| Multiomic-SFB | N-acetyl-3-methylhistidine* | 0,27 | 0,8 | Fecal | Comp3 |
| Multiomic-SFB | ethylmalonate | -0,24 | 0,77 | Fecal | Comp3 |
| Multiomic-SFB | N-acetylsphingosine | 0,26 | 0,76 | Fecal | Comp3 |
| Multiomic-SFB | caproate(6:0) | -0,18 | 0,66 | Fecal | Comp3 |
| Multiomic-SFB | guanosine | -0,12 | 0,63 | Fecal | Comp3 |
| Multiomic-SFB | ribose | -0,19 | 0,62 | Fecal | Comp3 |
| Multiomic-SFB | adenosine | -0,11 | 0,62 | Fecal | Comp3 |
| Multiomic-SFB | nervonate(24:1n9)* | -0,20 | 0,61 | Fecal | Comp3 |
| Multiomic-SFB | nicotinate | -0,14 | 0,6 | Fecal | Comp3 |
| Multiomic-SFB | stachydrine | -0,18 | 0,59 | Fecal | Comp3 |
| Multiomic-SFB | pseudouridine | -0,18 | 0,51 | Fecal | Comp3 |
| Multiomic-SFB | glutarate(pentanedioate) | -0,12 | 0,51 | Fecal | Comp3 |
| Multiomic-SFB | inosine | -0,08 | 0,51 | Fecal | Comp3 |
| Multiomic-SFB | xanthosine | -0,11 | 0,46 | Fecal | Comp3 |
| Multiomic-SFB | valerate | -0,12 | 0,45 | Fecal | Comp3 |
| Multiomic-SFB | carnitine | 0,07 | 0,39 | Fecal | Comp3 |
| Multiomic-SFB | stearoylethanolamide | -0,04 | 0,38 | Fecal | Comp3 |
| Multiomic-SFB | harmane | -0,03 | 0,38 | Fecal | Comp3 |
| Multiomic-SFB | deoxycarnitine | 0,04 | 0,36 | Fecal | Comp3 |
| Multiomic-SFB | spermidine | -0,02 | 0,36 | Fecal | Comp3 |
| Multiomic-SFB | benzoate | -0,01 | 0,32 | Fecal | Comp3 |
| Multiomic-SFB | pyridoxamine | -0,03 | 0,23 | Fecal | Comp3 |
| Multiomic-SFB | Bfe | -0,60 | 1 | Blood | Comp1 |
| Multiomic-SFB | BMg | -0,56 | 1 | Blood | Comp1 |
| Multiomic-SFB | BZn | -0,42 | 0,97 | Blood | Comp1 |
| Multiomic-SFB | Bca | 0,25 | 0,94 | Blood | Comp1 |
| Multiomic-SFB | BMn | -0,31 | 0,87 | Blood | Comp1 |
| Multiomic-SFB | Bco | 0,89 | 1 | Blood | Comp2 |
| Multiomic-SFB | Bmo | -0,42 | 0,9 | Blood | Comp2 |
| Multiomic-SFB | Bas | -0,13 | 0,66 | Blood | Comp2 |
| Multiomic-SFB | BMn | 0,06 | 0,52 | Blood | Comp2 |
| Multiomic-SFB | BCd | -0,71 | 1 | Blood | Comp3 |
| Multiomic-SFB | Bas | 0,41 | 1 | Blood | Comp3 |
| Multiomic-SFB | Bcu | -0,35 | 1 | Blood | Comp3 |
| Multiomic-SFB | Bca | -0,29 | 1 | Blood | Comp3 |
| Multiomic-SFB | Bmo | 0,18 | 1 | Blood | Comp3 |
| Multiomic-SFB | Bse | -0,16 | 1 | Blood | Comp3 |
| Multiomic-SFB | Bco | 0,15 | 1 | Blood | Comp3 |

| | | | | | |
|---|---|---|---|---|---|
| Multiomic-SFB | BZn | -0,14 | 1 | Blood | Comp3 |
| Multiomic-SFB | BPb | 0,12 | 1 | Blood | Comp3 |
| Multiomic-SFB | BHg | 0,11 | 1 | Blood | Comp3 |
| Multiomic-SFB | BMg | -0,05 | 1 | Blood | Comp3 |
| Multiomic-SFB | Bfe | 0,04 | 1 | Blood | Comp3 |
| Multiomic-SFB | BMn | -0,02 | 1 | Blood | Comp3 |
| Multiomic-UFB | unknown 677 | 0,91 | 1 | Urine | Comp1 |
| Multiomic-UFB | 2-hydroxyisobutyrate | 0,26 | 0,77 | Urine | Comp1 |
| Multiomic-UFB | 6-hydroxynicotinate | -0,21 | 0,75 | Urine | Comp1 |
| Multiomic-UFB | lactate | -0,17 | 0,66 | Urine | Comp1 |
| Multiomic-UFB | glucose | -0,18 | 0,64 | Urine | Comp1 |
| Multiomic-UFB | 3-hydroxyisovalerate | -0,80 | 0,63 | Urine | Comp2 |
| Multiomic-UFB | valine | -0,47 | 0,62 | Urine | Comp2 |
| Multiomic-UFB | HMB | -0,38 | 0,57 | Urine | Comp2 |
| Multiomic-UFB | isoleucine | -0,04 | 0,28 | Urine | Comp2 |
| Multiomic-UFB | unknown 33 | 0,00 | 0,25 | Urine | Comp2 |
| Multiomic-UFB | glycerol3-phosphate | -0,55 | 1 | Fecal | Comp1 |
| Multiomic-UFB | 3-carboxyadipate | -0,33 | 1 | Fecal | Comp1 |
| Multiomic-UFB | 2-acetamidobutanoate | 0,32 | 1 | Fecal | Comp1 |
| Multiomic-UFB | flavinmononucleotide(FMN) | -0,19 | 1 | Fecal | Comp1 |
| Multiomic-UFB | 1-stearoyl-2-oleoyl-GPC(18:0/18:1) | 0,27 | 0,99 | Fecal | Comp1 |
| Multiomic-UFB | 1-palmitoyl-2-oleoyl-GPC(16:0/18:1) | 0,26 | 0,99 | Fecal | Comp1 |
| Multiomic-UFB | 6-oxolithocholate | 0,26 | 0,99 | Fecal | Comp1 |
| Multiomic-UFB | 3,7-dimethylurate | 0,25 | 0,98 | Fecal | Comp1 |
| Multiomic-UFB | N-acetylalanine | 0,20 | 0,94 | Fecal | Comp1 |
| Multiomic-UFB | 5-methyluridine(ribothymidine) | -0,14 | 0,93 | Fecal | Comp1 |
| Multiomic-UFB | arabitol/xylitol | -0,12 | 0,91 | Fecal | Comp1 |
| Multiomic-UFB | aspartate | -0,13 | 0,9 | Fecal | Comp1 |
| Multiomic-UFB | 2'-deoxyuridine | -0,14 | 0,89 | Fecal | Comp1 |
| Multiomic-UFB | pseudouridine | -0,12 | 0,88 | Fecal | Comp1 |
| Multiomic-UFB | 13-methylmyristate | 0,16 | 0,87 | Fecal | Comp1 |
| Multiomic-UFB | 3-hydroxystearate | -0,09 | 0,82 | Fecal | Comp1 |
| Multiomic-UFB | stearate(18:0) | -0,09 | 0,68 | Fecal | Comp1 |
| Multiomic-UFB | gamma-aminobutyrate(GABA) | 0,06 | 0,67 | Fecal | Comp1 |
| Multiomic-UFB | 2,3-dimethylsuccinate | -0,07 | 0,66 | Fecal | Comp1 |
| Multiomic-UFB | daidzein | 0,03 | 0,62 | Fecal | Comp1 |
| Multiomic-UFB | 1H-quinolin-2-one | -0,04 | 0,59 | Fecal | Comp1 |
| Multiomic-UFB | 2'-deoxyinosine | -0,04 | 0,53 | Fecal | Comp1 |
| Multiomic-UFB | thymine | -0,03 | 0,5 | Fecal | Comp1 |
| Multiomic-UFB | 3,5-dihydroxybenzoicacid | -0,04 | 0,48 | Fecal | Comp1 |
| Multiomic-UFB | lithocholate | 0,01 | 0,43 | Fecal | Comp1 |
| Multiomic-UFB | pimelate(heptanedioate) | -0,02 | 0,41 | Fecal | Comp1 |
| Multiomic-UFB | thymidine | -0,01 | 0,41 | Fecal | Comp1 |
| Multiomic-UFB | N-acetylaspartate(NAA) | -0,01 | 0,34 | Fecal | Comp1 |
| Multiomic-UFB | tyrosol | 0,00 | 0,32 | Fecal | Comp1 |
| Multiomic-UFB | indole | 0,00 | 0,28 | Fecal | Comp1 |
| Multiomic-UFB | N-acetylmethioninesulfoxide | -0,76 | 0,45 | Fecal | Comp2 |
| Multiomic-UFB | 5-dodecenoate(12:1n7) | -0,24 | 0,34 | Fecal | Comp2 |
| Multiomic-UFB | N-formylmethionine | -0,61 | 0,31 | Fecal | Comp2 |
| Multiomic-UFB | 2,3-dihydroxyisovalerate | -0,05 | 0,23 | Fecal | Comp2 |
| Multiomic-UFB | 8-hydroxyguanine | 0,00 | 0,13 | Fecal | Comp2 |
| Multiomic-UFB | BMn | 0,82 | 1 | Blood | Comp1 |
| Multiomic-UFB | BHg | -0,43 | 1 | Blood | Comp1 |
| Multiomic-UFB | BMg | 0,36 | 0,97 | Blood | Comp1 |
| Multiomic-UFB | Bas | -0,12 | 0,86 | Blood | Comp1 |
| Multiomic-UFB | Bse | 0,61 | 1 | Blood | Comp2 |
| Multiomic-UFB | BCd | 0,56 | 1 | Blood | Comp2 |
| Multiomic-UFB | Bca | 0,33 | 1 | Blood | Comp2 |
| Multiomic-UFB | BPb | 0,27 | 1 | Blood | Comp2 |
| Multiomic-UFB | BZn | -0,22 | 1 | Blood | Comp2 |
| Multiomic-UFB | Bfe | -0,18 | 1 | Blood | Comp2 |
| Multiomic-UFB | Bmo | -0,14 | 1 | Blood | Comp2 |
| Multiomic-UFB | BMg | -0,14 | 1 | Blood | Comp2 |
| Multiomic-UFB | Bas | 0,10 | 1 | Blood | Comp2 |
| Multiomic-UFB | Bcu | 0,07 | 1 | Blood | Comp2 |
| Multiomic-UFB | BMn | 0,07 | 1 | Blood | Comp2 |
| Multiomic-UFB | Bco | -0,02 | 1 | Blood | Comp2 |
| Multiomic-UFB | BHg | -0,02 | 1 | Blood | Comp2 |

**Table S17. Missingness of key molecules identified in multiomics models.**

| Block | Molecules | Missingness |
|---|---|---|
| Blood metal | BMg | 0 |
| Blood metal | Bca | 0 |
| Blood metal | BMn | 0 |
| Blood metal | Bfe | 0 |
| Blood metal | Bco | 0 |
| Blood metal | Bcu | 0 |
| Blood metal | BZn | 0 |
| Blood metal | Bas | 0 |
| Blood metal | Bse | 0 |
| Blood metal | Bmo | 0 |
| Blood metal | BCd | 0 |
| Blood metal | BHg | 0,009090909 |
| Blood metal | BPb | 0 |
| Serum protein | 177_t-PA | 0 |
| Serum protein | 139_PRL | 0 |
| Serum protein | 181_VEGF-D | 0 |
| Serum protein | 117_Gal-3 | 0 |
| Serum protein | 107_CDCP1 | 0,134920635 |
| Serum protein | 114_CSTB | 0 |
| Serum protein | 190_CCL20 | 0 |
| Serum protein | 142_FasL | 0 |
| Serum protein | 143_BAFF | 0 |
| Serum protein | 110_OPG | 0 |
| Serum protein | 132_SCF | 0 |
| Serum protein | 179_MIC-A | 0,126984127 |
| Serum protein | 113_IL-6 | 0,007936508 |
| Serum protein | 164_CCL23 | 0,134920635 |
| Serum protein | 183_CXCL13 | 0 |
| Serum protein | 164_CTSD | 0 |
| Serum protein | 194_ADA | 0,134920635 |
| Serum protein | 157_LITAF | 0,079365079 |
| Serum protein | 134_IL-6RA | 0 |
| Serum protein | 112_IL-1ra | 0 |
| Serum protein | 172_TNFRSF4 | 0 |
| Serum protein | 153_ErbB2/HER2 | 0 |
| Serum protein | 195_IL-17RB | 0,087301587 |
| Serum protein | 135_TNF-R2 | 0 |
| Serum protein | 165_CDH3 | 0 |
| Serum protein | 154_ErbB3/HER3 | 0 |
| Serum protein | 131_FGF-23 | 0 |
| Serum protein | 174_AGRP | 0 |
| Serum protein | 177_AR | 0 |
| Serum protein | 175_REG-4 | 0 |
| Serum protein | 183_SPON1 | 0 |
| Serum protein | 150_CHI3L1 | 0,007936508 |
| Serum protein | 179_ESM-1 | 0 |
| Serum protein | 145_CCL19 | 0 |
| Serum protein | 166_MIP-1 alpha | 0,134920635 |
| Serum protein | 182_MMP-12 | 0 |
| Serum protein | 189_TWEAK | 0,134920635 |
| Serum protein | 155_ErbB4/HER4 | 0 |
| Serum protein | 171_Dkk-1 | 0 |
| Serum protein | 152_Ep-CAM | 0 |
| Serum protein | 156_HGF | 0 |
| Serum protein | 103_AM | 0 |
| Serum protein | 101_IL-8 | 0 |
| Serum protein | 148_PTX3 | 0 |
| Serum protein | 148_PTPN22 | 0,023809524 |
| Serum protein | 145_PAPPA | 0,031746032 |
| Serum protein | 142_MMP-1 | 0,007936508 |
| Serum protein | 159_MIA | 0 |
| Serum protein | 173_GAL | 0 |
| Serum protein | 151_ST2 | 0 |
| Serum protein | 178_IFN-gamma | 0,126984127 |
| Serum protein | 158_MB | 0 |
| Serum protein | 149_EMMPRIN | 0 |

| | | |
|---|---|---|
| Serum protein | 170_4E-BP1 | 0,134920635 |
| Serum protein | 182_HE4 | 0 |
| Serum protein | 178_HB-EGF | 0 |
| Serum protein | 168_VEGFR-2 | 0 |
| Serum protein | 137_CCL11 | 0,134920635 |
| Serum protein | 116_KLK6 | 0 |
| Serum protein | 184_EGFR | 0 |
| Serum protein | 121_hK11 | 0,015873016 |
| Serum protein | 128_CXCL1 | 0 |
| Serum protein | 136_MCP-4 | 0,134920635 |
| CSF protein | 141_GH | 0,034883721 |
| CSF protein | 140_FUR | 0 |
| CSF protein | 131_IL-7 | 0,034883721 |
| CSF protein | 160_CXCL5 | 0 |
| CSF protein | 110_ILT-3 | 0 |
| CSF protein | 193_FS | 0,034883721 |
| CSF protein | 157_PSGL-1 | 0,197674419 |
| CSF protein | 171_Dkk-1 | 0,034883721 |
| CSF protein | 187_TNFRSF9 | 0 |
| CSF protein | 185_ITGA1 | 0 |
| CSF protein | 151_IL-18R1 | 0 |
| CSF protein | 182_HE4 | 0 |
| CSF protein | 120_TRAIL | 0,034883721 |
| CSF protein | 113_IL-6 | 0,034883721 |
| CSF protein | 151_CXCL10 | 0 |
| CSF protein | 164_CTSD | 0,034883721 |
| CSF protein | 179_ESM-1 | 0,034883721 |
| CSF protein | 194_ADA | 0 |
| CSF protein | 152_Ep-CAM | 0 |
| CSF protein | 177_t-PA | 0,034883721 |
| CSF protein | 149_IL-10RB | 0 |
| CSF protein | 166_CCL3 | 0,151162791 |
| CSF protein | 196_FR-alpha | 0 |
| CSF protein | 115_MCP-1 | 0,034883721 |
| Fecal metabolite | 3-methyladipate | 0,08994709 |
| Fecal metabolite | 2'-deoxyinosine | 0,010582011 |
| Fecal metabolite | N-acetylglutamate | 0 |
| Fecal metabolite | taurochenodeoxycholate | 0,380952381 |
| Fecal metabolite | 9,10-DiHOME | 0,021164021 |
| Fecal metabolite | cis-4-hydroxycyclohexylaceticacid | 0,021164021 |
| Fecal metabolite | pyridoxamine | 0 |
| Fecal metabolite | stachydrine | 0,005291005 |
| Fecal metabolite | nervonate(24:1n9)* | 0,296296296 |
| Fecal metabolite | cysteines-sulfate | 0,021164021 |
| Fecal metabolite | 3-methylglutaconate | 0,037037037 |
| Fecal metabolite | valine | 0 |
| Fecal metabolite | riboflavin(VitaminB2) | 0 |
| Fecal metabolite | arachidonate(20:4n6) | 0 |
| Fecal metabolite | leucine | 0 |
| Fecal metabolite | thioproline | 0,211640212 |
| Fecal metabolite | flavinmononucleotide(FMN) | 0,132275132 |
| Fecal metabolite | 2-aminoadipate | 0 |
| Fecal metabolite | 4-imidazoleacetate | 0 |
| Fecal metabolite | 2-hydroxydecanoate | 0,010582011 |
| Fecal metabolite | 1,3,7-trimethylurate | 0,047619048 |
| Fecal metabolite | 5-dodecenoate(12:1n7) | 0,026455026 |
| Fecal metabolite | myristoleate(14:1n5) | 0 |
| Fecal metabolite | 3-hydroxylaurate | 0,005291005 |
| Fecal metabolite | 1-palmitoyl-2-oleoyl-GPC(16:0/18:1) | 0 |
| Fecal metabolite | thymidine | 0 |
| Fecal metabolite | N-carbamoylaspartate | 0,021164021 |
| Fecal metabolite | glutamine | 0 |
| Fecal metabolite | 2'-deoxyuridine | 0 |
| Fecal metabolite | stearoylcarnitine | 0,312169312 |
| Fecal metabolite | deoxycarnitine | 0 |
| Fecal metabolite | uridine-2',3'-cyclicmonophosphate | 0,291005291 |
| Fecal metabolite | citramalate | 0 |
| Fecal metabolite | N-acetyl-3-methylhistidine* | 0,126984127 |
| Fecal metabolite | N-acetylsphingosine | 0,44973545 |
| Fecal metabolite | ethylmalonate | 0 |

| | | |
|---|---|---|
| Fecal metabolite | ribose | 0 |
| Fecal metabolite | caproate(6:0) | 0,074074074 |
| Fecal metabolite | pseudouridine | 0 |
| Fecal metabolite | nicotinate | 0 |
| Fecal metabolite | valerate | 0 |
| Fecal metabolite | glutarate(pentanedioate) | 0 |
| Fecal metabolite | guanosine | 0 |
| Fecal metabolite | xanthosine | 0,227513228 |
| Fecal metabolite | adenosine | 0 |
| Fecal metabolite | inosine | 0 |
| Fecal metabolite | carnitine | 0 |
| Fecal metabolite | stearoylethanolamide | 0,010582011 |
| Fecal metabolite | harmane | 0,264550265 |
| Fecal metabolite | spermidine | 0,031746032 |
| Fecal metabolite | benzoate | 0,26984127 |
| Fecal metabolite | glycerol3-phosphate | 0,010582011 |
| Fecal metabolite | 3-carboxyadipate | 0,005291005 |
| Fecal metabolite | 2-acetamidobutanoate | 0,137566138 |
| Fecal metabolite | 1-stearoyl-2-oleoyl-GPC(18:0/18:1) | 0,132275132 |
| Fecal metabolite | 6-oxolithocholate | 0,148148148 |
| Fecal metabolite | 3,7-dimethylurate | 0,111111111 |
| Fecal metabolite | N-acetylalanine | 0 |
| Fecal metabolite | 13-methylmyristate | 0,195767196 |
| Fecal metabolite | 5-methyluridine(ribothymidine) | 0,021164021 |
| Fecal metabolite | aspartate | 0 |
| Fecal metabolite | arabitol/xylitol | 0,238095238 |
| Fecal metabolite | stearate(18:0) | 0 |
| Fecal metabolite | 3-hydroxystearate | 0,211640212 |
| Fecal metabolite | 2,3-dimethylsuccinate | 0 |
| Fecal metabolite | gamma-aminobutyrate(GABA) | 0 |
| Fecal metabolite | 3,5-dihydroxybenzoicacid | 0,169312169 |
| Fecal metabolite | 1H-quinolin-2-one | 0 |
| Fecal metabolite | thymine | 0 |
| Fecal metabolite | daidzein | 0,264550265 |
| Fecal metabolite | pimelate(heptanedioate) | 0,169312169 |
| Fecal metabolite | N-acetylaspartate(NAA) | 0 |
| Fecal metabolite | lithocholate | 0,005291005 |
| Fecal metabolite | indole | 0,285714286 |
| Fecal metabolite | tyrosol | 0,021164021 |
| Fecal metabolite | N-acetylmethioninesulfoxide | 0 |
| Fecal metabolite | N-formylmethionine | 0 |
| Fecal metabolite | 2,3-dihydroxyisovalerate | 0,015873016 |
| Fecal metabolite | 8-hydroxyguanine | 0,079365079 |
| urine metabolite | unknown 677 | 0 |
| urine metabolite | 2-hydroxyisobutyrate | 0 |
| urine metabolite | 6-hydroxynicotinate | 0 |
| urine metabolite | glucose | 0 |
| urine metabolite | lactate | 0 |
| urine metabolite | 3-hydroxyisovalerate | 0 |
| urine metabolite | valine | 0 |
| urine metabolite | HMB | 0 |
| urine metabolite | isoleucine | 0 |
| urine metabolite | unknown 33 | 0 |
